# Biventricular cardiac dynamic shape: genetics and cardiometabolic disease associations

**DOI:** 10.64898/2026.04.19.26350940

**Authors:** Richard Burns, William J. Young, Kulsuma Uddin, Steffen E Petersen, Julia Ramírez, Alistair A Young, Patricia B Munroe

**Affiliations:** School of Biomedical Engineering and Imaging Sciences, King’s College London, London, UK; William Harvey Research Institute, Queen Mary University London, Charterhouse Square, London, UK; Barts Heart Centre, St Bartholomew’s Hospital, Barts Health NHS Trust, West Smithfield, London, UK; NIHR Barts Biomedical Research Centre, Queen Mary of University London, Charterhouse Square, London, UK; Aragon Institute of Engineering Research, University of Zaragoza, Zaragoza, Spain; Centro de Investigación Biomédica en Red – Bioingeniería, Biomateriales y Nanomedicina, Zaragoza, Spain

**Author notes:** Indicates joint supervisors (Julia Ramírez, Alistair A Young, Patricia B Munroe). indicates corresponding authors.

**Keywords:** biventricular, dynamic cardiac shape and function, cardiometabolic disease, genome-wide association study

## Abstract

**Background:** Genetic studies using cardiac magnetic resonance (CMR) imaging have identified loci related to cardiac shape, but most focus on static morphology. The value of a dynamic cardiac shape atlas capturing both shape and function remains unknown.

**Methods:** A dynamic shape atlas comprising CMR-derived shape models at end-diastole and end-systole was combined with genetic and outcome data in 36,992 UK Biobank participants. Dynamic shape principal components (PCs) describing >1% of variance were characterized, and tested for associations with prevalent and incident cardiometabolic diseases, including ischemic heart disease (IHD), heart failure (HF), significant atrioventricular block (AVB), and atrial fibrillation (AF), and independent predictive power alongside standard CMR measures. Genome-wide association studies (GWAS) were performed to identify candidate genes and biological pathways, and polygenic risk scores (PRS) were assessed for disease associations. Mendelian randomization (MR) was performed to test causality of observed disease associations.

**Results:** We identified 14 dynamic cardiac shape PCs capturing 83.3% of total dynamic cardiac shape variance. These PCs captured distinct functional remodeling patterns such as variation in annular plane systolic excursion, while remaining only modestly correlated with standard CMR measures. All 14 PCs were associated with at least one incident cardiometabolic disease, with the strongest associations observed for incident IHD, HF, and AVB. Notably, incorporating dynamic shape PCs improved the prediction of incident IHD beyond standard CMR measures. GWAS identified 75 genetic loci associated with dynamic shape, including 14 variants previously unreported for cardiac traits, and candidate genes demonstrated enrichment in pathways related to cardiac development and contractile function. PRS derived from dynamic shape loci were significantly associated with multiple outcomes, most prominently HF. MR identified significant causal relationships between several PCs and cardiometabolic disease.

**Conclusions:** Dynamic cardiac shape features capture aspects of cardiac structure and function not fully represented by standard CMR measures. These features are strongly associated with incident cardiometabolic disease and provide new insights into the genetic architecture of cardiac remodeling.

**Clinical perspective:** *What is new?:*

- Genetic and outcome relationships with a dynamic statistical shape model capturing both left and right ventricles at end-diastole and end-systole.
- Demonstration of incremental value over existing cardiac shape models, through capture of functional remodeling not represented by standard imaging measures.
- Identification of genetic susceptibility loci for dynamic cardiac shape, including 14 variants not previously reported for cardiac traits.

*What are the clinical implications?:*

- The results enhance our understanding of the genetic architecture of dynamic cardiac shape and function in the general population and clarify their relationships with other cardiovascular endophenotypes and incident cardiometabolic diseases.
- Newly identified candidate genes expand the biological pathways implicated in cardiac remodeling and provide targets for future functional and mechanistic studies.
- The improved prediction of incident cardiometabolic disease, particularly ischemic heart disease, achieved by adding dynamic shape PCs to traditional CMR measures suggests potential value for their inclusion in evaluation of patients.

## Introduction

The relationships between disease and cardiac morphology are bidirectional, in that disease can affect cardiac morphology through remodeling, and morphology can influence risk of cardiac disease incidence through physiological feedback mechanisms. For example, in ischemic heart disease (IHD) impaired blood flow may trigger myocardial remodeling, including hypertrophy or fibrosis^1–4^, but pre-existing ventricular hypertrophy may conversely increase myocardial oxygen demand leading to ischemic conditions ^5,6^. Cardiac structure and function measurements are therefore critical not only for diagnosing cardiovascular disease, but also for understanding disease risk mechanisms, pathogenesis, and prognosis^7,8^. However, standard measures such as ventricular volumes, masses, wall thicknesses, and ejection fraction do not capture all the morphological information present in modern cardiac imaging examinations.

Statistical cardiac shape atlas scores have established relationships with clinically meaningful measures^9–12^, demonstrate stronger associations with prevalent disease than standard measures^11,13^, and independently predict incident cardiovascular events^14^. However, the biological interpretation of the main drivers of shape variation, disease-shape relationships, and their genetic underpinnings remain limited. In particular, understanding the genetic basis of three-dimensional cardiac shape and motion may provide important insights into mechanisms underlying disease development.

Standard imaging-derived measures have a substantial genetic basis, including left and right ventricular end-diastolic (ED) and end-systolic (ES) volumes, myocardial mass, ejection fraction, myocardial strain, wall thickness, and sphericity^15–25^. Polygenic risk scores (PRS) derived from these traits have shown predictive value for cardiometabolic diseases, including cardiomyopathies^26–28^, heart failure^29,30^, atrial fibrillation^31–33^, and type 2 diabetes mellitus^31,34,35^. More complex imaging-derived phenotypes have further expanded locus discovery, including regional wall thickness^25,36^, trabecular complexity quantified by fractal dimension^37^, and latent phenotypes derived from machine learning approaches^38–40^. Our previous work demonstrated that ED biventricular shape atlas scores have a significant genetic basis, with 43 associated loci^41^. However, analysis of coupled ED-ES shape and function, which utilizes information across the cardiac cycle and can better capture associations with cardiac functional disease, remains unexplored.

In this study, we investigated the relationships between a dynamic shape atlas, combining ED and ES shape in both left and right ventricles^42^, and both prevalent and incident cardiometabolic disease in 36,992 participants of the UK Biobank, and evaluated the additive value of these phenotypes beyond standard measures in predictive models. We performed genome-wide association studies (GWAS), gene-burden testing, and downstream bioinformatic analyses to characterize the genetic basis of dynamic cardiac shape and compared findings with our previously published ED-only shape GWAS^41^. We constructed PRS and conducted Mendelian randomization (MR) analyses to explore potential causal relationships between genetically determined shape variation and cardiometabolic disease. We hypothesized that dynamic cardiac shape phenotypes capture complementary structural and functional information that enables identification of novel loci, provides insight into cardiac remodeling mechanisms, and improves prediction of incident cardiometabolic disease beyond standard imaging measures.

## Methods

### Data availability

All UK Biobank (UKB) data are available upon application (www.ukbiobank.ac.uk). Summary GWAS statistics will be publicly available through the GWAS catalog (https://www.ebi.ac.uk/gwas) on publication.

### Study population and cardiac phenotyping

UK Biobank is a large prospective cohort of approximately 500,000 participants aged 40-69 years at recruitment, with extensive demographic, clinical, lifestyle, and genetic data. Genotype data were available for 488,377 individuals, generated using the UK BiLEVE and UK Biobank Axiom arrays. Study design, genotyping, and quality control procedures have been described previously^43,44^.

This study included cardiovascular magnetic resonance (CMR) imaging data from 45,683 participants, which has been described previously^45^. Briefly, images were acquired on a 1.5T wide-bore scanner (MAGNETOM Aera, Siemens Healthcare) using ECG-gating. Short-axis cine images covered both ventricles with 8 mm slice thickness and 2 mm inter-slice gap (TR=2.6msec, TE=1.10msec), interpolated to 50 phases per cardiac cycle. Long-axis cine images (2-, 3-, and 4- chamber views) were acquired with 6 mm slice thickness (TR 2.7 ms, TE 1.16 ms).

### Image analysis and dynamic shape atlas construction

The automated image analysis and shape atlas pipeline has been described previously^42^. Briefly, short- and long-axis cine images were analyzed using Circle cvi42 (version 5.11 1505, Circle Cardiovascular Imaging Inc., Calgary, Canada), using deep learning convolutional neural networks to segment left ventricular (LV) endocardial and epicardial surfaces and right ventricular (RV) endocardial surfaces. Valve landmarks for the mitral, tricuspid, and aortic valves were also automatically extracted from long-axis views.

Biventricular subdivision surface meshes were generated at ED and ES using a non-rigid diffeomorphic registration framework^46^. Short- and long-axis contours were first aligned to correct for breath-hold misregistration, after which a template mesh was iteratively deformed to minimize contour-to-surface distances while preserving diffeomorphic properties.

A statistical shape atlas was constructed by aligning ED meshes using generalized Procrustes alignment^47^ (excluding scaling to preserve size variation). The same transformation was applied to both ED and ES meshes, preserving individual-specific ED-ES displacement patterns. ED and ES were concatenated into a single vector per individual, and principal component analysis (PCA) was applied. Fourteen dynamic cardiac shape PCs explaining >1% of total variance each (83.3% cumulative variance) were retained, capturing joint morphological and functional variation across the cohort.

### Shape quality control

Quality control procedures were performed to exclude cases in which the shape modeling process failed, as described previously^42^. Briefly, cases with incomplete information needed to construct a 3D shape model were excluded. Outlier shape models (chamber volumes exceeding 5 IQR) at ED or ES were excluded prior to PCA. After PCA, shape score outliers exceeding 3 IQR in Mahalanobis distance were excluded, and models with residual distance between the model surface and the PCA representation exceeding 3 IQR were excluded.

### Associations with conventional CMR measures and ED shape atlas

To assess relationships between dynamic shape PCs and standard CMR-derived measures, Pearson correlations were computed for left and right ventricular volumes, masses, and ejection fractions. Associations between dynamic shape PCs and cardiometabolic disease risk factors were calculated similarly. Correlations with previously reported ED-only shape PCs^41^ were also calculated to compare static and dynamic atlases. Analyses were performed in R (version 4.3.1).

### Disease association analyses

Associations between dynamic shape PCs and cardiometabolic disease were assessed for six well-represented cardiometabolic diseases encompassing the different aspects of cardiac pathology in the UK Biobank: atrial fibrillation (AF), significant atrioventricular block (AVB), heart failure (HF), IHD, stroke, and type 2 diabetes (T2D). Multivariable logistic regression was used for prevalent disease and Cox proportional hazards models for incident disease. Covariates, selected through stepwise regression, included age at imaging, sex, height, body mass index (BMI), systolic and diastolic blood pressure (averaged between automated and manual readings and adjusted for medication use), serum high-density cholesterol (HDL), low-density lipoprotein (LDL) and mean triglyceride levels, smoking and alcohol consumption (Supplementary Table 1). Missing covariate data were imputed using the MICE^48^, version 3.16.0) imputation package in R. Significant associations were defined at P < 0.0083 (Bonferroni corrected at 0.05/6 diseases tested). Disease information was obtained from hospital episode statistics data through UKB. ICD9, ICD10 and OPCS-4 codes were used to define each disease (Supplementary Table 2). Prevalent and incident disease were defined using the date of each participant’s CMR imaging and the date of disease diagnosis.

### Incident disease prediction: contribution of dynamic shape PCs beyond standard CMR measures

We tested whether incorporating dynamic cardiac shape PCs alongside standard CMR measures improves prediction of incident cardiometabolic disease compared with models based on standard CMR measures alone. Cox proportional hazards regression was used to construct two models for each outcome.

The baseline model included standard CMR measures (left and right ventricular ED and ES volumes, myocardial masses, ejection fractions, stroke volumes, and global circumferential and longitudinal strain), together with established clinical covariates: sex, age, height, BMI, SBP and DBP, smoking status, alcohol consumption, serum HDL, LDL and triglyceride levels. The extended model included dynamic cardiac shape PCs in addition to the CMR measures and covariates retained from the baseline model.

Separate prediction cohorts were defined within UK Biobank for each cardiometabolic outcome (AF, AVB, HF, IHD, stroke, and T2D). Participants with a diagnosis of the relevant disease prior to their UK Biobank imaging assessment date were excluded. Incident disease was defined using linked electronic health record ICD codes (Supplementary Table 2).

A penalized Cox proportional hazards regression model framework with elastic net regularization within R glmnet^49,50^ was used to model time to event for incident cardiometabolic diseases with equal lasso and ridge penalties to balance variable selection and coefficient shrinkage. The optimal regularization parameter was selected using internal cross-validation within the training data. Two models were created; the first a CMR model which combined clinical covariates identified to be significantly associated with each disease through stepwise regression (Supplementary Table 1) and CMR measures; and the second with these same variables as well as dynamic cardiac shape PCs.

Model performance was assessed using five-fold cross validation, calculating predictive performance using concordance index (C-index). C-index variability, non-parametric bootstrap resampling was performed (with 1,000 samples), and the 95% confidence intervals are reported alongside the C-index.

### Genetic analyses

A more detailed description of genetic analyses can be found in Supplementary Methods. Heritability of dynamic shape PCs was estimated in 36,992 unrelated white European participants with high-quality imaging data, no previous history of major cardiovascular disease, and a left ventricular ejection fraction of >40%, using BOLT-REML^51^ version 2.3. GWAS were performed for each PC using a linear mixed model method (BOLT-LMM^52^, version 2.4.1), testing 558,994 variants (minor allele frequency ≥1% and INFO >0.3), adjusting for age, sex, height, BMI, blood pressure, heart rate, array type and the top 10 genetic principal components. Genome-wide significance was defined as P < 5×10^-8^. A genomic signal was defined as the most significant (lead) SNV and all SNVs within a 1mb window in linkage disequilibrium (LD) r^2^ > 0.1. Population stratification and confounding were assessed using LD score software (LDSC)^53^, v1.0.1. Secondary signals were identified using GCTA conditional analysis^54^, version 1.94.1.

### Functional annotation and pathway analyses

Candidate genes were prioritized using variant effect prediction (VEP)^55–57^, expression quantitative trait locus (eQTL) colocalization analysis^58^, transcriptome-wide association studies^59^, Hi-C long-range chromatin interactions^60–62^, Polygenic Priority Score (PoPS)^63^, literature review, and mouse knockout phenotypes (Supplementary methods)^64^. Candidate genes were queried using Functional Mapping and Annotation of Genome-Wide Association Studies (FUMA GWAS)^65^ to perform functional enrichment analysis. Significant enrichment was defined as an adjusted False Discovery Rate P < 0.05.

### Rare variant analysis

We used Regenie^66^ (v4.0) to assess the contribution of rare genetic variants to dynamic cardiac shape PCs, and prespecified variant masks and annotations were used. Leave one variant out (LOVO) analysis was performed to assess whether significant results were driven by single or multiple rare variants (Supplementary methods).

### Genetic relationship with end-diastolic shape atlas

We assessed the genetic correlation between the dynamic cardiac shape atlas and previously published end-diastolic shape atlas using the software tool LDSC^53^ (v1.0.1). All dynamic cardiac shape lead variants and high LD proxies (r^2^ > 0.8) were also searched within the summary statistics of the end-diastolic shape GWAS, and results of variants with the lowest P-value among the 11 end-diastolic PCs are reported and shared signals across the cardiac atlases are indicated.

### Pleiotropy and PheWAS analyses

To assess novelty of the EDES GWAS results and identify which genetic signals were not previously reported for cardiovascular traits, we collated a list of all genome-wide significant variants and high LD proxies (r^2^ > 0.8) from a literature review of cardiac trait (cardiovascular diseases, electrocardiogram and measures of cardiac structure and function) GWAS and in the GWAS Catalog^67^. If an EDES GWAS lead variant was within 500kb of a previously reported SNV associated with a cardiovascular trait, and is in LD r^2^ ≥ 0.1, the association is reported.

We performed a phenome-wide association study (PheWAS) using the R package PheWAS^68^ (version 0.99.6.1) to assess relationships between dynamic cardiac shape PC variants and curated phecodes for clinical conditions (only including case numbers ≥200), and to test PRS for these PCs with the same phecodes. The analysis was performed on UK Biobank individuals not included in the GWAS, and was adjusted for age, sex and the first 10 genetic principal components. A Bonferroni corrected threshold (number of phecodes tested 0.05/325 = 1.54 x 10^-4^) was used to declare significance.

### Polygenic risk scoring

We used PRSice-2^69^ (version 2.3.5) to calculate each PRS, summing the dosage of each dynamic cardiac shape lead variant allele weighted by effect size (from GWAS) in the full UK Biobank genotyping cohort of 487,409 individuals. Associations were calculated through logistic regression of standard deviation increase in standardized PRS against cardiometabolic disease incidence in 447,408 unrelated (kinship pairs < 0.0884) individuals not included in the GWAS cohort. Disease outcomes were AF, AVB, HF, IHD, stroke and T2D (Supplementary Methods). Logistic regression models were adjusted for covariates identified to be significant for each disease through stepwise regression, selecting from age, sex, height, BMI, alcohol consumption, systolic and diastolic blood pressures, smoking status, LDL and mean triglyceride levels (Supplementary Table 1), the genetic array used and the first 15 genetic principal components. For the analysis the first visit covariate data was used. A P-value threshold of 0.00833 was used to define significant associations (Bonferroni corrected 0.05/number of diseases).

### Mendelian Randomization

To explore the directionality and potential causality underlying significant associations between dynamic cardiac shape PCs and cardiometabolic diseases, Mendelian randomization (MR) analyses were performed using the TwoSampleMR^70^ R package (version 0.6.6). PC-disease pairs selected for MR were based on statistically significant associations identified in regression analyses of dynamic shape PCs with prevalent and/or incident disease (Supplementary methods).

For associations with prevalent disease, where disease-related remodeling could influence cardiac shape, MR analyses were conducted with disease status as the exposure and dynamic cardiac shape PCs as the outcome. Conversely, for associations with incident disease, where cardiac shape may confer risk of future disease, MR analyses were performed with dynamic cardiac shape PCs as the exposure and disease incidence as the outcome. Additional methodological details, including instrument selection, sensitivity analyses, and assessment of pleiotropy and heterogeneity, are provided in the Supplementary methods.

## Results

An overview of the study design is shown in Figure 1, with demographic details of the imaging and full UKB cohorts provided in Table 1. Of 45,683 individuals with suitable CMR images, 41,659 passed shape quality control and were included in the dynamic cardiac shape atlas. Fourteen PCs, each explaining > 1% of total variance were retained, jointly accounting for 83.3% of overall shape variability (Table 2). Genetic and shape data were available in 36,992 participants with complete covariate data.

**Figure 1.**
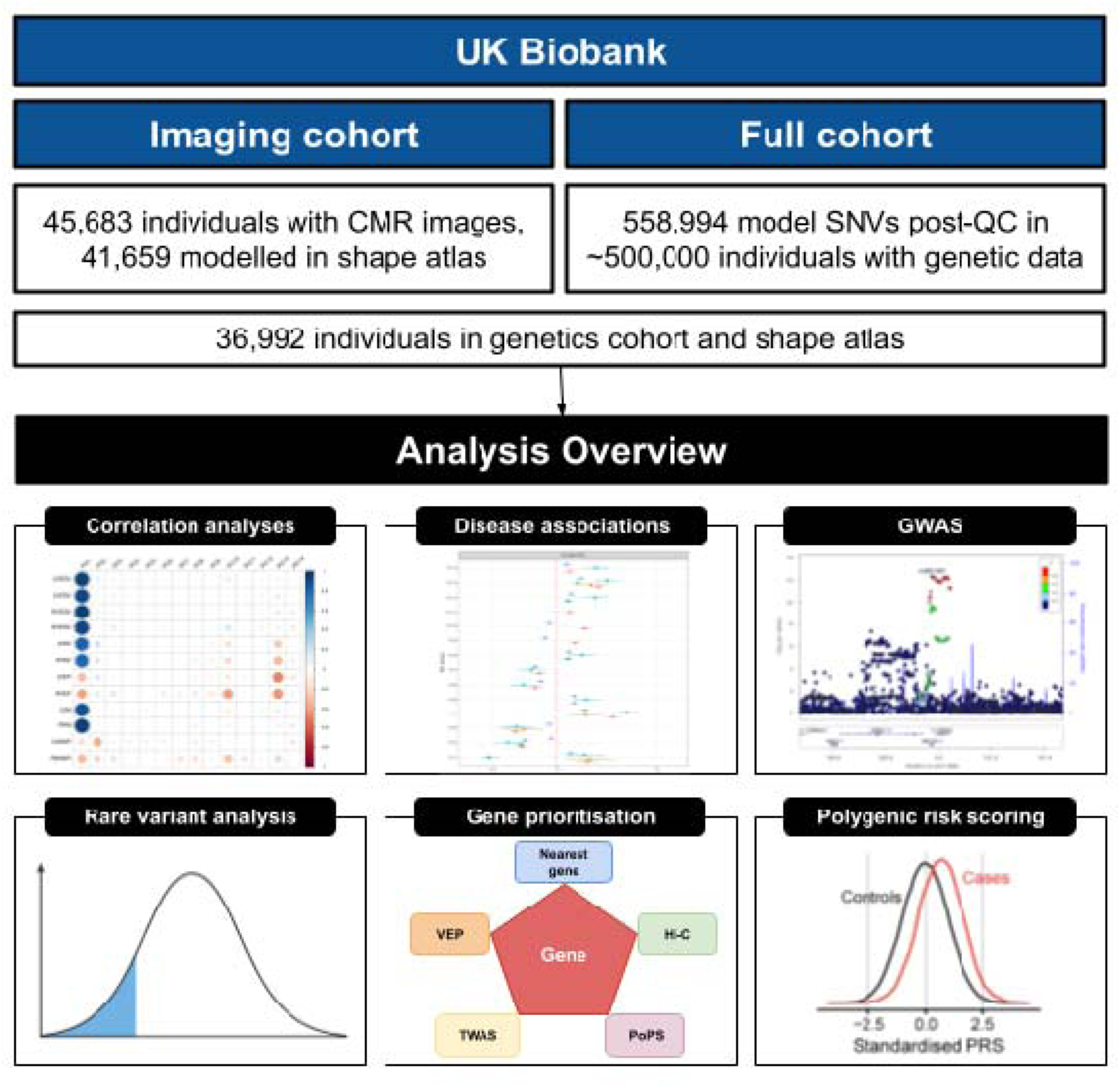
Study overview. Graphical overview of the study cohorts and analyses performed. In blue the cohorts used in the imaging and genetics analyses with the sample size for the GWAS indicated. In black there are graphics of the main analyses performed; correlation analyses between dynamic shape PCs and CMR measures, cardiometabolic disease risk factors and end-diastolic PCs; regression analyses between dynamic shape PCs and prevalent and incident cardiometabolic disease; GWAS for dynamic shape PCs; rare variant analysis to identify rare frequency variants associated with dynamic cardiac shape; a gene prioritization bioinformatics pipeline for identifying candidate genes at genome-wide significant loci; polygenic risk scoring and regression analyses to calculate associations between dynamic shape PCs and prevalent and incident cardiometabolic disease. Abbreviations: CMR, cardiac magnetic resonance (imaging); SNV, single nucleotide variant; QC, quality control; GWAS, genome-wide association study; VEP, variant effect prediction; TWAS, transcriptome-wide association study; PRS, polygenic risk score.

**Table 1.**
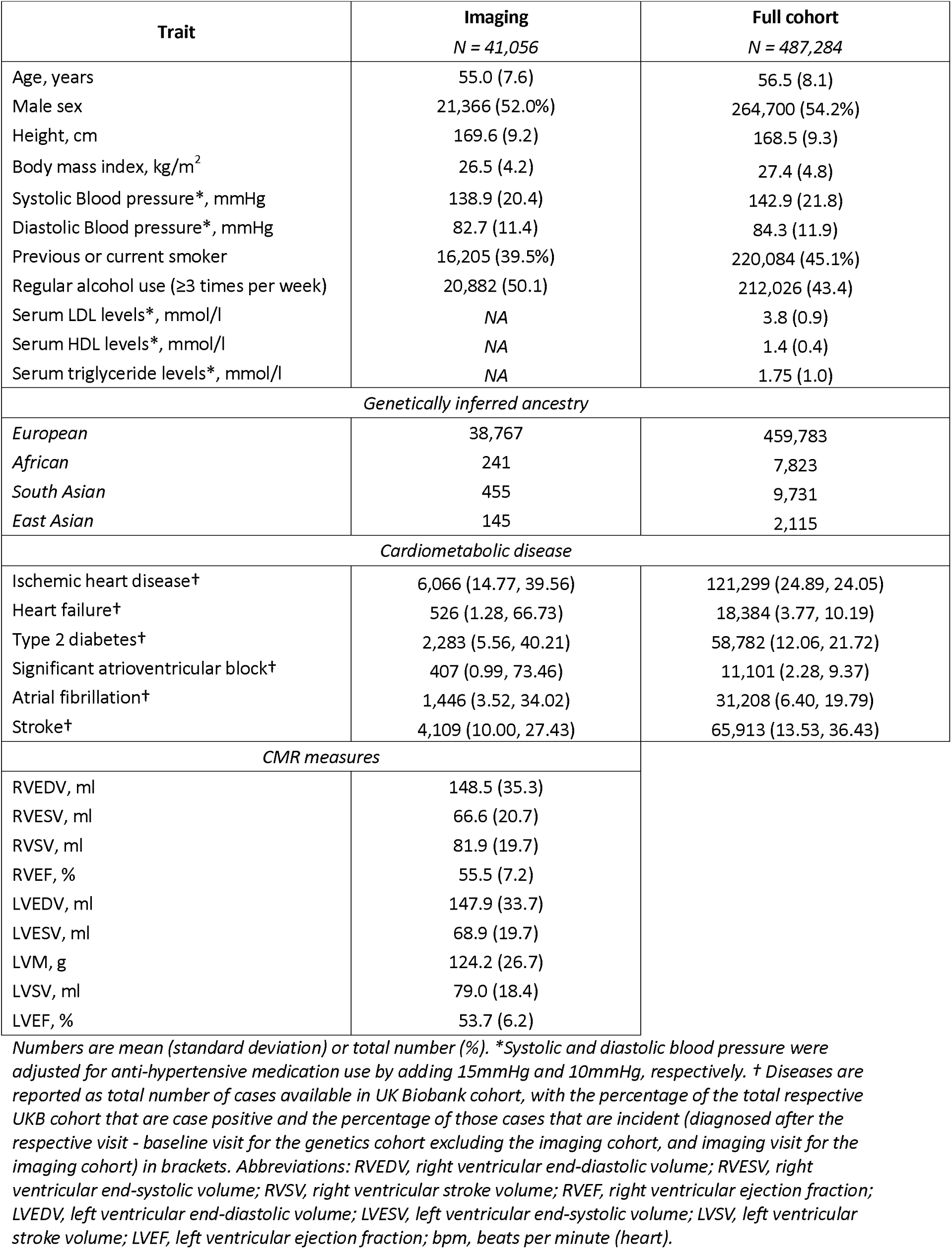
Study demographics of the imaging and full cohorts in UK Biobank.

**Table 2.** Summary of dynamic shape PCs.

| PC | Biological feature | Variation direction (Higher/Lower) |  | Variance explained (%) |
| --- | --- | --- | --- | --- |
|  |  | Higher | Lower |  |
| 1 | Overall heart size | Bigger heart | Smaller heart | 34.07 |
| 2 | Mitral and tricuspid annular plane systolic excursion | Greater degree of MAPSE/TAPSE | Lesser degree of MAPSE/TAPSE | 10.62 |
| 3 | Ventricular sphericalisation | Thinner and longer ventricles | Shorter and wider ventricles in anterior posterior direction | 7.81 |
| 4 | Left ventricular basal position | Lower mitral plane and wider RV in anterior-posterior direction | Higher mitral plane and higher angled tricuspid valve plane | 5.52 |
| 5 | Lateral-septal width | Narrower in the lateral septal direction | Wider in lateral septal direction | 5.46 |
| 6 | Left ventricular length | Longer LV length | Shorter LV length | 4.22 |
| 7 | Septal position | Septal position more shifted to the left | Septal position more shifted towards the right | 4.02 |
| 8 | Tricuspid valve opening size | Smaller tricuspid valve opening | Wider tricuspid valve opening | 2.41 |
| 9 | LV basal systolic excursion | Reduced MAPSE | Greater MAPSE | 2.08 |
| 10 | Right ventricular ejection fraction | Greater RV ejection fraction | Less RV ejection fraction | 2 |
| 11 | Rounder ventricular apex | Rounder apex of the ventricles | More elongated and pointed ventricular apex | 1.6 |
| 12 | Relative left ventricular size | Greater relative size of the LV | Smaller LV relative to the RV | 1.3 |
| 13 | Ejection fraction | Reduced LV and RV ejection fraction | Greater LV and RV ejection fraction | 1.14 |
| 14 | Relative ventricular size and ejection fraction | smaller RV, with reduced LV ejection fraction | Larger RV, greater LV ejection fraction | 1.04 |

Abbreviations: PC, principal component; LV, left ventricle; RV, right ventricle; TAPSE, tricuspid (valve) annular plane systolic excursion.

Since PCA is an unsupervised dimension reduction method, individual PCs are not directly interpretable in terms of traditional measures. However, visual inspection of the PCs revealed that most can be associated with biologically interpretable patterns of cardiac structure and function (Figure 2). PC1 reflected overall heart size and was the dominant source of morphological variation in the population, consistent with the absence of size correction during Procrustes alignment^47^.

**Figure 2.**
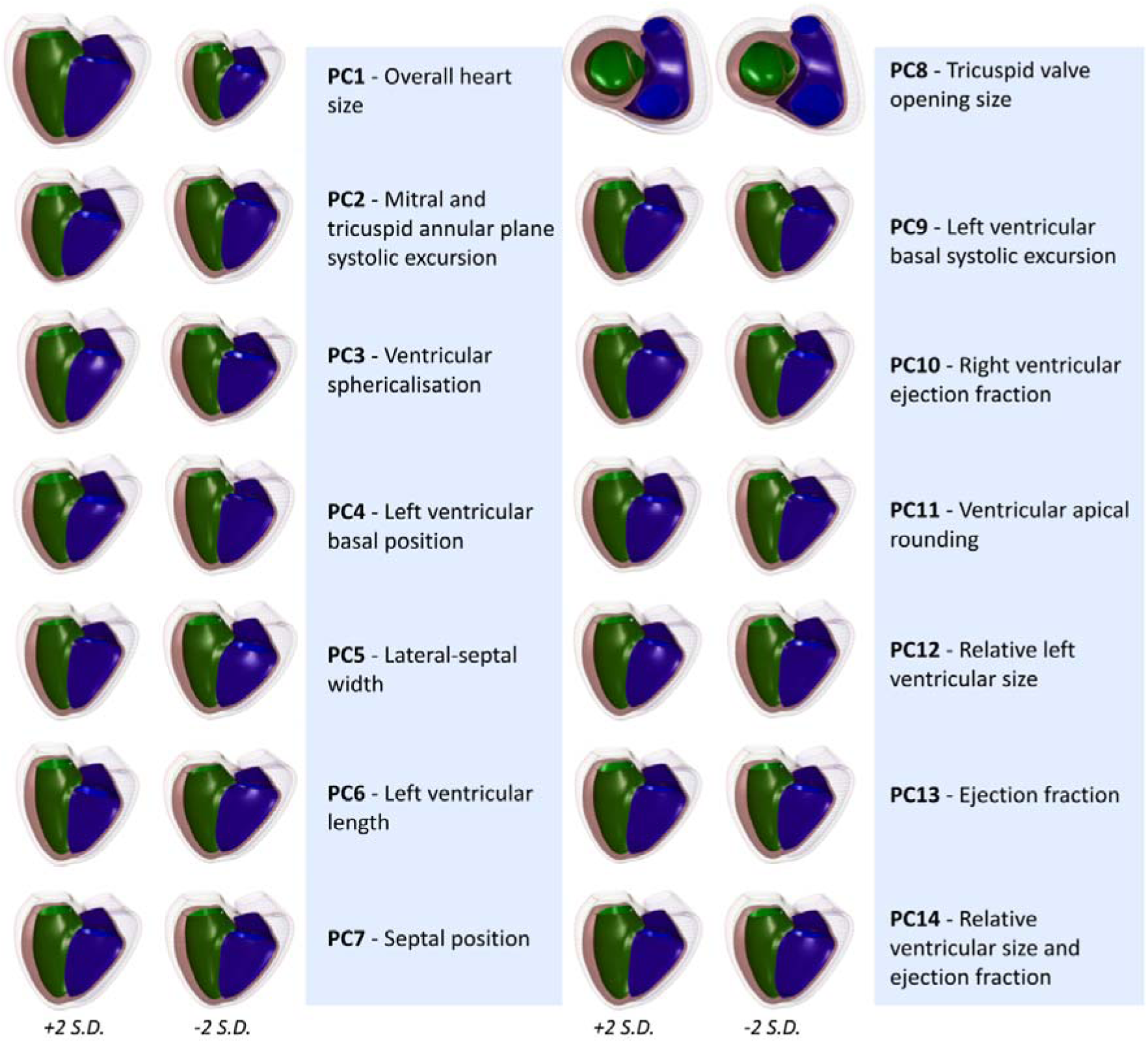
Dynamic shape principal components. Visual summaries of the dynamic shape variation captured by each PC with a text description. For each PC, the shape of the biventricular model at ±2 standard deviations from the mean shape in the direction of that PC is represented. Within each biventricular model represented, green indicates the left ventricular endocardium at end-systole, blue indicates the right ventricular endocardium at end-systole, pink indicates the epicardium at end-systole, and these same structures and end-diastole are indicated in wireframe. PC, principal component; S.D., standard deviation.

Subsequent PCs therefore captured dynamic shape variation independent of global size. We observed several PCs that reflect functional remodeling: PC2 (10.62% variance) was associated with systolic excursion of the mitral and tricuspid annular planes (MAPSE/TAPSE); PC9 (2.08%) was related to LV basal systolic excursion; PC13 (1.14%) represented global biventricular ejection fraction and PC14 (1.04%) reflected variation in relative ventricular size and ejection fraction. These interpretations are summarized in Table 2, Figure 2, and can be further visualized in Supplementary Movies 1-42.

### Relationships with standard CMR measures, cardiovascular risk factors and biventricular ED shape atlas

Correlations between dynamic shape PCs and standard CMR measures demonstrated both shared and complementary information (Supplementary Figure 1). PC1 showed strong correlations with volumetric measures and LV mass (r = 0.88-0.95). In contrast, function-related PCs exhibited only moderate or weak correlations with standard metrics. PC13 (biventricular ejection fraction) correlated moderately with LVEF and RVEF (r = −0.49 and −0.44), while PC2 (MAPSE/TAPSE) showed weak correlations (r = 0.18 and 0.15) with LVEF and RVEF, indicating that dynamic shape features capture aspects of function not fully represented by standard CMR measures

Associations with cardiometabolic risk factors were heterogeneous (Supplementary Figure 2). PC1 was strongly associated with sex and age (r = 0.73 and 0.72), whereas several PCs, such as PC9 (LV basal systolic excursion) showed weaker relationships with classical risk factors (r<0.3), indicating that established epidemiological factors do not explain variation in biventricular shape and motion.

Comparisons with our previously published ED shape atlas^41^ showed high concordance for size-related PCs (PC1, r = 0.98) and moderate correlations for selected geometric features (Supplementary Figure 3). In contrast, key functional PCs (e.g. PC2 and PC13) showed weak correlations with ED-only PCs, confirming that the dynamic atlas captures additional information derived from systolic deformation.

### Associations with cardiometabolic diseases

Dynamic shape PCs were significantly associated with all six prevalent cardiometabolic diseases (Supplementary Table 3) after Bonferroni correction (Figure 3). Multiple PCs were associated with prevalent IHD, AF, and HF, often with patterns of risk-increasing and protective effects consistent with clinical expectations. Associations were also observed for AVB and T2D.

**Figure 3.**
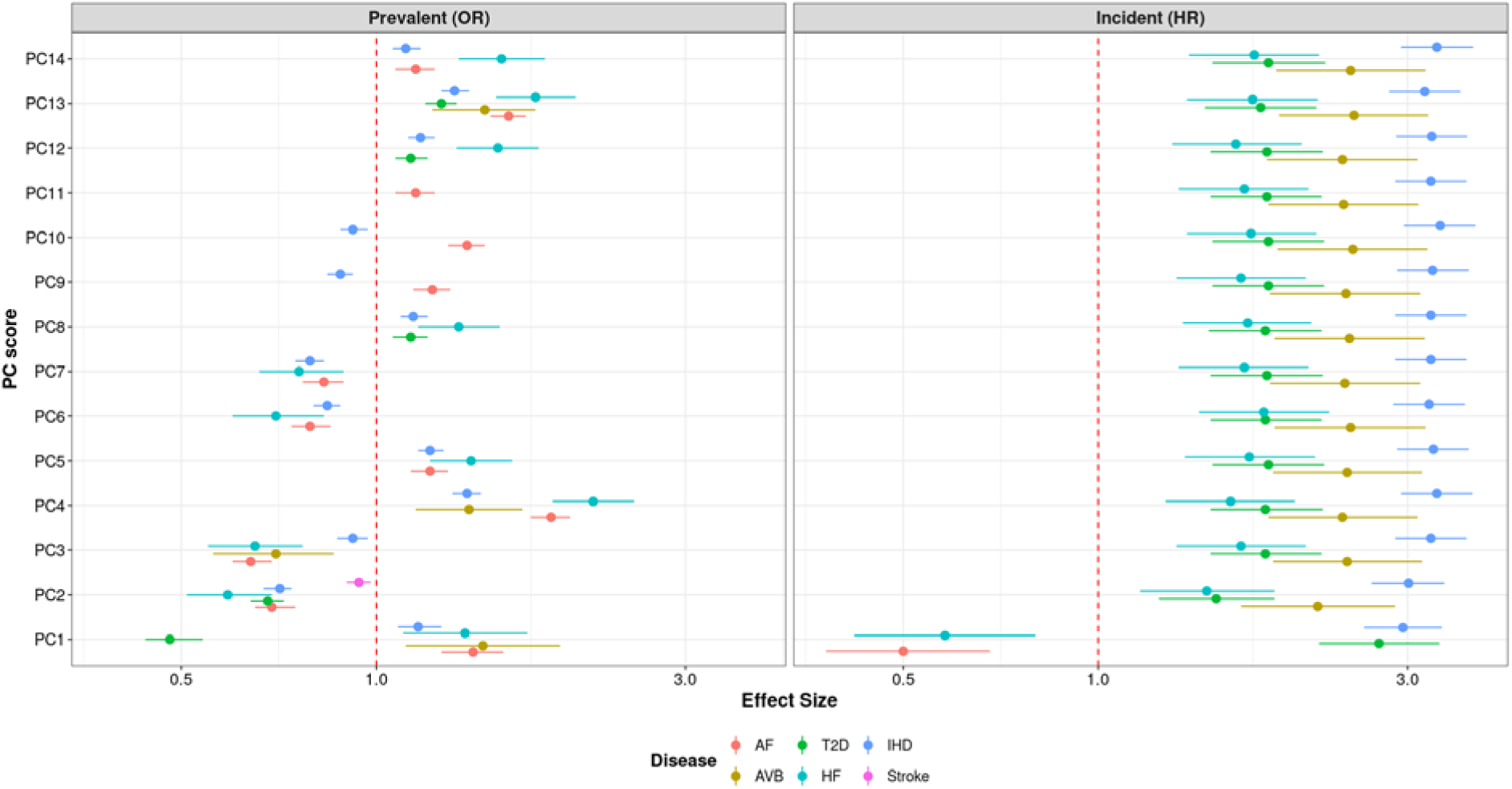
Disease associations between dynamic cardiac shape and cardiometabolic disease. Summary plots of the significant associations between dynamic shape PCs and prevalent and incident cardiometabolic disease. On the X-axis is the relative effect of change in the PC, represented as odds-ratio for prevalent disease and hazard ratio for incident disease. Each association between PC and disease is color coded, with the effect represented as a dot and the 95% confidence intervals as a line. Only disease associations which are significant (Bonferroni corrected P value < 0.05/6) are shown. The dashed red line on the X-axis represents no effect. Abbreviations: OR, odds ratio; HR, hazard ratio; AF, atrial fibrillation; AVB, significant 2^nd^ or 3^rd^ degree atrioventricular block; HF, heart failure; T2D, type 2 diabetes mellitus; IHD, ischemic heart disease; PC, principal component; PRS, polygenic risk score.

For incident outcomes, five diseases showed significant associations, excluding stroke (Figure 3, Supplementary Table 4). Interestingly, 13 PCs were associated with incident AVB, all in a risk-increasing direction. Several PCs showed discordant directions of association between prevalent and incident disease, consistent with a distinction between disease-related and shape-mediated disease susceptibility.

Functionally interpretable PCs showed particularly strong effects. Increased systolic annular excursion (PC2) was protective for prevalent IHD (OR 0.71, CI 0.67-0.74) but strongly associated with increased risk of incident IHD (HR 3.01, CI 2.64-3.42). Similarly, reduced ejection fraction captured by PC13 and PC14 was associated with markedly increased risk of incident HF and IHD (Figure 3).

### Modeling incident cardiometabolic disease risk prediction

Inclusion of dynamic shape information from PC2 (valve annular plane systolic excursion) improved prediction accuracy of time to event for incident IHD beyond standard CMR and clinical measures alone (C-index 0.67 [CI 0.66-0.68], 0.63 [CI 0.62-0.64] respectively). The combined model includes PC2 alongside sex, age, SBP, HDL levels, LVESV, LVM_systolic_ and global longitudinal strain measured in both the 3 and 4 chamber views. The accuracy for predicting time to incident diabetes also improved when including PC2 in a model alongside BMI and triglyceride levels (C-index 0.65 [CI 0.63-0.67]), where the CMR model selected BMI, triglyceride levels, HDL levels and right ventricular septal global circumferential strain (C-index 0.59 [CI 0.56-0.61]). Cox regression models for predicting time to incident AF, stroke and AVB failed to meaningfully predict time to event with either model. The model for HF did not include dynamic shape information as a predictor in the final model. Variables selected for all models are reported in Supplementary Table 5. Results for all models are reported in Supplementary Table 6.

### Genetic architecture of dynamic cardiac shape

All 14 PCs were heritable (h^2^ = 12.3-36.9%, Supplementary Table 7). GWAS identified 75 genome-wide significant loci across the PCs (Table 3, Figure 4), with minimal evidence of confounding (λ = 1.05 - 1.15). Manhattan, locus zoom, and QQ plots are available in Supplementary Figures. Genetic correlations among PCs were modest to moderate (Supplementary Table 8), reflecting partially shared but distinct genetic influences.

**Figure 4.**
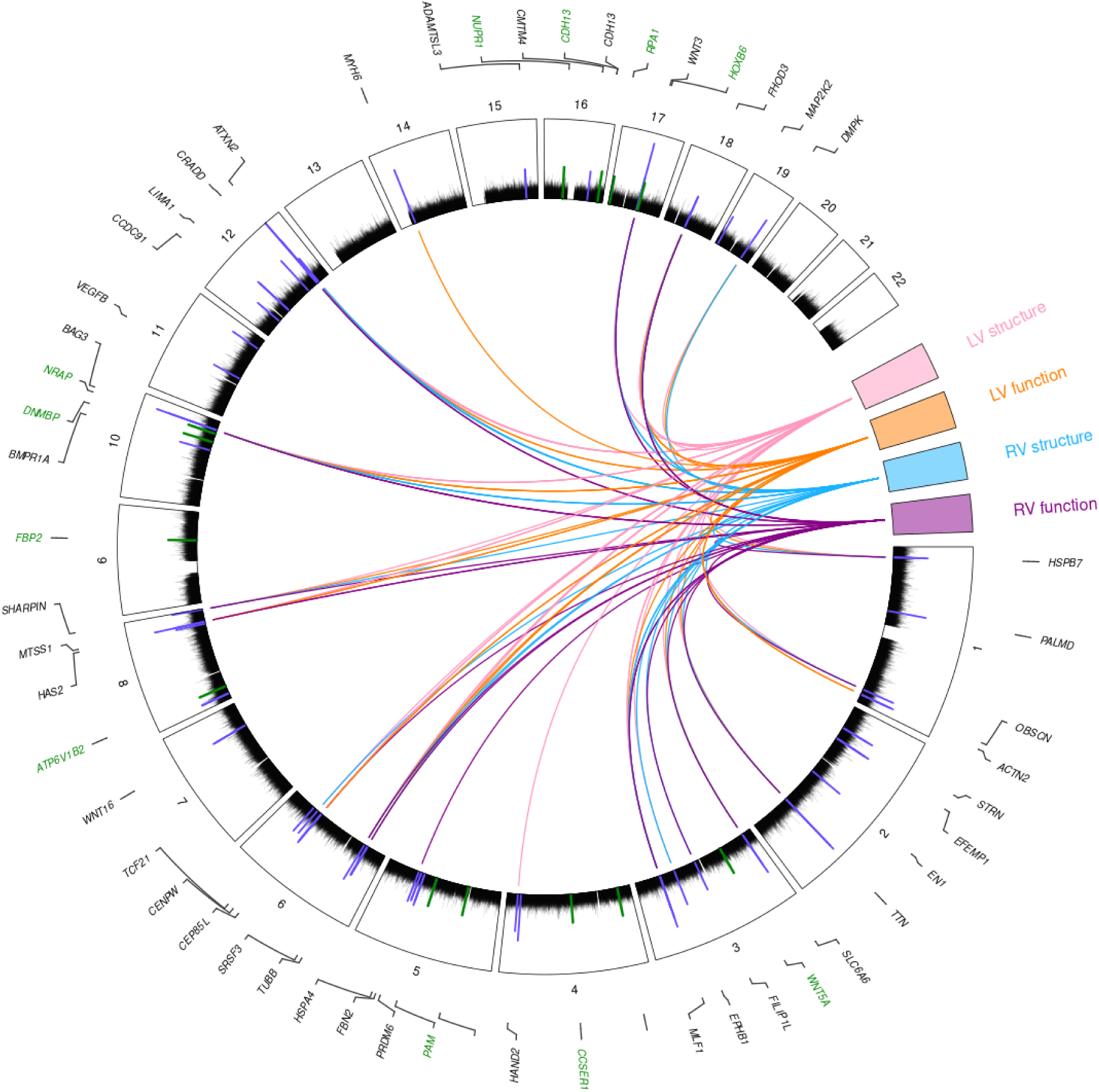
Circos plot of GWAS results and associations with CMR measures. A circos plot summarizing the genetic basis of dynamic cardiac shape. Visualized is a circular Manhattan plot with genome-wide association study summary statistics for each of the 14 dynamic shape principal components overlaid. Chromosomes are divided into segments numbered on the outside of the Manhattan plot; genomic position is along the X-axis and Log10 P-Value on the Y-axis. The genome-wide significant variants (P ≤ 5×10^-8^) which have been previously reported for CMR ventricular structural and functional traits are highlighted in blue, and genome-wide significant variants which have not been previously identified for any cardiovascular-related trait (n=14) are highlighted in green. For each genome-wide significant variant, the candidate gene annotated by the PoPS tool is indicated on the outside of the plot with a line pointing to the variant position. The plot also illustrates genome-wide significant variants which have been identified previously for CMR structural and functional measures; divided into left and right ventricular structural and functional CMR measures. These have segments in the plot alongside the chromosomes, color coded with lines leading to the significant variants that have been associated with the trait. LV, left ventricular; RV, right ventricular.

**Table 3.** Lead genetic variants and secondary signals from genome-wide association study.

| PC | Nearest gene | Lead variant | CHR | Position (Hg19) | EA | NEA | MAF | BETA | SE | P value |
| --- | --- | --- | --- | --- | --- | --- | --- | --- | --- | --- |
| 1 | ATXN2 | rs653178 | 12 | 112007756 | C | T | 0.481 | -0.037 | 0.004 | 2.70E-21 |
| 1 | TTN | rs2042995 | 2 | 179558366 | T | C | 0.223 | 0.042 | 0.005 | 1.30E-18 |
| 1 | BAG3 | rs72840788 | 10 | 121415685 | G | A | 0.218 | 0.041 | 0.005 | 3.40E-17 |
| 1 | RSPH6A | rs7257103 | 19 | 46306904 | C | T | 0.344 | -0.03 | 0.004 | 1.30E-12 |
| 1 | LSM3 | rs73028849 | 3 | 14272766 | G | C | 0.34 | 0.026 | 0.004 | 2.90E-10 |
| 1 | EPHB1 | rs77709005 | 3 | 134444297 | C | T | 0.148 | 0.035 | 0.006 | 4.00E-10 |
| 1 | MLF1 | rs9810339 | 3 | 158302517 | T | C | 0.496 | -0.025 | 0.004 | 4.30E-10 |
| 1 | GOSR2 | rs533030436 | 17 | 45091770 | A | G | 0.12 | -0.038 | 0.006 | 2.20E-09 |
| 1 | OBSCN | rs78529941 | 1 | 228551488 | G | A | 0.38 | -0.024 | 0.004 | 3.00E-09 |
| 1 | HLA-DRB5 | rs6939900 | 6 | 31228357 | C | T | 0.357 | 0.025 | 0.004 | 3.00E-09 |
| 1 | FHOD3 | rs2644262 | 18 | 34223566 | T | C | 0.271 | 0.026 | 0.004 | 3.80E-09 |
| 1 | LINC00964 | rs34123659 | 8 | 125862029 | T | TA | 0.186 | 0.03 | 0.005 | 4.90E-09 |
| 1 | CLCNKA | rs28579893 | 1 | 16347534 | A | G | 0.327 | -0.024 | 0.004 | 5.20E-09 |
| 1 | HSPA4 | 5:132409975_CT_C | 5 | 132409975 | CT | C | 0.267 | 0.027 | 0.005 | 5.90E-09 |
| 1 | PLEC | rs11786896 | 8 | 145018354 | C | T | 0.051 | 0.052 | 0.009 | 1.10E-08 |
| 1 | PLCB3 | rs71468663 | 11 | 64018104 | A | AC | 0.047 | 0.052 | 0.009 | 2.20E-08 |
| 1 | CENPW | rs373980643 | 6 | 126699611 | GT | G | 0.457 | -0.022 | 0.004 | 2.50E-08 |
| 2 | EFEMP1 | rs3791679 | 2 | 56096892 | A | G | 0.228 | -0.046 | 0.008 | 1.20E-09 |
| 2 | PTPN11 | rs11066320 | 12 | 112906415 | A | G | 0.423 | -0.037 | 0.006 | 7.40E-09 |
| 3 | PALMD | rs1890753 | 1 | 100039519 | G | C | 0.478 | 0.043 | 0.007 | 1.00E-10 |
| 3 | EN1 | rs332659 | 2 | 119456238 | A | C | 0.326 | 0.044 | 0.007 | 5.30E-10 |
| 3 | PALLD | rs10155248 | 4 | 169666162 | T | A | 0.474 | -0.048 | 0.007 | 9.40E-13 |
| 3 | PRDM6 | rs10077410 | 5 | 122450771 | G | A | 0.136 | 0.054 | 0.01 | 2.30E-08 |
| 3 | CDKN1A | rs3176326 | 6 | 36647289 | G | A | 0.199 | -0.055 | 0.008 | 8.30E-11 |
| 3 | DNMBP* | rs11190360 | 10 | 101760736 | T | C | 0.058 | -0.082 | 0.014 | 1.30E-08 |
| 3 | NRAP* | rs10749139 | 10 | 115415214 | T | C | 0.137 | -0.055 | 0.01 | 3.80E-08 |
| 3 | DIP2B | rs112716157 | 12 | 50907656 | C | CA | 0.414 | 0.051 | 0.007 | 1.30E-11 |
| 3 | CRADD | rs4144499 | 12 | 94151844 | C | T | 0.496 | -0.044 | 0.007 | 3.20E-11 |
| 3 | GOSR2 | rs17608766 | 17 | 45013271 | T | C | 0.145 | 0.084 | 0.009 | 1.30E-18 |
| 3 | MIR196A1* | rs71366902 | 17 | 46719157 | A | AT | 0.206 | 0.045 | 0.008 | 4.20E-08 |
| 4 | PRDM6 | rs186749 | 5 | 122454305 | A | G | 0.356 | 0.039 | 0.007 | 2.00E-08 |
| 5 | WNT5A* | rs12494109 | 3 | 55491571 | C | A | 0.456 | 0.038 | 0.007 | 4.70E-08 |
| 5 | HAND2-AS1 | rs12502027 | 4 | 174600223 | A | G | 0.362 | -0.046 | 0.007 | 2.40E-10 |
| 5 | PRDM6 | rs398028 | 5 | 122454550 | C | T | 0.203 | -0.052 | 0.009 | 2.20E-09 |
| 5 | HLA-DRB5 | rs2284174 | 6 | 30713580 | T | C | 0.224 | -0.05 | 0.008 | 2.90E-09 |
| 5 | HLA-DRB5 | 6:31588020_CCTCT_C | 6 | 31588020 | CCTCT | C | 0.362 | 0.043 | 0.007 | 3.30E-09 |
| 5 | CCDC91 | rs2638950 | 12 | 28551567 | A | G | 0.244 | 0.044 | 0.008 | 4.30E-08 |
| 5 | ADAMTSL3 | rs6602994 | 15 | 84490757 | T | C | 0.279 | 0.043 | 0.008 | 2.20E-08 |
| 6 | EFEMP1 | rs201699038 | 2 | 56075676 | AT | A | 0.156 | 0.06 | 0.009 | 1.30E-10 |
| 6 | RSRC1 | rs11710570 | 3 | 158056654 | T | C | 0.448 | 0.052 | 0.007 | 1.00E-14 |
| 6 | CEP85L | rs11756440 | 6 | 118993642 | C | A | 0.473 | 0.041 | 0.007 | 8.00E-10 |
| 6 | TCF21 | rs162185 | 6 | 134226147 | T | C | 0.409 | -0.039 | 0.007 | 9.80E-09 |
| 6 | *FAM3C* | rs12531355 | 7 | 121027434 | C | T | 0.271 | 0.048 | 0.008 | 2.00E-10 |
| 6 | *HAS2-AS1* | rs62526480 | 8 | 122681022 | T | C | 0.406 | -0.038 | 0.007 | 2.00E-08 |
| 6 | *DDX6\** | rs12223220 | 11 | 118587888 | A | G | 0.175 | -0.053 | 0.009 | 1.10E-08 |
| 6 | *ZBTB7A* | rs66534382 | 19 | 4056366 | G | A | 0.19 | 0.049 | 0.009 | 9.70E-09 |
| 7 | *HEATR1* | rs531997 | 1 | 236803402 | A | G | 0.179 | -0.053 | 0.009 | 3.00E-09 |
| 7 | *PPIP5K2\** | rs32841 | 5 | 102478258 | A | G | 0.347 | 0.041 | 0.007 | 2.40E-08 |
| 7 | *TBX3* | rs10850409 | 12 | 115381740 | G | A | 0.268 | 0.044 | 0.008 | 1.30E-08 |
| 8 | *SGF29\** | rs151091024 | 16 | 28569054 | A | G | 0.026 | -0.13 | 0.022 | 6.40E-09 |
| 8 | *DPH1\** | rs12939170 | 17 | 1937526 | C | T | 0.028 | -0.115 | 0.021 | 4.60E-08 |
| 9 | *SLIT2\** | rs6821503 | 4 | 19609401 | C | T | 0.263 | 0.042 | 0.008 | 3.90E-08 |
| 9 | *LZTS1-AS1\** | rs7014186 | 8 | 20318812 | A | C | 0.115 | -0.057 | 0.01 | 4.30E-08 |
| 9 | *MFSD14B\** | 9:97209762_TG_T | 9 | 97209762 | TG | T | 0.108 | -0.076 | 0.014 | 4.40E-08 |
| 10 | *MYH6* | rs422068 | 14 | 23864804 | T | C | 0.356 | -0.057 | 0.007 | 1.50E-15 |
| 10 | *FHOD3* | 18:34219777_G_GTT | 18 | 34219777 | G | GTT | 0.248 | 0.048 | 0.008 | 1.80E-09 |
| 10 | *BAG3* | rs17617337 | 10 | 121426884 | C | T | 0.218 | 0.047 | 0.008 | 9.70E-09 |
| 10 | *CMTM4* | 16:66690988_AT_A | 16 | 66690988 | AT | A | 0.489 | -0.039 | 0.007 | 2.30E-08 |
| 11 | *CDKN1A* | rs3176326 | 6 | 36647289 | G | A | 0.199 | 0.058 | 0.009 | 4.20E-11 |
| 11 | *STRN* | rs138934670 | 2 | 37136145 | A | AAAC | 0.356 | 0.048 | 0.008 | 1.50E-09 |
| 11 | *CDH13\** | rs9932314 | 16 | 82881172 | C | T | 0.498 | 0.04 | 0.007 | 1.00E-08 |
| 11 | *ITGA1\** | rs60757101 | 5 | 51603171 | T | A | 0.168 | -0.056 | 0.01 | 2.50E-08 |
| 11 | *CCSER1\** | rs77504602 | 4 | 92260138 | C | A | 0.288 | -0.042 | 0.008 | 4.40E-08 |
| 12 | *FBN2* | rs13185878 | 5 | 127962298 | A | G | 0.273 | -0.043 | 0.008 | 1.90E-08 |
| 12 | *HEATR5B* | rs1105014 | 2 | 37265727 | G | C | 0.479 | -0.038 | 0.007 | 2.10E-08 |
| 13 | *BAG3* | rs2234962 | 10 | 121429633 | T | C | 0.218 | 0.061 | 0.008 | 4.60E-14 |
| 13 | *LINC00964* | rs34866937 | 8 | 125859850 | G | A | 0.314 | 0.054 | 0.007 | 5.90E-14 |
| 13 | *LSM3* | rs56242556 | 3 | 14273472 | C | T | 0.215 | -0.058 | 0.008 | 1.30E-12 |
| 13 | *HSPB7\** | rs1048238 | 1 | 16341649 | C | T | 0.428 | -0.041 | 0.007 | 1.30E-09 |
| 13 | *LDB3* | rs35605324 | 10 | 88443703 | G | A | 0.081 | 0.071 | 0.012 | 6.60E-09 |
| 13 | *FAM86B3P* | rs1915986 | 8 | 8114543 | A | G | 0.499 | 0.038 | 0.007 | 2.10E-08 |
| 14 | *SLC35F1* | rs17079881 | 6 | 118566187 | A | G | 0.134 | 0.063 | 0.01 | 2.90E-10 |
| 14 | *ACTN2* | rs12724121 | 1 | 236852282 | A | T | 0.371 | -0.044 | 0.007 | 6.70E-10 |
| 14 | *CMSS1* | rs12488245 | 3 | 99835254 | T | C | 0.416 | -0.04 | 0.007 | 1.00E-08 |
| 14 | *FHOD3* | 18:34219777_G_GTT | 18 | 34219777 | G | GTT | 0.248 | 0.044 | 0.008 | 4.00E-08 |
| Conditionally independent secondary signals |  |  |  |  |  |  |  |  |  |  |
| 1 | *SLC6A6* | rs34234056 | 3 | 14418444 | A | G | 0.494 | 0.021 | 0.004 | 1.10E-07 |
| 11 | *CDH13* | rs4366697 | 16 | 83067983 | G | C | 0.322 | 0.038 | 0.007 | 2.80E-07 |

Abbreviations: PC, Principal component; CHR, chromosome; Hg19, Genome Reference Consortium Human Build 37 (GRCh37); EA, effect allele; NEA, non-effect allele; Beta, effect size; SE, standard error; p, probability value.
\* indicates variant not previously reported for a cardiac trait.

Comparison with ED shape GWAS demonstrated substantial overlap for size and geometry-related PCs, but limited genetic correlation for functional PCs such as PC2 (MAPSE/TAPSE) and PC13 (ejection fraction (Supplementary Table 9). Of the 77 lead signals (two independent signals at discovered loci), 20 were genome-wide significant in the ED shape GWAS, and 13 demonstrated suggestive significance (5×10^-8^ < P < 1×10^-6^). Thirty lead signals were 1×10^-6^ < P < 1×10^-3^ and 14 had nominal significance 1×10^-3^ < P < 0.05 (Supplementary Table 10). Moreover, of the 77 lead signals, 14 of which had not previously been implicated in cardiac traits, and 8 were entirely novel across published GWAS at time of analysis (Supplementary Table 11, Supplementary Table 12).

Functional annotation identified two non-synonymous missense variants, one was predicted to be possibly damaging with PolyPhen (rs2234962, chr10:121429633:T>C, BAG3, PCs 1, 10 and 13) and one was predicted to be probably damaging (rs26821, chr5:102520400:A>G, PPIP5K2, PC7) (Supplementary Table 13). Twenty-two variants were identified as eQTLs in aortic artery, coronary artery, left ventricle and atrial appendage tissues^71^, of which twenty had support for colocalization (PP4 > 0.75)^58^ (Supplementary Table 14). Transcriptome-wide analysis identified 103 genes (P < 3.1 x 10^-6^, Bonferroni corrected) mapping to the boundaries of 40 loci (Supplementary Table 15). Finally, 280 Hi-C interactions were identified at 45 of the lead GWAS variants, summarized in Supplementary Table 16. Integrative gene prioritization including those genes identified by the aforementioned tools alongside results from PoPs, identified 77 candidate genes (Supplementary Table 17), enriched for pathways related to cardiac development (P_adj_ = 2.79×10^-5^), muscle structure (P_adj_ = 3.53×10^-4^), contractile function (P_adj_ = 4.45×10^-4^, Supplementary Table 18).

Rare variant burden testing implicated established cardiomyopathy genes (e.g. TTN for PC1, FHOD3 for PC10), providing convergent evidence with common variant findings (Supplementary Table 19). Five additional genes not identified in the common-variant GWAS showed evidence of rare variants driving associations (P < 2.79×10^-6^); RBM20 and VPS25 for PC1 (overall heart size), SMARCAD1 for PC3 (ventricular sphericity), NKAPD1 for PC7 (septal position), CSRP3 for PC14 (relative ventricular size and ejection fraction). There was significant evidence that multiple rare missense variants were driving the associations at TTN and SMARCAD1 (Supplementary Table 19).

### Polygenic risk and causal inference

PRS derived from dynamic shape PCs were associated with a broad range of cardiovascular phenotypes in PheWAS, predominantly within the circulatory system (Supplementary Table 20). PRS for reduced biventricular ejection fraction (PC13) showed strong associations with HF phecodes (OR 1.06, 1.07, P = 2.74×10^-8^ – 6.19×10^-7^), while PRS for reduced annular excursion (PC2) was associated with ischemic and hypertensive phenotypes, despite comprising only two variants (Supplementary Table 21).

We also explored relationships between dynamic cardiac shape PC PRS with prevalent and incident cardiometabolic diseases (Figure 5). There were seven significant associations between PC PRS and prevalent disease (P < 0.0083). Four were associated with prevalent IHD; PC12 (relative left ventricular size) in a risk conferring direction (OR 2.46, CI 1.60-3.78) and PCs 2 (MAPSE/TAPSE), 5 (lateral-septal width, OR 0.45, CI 0.29-0.71) and 6 (left ventricular length, OR 0.75, CI 0.62-0.92) as protective, two with prevalent AF; PCs 6 (LV length) and 10 (right ventricular ejection fraction) (OR 1.73 [CI 1.28-2.37]; OR 1.92 [CI 1.26-2.93]), and one with HF; PC8 (tricuspid valve opening size) (OR 4.71, CI 1.53-14.48) (Supplementary Table 22).

**Figure 5.**
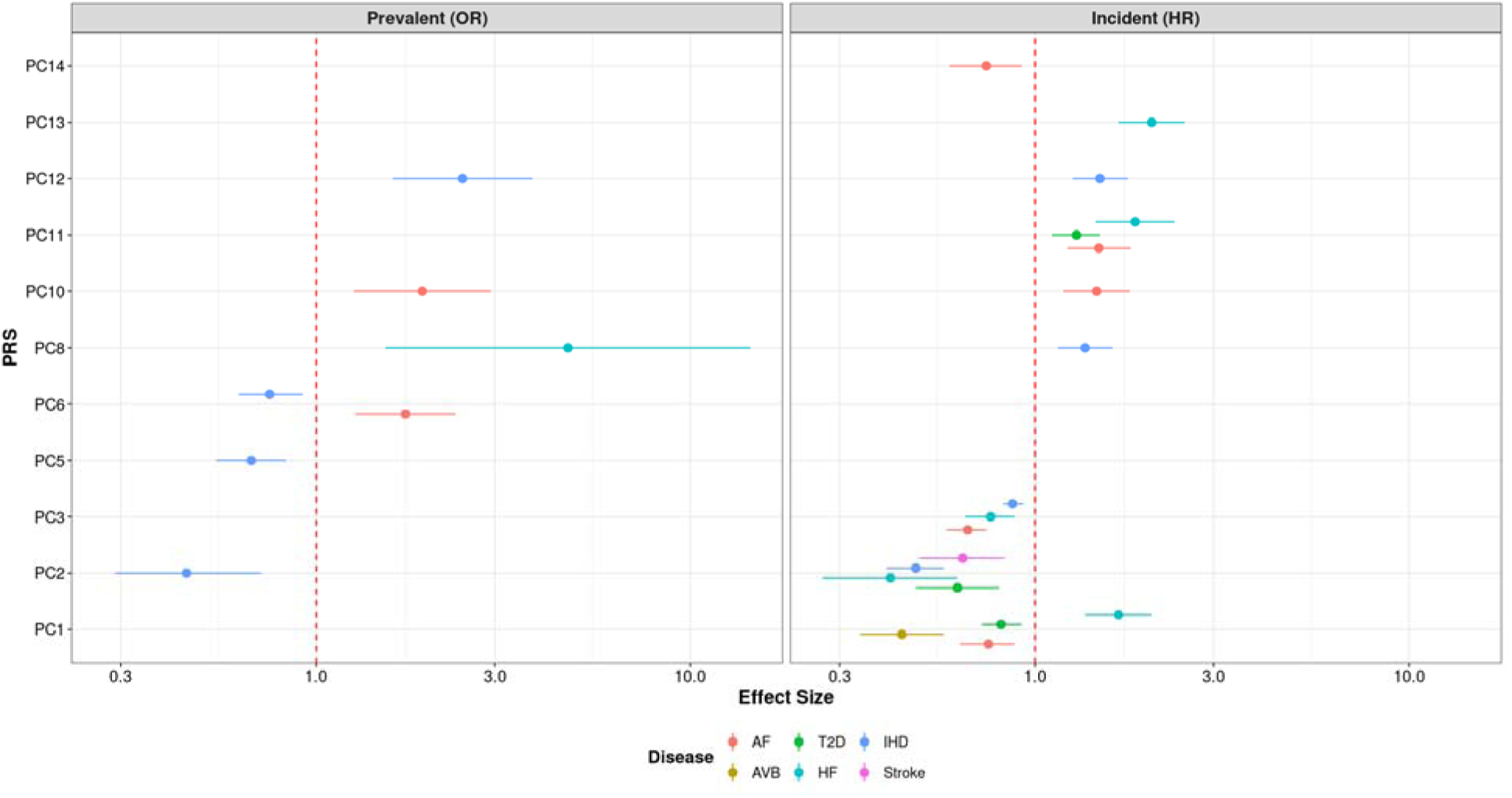
Disease associations between dynamic cardiac shape polygenic risk scores and cardiometabolic disease. Summary plots of the significant associations between dynamic shape PC PRS and prevalent and incident cardiometabolic disease. On the X-axis is the relative effect of change in the PC PRS, represented as odds-ratio for prevalent disease and hazard ratio for incident disease. Each association between PC PRS and disease is color coded, with the effect represented as a dot and the 95% confidence intervals as a line. Only disease associations which are significant (Bonferroni corrected P value < 0.05/6) are shown. The dashed red line on the X-axis represents no effect. Abbreviations: OR, odds ratio; HR, hazard ratio; AF, atrial fibrillation; AVB, significant 2^nd^ or 3^rd^ degree atrioventricular block; HF, heart failure; T2D, type 2 diabetes mellitus; IHD, ischemic heart disease; PC, principal component; PRS, polygenic risk score.

Reviewing results from testing of PC PRS with incident disease highlighted that the strongest associations were with incident HF. There were five PC PRS significantly associated with HF: PCs 1 (heart size, OR 1.67, CI 1.36-2.05), 11 (rounder ventricular apex, OR 1.85, CI 1.45-2.36) and 13 (global biventricular ejection fraction, OR 2.05, CI 1.67-2.51) in a risk-conferring direction and PCs 2 (MAPSE/TAPSE, OR 0.41, CI 0.27-0.62) and 3 (ventricular sphericalization, OR 0.76, CI 0.65-0.88) as protective. Four PRS were significantly associated with incident AF; PCs 10 (right ventricular ejection fraction, OR 1.46, CI 1.19-1.79) and 11 (OR 1.48, CI 1.22-1.80) in a risk-conferring direction and PCs 1 (heart size, OR 0.75, CI 0.63-0.88) and 3 (OR 0.66, CI 0.58-0.75) as protective. A full summary of the significant PRS associations can be found in Supplementary Table 23.

MR supported a robust causal effect of reduced biventricular ejection fraction (PC13) on HF risk, with consistent estimates across multiple methods (including Inverse variance weighted β 0.492, P 6×10^-7^ and Weighted median β 0.408, P 4.2×10^-5^) and no evidence of horizontal pleiotropy. Additional significant causal relationships were observed with disease as the exposure and dynamic cardiac shape as the outcome (reporting inverse variance weighted results, full results in Table 4); between T2D and PCs 1 (overall heart size) (β −0.05, P 0.003) and 12 (relative LV size) (β 0.04, P 0.008); between IHD and PC9 (LV basal systolic excursion) (β −0.06, P 0.008), and between HF and PC14 (relative ventricular size and ejection fraction) (β 0.1, P 0.004) (sensitivity analyses in Supplementary Table 24). Full MR results are presented in Supplementary Tables 25-27.

**Table 4.** Significant Mendelian Randomization results.

| Exposure | Outcome | Method | n<br>SNV | Beta | SE | P value | MR Egger<br>Het P | Inverse<br>variance<br>weighted<br>Het P | Pleiotropy<br>P |
| --- | --- | --- | --- | --- | --- | --- | --- | --- | --- |
| PC13 | HF | Inverse variance weighted | 5 | 0.49 | 0.10 | 6.00E-07 | 0.04 | 0.08 | 0.86 |
| PC13 | HF | Weighted median | 5 | 0.41 | 0.10 | 4.18E-05 | 0.04 | 0.08 | 0.86 |
| T2D | PC12 | Inverse variance weighted | 138 | 0.04 | 0.01 | 0.008 | 0.01 | 0.01 | 0.28 |
| T2D | PC12 | Weighted median | 138 | 0.07 | 0.02 | 3.64E-04 | 0.01 | 0.01 | 0.28 |
| T2D | PC12 | Weighted mode | 138 | 0.09 | 0.03 | 0.001 | 0.01 | 0.01 | 0.28 |
| T2D | PC1 | Weighted mode | 154 | -0.05 | 0.02 | 0.003 | <1E-7 | <1E-7 | 0.14 |
| HF | PC14 | Inverse variance weighted | 37 | 0.10 | 0.04 | 0.004 | 0.05 | 0.05 | 0.36 |
| IHD | PC9 | Inverse variance weighted | 175 | -0.06 | 0.02 | 0.008 | 0.20 | 0.21 | 0.75 |

Abbreviations: PC, principal component; T2D, type 2 diabetes mellitus; HF, heart failure; IHD, ischemic heart disease; SNV, single nucleotide variants included in analysis; SE, standard error.

## Discussion

Quantifying dynamic cardiac shape through statistical shape atlasing provides a complementary framework for studying cardiac disease-morphology relationships. By jointly modeling ventricular geometry at ED and ES, dynamic shape phenotypes capture not only static morphology but also functional deformation during contraction. Previously, we showed that these static shape phenotypes have substantially stronger associations with prevalent cardiometabolic diseases than conventional CMR measures, including ejection fraction, strain, and valve excursions^42^. Here, we identified dynamic shape components that are significantly associated with prevalent and incident cardiometabolic disease, influenced by expression of novel genetic loci, PRS of which are also predictive of incident HF and AF, and which are causal for HF by Mendelian randomization. We also improve prediction of incident IHD when adding PC13 (ejection fraction) to standard CMR measures and covariates versus CMR measures and covariates alone.

The dynamic shape atlas builds directly on previous work based on ED shape alone. As expected, significant correlations were observed between ED and dynamic shape PCs, particularly for components capturing global cardiac size. However, beyond the first PC, many dynamic shape PCs showed only modest phenotypic and genetic correlation with ED PCs, indicating that the inclusion of ES information introduces genuinely new variation related to cardiac function. This is further supported by the observation that all dynamic shape PCs demonstrated some degree of genetic signal in the ED GWAS, while the dynamic atlas substantially increased discovery power. Several dynamic shape PCs captured functional features closely aligned with established clinical metrics. PC2 reflected longitudinal annular motion analogous to MAPSE and TAPSE, while PC13 captured variation resembling biventricular ejection fraction. Despite these similarities, correlations with standard measures were incomplete, indicating that dynamic shape phenotypes encode complementary information about cardiac mechanics that is not fully captured by standard CMR metrics.

Strong associations were observed between both prevalent and incident IHD and several dynamic cardiac shape PCs, particularly those capturing functional variation (PCs 2, 9, 13 and 14). Importantly, the direction of these associations consistently reflected impaired cardiac function, including reduced annular plane systolic excursion (PC2) and reduced biventricular ejection fraction (PC13). This association is also seen in the PRS for PC2, however there is disagreement in the direction of effect between the PC and IHD and the PC PRS and IHD which could be attributed to the low percent heritability explained by the two genome-wide significant variants identified for PC2 (0.5%). While IHD is primarily driven by coronary artery pathology, which is not directly captured by ventricular shape atlases, these findings demonstrate that ventricular geometry and systolic deformation encode meaningful information about IHD risk. There is also evidence that PC2 provides complementary information to CMR measures when predicting incident IHD.

Diabetes has long been associated with characteristic patterns of cardiac remodeling, including reduced ventricular volumes, increased myocardial mass, and altered systolic performance^72,73^. Consistent with prior studies, we observed significant associations between multiple dynamic shape PCs and prevalent T2D (PCs 1, 2, 8, 12, 13), extending our previous findings based on ED shape alone^41^. Some of these observations are supported by MR, with a significant causal relationship observed between T2D on PCs 1 (overall heart size) and 12 (relative LV size). Importantly, dynamic shape PCs capturing both size and functional variation were associated with T2D, highlighting the importance of systolic remodeling in metabolic heart disease. Associations with incident T2D were also observed for all dynamic shape PCs, as well as between T2D and the PRS for PCs 2 and 11, reinforcing the close link between cardiac morphology and metabolic dysfunction ^72,74–77^. Together, these findings support the concept that cardiac shape and function reflect a continuum of metabolic stress and insulin resistance^78–82^. Given the difficulty of directly measuring insulin resistance, dynamic cardiac shape phenotypes may serve as informative imaging-derived biomarkers of subclinical metabolic dysfunction.

There were substantial associations observed between cardiac shape PCs and HF in the models using both the PCs and PC PRS. PCs 1-8, 12-14 had significant association with prevalent HF, of which the association between prevalent HF and PC8 (tricuspid valve opening size) is mirrored in the PRS results, suggesting a remodeling effect of HF reflecting in dynamic shape. All PCs were significantly associated with incident HF, relationships that were also observed in PRS for PCs 1, 2, 3, 11 and 13, suggesting that dynamic cardiac shape can predispose future HF events. In testing the causality of these associations in MR, a significant causal effect was observed of HF on PC14 (relative ventricular size and ejection fraction), and of PC13 (ejection fraction) on HF. Importantly, the incomplete correlation between PC13 and imaging-derived ejection fraction suggests that this phenotype captures additional functional information relevant to HF beyond standard measures.

Dynamic cardiac shape PCs demonstrated moderate-to-high heritability and yielded substantially more genome-wide significant loci than previous ED shape analysis. In total, 75 loci were identified, with 14 genetic signals being novel for cardiac traits. Importantly, even for PCs closely related to ED morphology, such as overall heart size, the inclusion of systolic information increased the number of associated loci, highlighting the added genetic signal captured by dynamic phenotyping.

Several novel loci provided candidate genes with established roles in cardiac development, structure, and function, including pathways related to myocardial architecture and contractile mechanisms. In particular, PCs capturing smaller proportions of total variance, reflecting more localized or functional features, had a higher number of associations with novel loci, whereas PCs explaining larger variance overlapped more strongly with known cardiac structure and function traits. This pattern suggests that dynamic shape atlasing enables the discovery of genetic influences on subtler aspects of cardiac mechanisms that are not well represented by traditional imaging phenotypes.

Novel signals for cardiac traits included: rs12494109 identified for PC5 (lateral-septal width), the candidate gene is Wingless-Type MMTV Integration Site Family, Member 5A, this is a gene implicated in developmental processes during embryogenesis, associated with congenital right ventricular outflow tract obstruction in autosomal recessive Robinow syndrome^83,84^, and mouse models exhibit a double outlet right ventricle, atrioventricular septal defect and left ventricular and pulmonary arterial hypoplasia^85^.

Two variants, rs32841 identified for PC7 (septal position) with potential candidate gene Gypsy Retrotransposon Integrase 1 and rs12939170 identified for PC8 (tricuspid valve opening size) with candidate gene Replication Protein A1 had knockout mouse models with abnormal cardiac morphology^86^ – knockout mice with Gypsy Retrotransposon Integrase 1 had abnormal heart shape, and knockout mice with Replication Protein A1 had increased heart weight.

Several additional candidate genes were indicated from gene-burden testing, of these some are known cardiomyopathy genes. One example is Cysteine And Glycine Rich Protein 3 (CRSP3), rare variants in this gene are associated with PC14 (left ventricular size and reduced ejection fraction). This gene encodes a muscle LIM protein which is important for cardiac and skeletal muscle structure and function and is a regulator in sarcomere assembly and contraction^87^. A rare variant was associated in Vacuolar Protein Sorting 25 Homolog (VPS25), VPS25 encodes a protein that functions in sorting of ubiquitinated membrane proteins during endocytosis, has mouse models with decreased heart weight and hemorrhaging, and has been associated with frontotemporal dementia^88^.

## Conclusion

In conclusion, dynamic statistical shape atlasing captures cardiac shape variation across the cardiac cycle together with functional deformation, providing robust and interpretable imaging-derived phenotypes that encapsulate substantially more information from CMR than traditional summary measures. Building on previous ED biventricular atlases, this work demonstrates the added value of incorporating systolic information to define shape components that jointly reflect cardiac morphology and function. These dynamic shape PCs show biologically plausible associations with both prevalent and incident cardiometabolic disease and improve disease prediction when added to standard CMR-derived metrics. In genetic analyses, dynamic shape PCs enhance locus discovery compared with ED shape alone, identifying additional variants, including loci not previously implicated in cardiovascular traits. PRS constructed from these variants were also predictive of cardiometabolic outcomes. Together, these findings establish dynamic cardiac shape atlasing as a powerful and interpretable framework for integrating cardiac structure, function, genetics, and disease risk.

In future work this statistical shape atlasing methodology can be applied to more varied cohorts to explore diseases such as cardiomyopathies which are under-represented in UK Biobank, as well as exploring dynamic shape variation within the population due to factors such as sex, age or ancestry. The capacity for dynamic cardiac shape to predict incident cardiometabolic disease can be further explored with more specific datasets, and leveraging the longitudinal repeat imaging data from UK Biobank to assess the clinical utility of these imaging-derived phenotypes.

## Ethics approval

This study complies with the Declaration of Helsinki; the work was covered by the ethical approval for UK Biobank studies from the NHS National Research Ethics Service on 17th June 2011 (Ref 11/NW/0382) and extended on 18 June 2021 (Ref 21/NW/0157) with written informed consent obtained from all participants.

## Acknowledgements

This research was conducted using the UK Biobank Resource under Application Number 2964. The work uses data provided by patients and collected by the NHS as part of their care and support. This research used data assets made available by National Safe Haven as part of the Data and Connectivity National Core Study, led by Health Data Research UK in partnership with the Office for National Statistics and funded by UK Research and Innovation (grant ref MC_PC_20029). Copyright © (2022), NHS Digital. Re-used with the permission of the NHS Digital [and/or UK Biobank]. All rights reserved. Barts Charity (G-002346) contributed to fees required to access UK Biobank data [access application #2964]. JR acknowledges funding by the Spanish Ministry of Science and Innovation (MCIN/AEI/10.13039/501100011033) for fellowship RYC2021-031413-I. RB, SEP and PBM acknowledge the support of the National Institute for Health and Care Research (NIHR) Barts Biomedical Research Centre (NIHR203330); a delivery partnership of Barts Health NHS Trust, Queen Mary University of London, St George’s University Hospitals NHS Foundation Trust and St George’s University of London. WJY acknowledges the NIHR Integrated Academic Training program, which supports his Academic Clinical Lectureship post. KU acknowledges support from a British Heart Foundation 4-year PhD studentship (FS/4yPhD/F/23/34196).

## Contributions

R.B., J.R., A.Y., and P.B.M. conceived and designed the experiments, R.B., performed the experiments, R.B., and K.U. performed statistical analysis, R.B., J.R., A.Y., and P.B.M. analysed the data, W.J.Y., J.R., A.Y., and P.B.M. contributed reagents/materials/analysis tools, J.R., A.Y., and P.B.M. jointly supervised the research, R.B., J.R., A.Y., and P.B.M. wrote the paper and W.J.Y and S.E.P. provided critical review of the manuscript.

