## Supplementary Figures for "Biventricular cardiac dynamic shape: genetics and cardiometabolic disease associations"

Supplementary Figure 1 – Correlation matrix between dynamic shape PCs and CMR measures

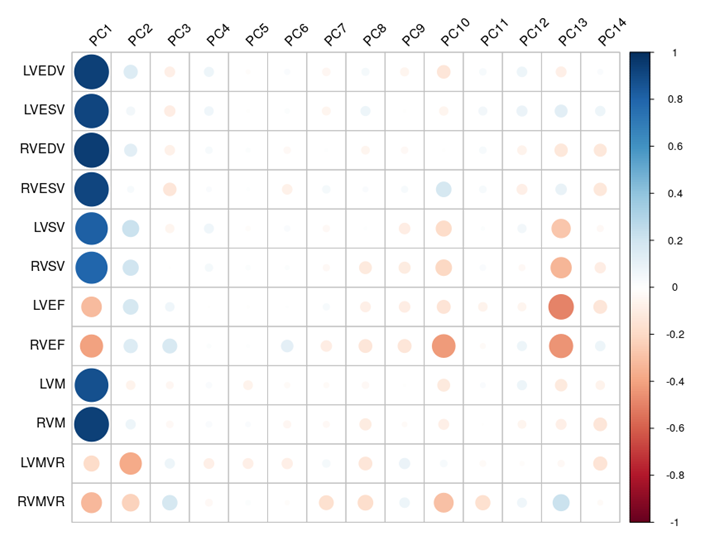
*PC, principal component; CMR, cardiac magnetic resonance (imaging); LVEDV, left ventricular end-diastolic volume; LVESV, left ventricular end-systolic volume; LVSV, left ventricular stroke volume; LVEF, left ventricular ejection fraction; LVM, left ventricular (myocardial) mass; left ventricular mass to volume ratio; RVEDV, right ventricular end-diastolic volume; RVESV, right ventricular end-systolic volume; RVSV, right ventricular stroke volume; RVEF, right ventricular ejection fraction; RVM, right ventricular (myocardial) mass; right ventricular mass to volume ratio.*

Supplementary Figure 2 - Correlation matrix between dynamic shape PCs and cardiometabolic disease risk factors
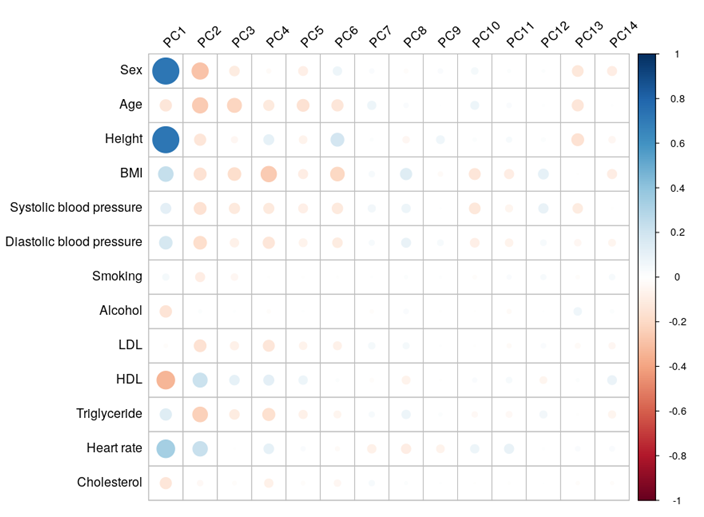
*PC, principal component; BMI, body mass index; LDL, serum low-density lipoprotein; HDL, serum high-density lipoprotein.*

Supplementary Figure 3 – Correlation matrix between dynamic shape PCs and end-diastolic shape PCs

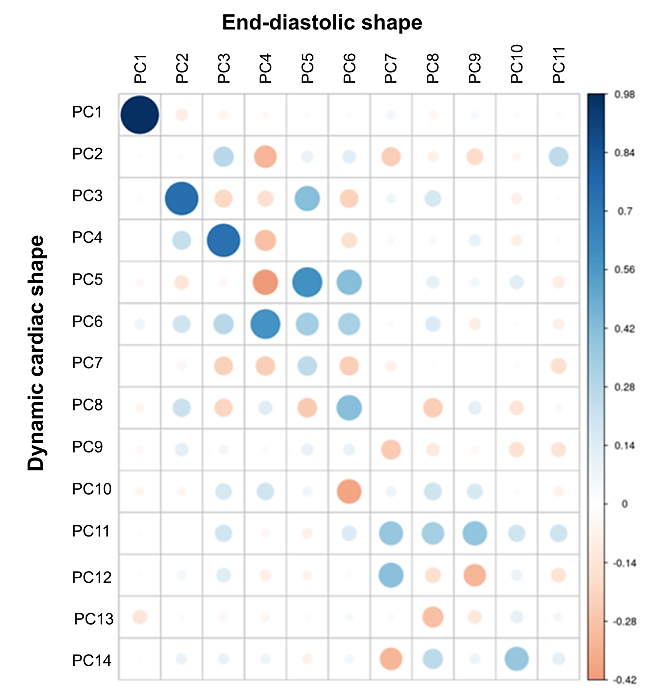

*PC, principal component; ED, end-diastolic; EDES, end-diastolic end-systolic dynamic cardiac shape.*

Supplementary Figure 4 – PC1 Manhattan plot

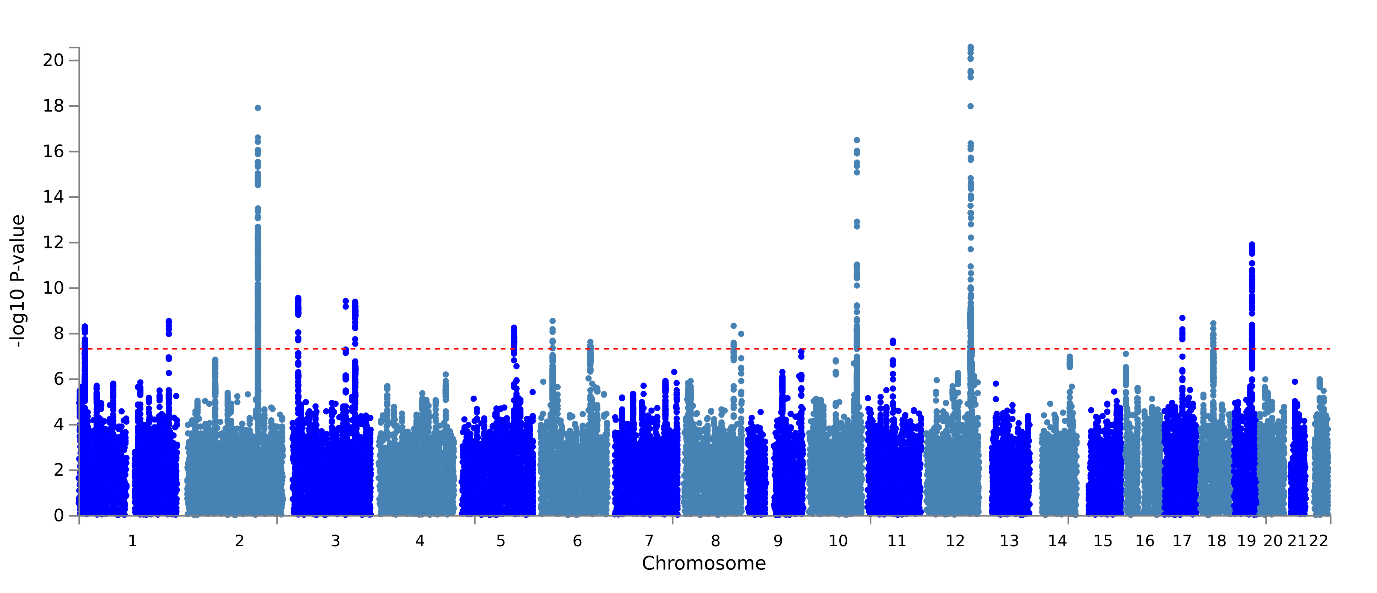

Supplementary Figure 5 – PC2 manhattan plot

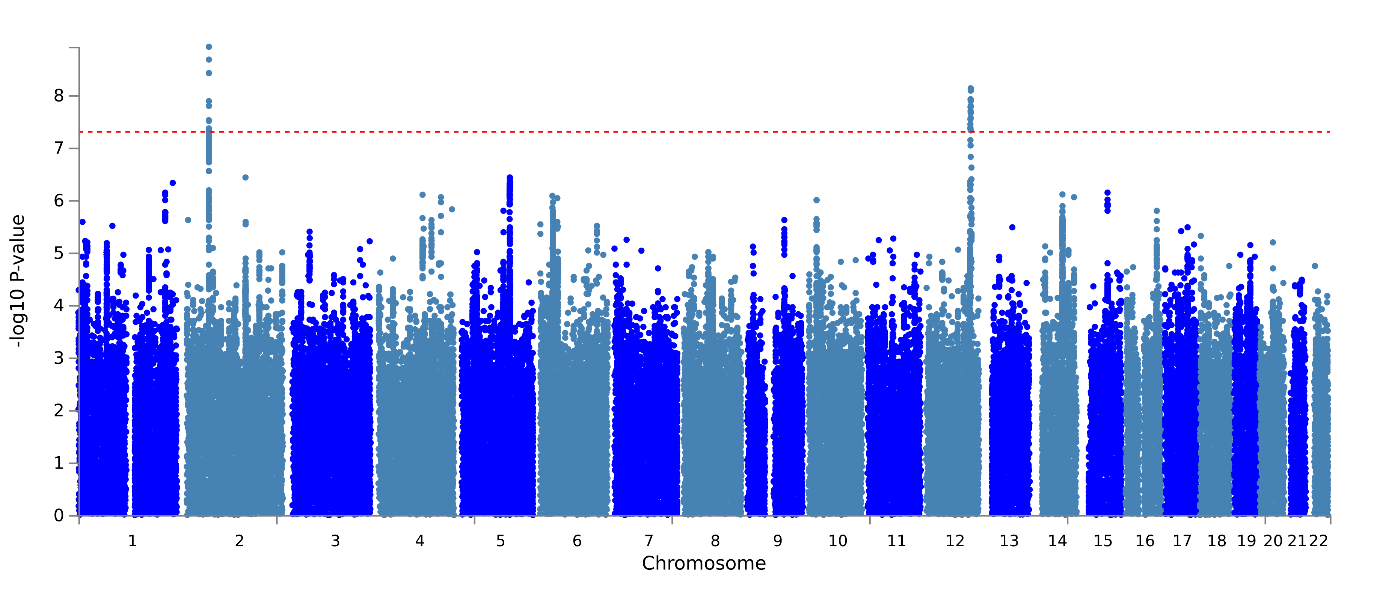

Supplementary Figure 6 – PC3 Manhattan plot

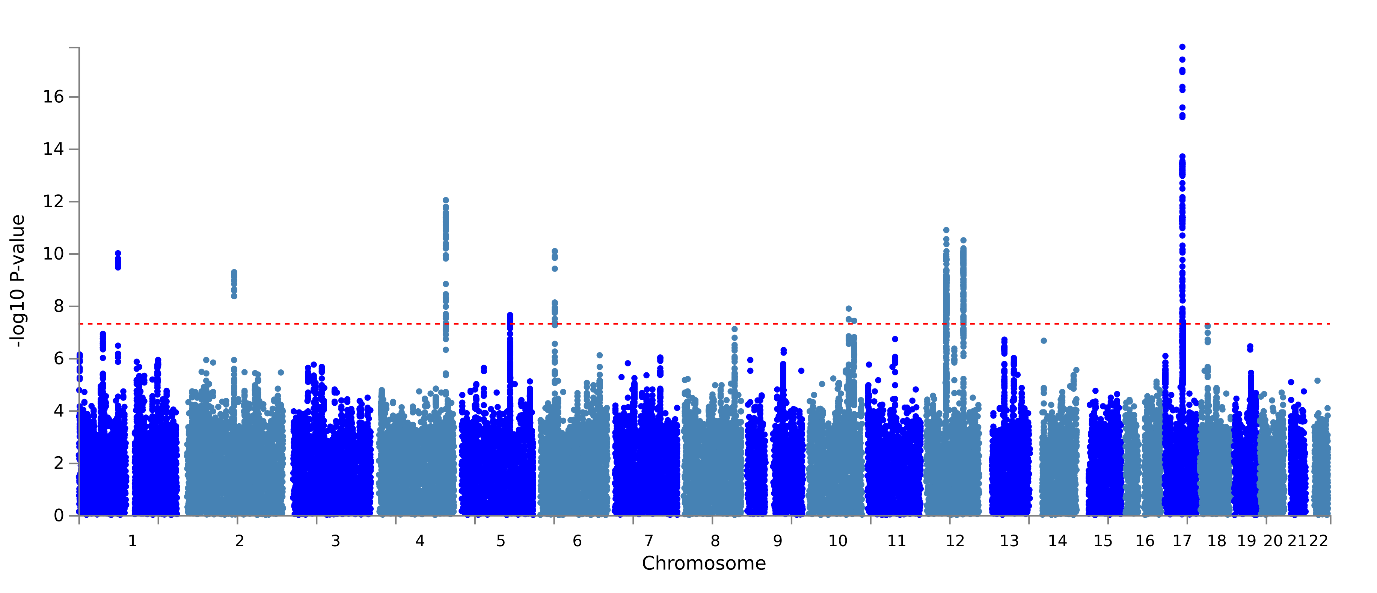

Supplementary Figure 7 – PC4 Manhattan plot
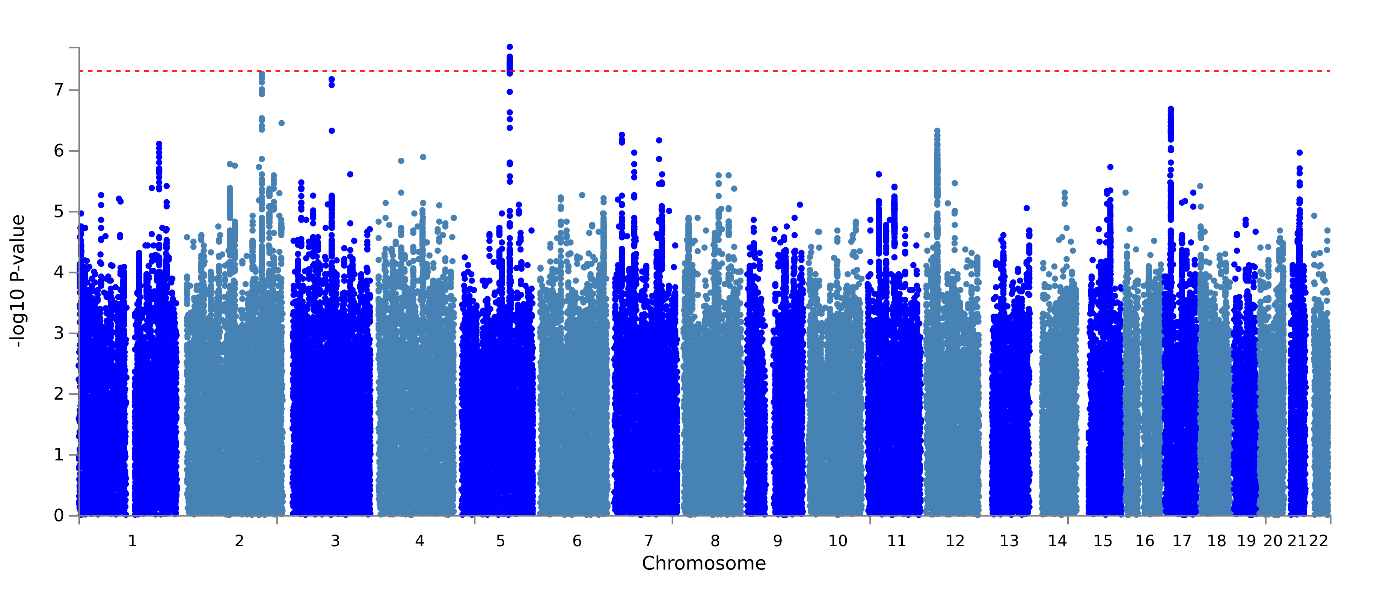

Supplementary Figure 8 – PC5 Manhattan plot

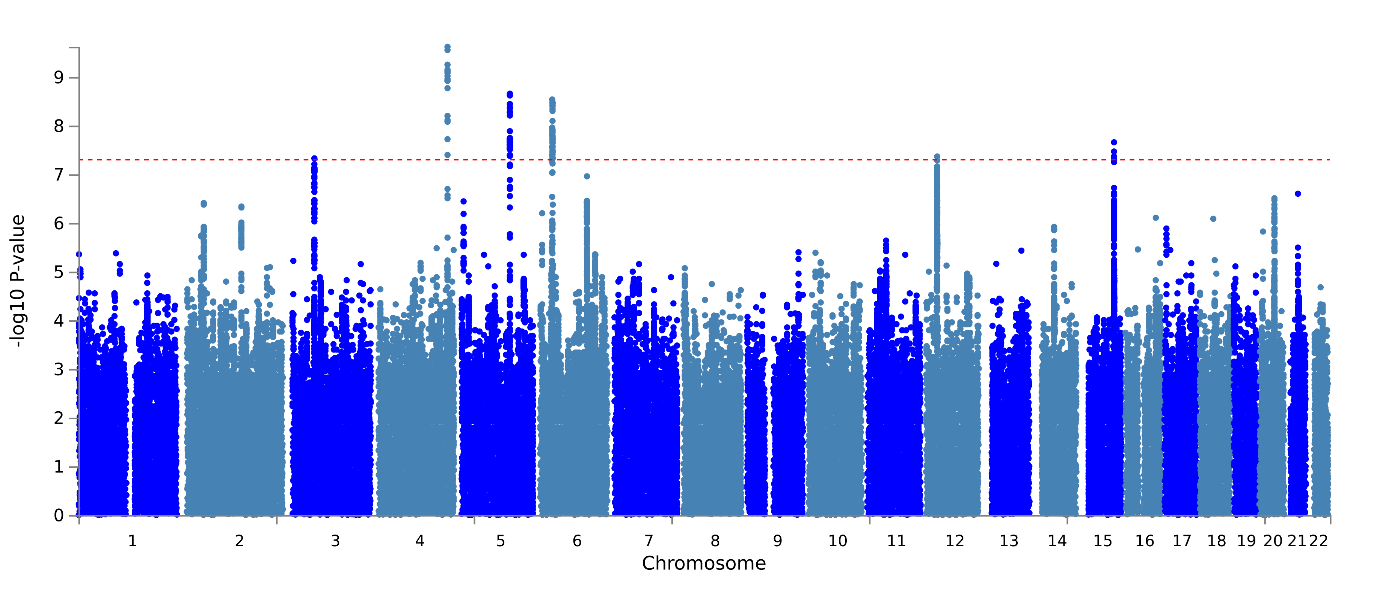

Supplementary Figure 9 – PC6 Manhattan plot

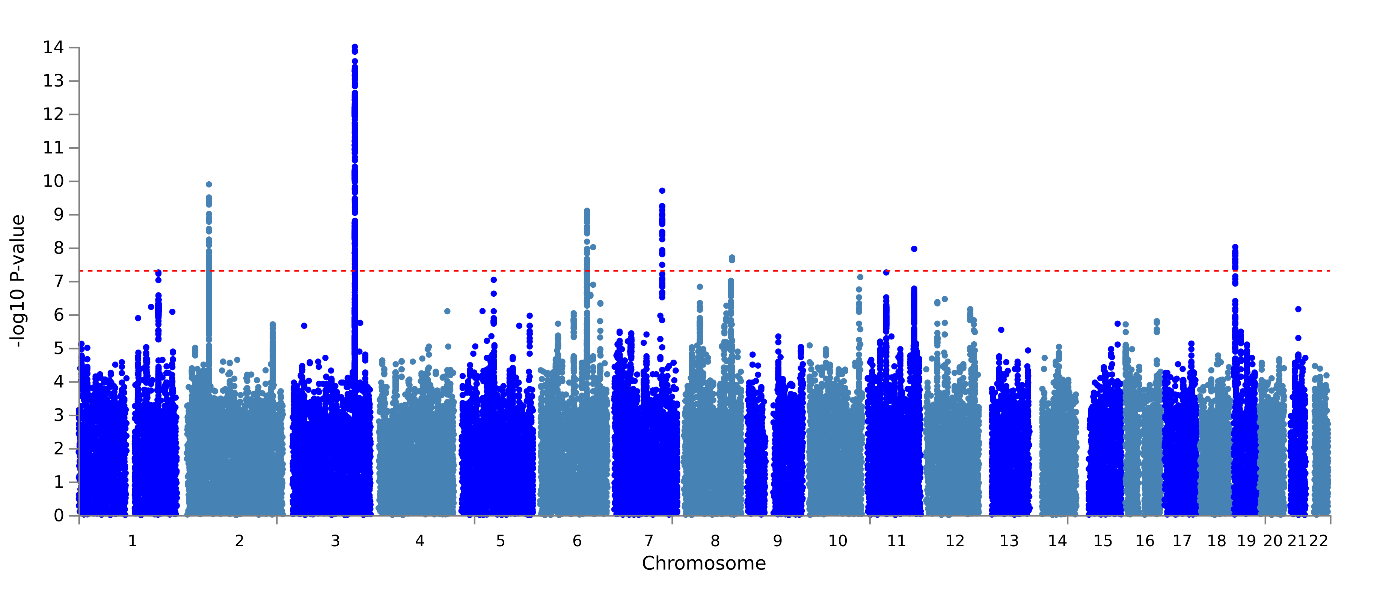

Supplementary Figure 10 – PC7 Manhattan plot

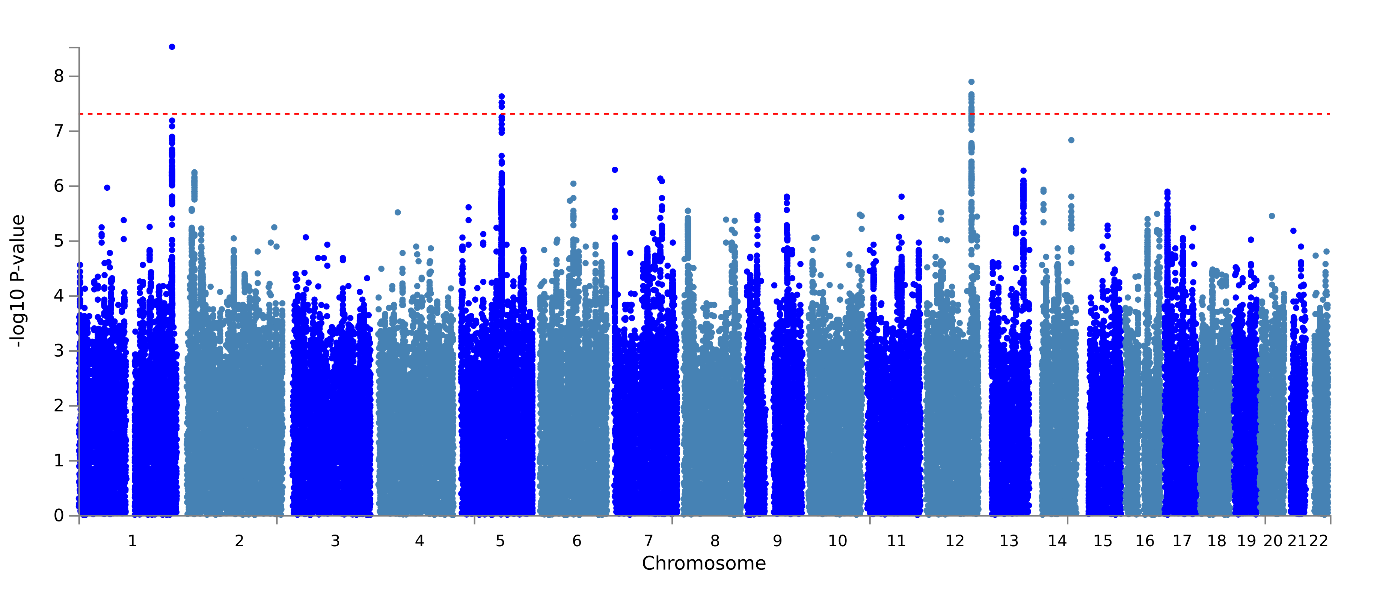

Supplementary Figure 11 – PC8 Manhattan plot

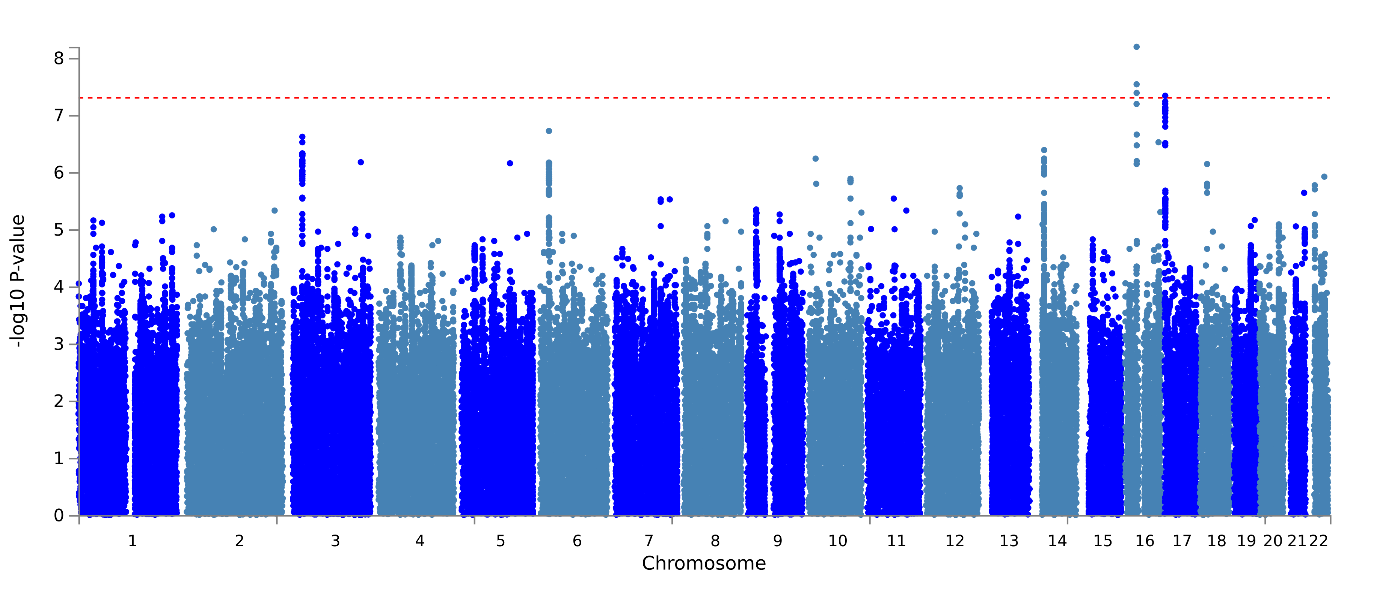

Supplementary Figure 12 – PC9 Manhattan plot

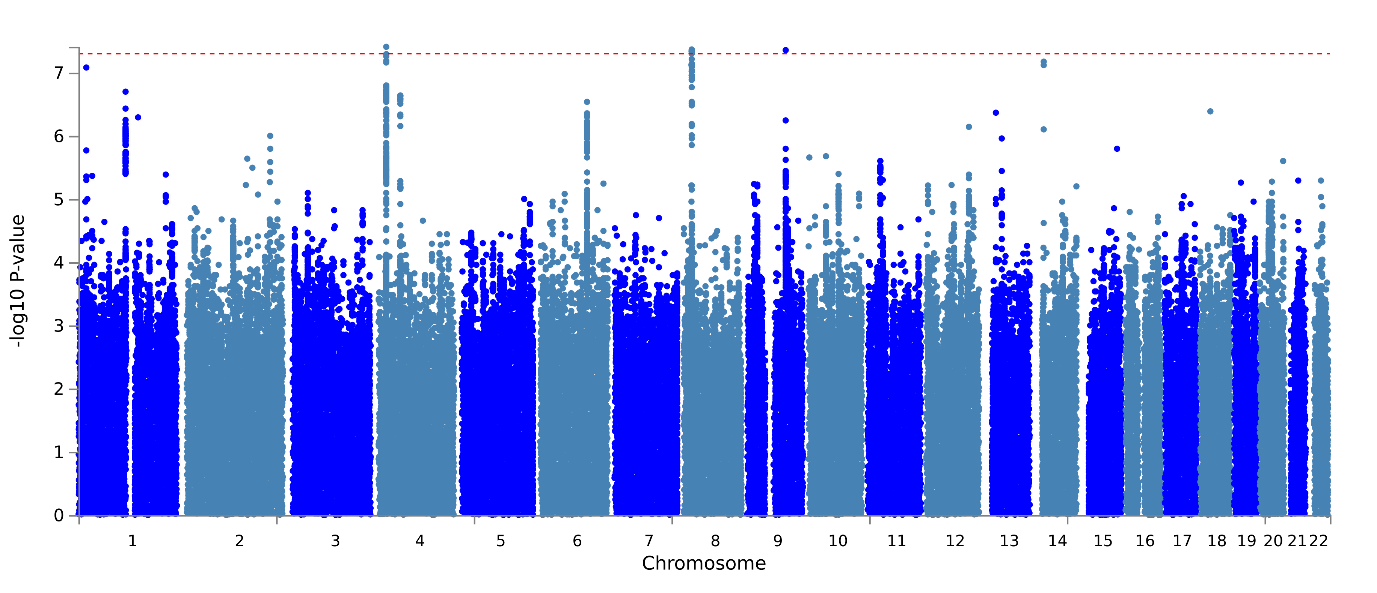

Supplementary Figure 13 – PC10 Manhattan plot

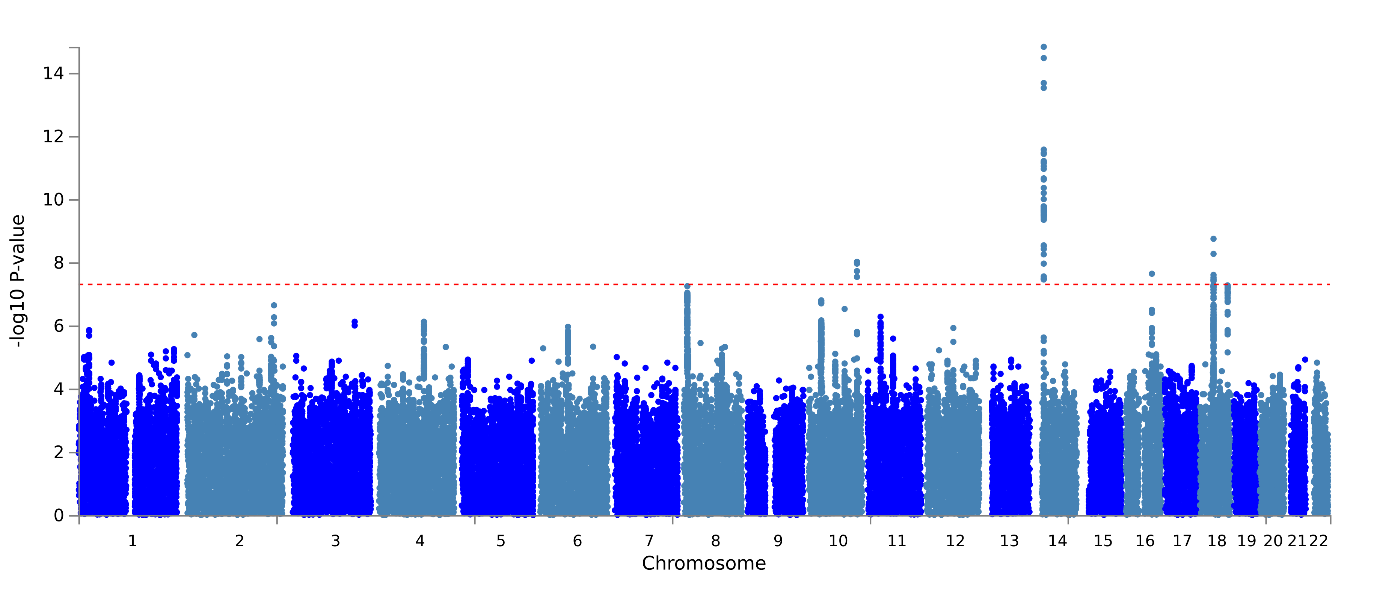

Supplementary Figure 14 – PC11 Manhattan plot

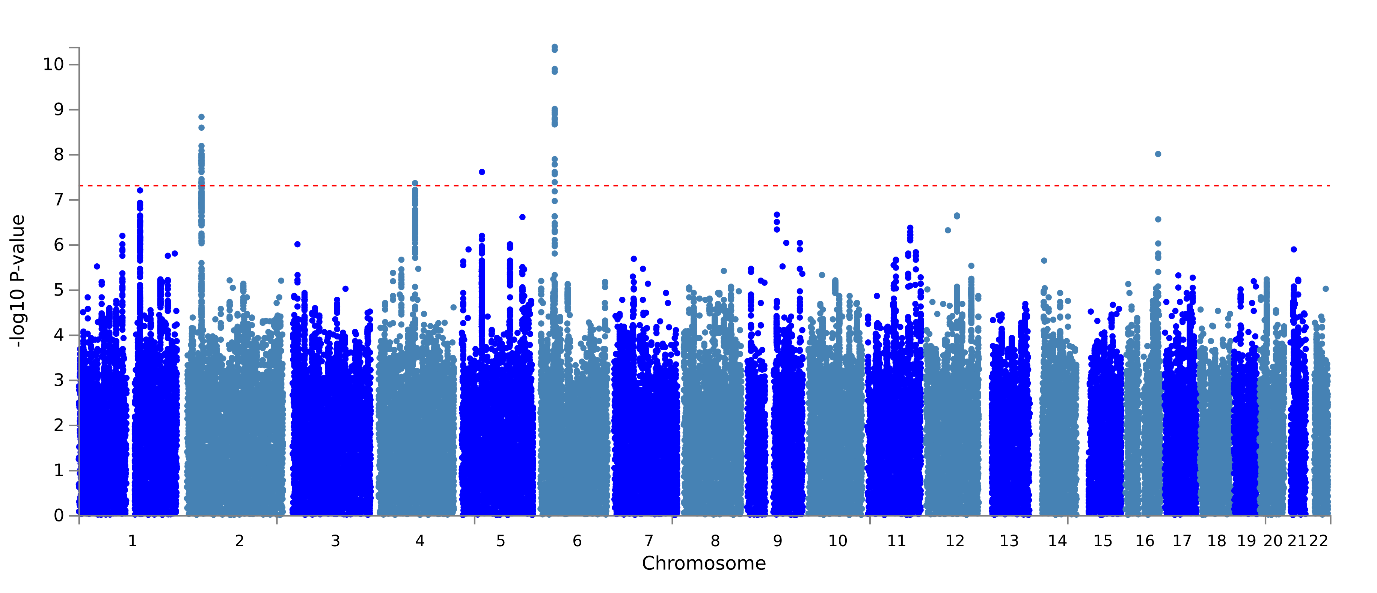

Supplementary Figure 15 – PC12 Manhattan plot

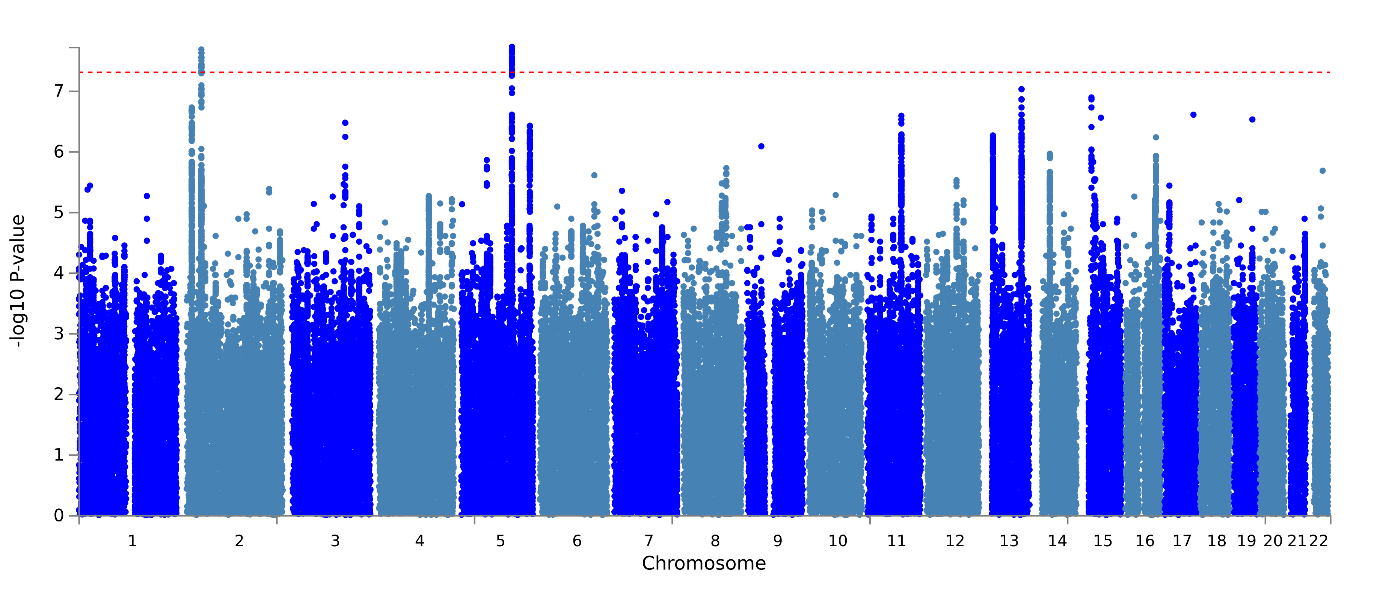

Supplementary Figure 16 – PC13 Manhattan plot

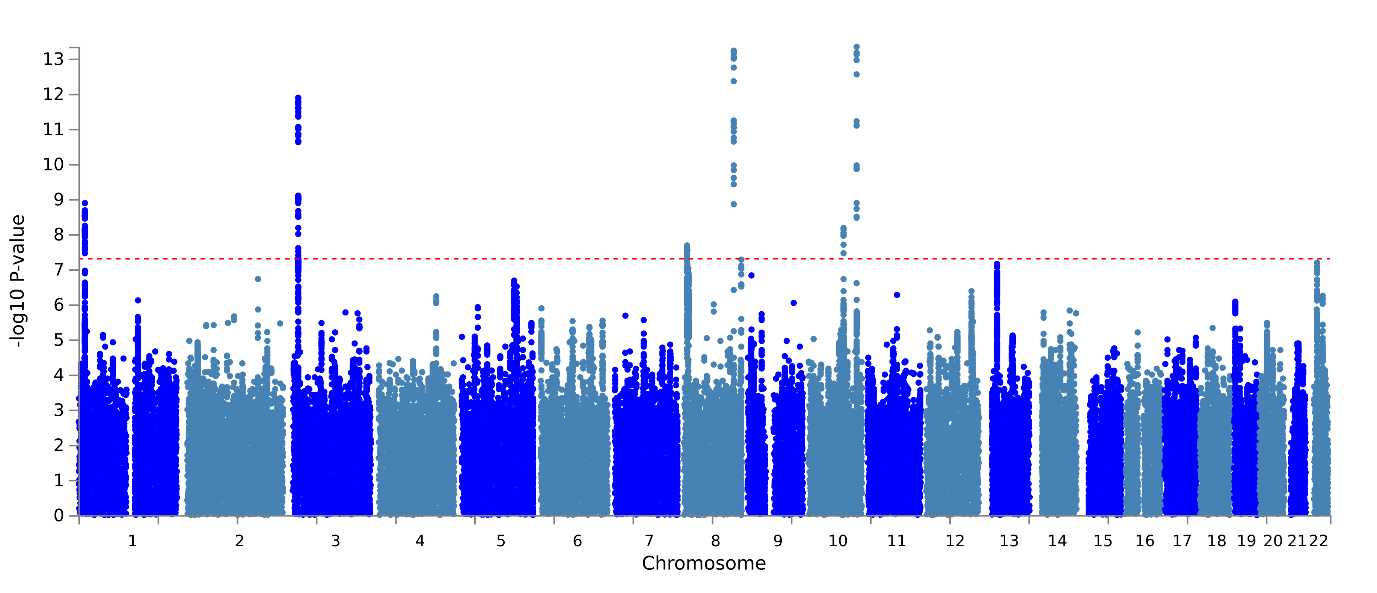

Supplementary Figure 17 – PC14 Manhattan plot

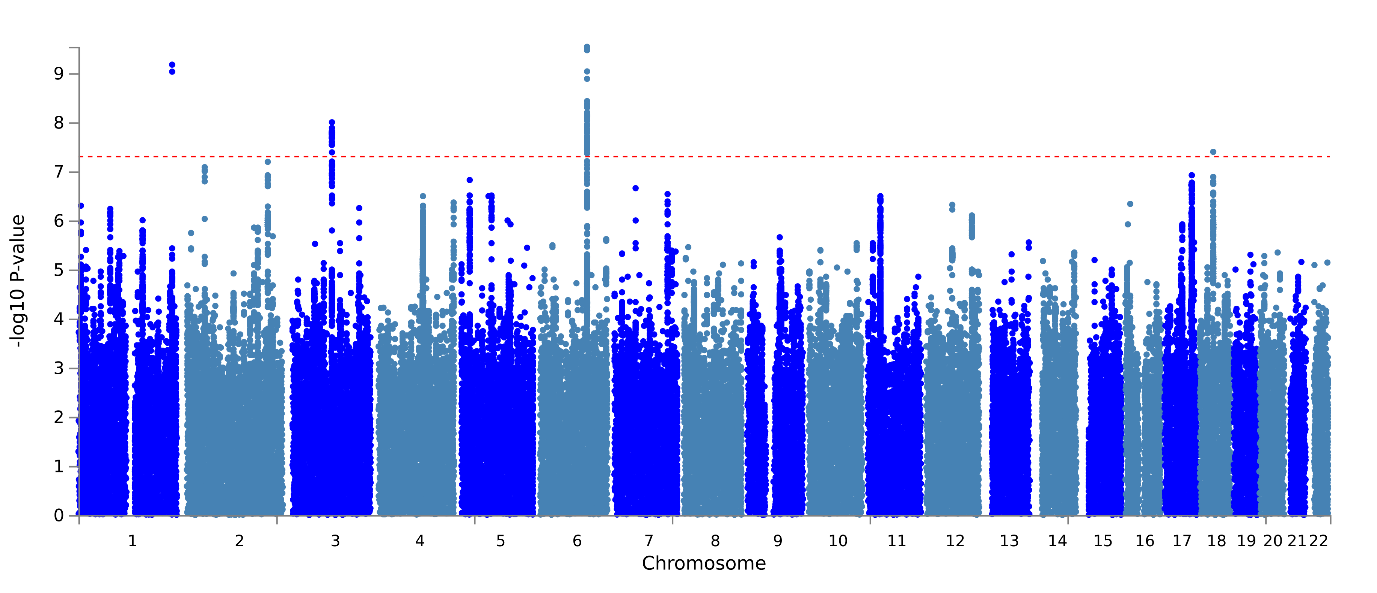

Supplementary Figure 18 – QQ PC1

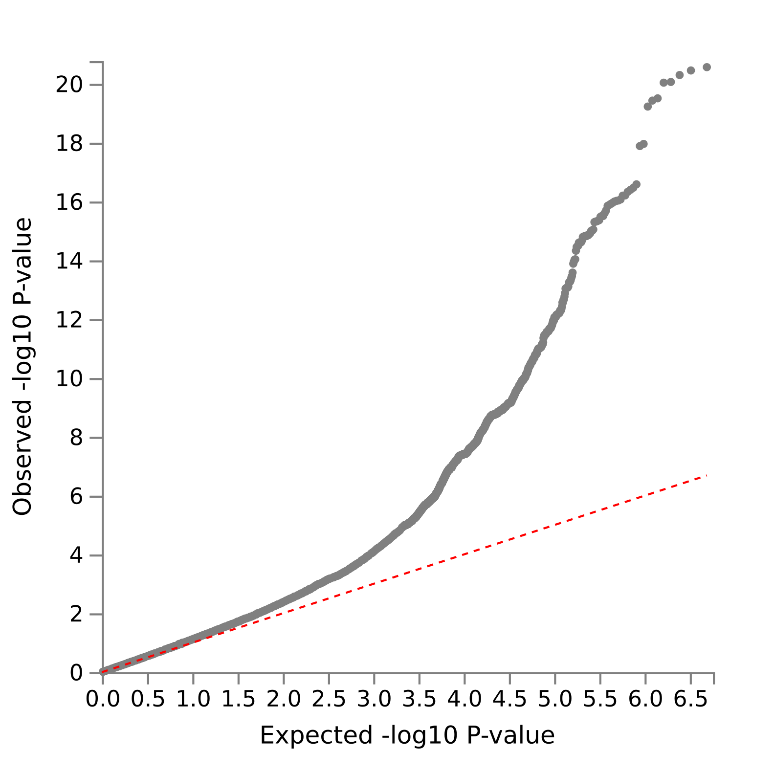

Supplementary Figure 19 – QQ PC2

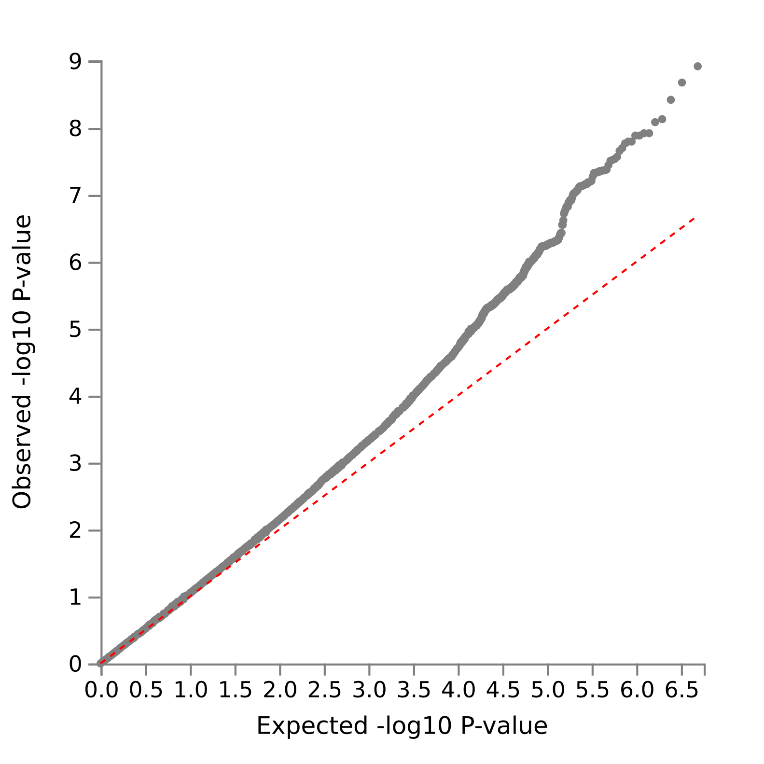

Supplementary Figure 20 – QQ PC3

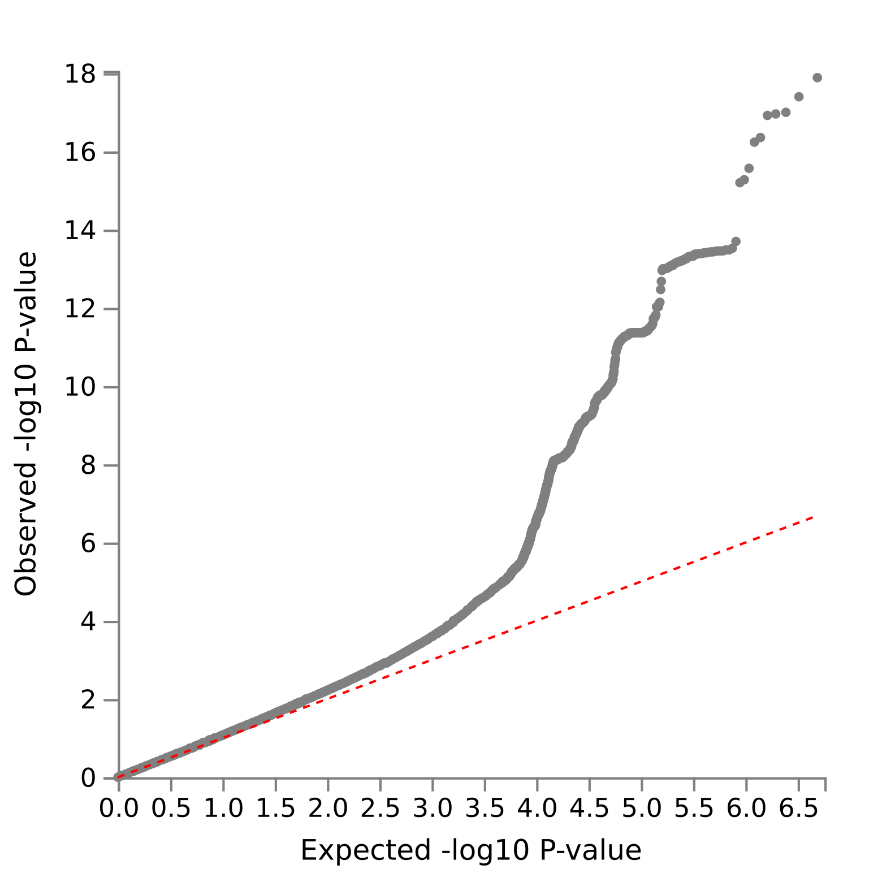

Supplementary Figure 21 - QQ PC4

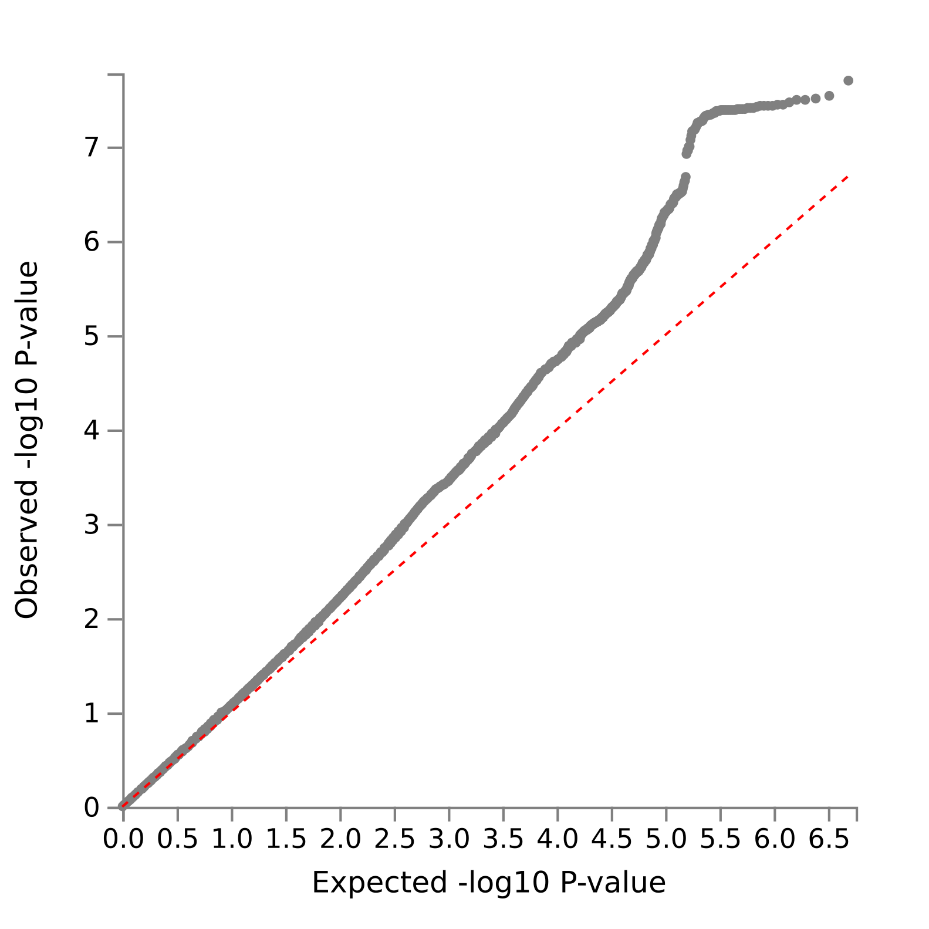

Supplementary Figure 22 – QQ PC5

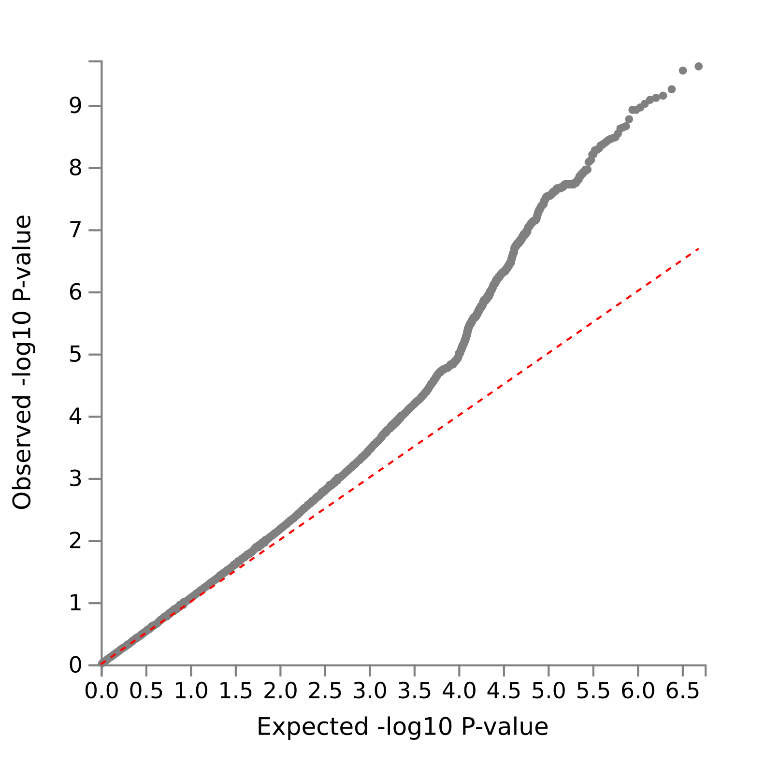

Supplementary Figure 23 – QQ PC6

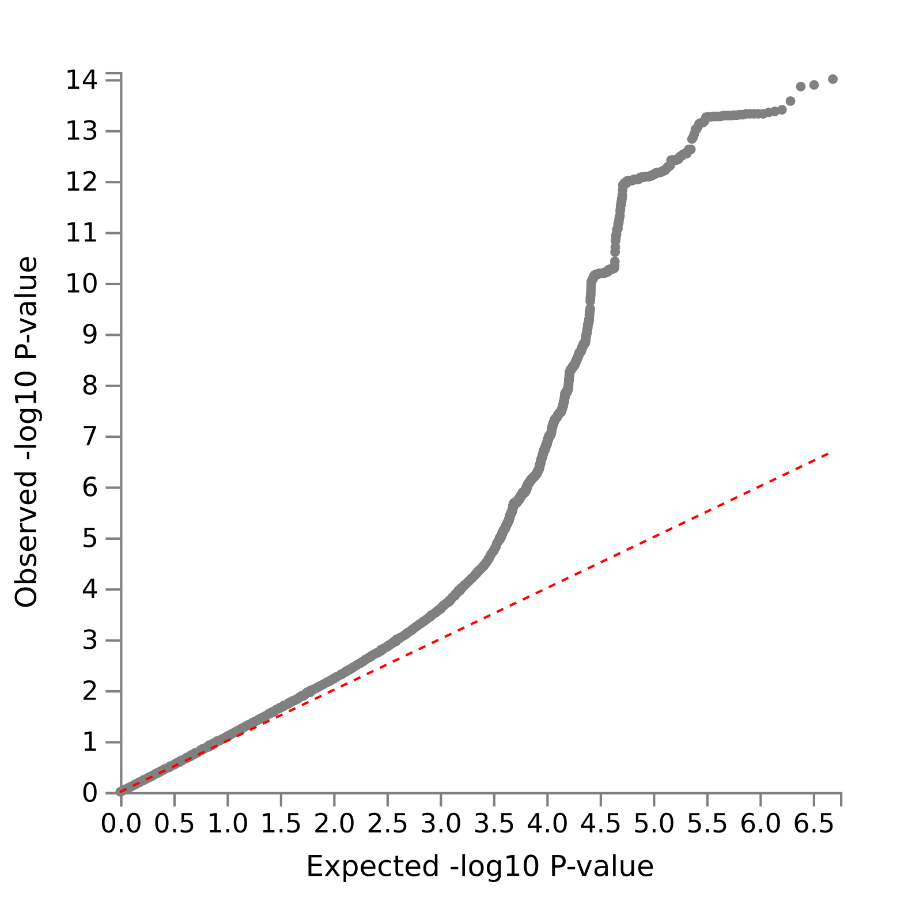

Supplementary Figure 24 – QQ PC7

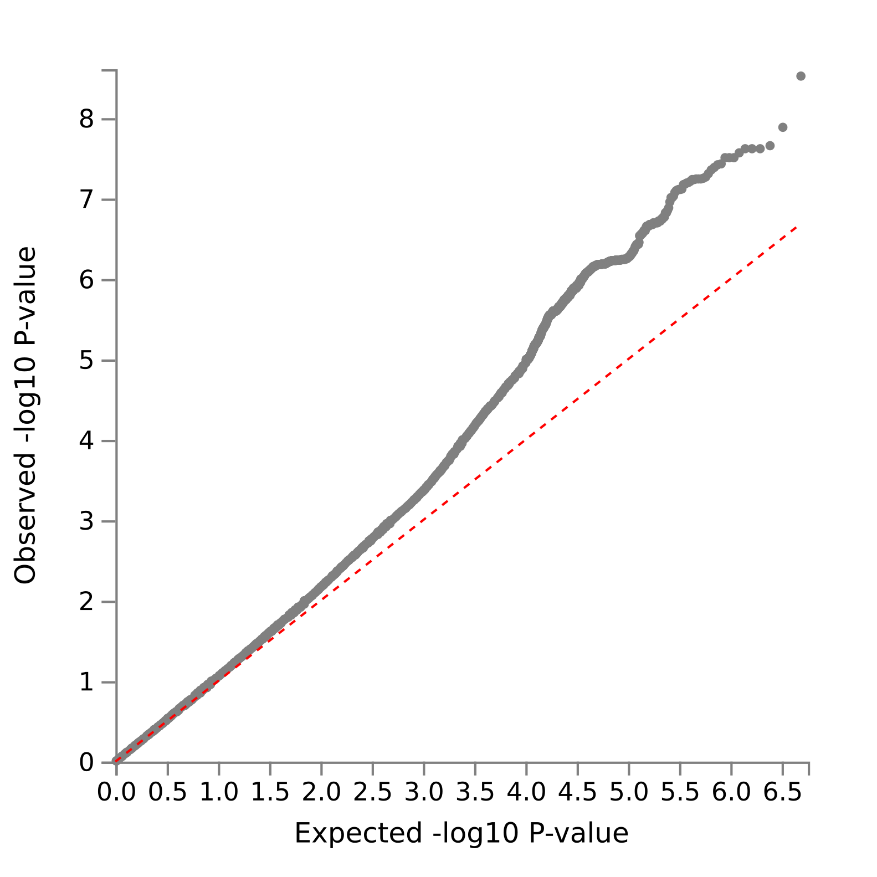

Supplementary Figure 25 – QQ PC8

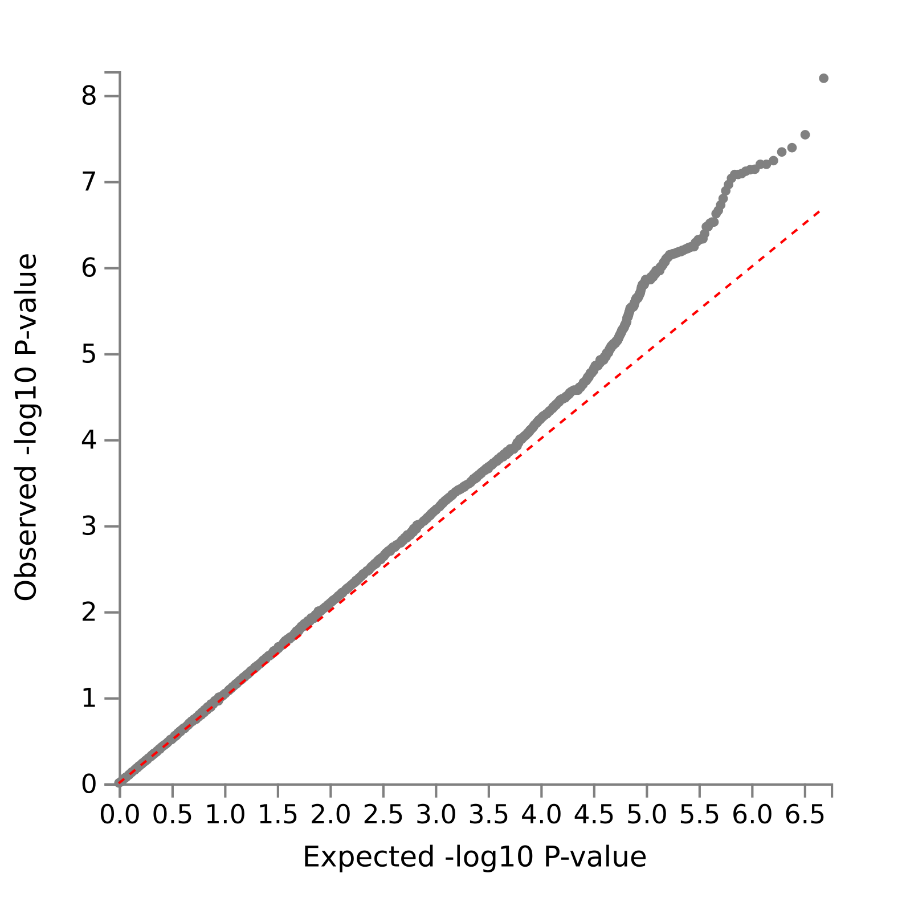

Supplementary Figure 26 - QQ PC9

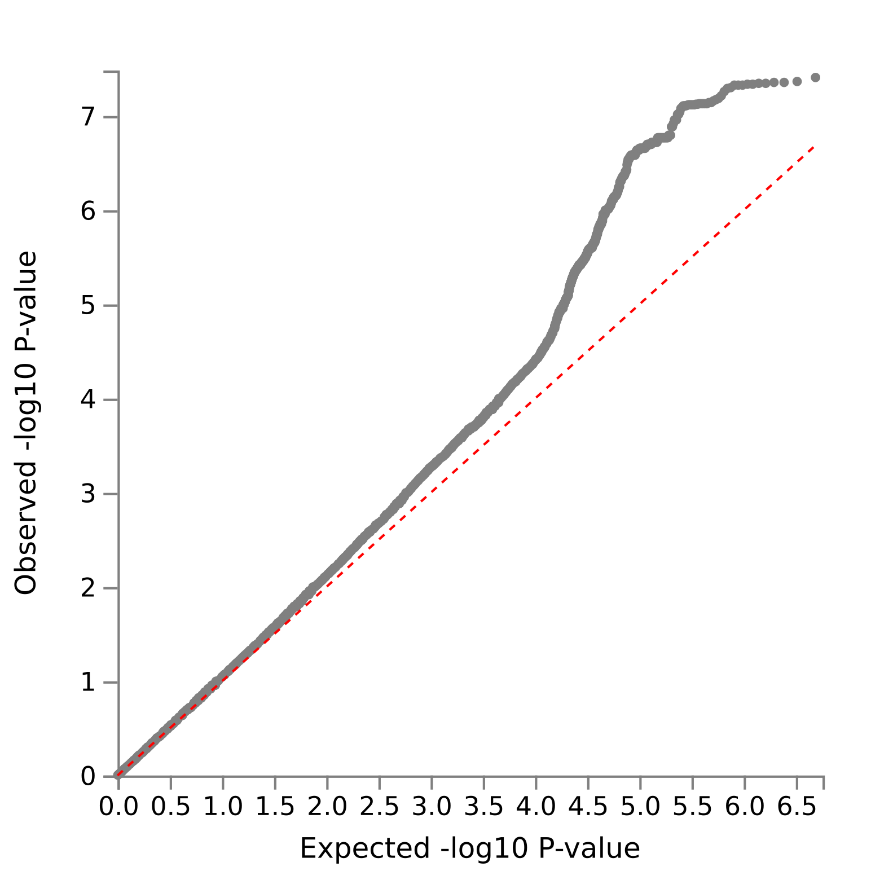

Supplementary Figure 27 – QQ PC10

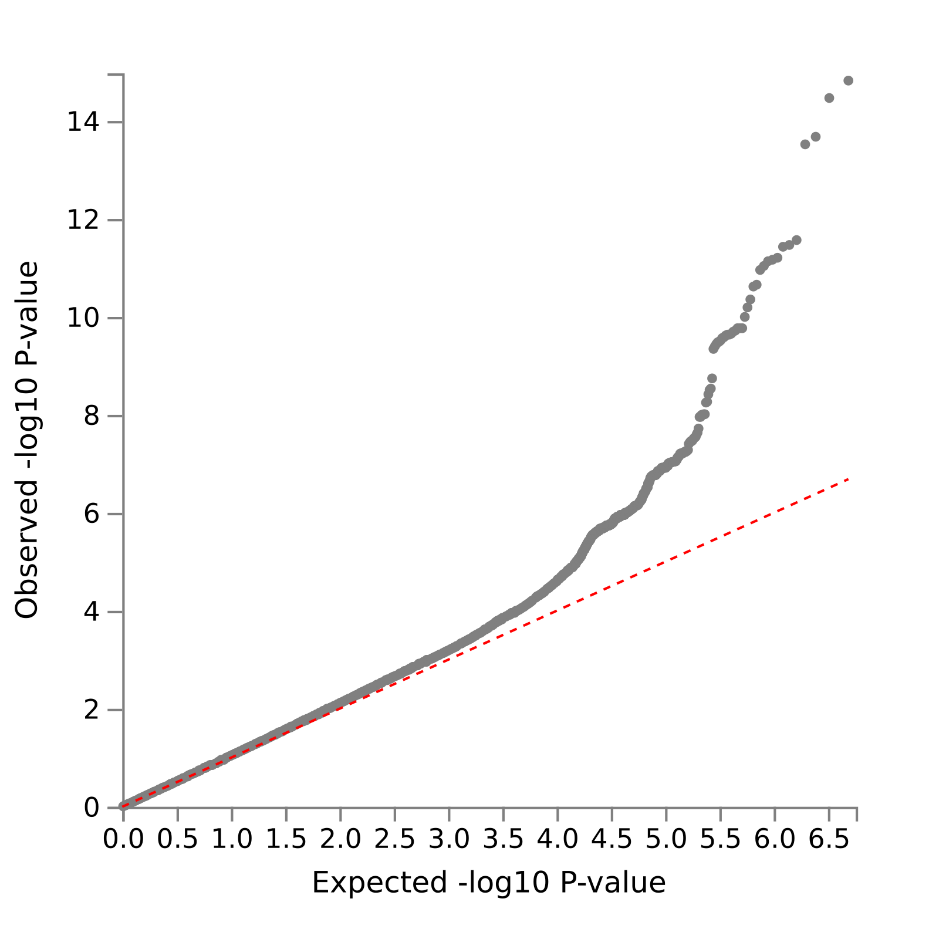

Supplementary Figure 28 - QQ PC11

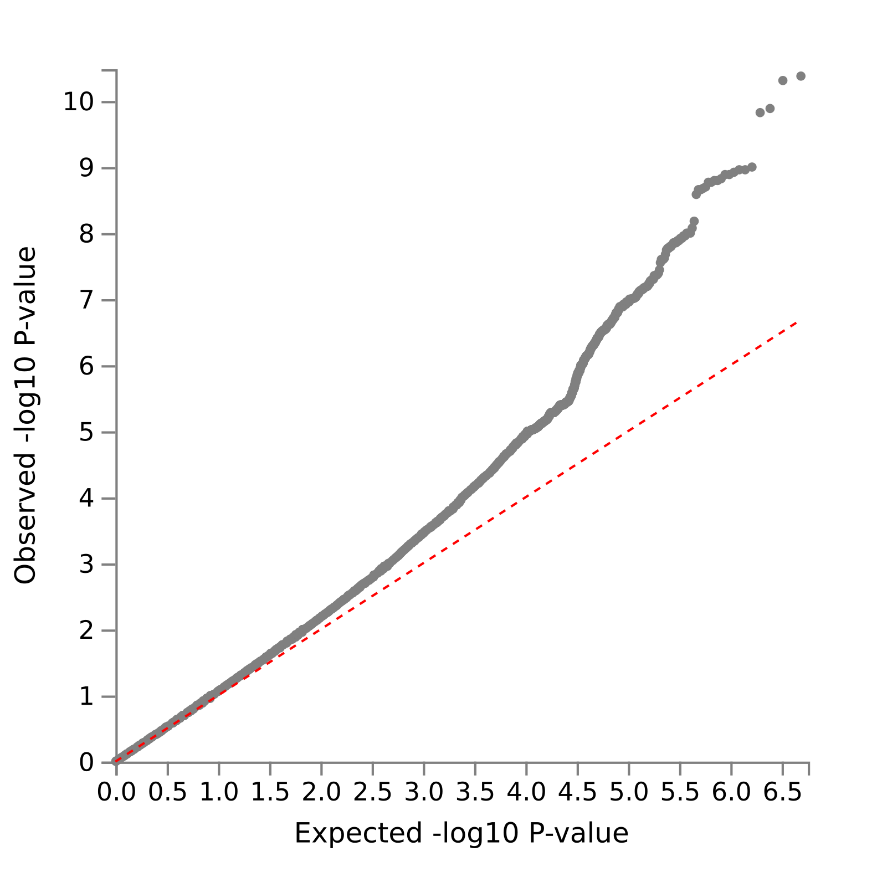

Supplementary Figure 29 - QQ PC12

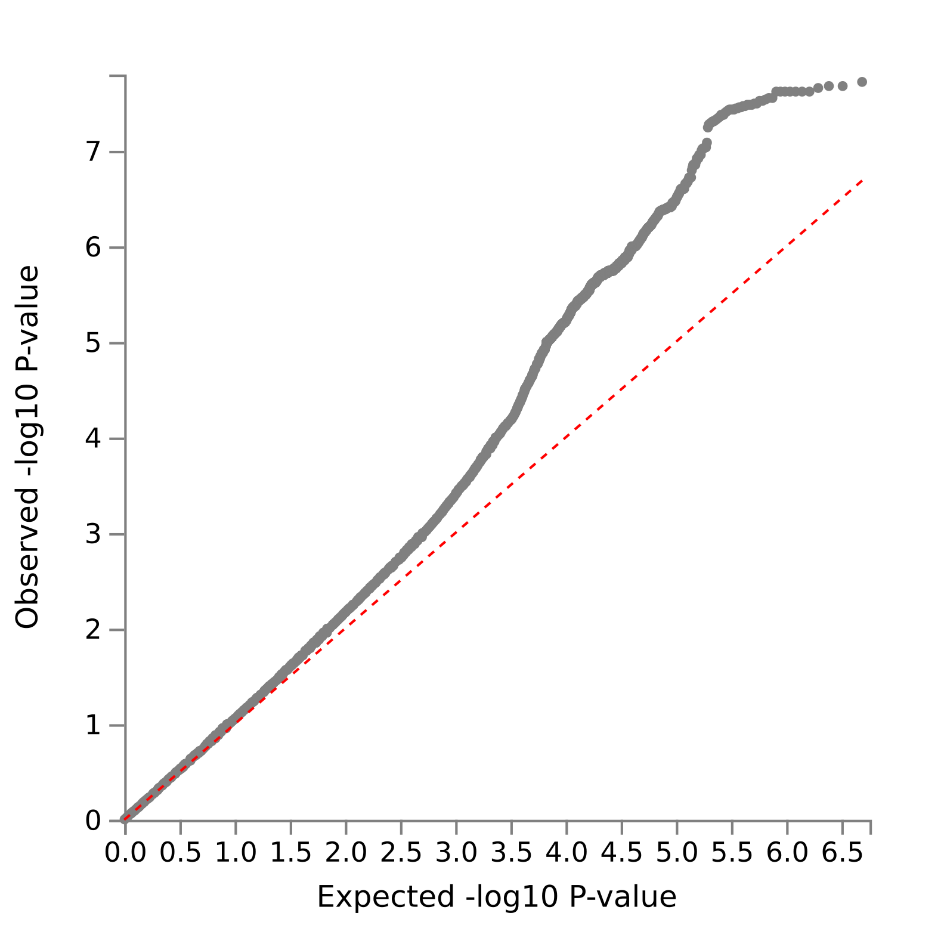

Supplementary Figure 30 – QQ PC13

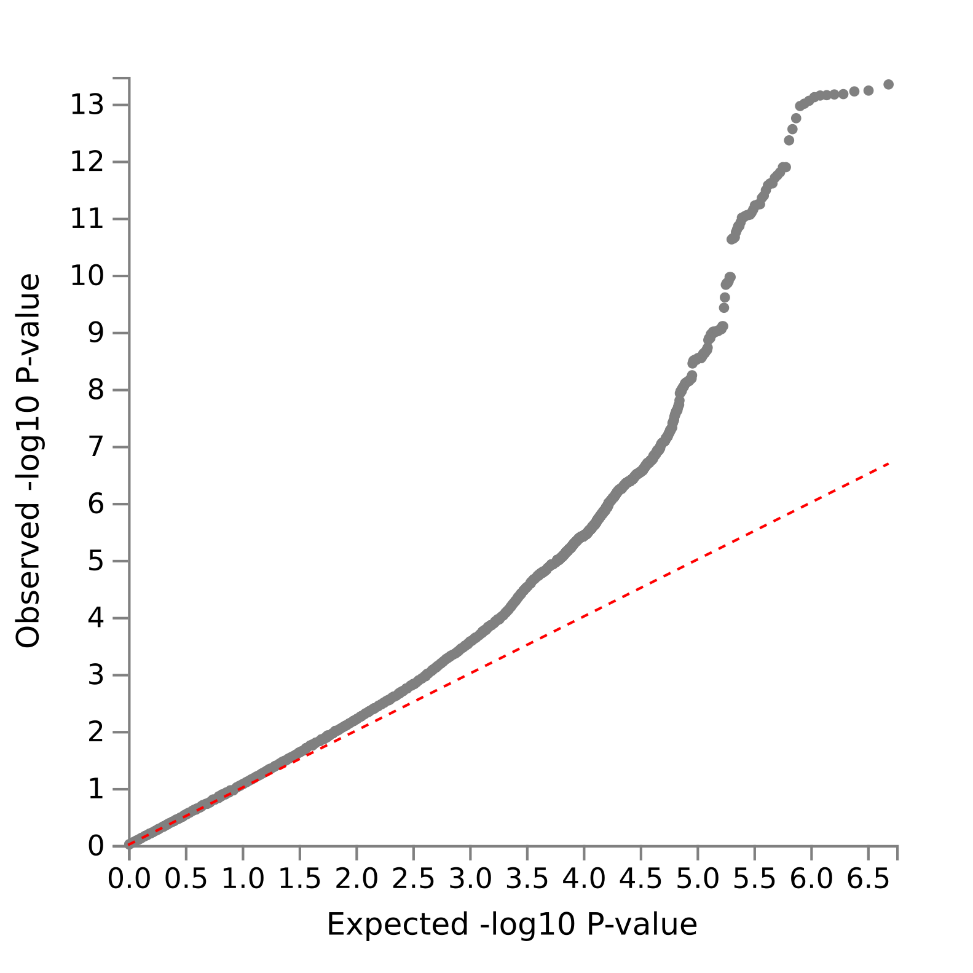

Supplementary Figure 31 - QQ PC14

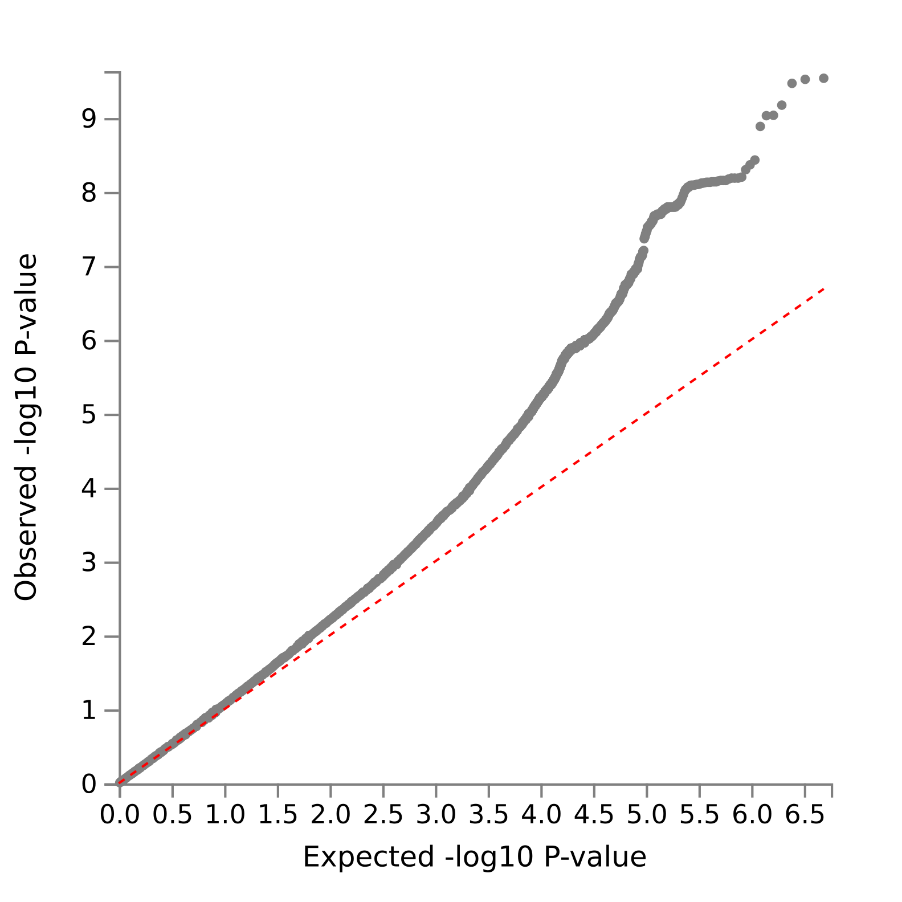

Supplementary Figure 32 - Locus zoom rs653178 PC1

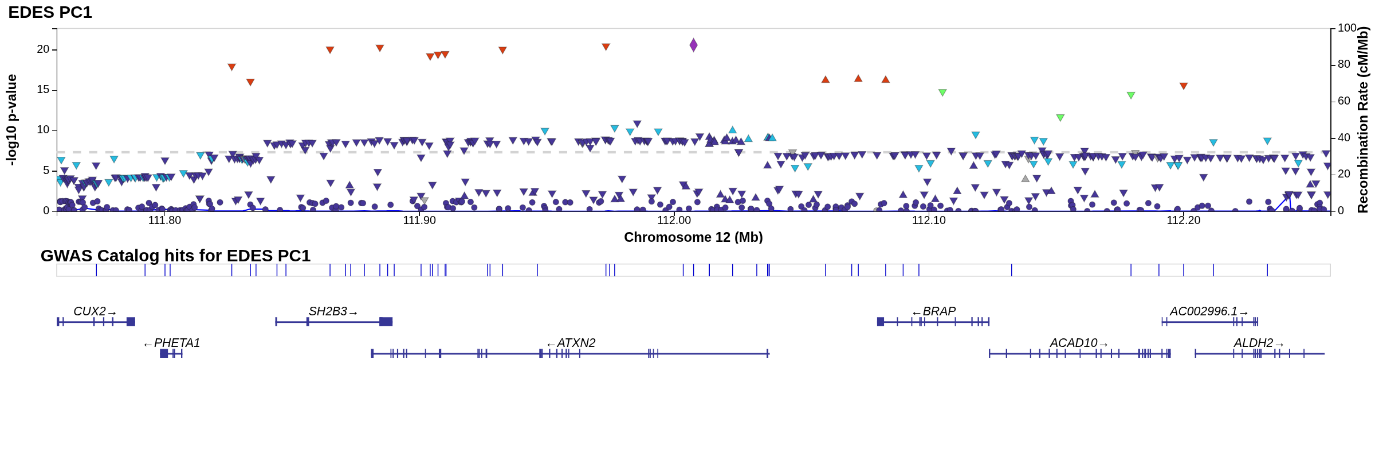

Supplementary Figure 33 – Locus zoom rs2042995 PC1

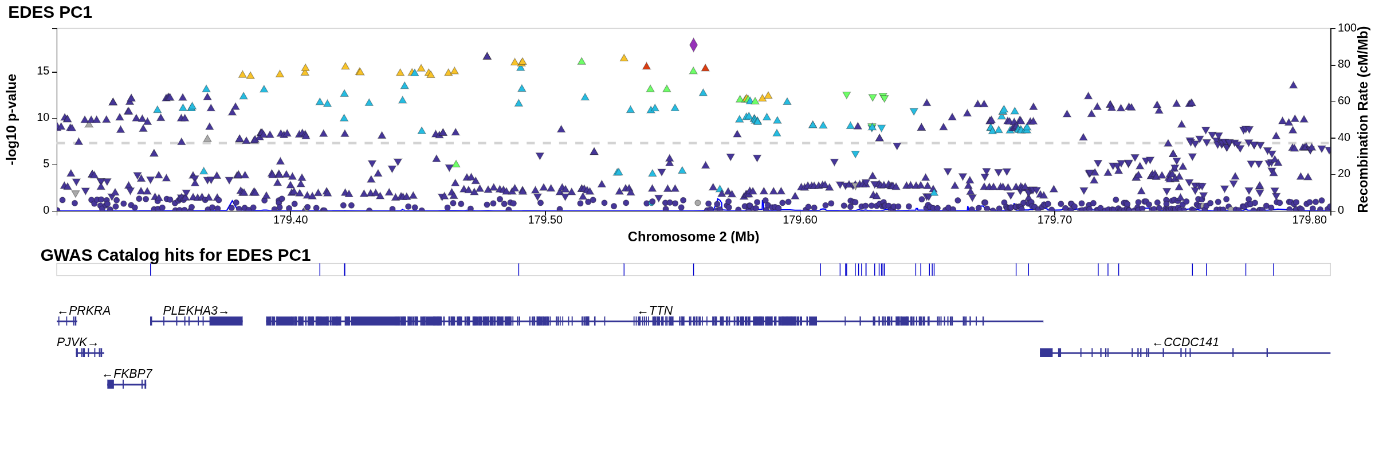
Supplementary Figure 34 – Locus zoom rs72840788 PC1

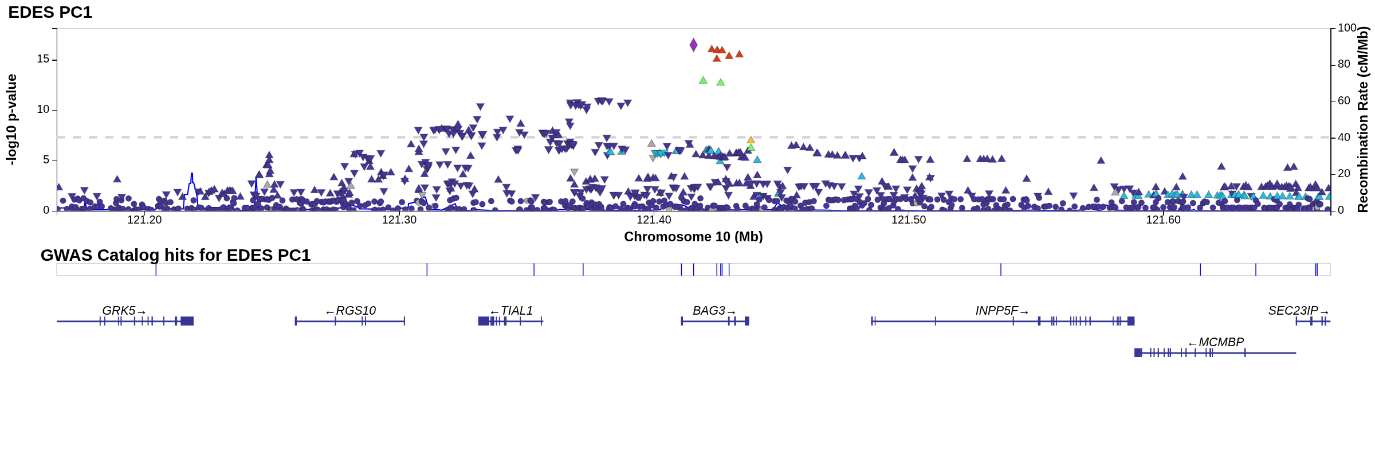

Supplementary Figure 35 – Locus zoom rs7257103 PC1

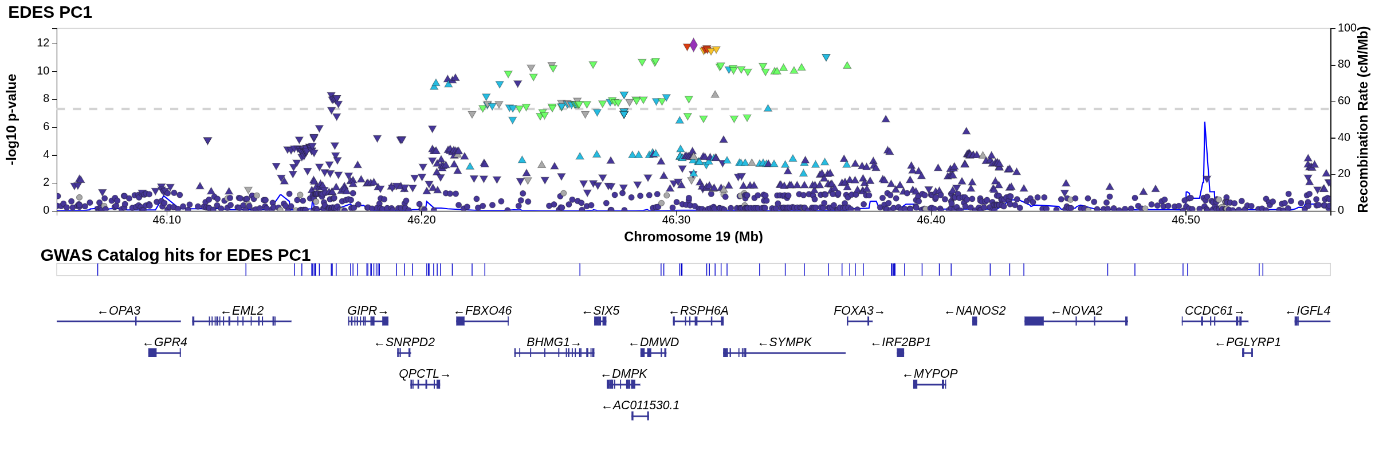
Supplementary Figure 36 – Locus zoom rs73028849 PC1

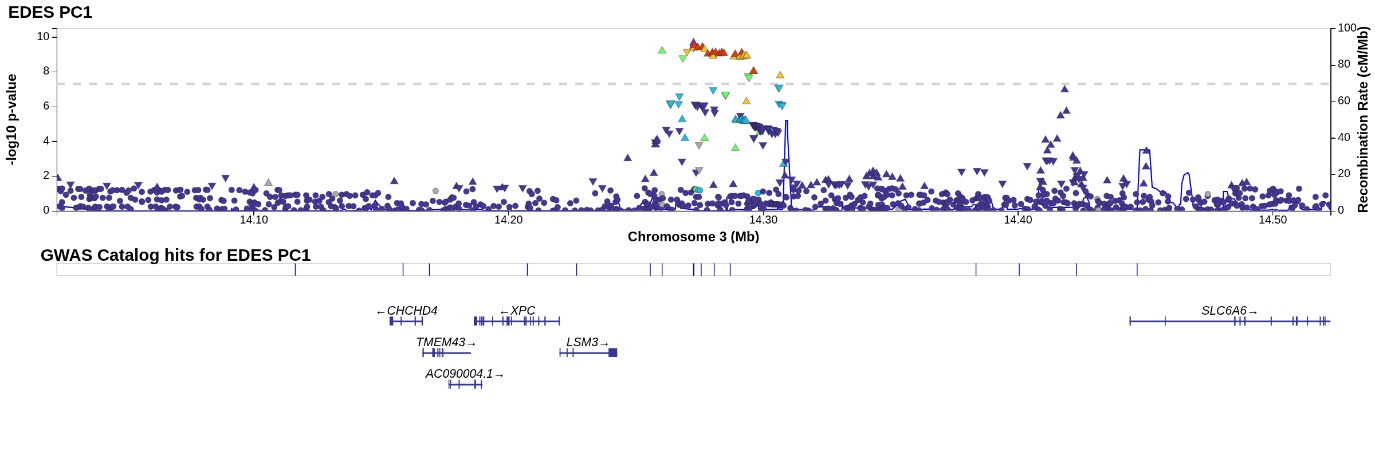
Supplementary Figure 37 – Locus zoom rs77709005 PC1

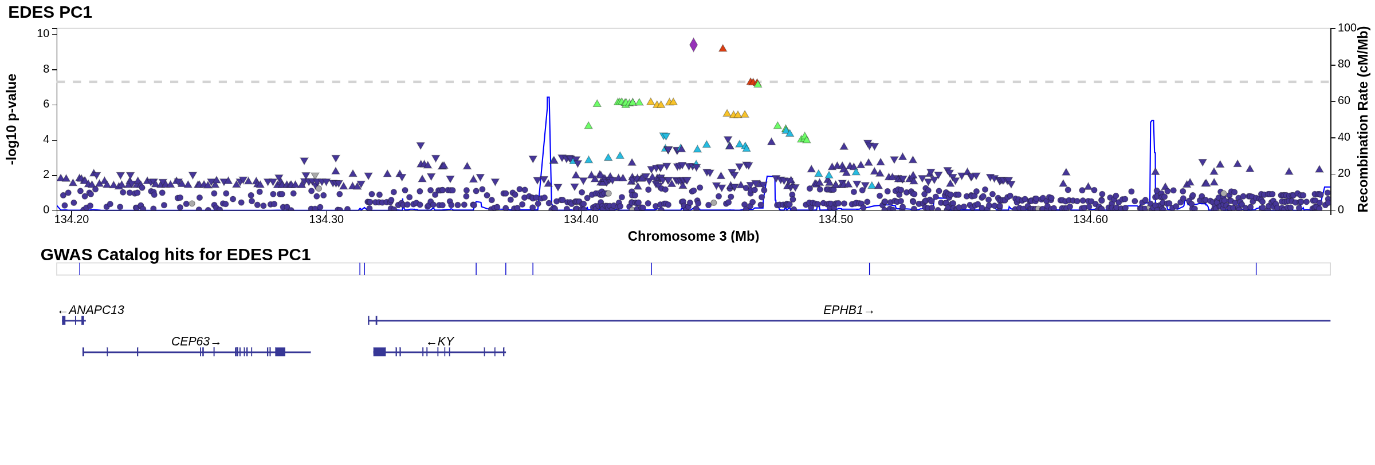

Supplementary Figure 38 – Locus zoom rs9810339 PC1

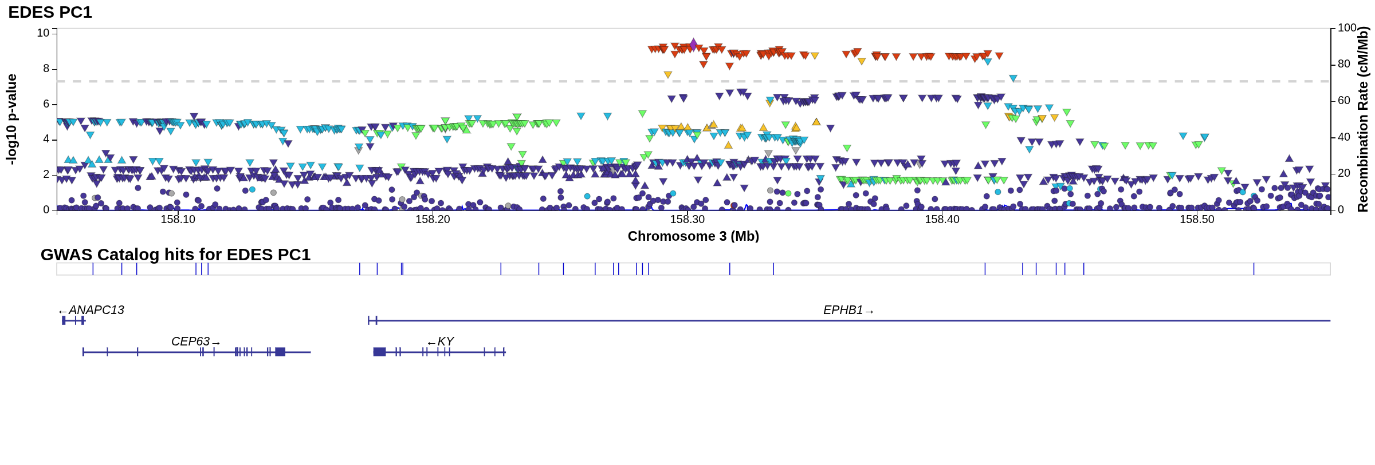

Supplementary Figure 39 – Locus zoom rs533030436 PC1

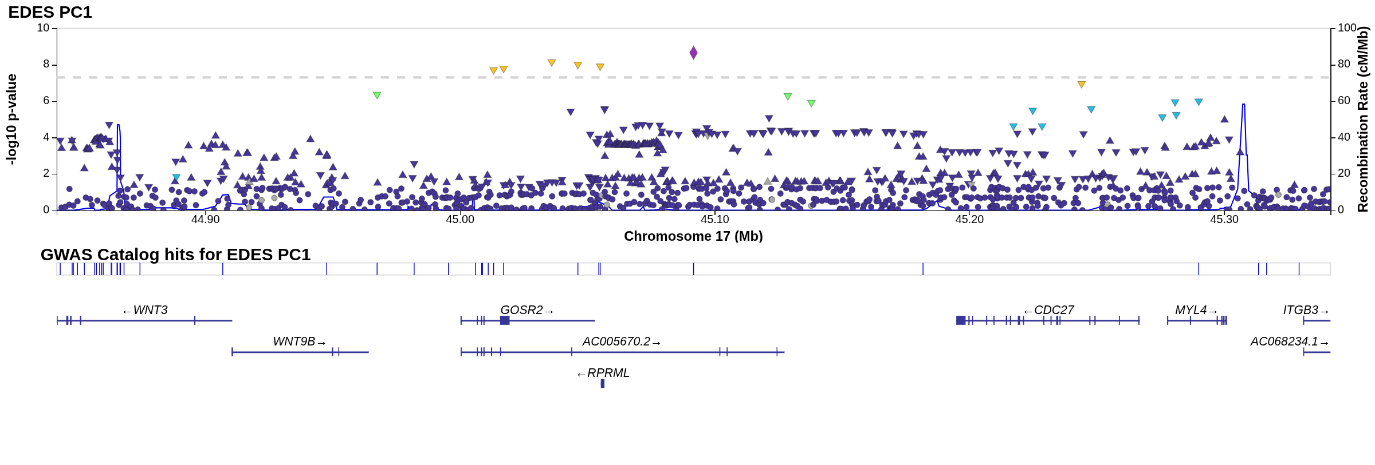

Supplementary Figure 40 – Locus zoom rs78529941 PC1
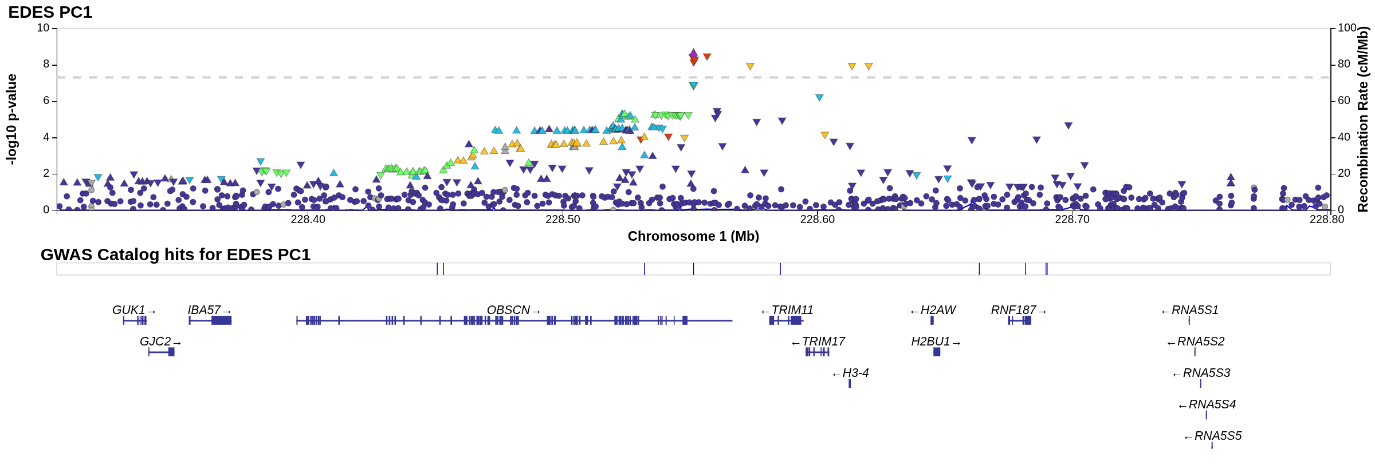

Supplementary Figure 41 – Locus zoom rs6939900 PC1

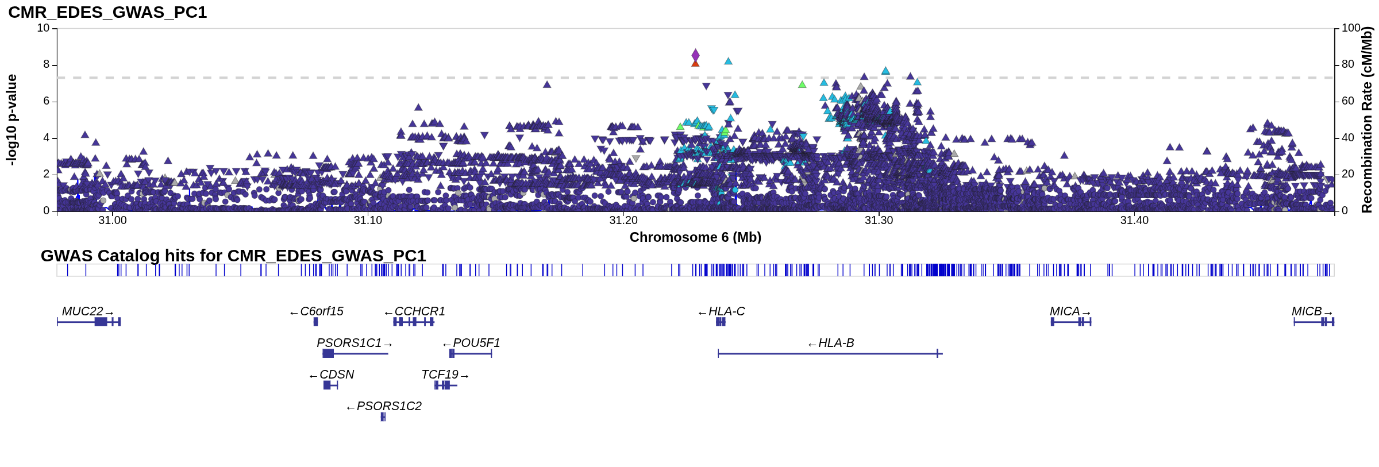

Supplementary Figure 42 – Locus zoom rs2644262 PC1

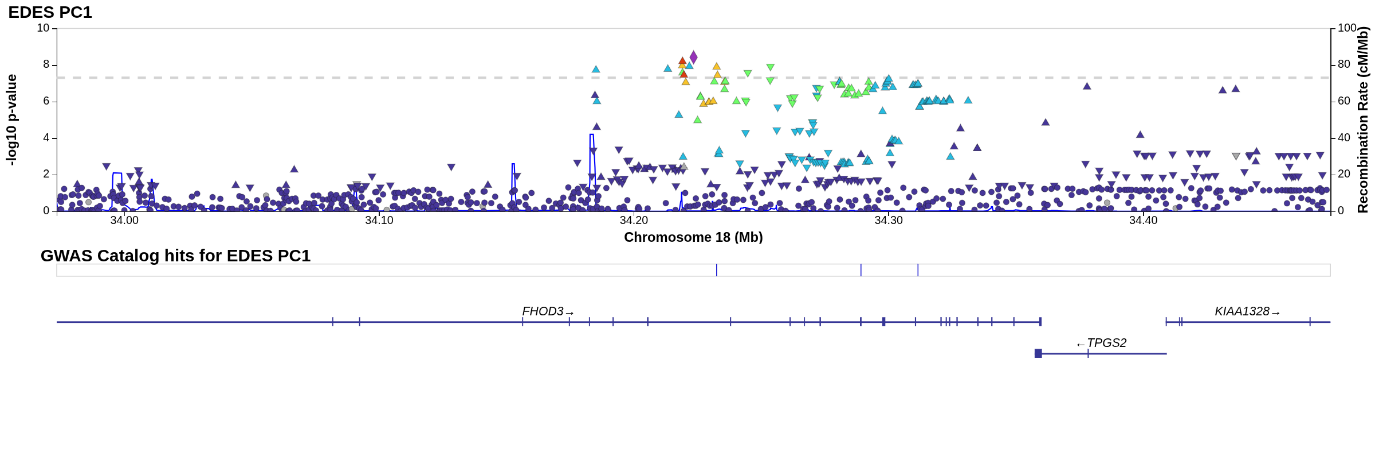

Supplementary Figure 43 – Locus zoom rs34123659 PC1

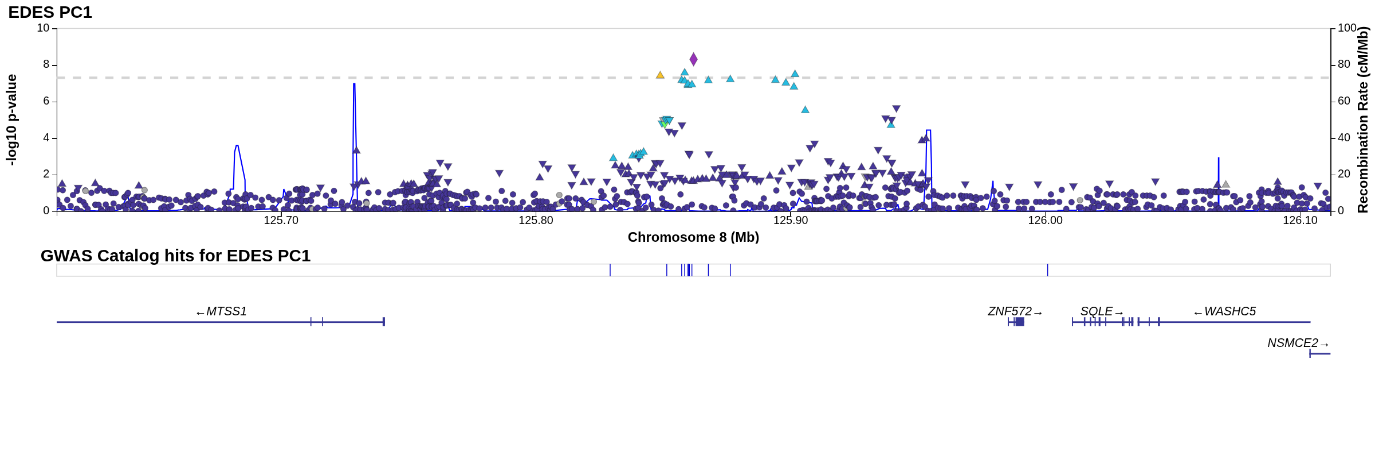

Supplementary Figure 44 – Locus zoom rs28579893 PC1

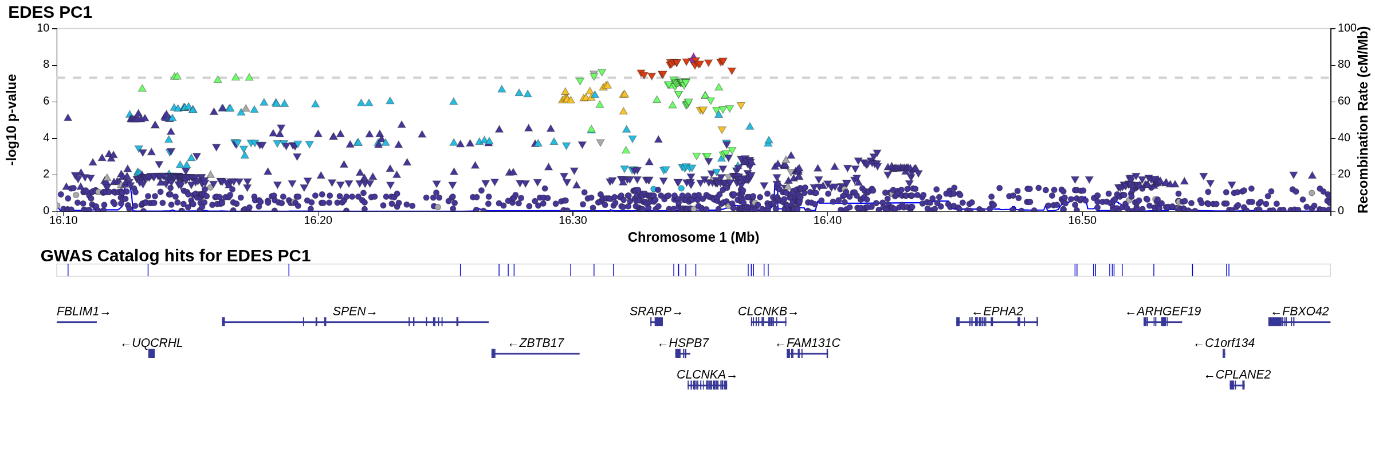

Supplementary Figure 45 – Locus zoom 5:132409975_CT_C PC1

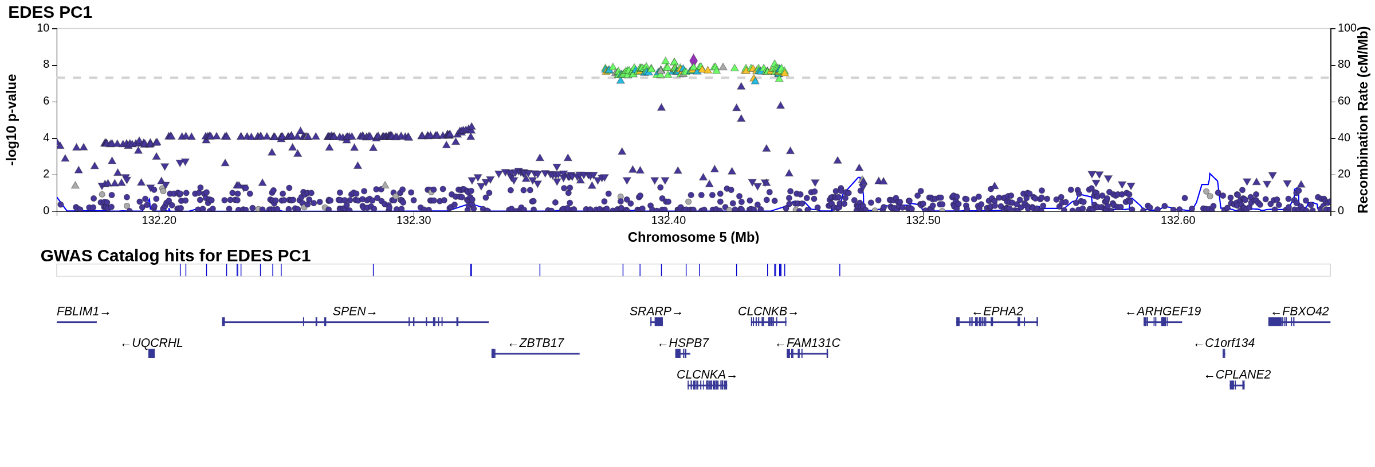

Supplementary Figure 46 – Locus zoom rs11786896 PC1

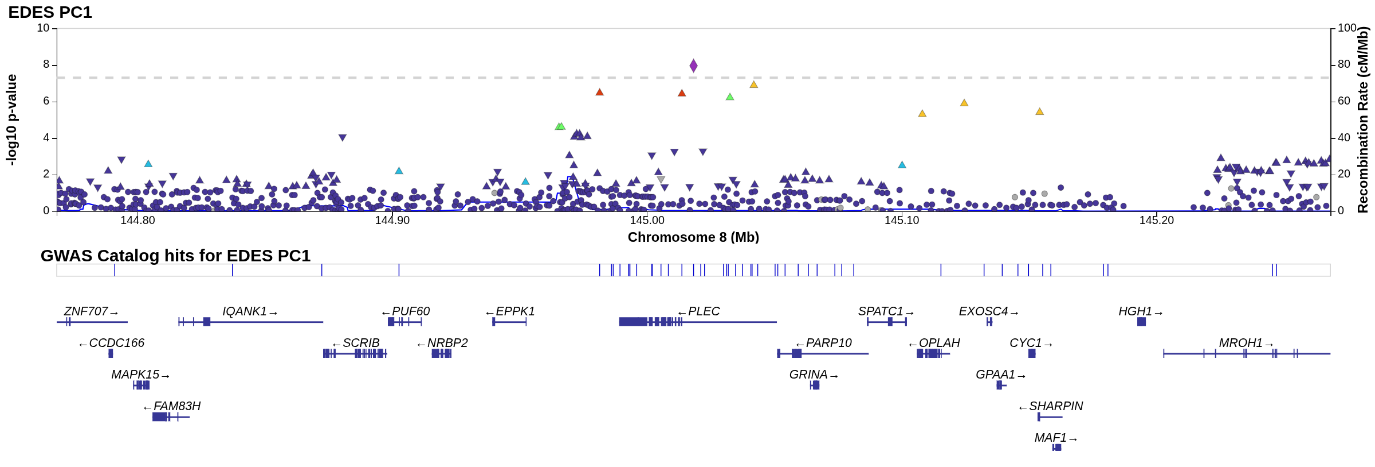

Supplementary Figure 47 – Locus zoom rs71468663 PC1

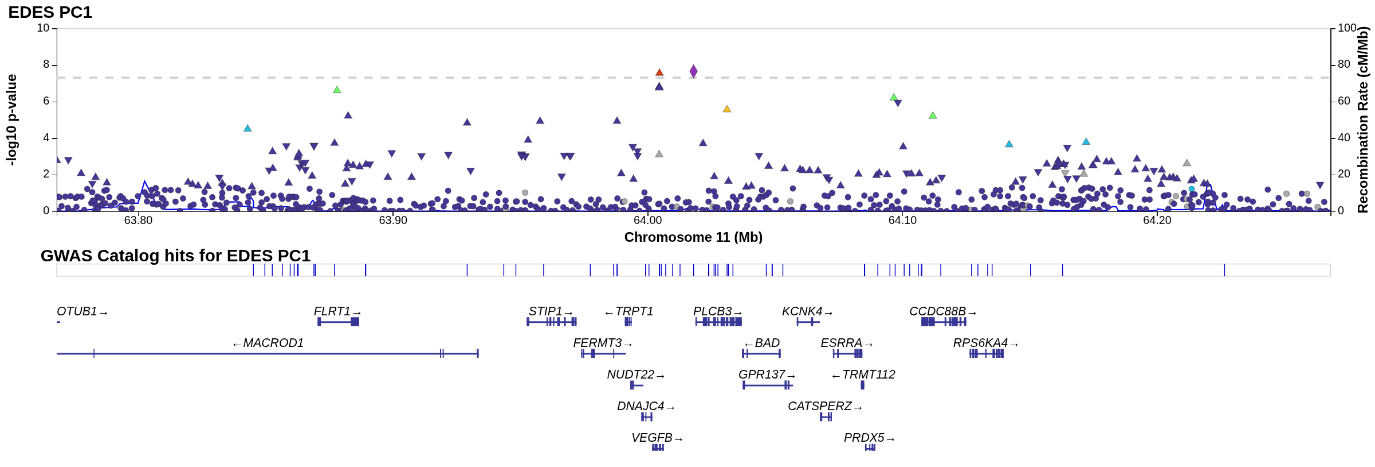

Supplementary Figure 48 – Locus zoom rs373980643 PC1

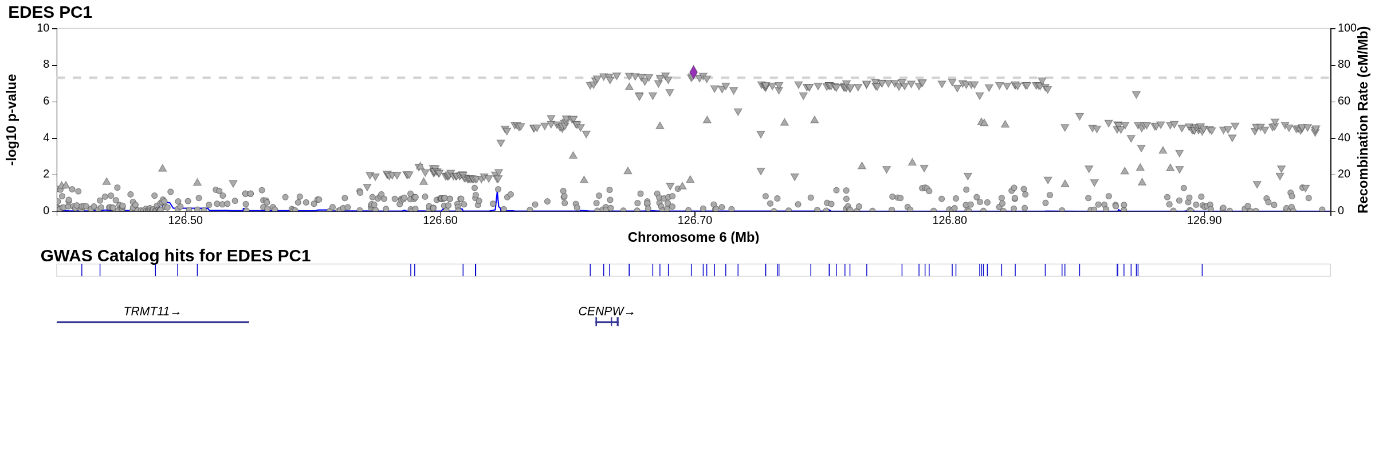

Supplementary Figure 49 – Locus zoom rs3791679 PC2

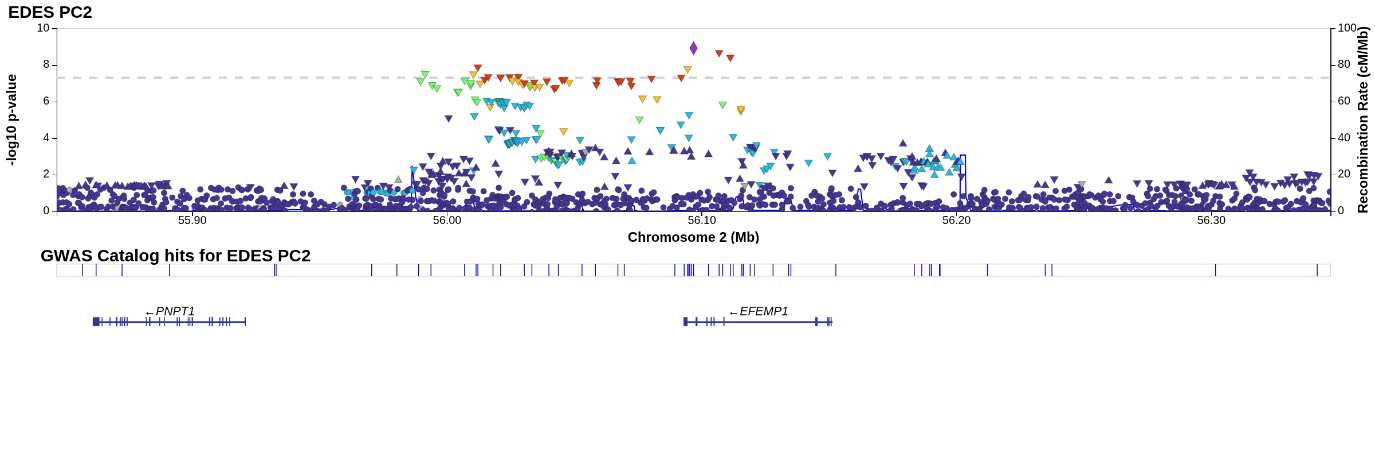

Supplementary Figure 50 – Locus zoom rs11066320 PC2

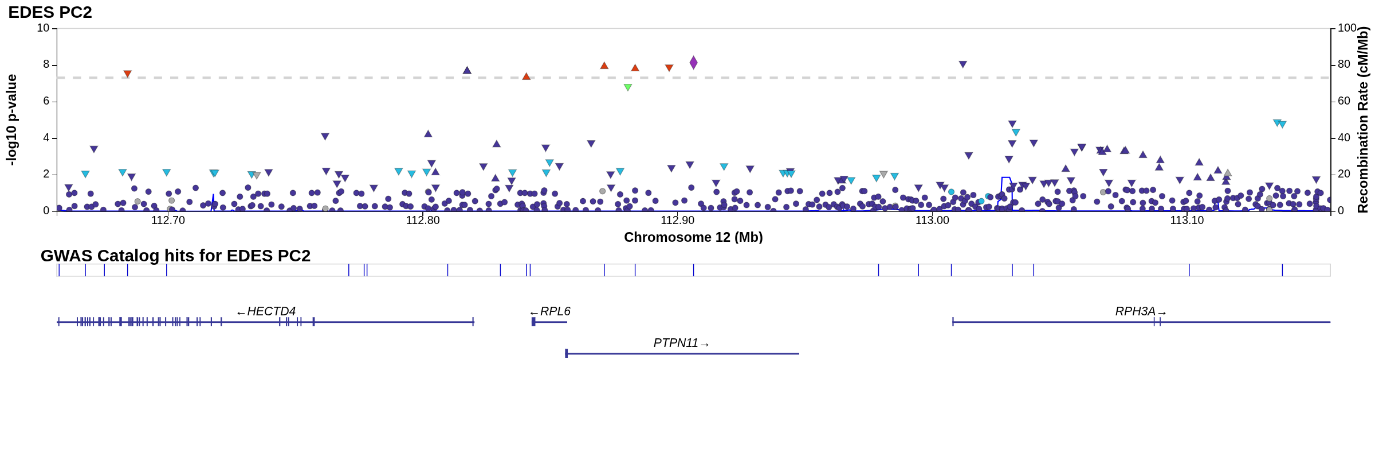

Supplementary Figure 51 – Locus zoom rs1890753 PC3

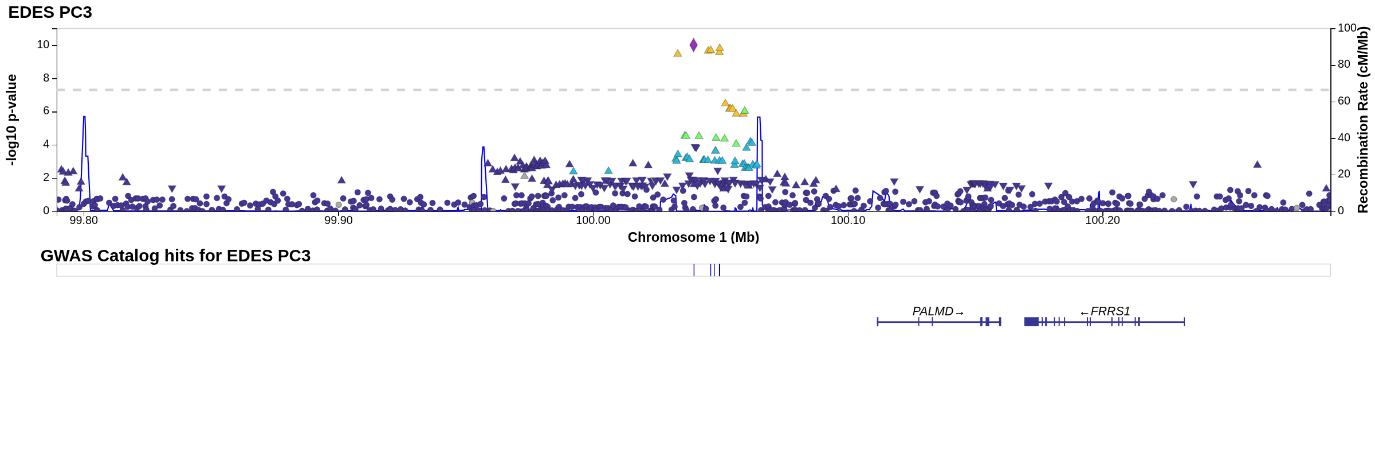

Supplementary Figure 52 – Locus zoom rs332659 PC3

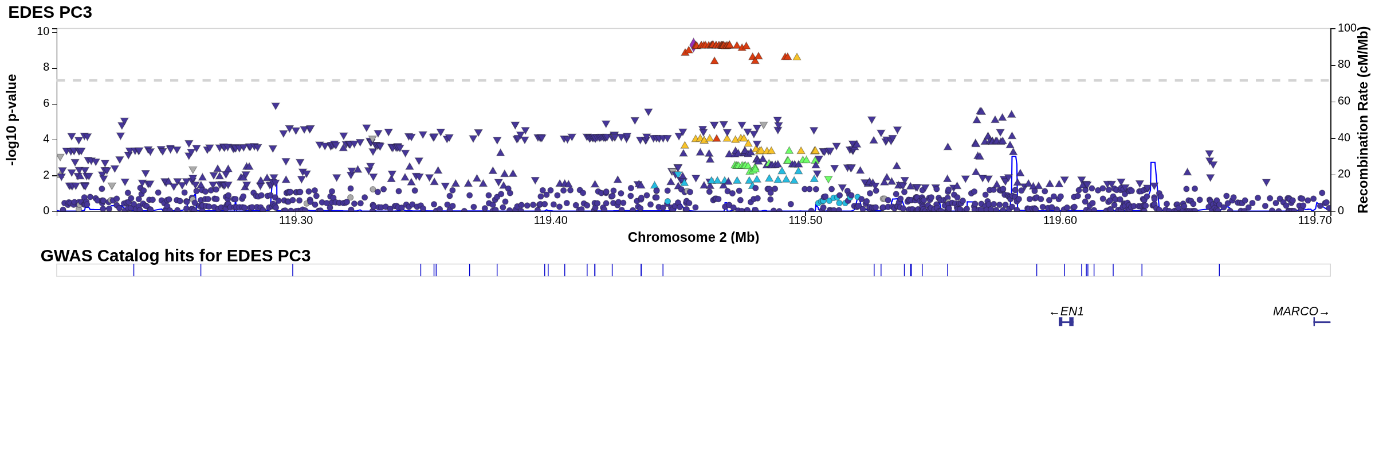

Supplementary Figure 53 – Locus zoom rs10155248 PC3

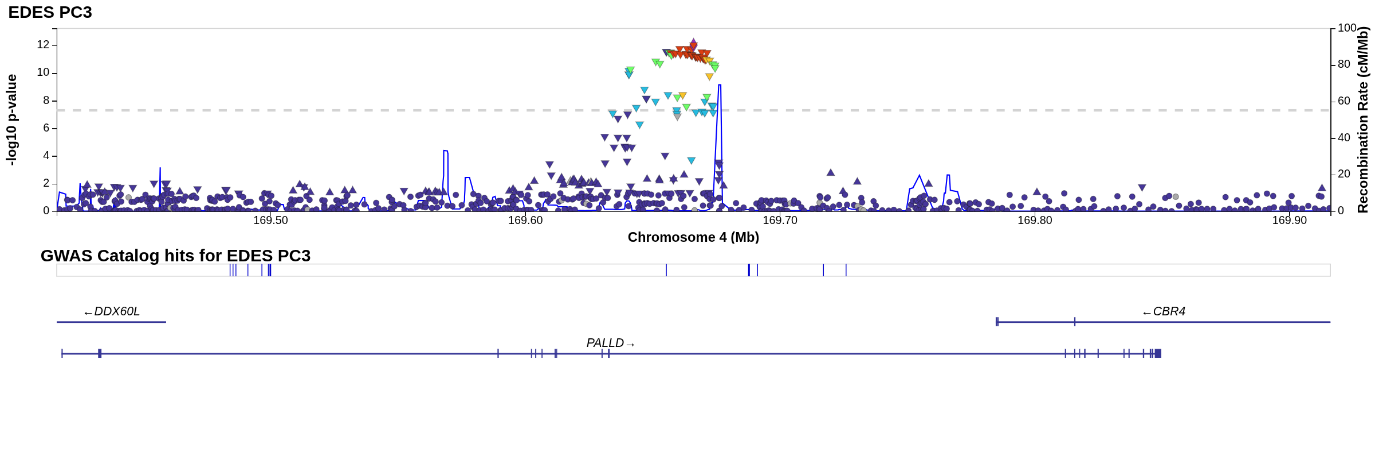

Supplementary Figure 54 – Locus zoom rs10077410 PC3

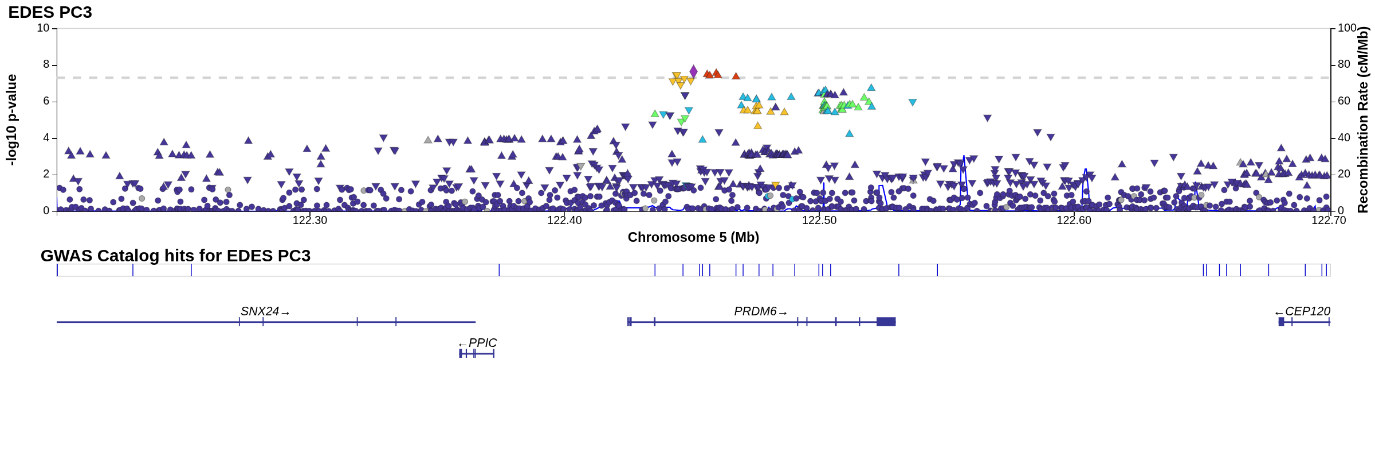

Supplementary Figure 55 – Locus zoom rs3176326 PC3

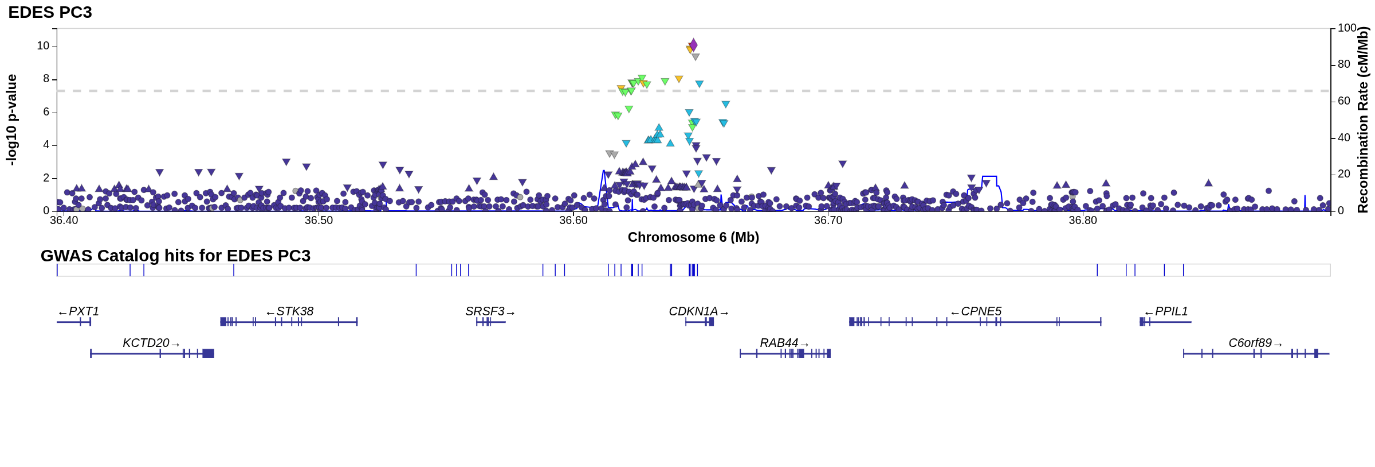

Supplementary Figure 56 – Locus zoom rs11190360 PC3

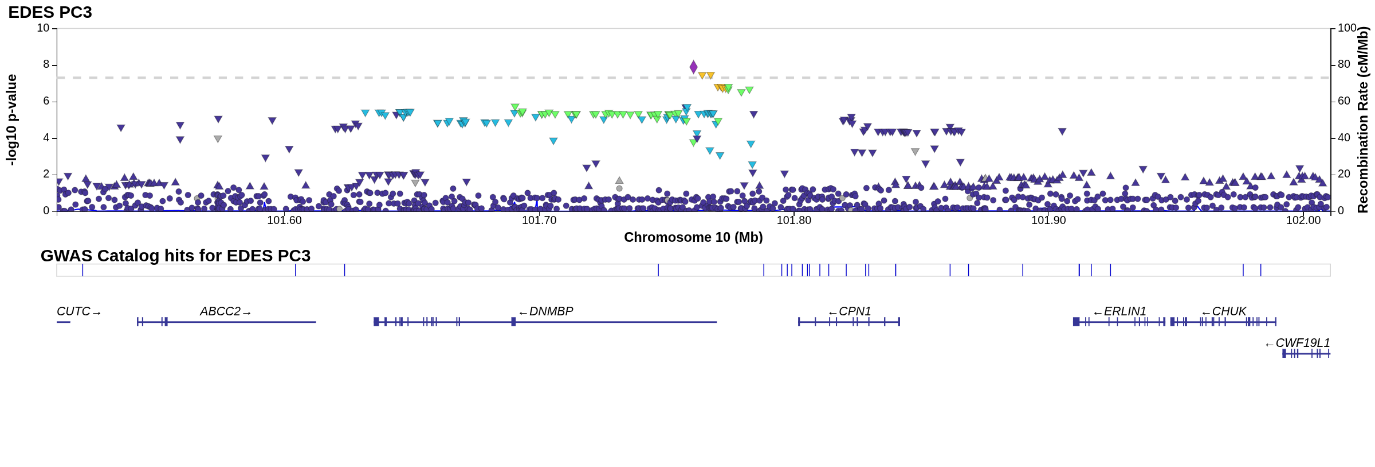

Supplementary Figure 57 – Locus zoom rs10749139 PC3

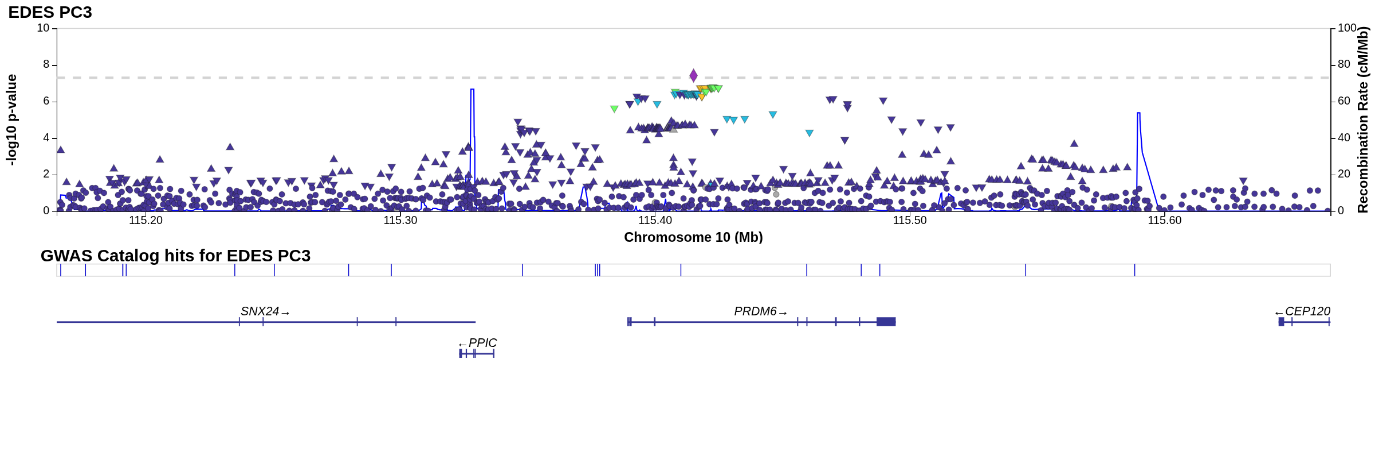

Supplementary Figure 58 – Locus zoom rs112716157	PC3

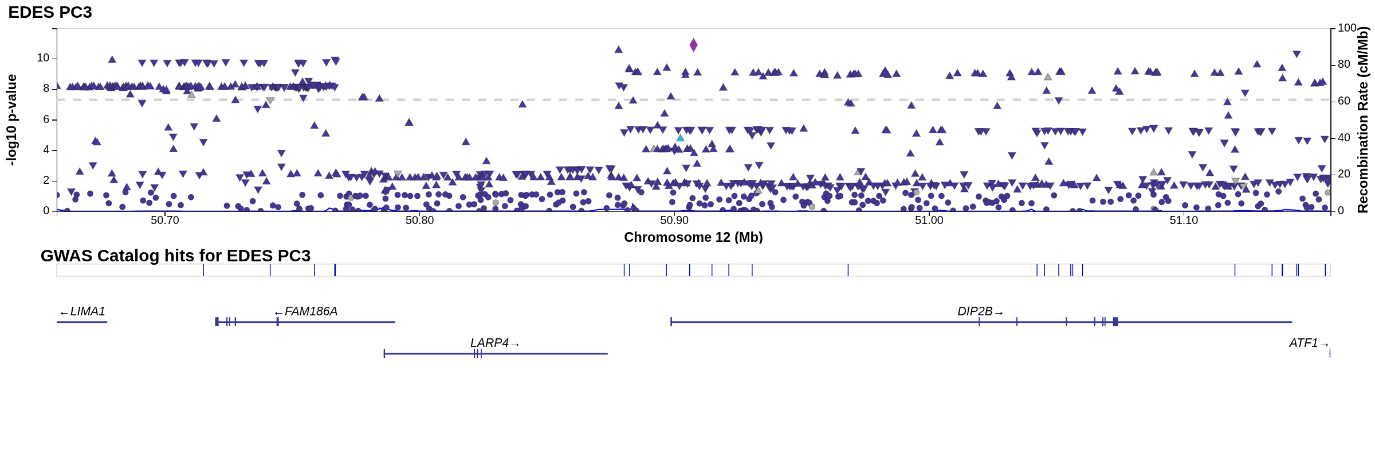

Supplementary Figure 59 – Locus zoom rs4144499 PC3

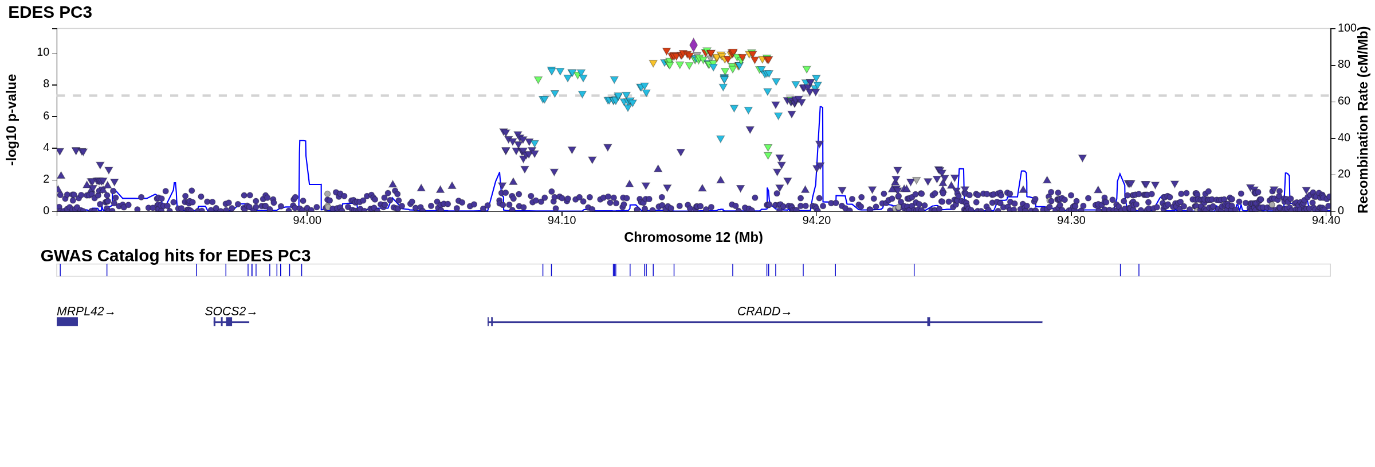

Supplementary Figure 60 – Locus zoom rs17608766 PC3

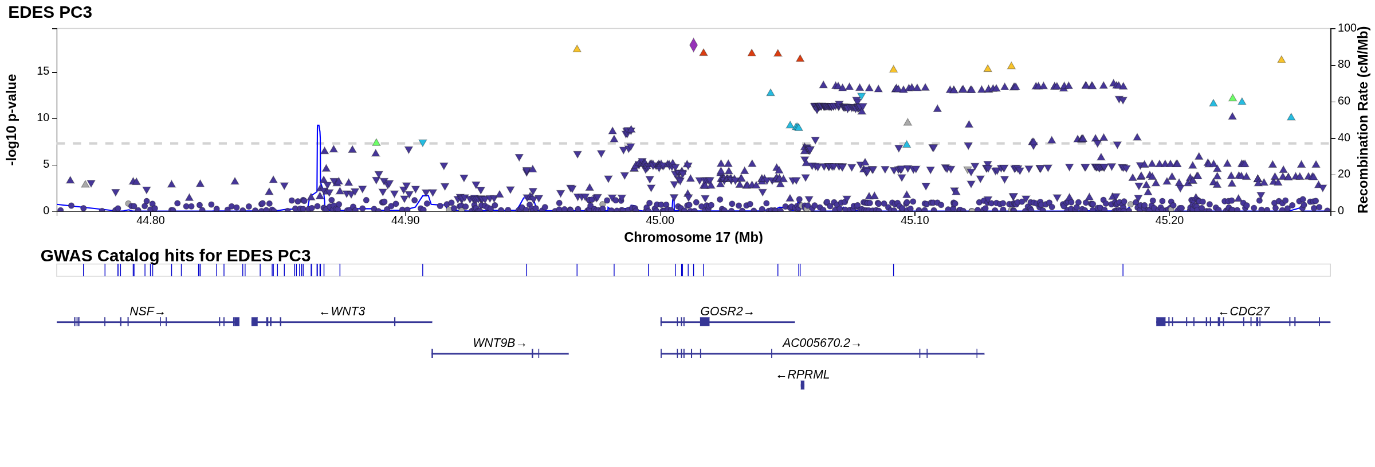

Supplementary Figure 61 – Locus zoom rs71366902 PC3

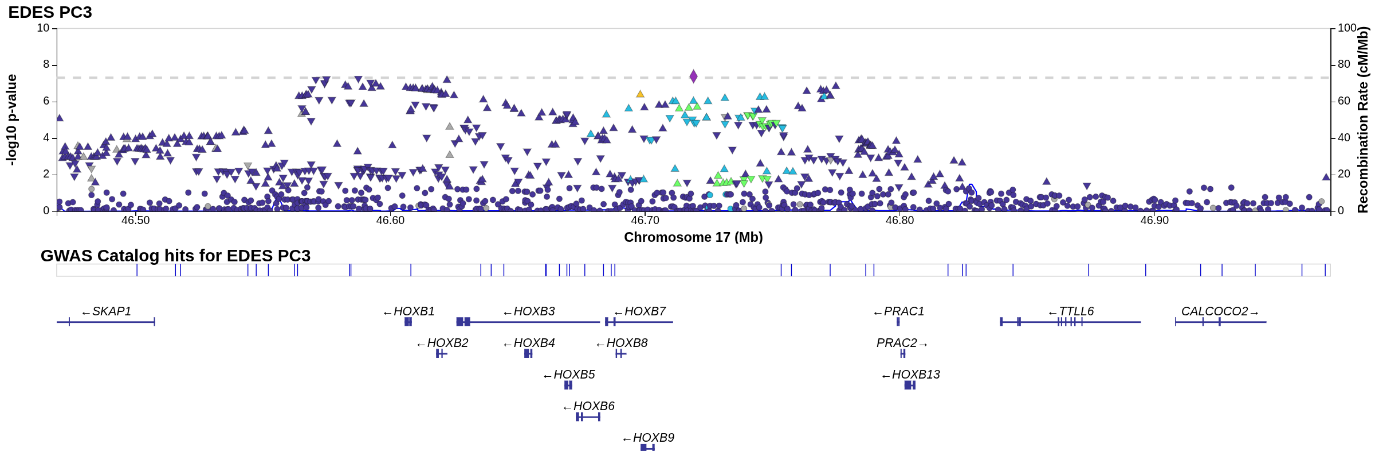

Supplementary Figure 62 – Locus zoom rs186749 PC4
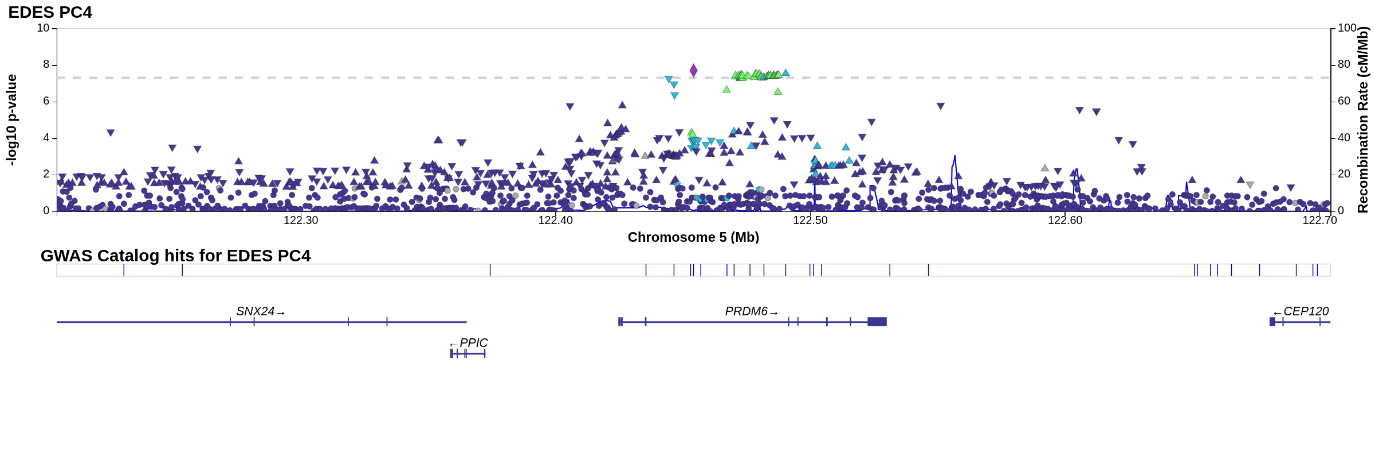

Supplementary Figure 63 – Locus zoom rs12494109 PC5

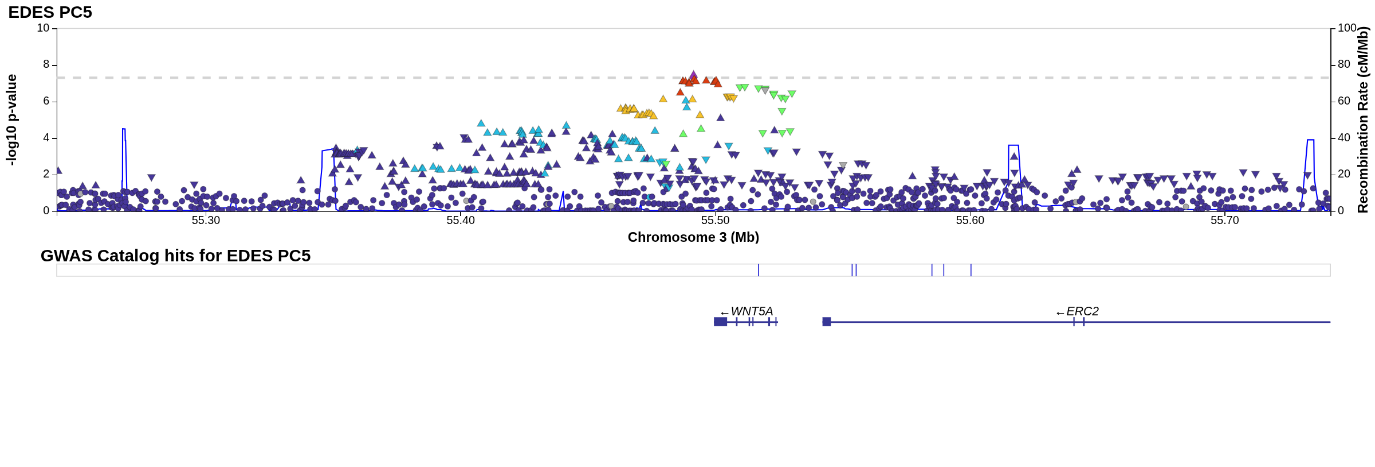

Supplementary Figure 64 – Locus zoom rs12502027 PC5

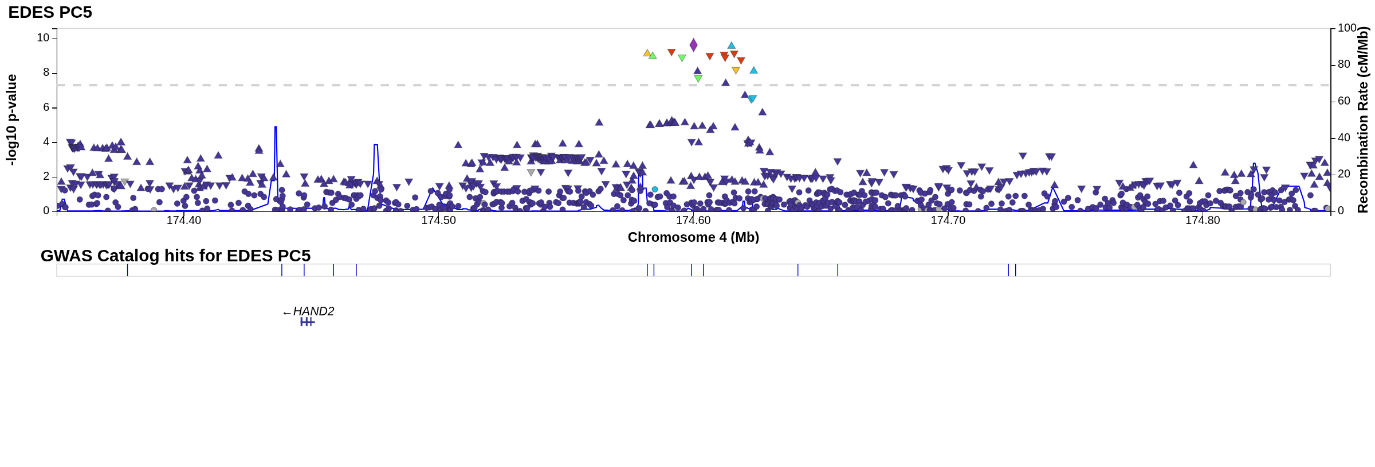

Supplementary Figure 65 – Locus zoom rs398028 PC5

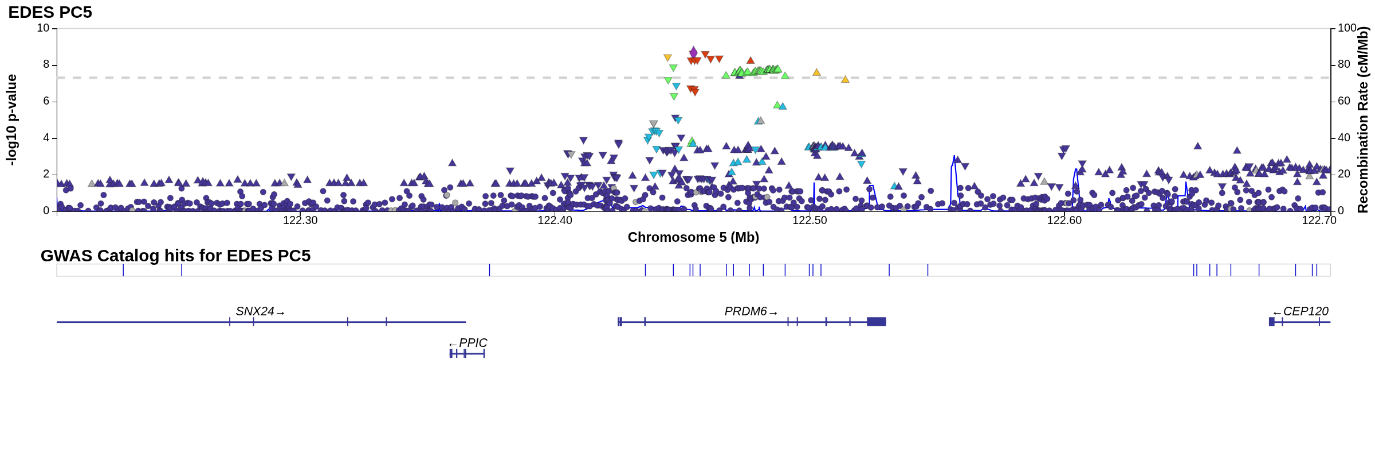

Supplementary Figure 66 – Locus zoom rs2284174 PC5

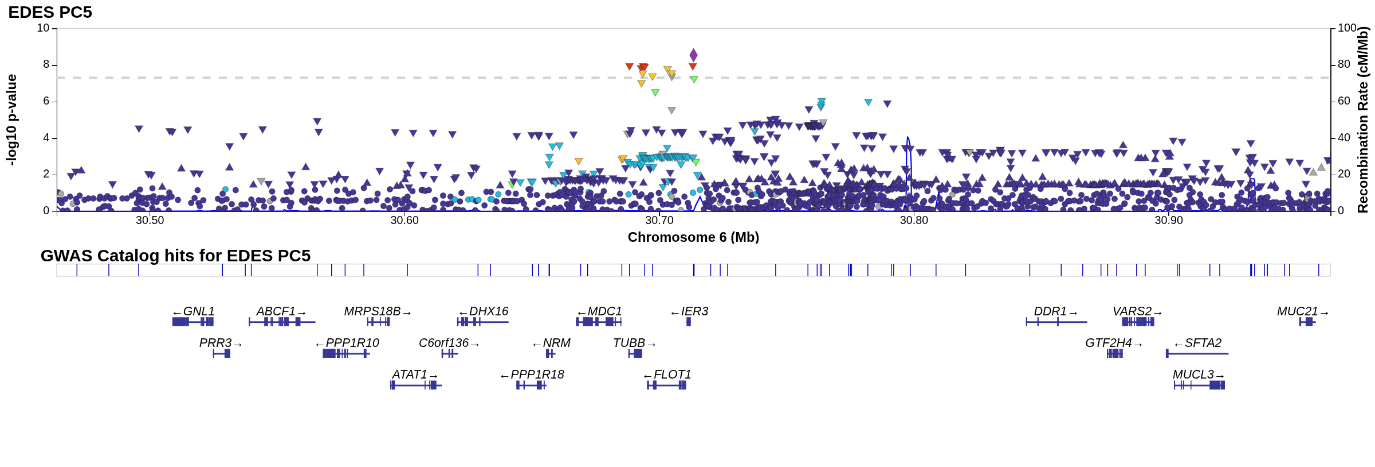

Supplementary Figure 67 – Locus zoom 6:31588020_CCTCT_C	PC5

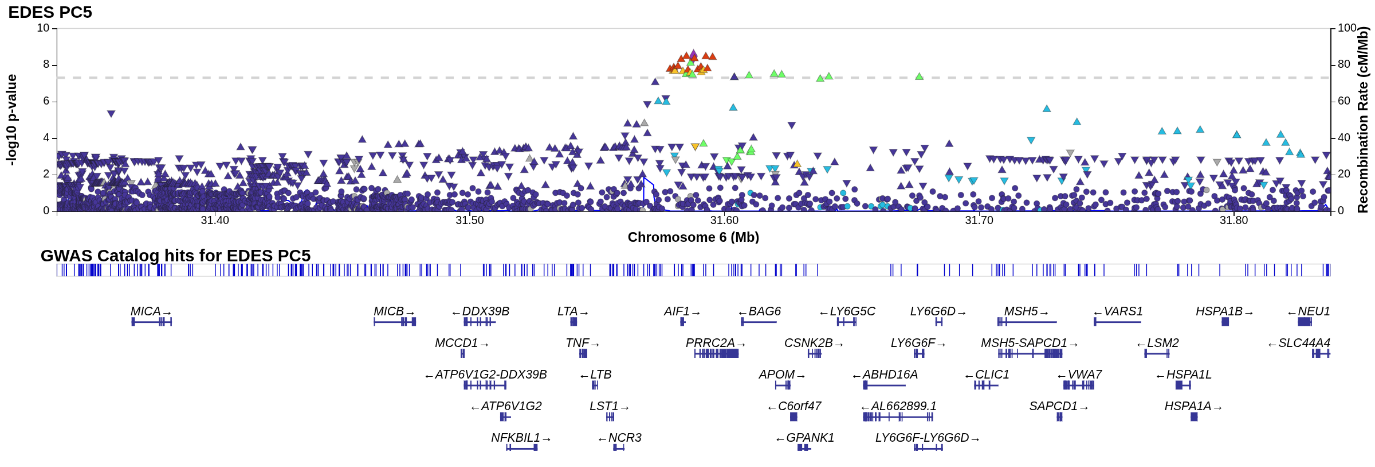

Supplementary Figure 68 – Locus zoom rs2638950 PC5

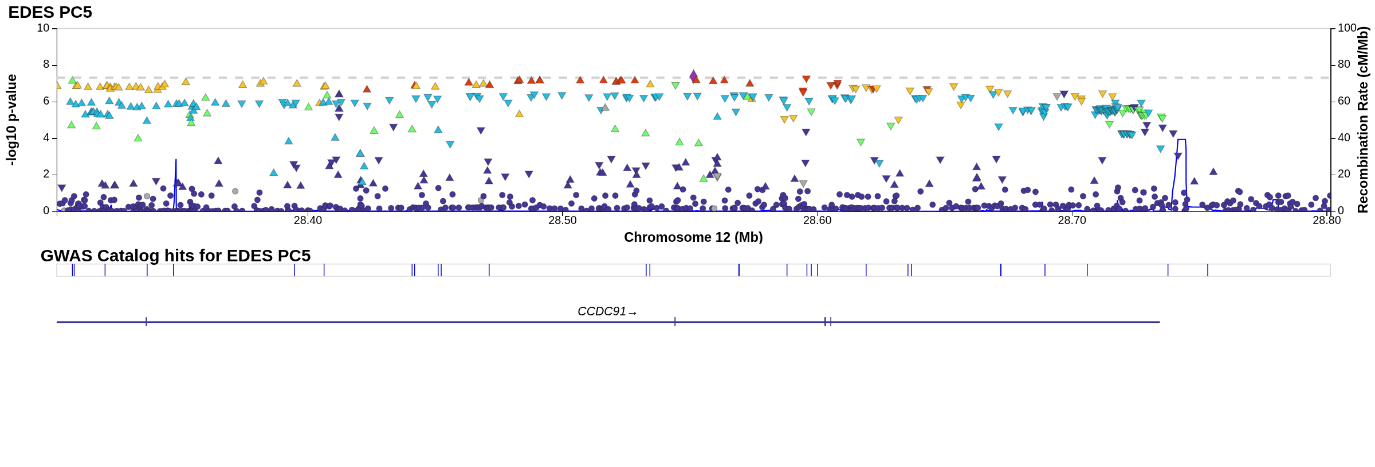

Supplementary Figure 69 – Locus zoom rs6602994 PC5

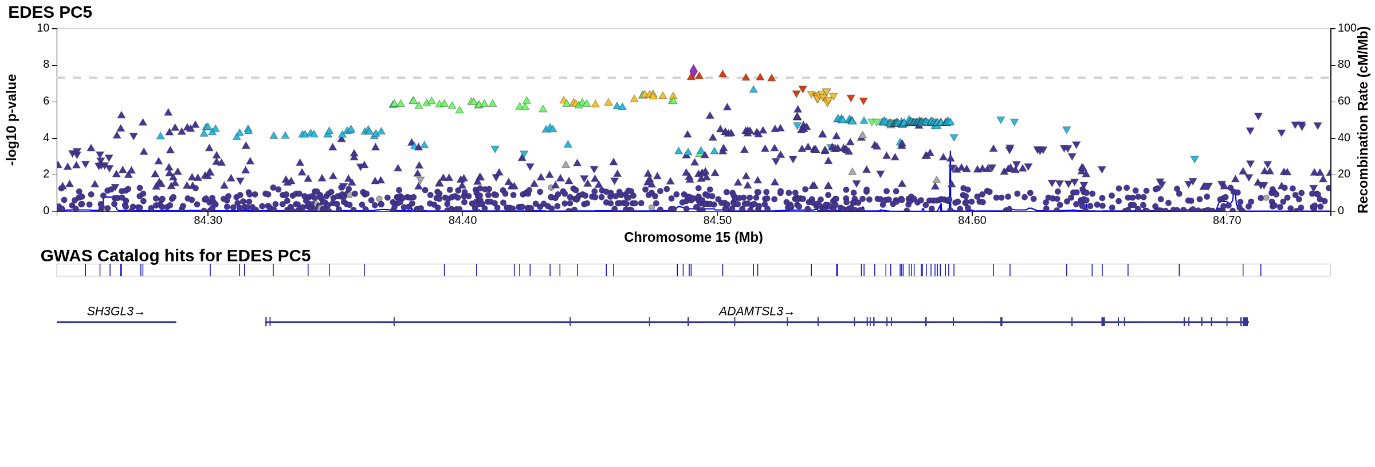

Supplementary Figure 70 – Locus zoom rs201699038 PC6

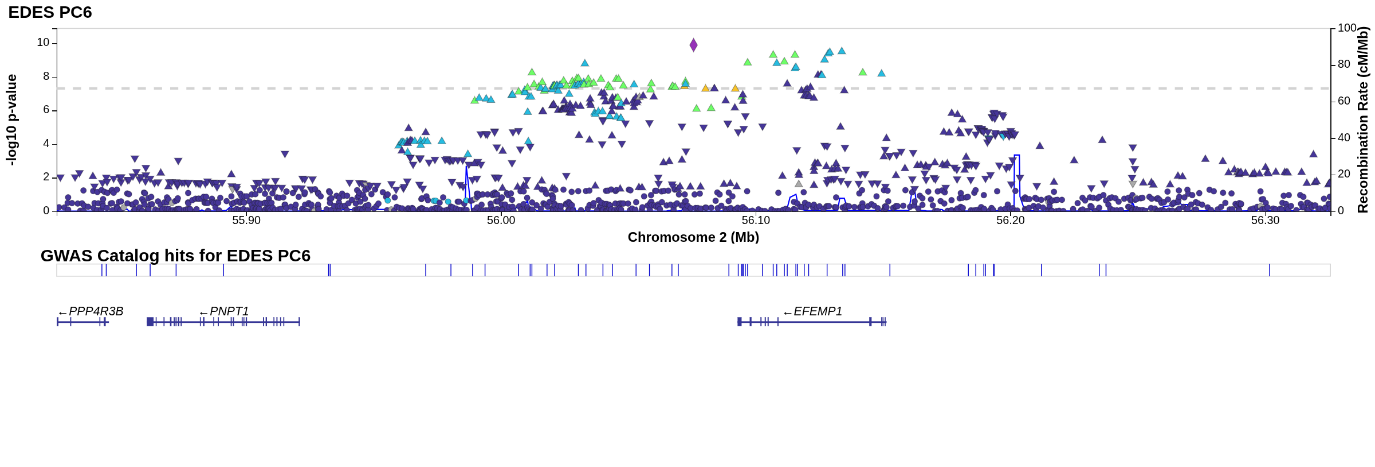

Supplementary Figure 71 – Locus zoom rs11710570 PC6

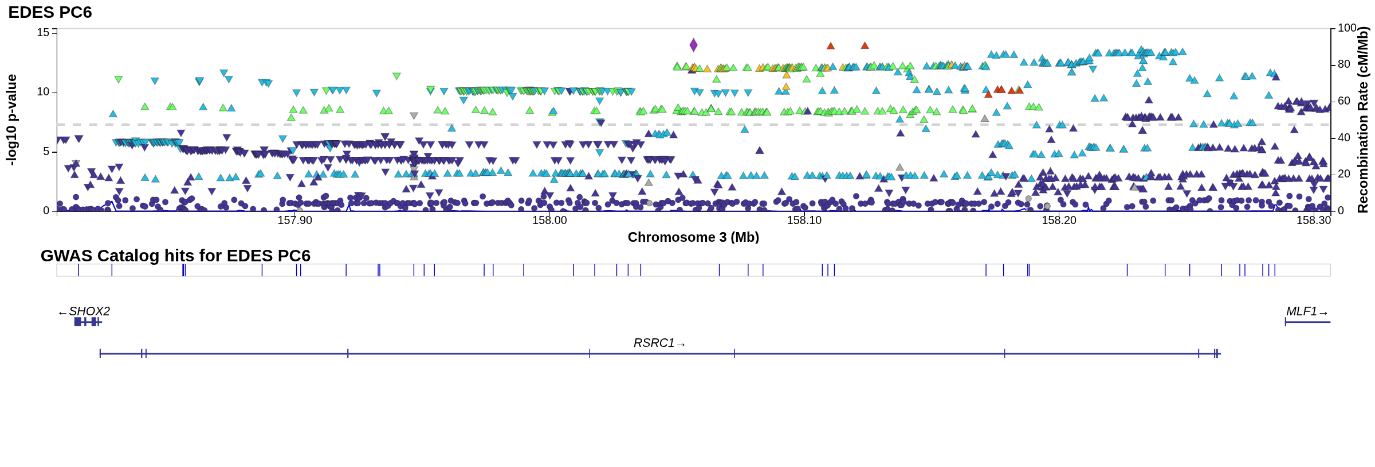

Supplementary Figure 72 – Locus zoom rs11756440 PC6

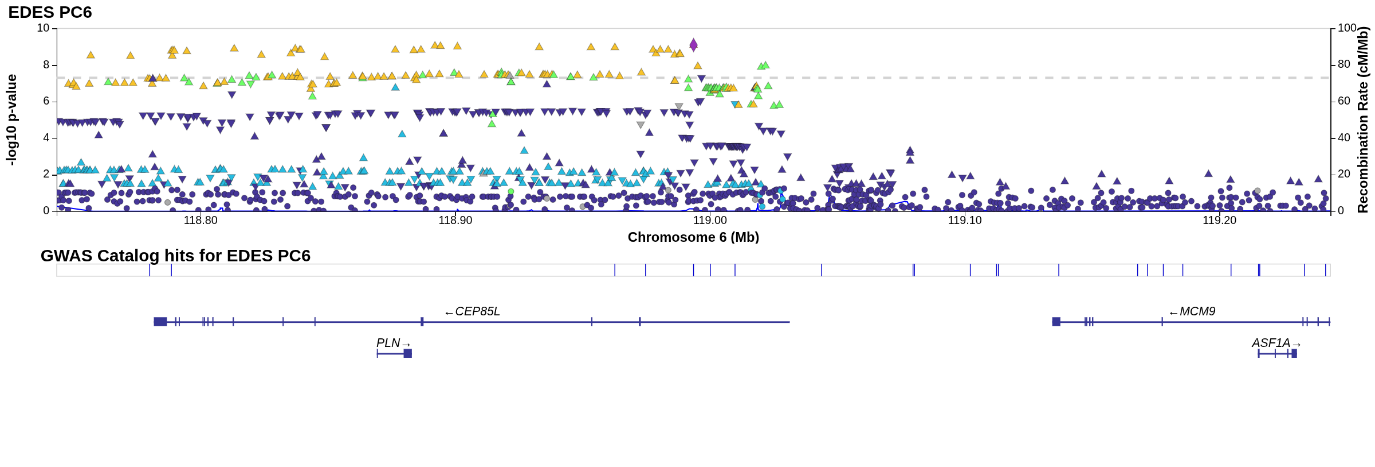

Supplementary Figure 73 – Locus zoom rs162185 PC6

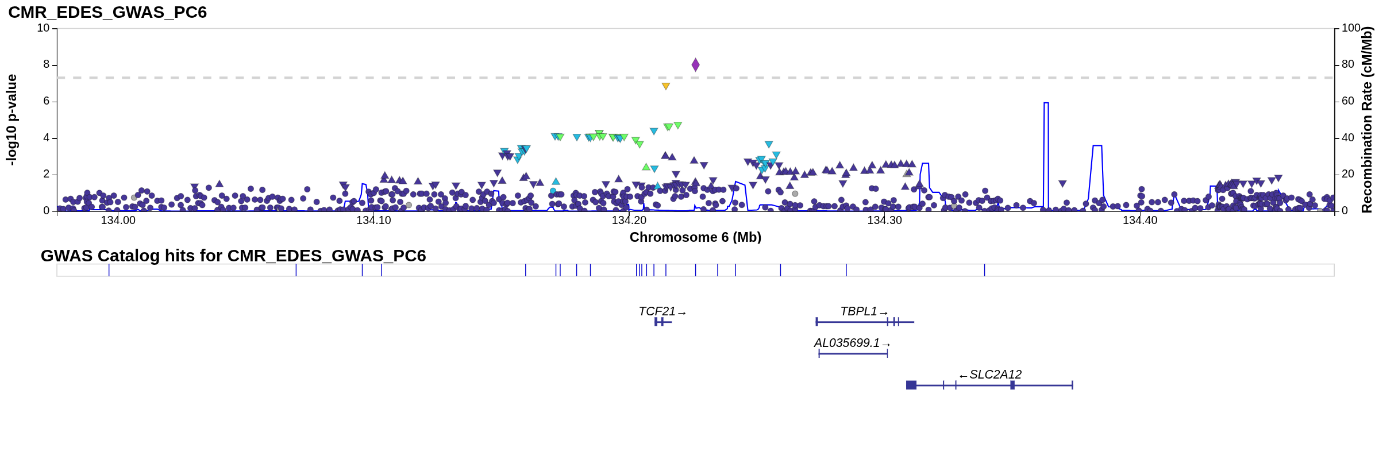

Supplementary Figure 74 – Locus zoom rs12531355 PC6

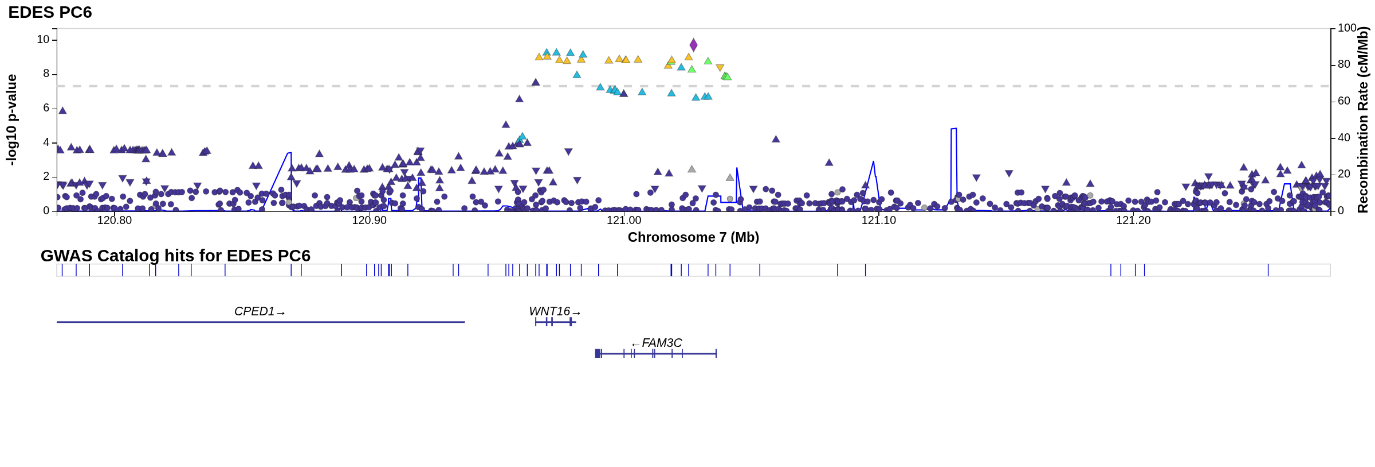

Supplementary Figure 75 – Locus zoom rs62526480 PC6

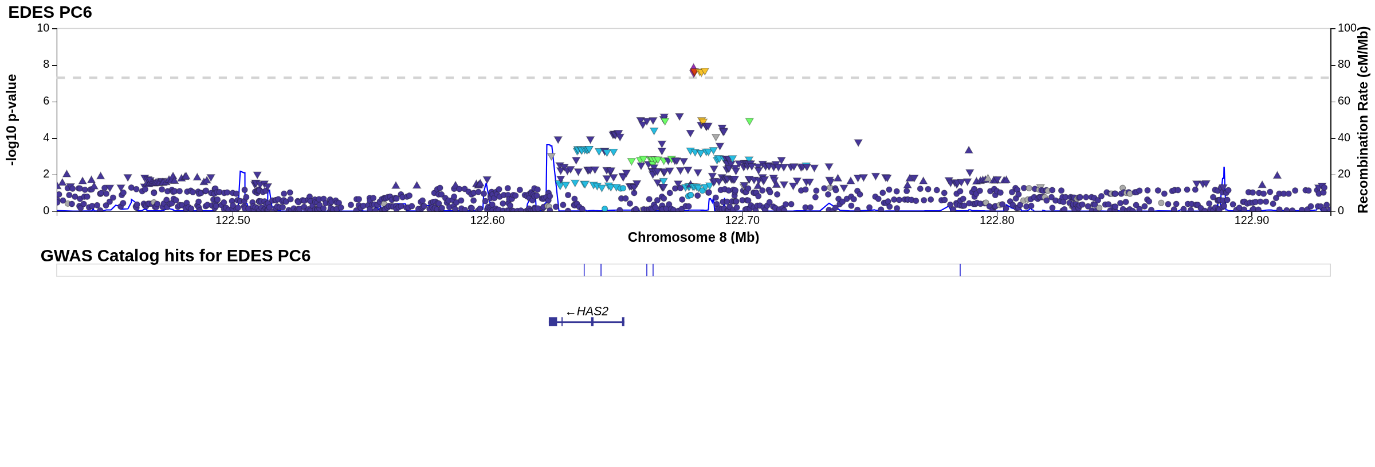

Supplementary Figure 76 – Locus zoom rs12223220 PC6

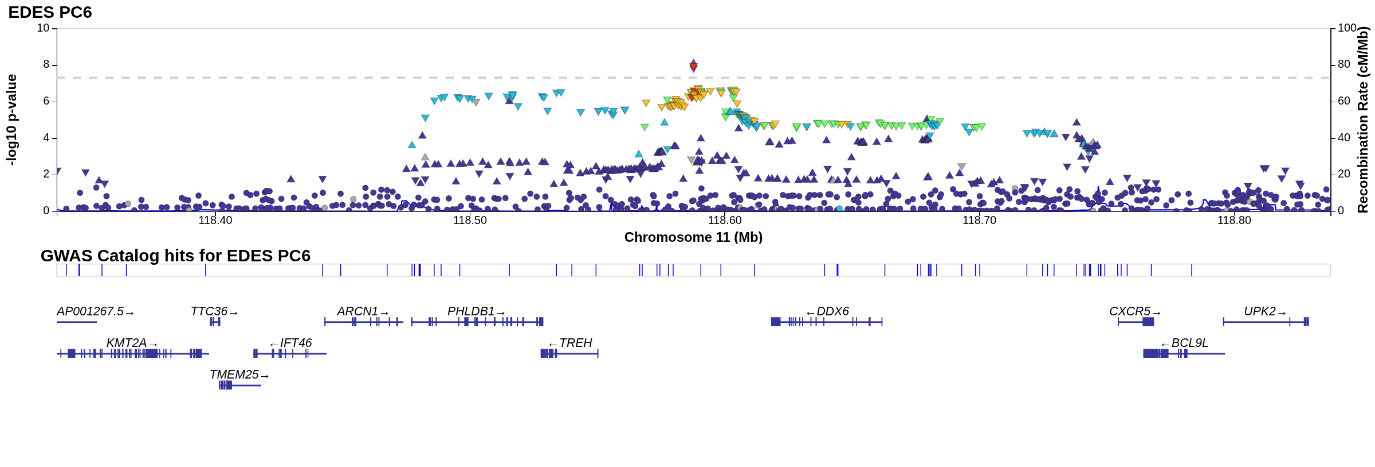

Supplementary Figure 77 – Locus zoom rs66534382 PC6

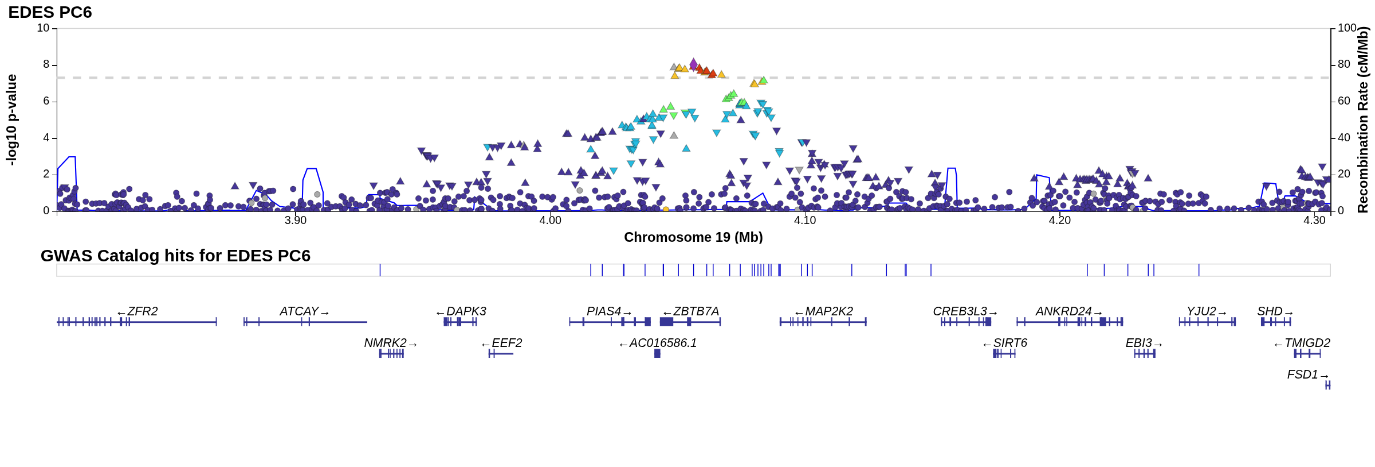

Supplementary Figure 78 – Locus zoom rs531997 PC7

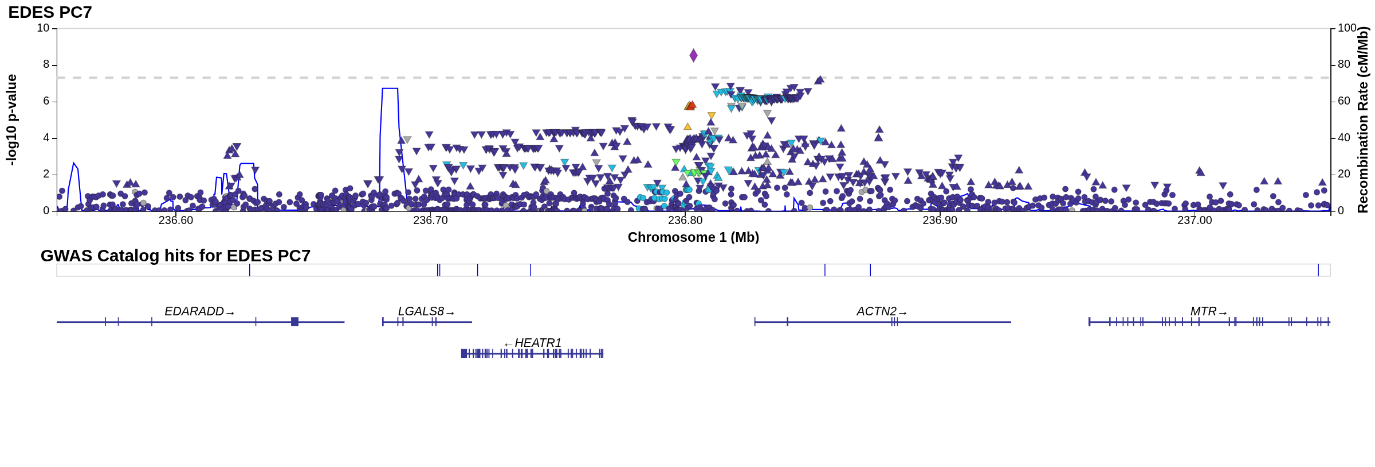

Supplementary Figure 79 – Locus zoom rs32841 PC7

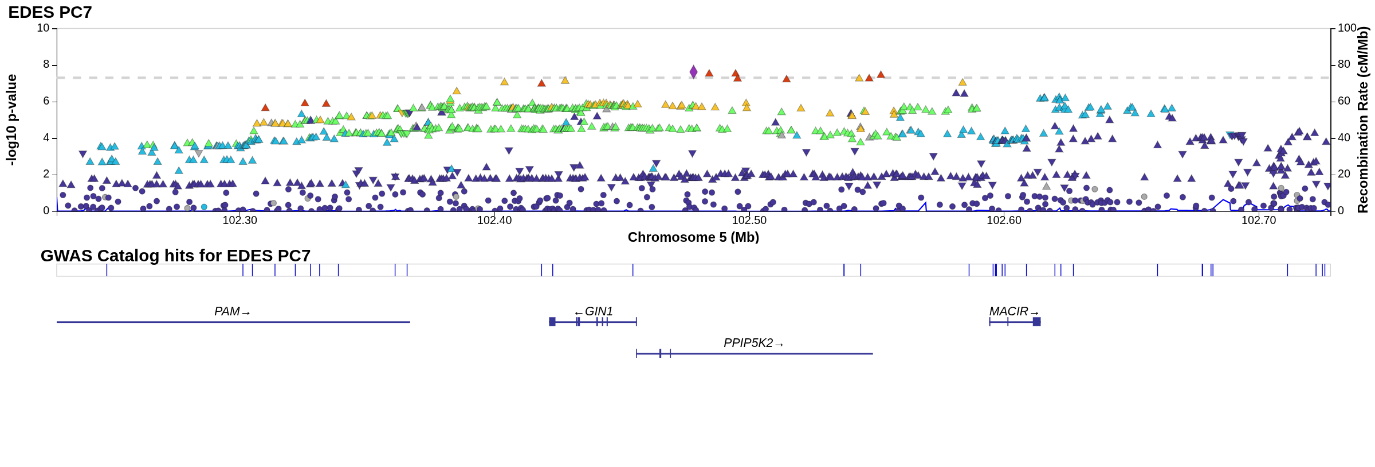

Supplementary Figure 80 – Locus zoom rs10850409 PC7

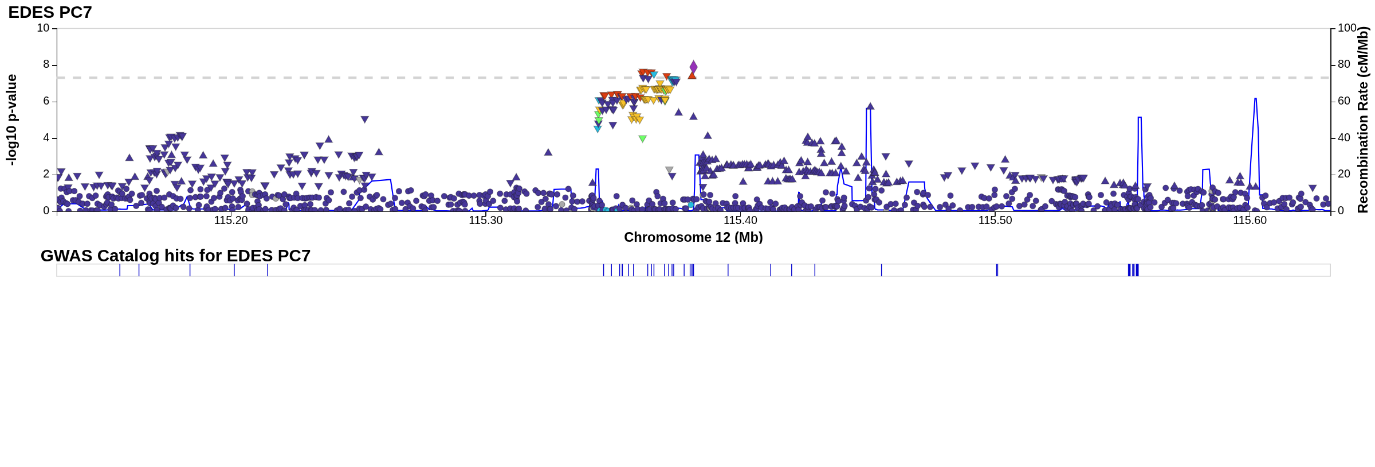

Supplementary Figure 81 – Locus zoom rs151091024 PC8

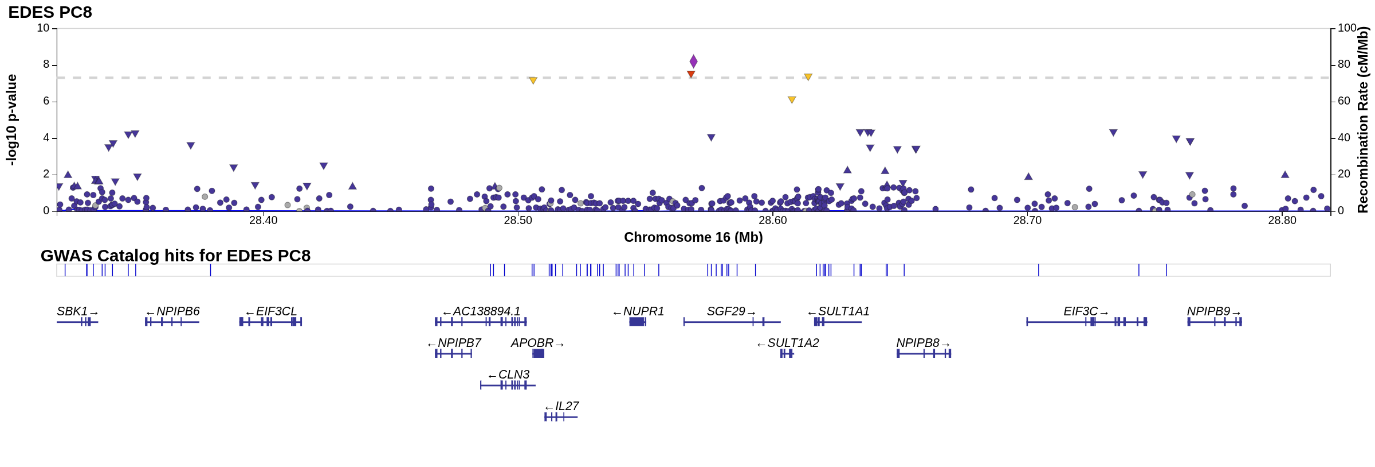

Supplementary Figure 82 – Locus zoom rs12939170 PC8

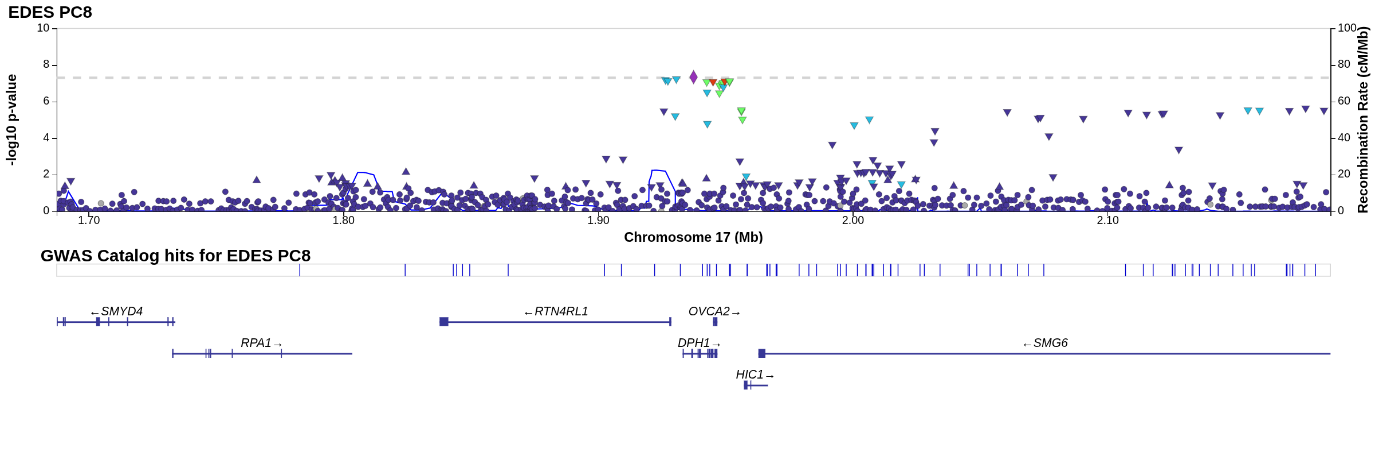

Supplementary Figure 83 – Locus zoom rs6821503 PC9

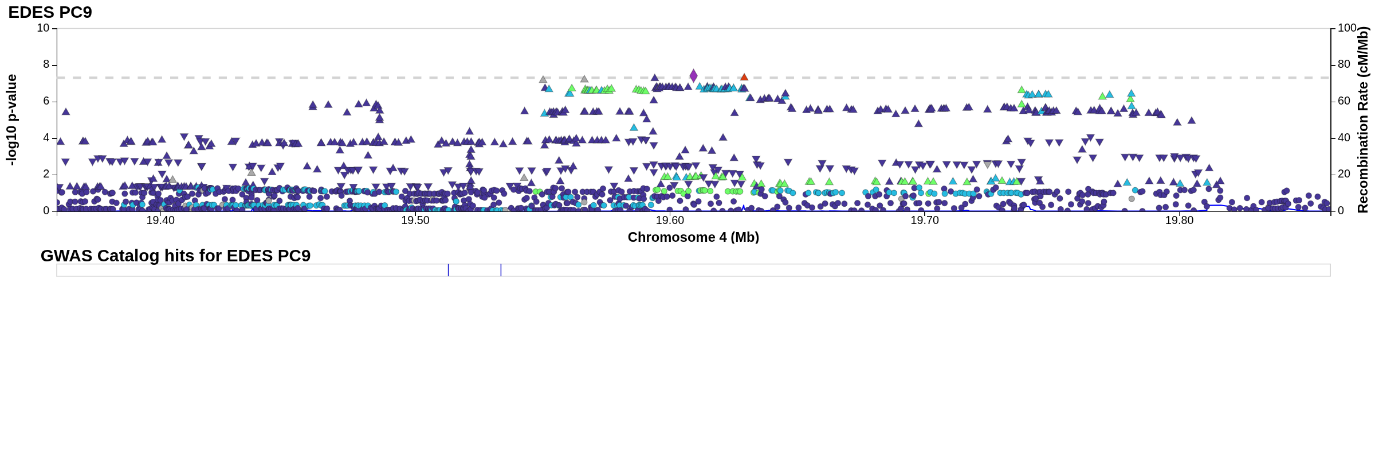

Supplementary Figure 84 – Locus zoom rs7014186 PC9

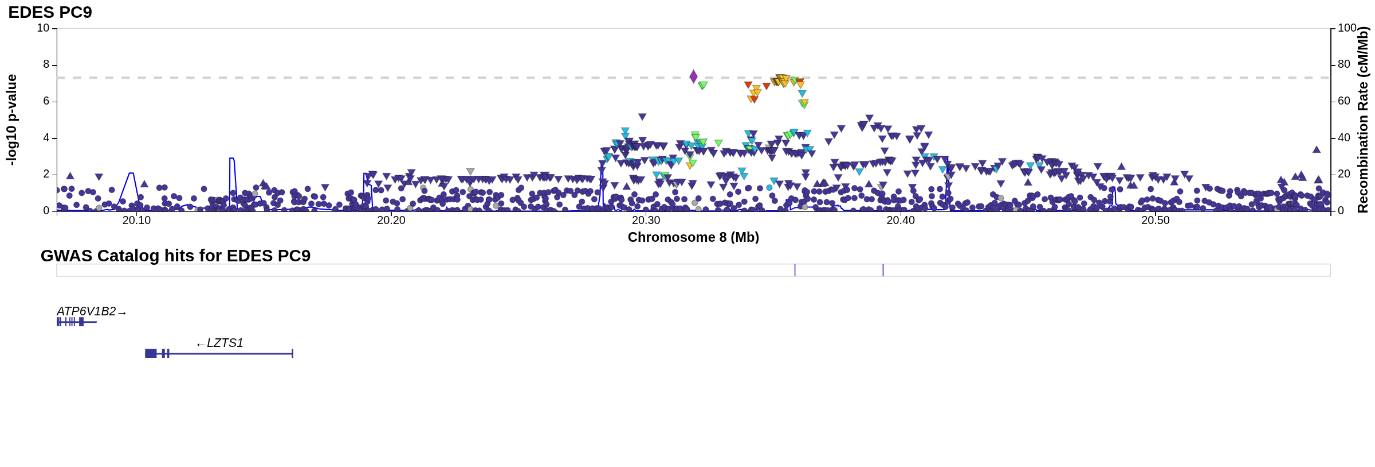

Supplementary Figure 85 – Locus zoom 9:97209762_TG_T PC9

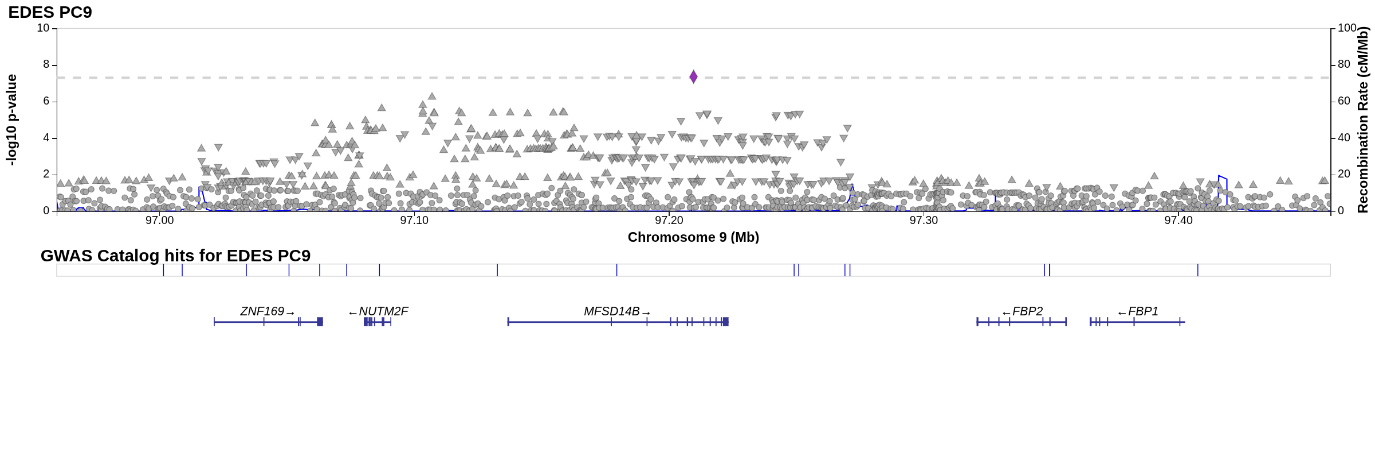

Supplementary Figure 86 – Locus zoom rs422068 PC10

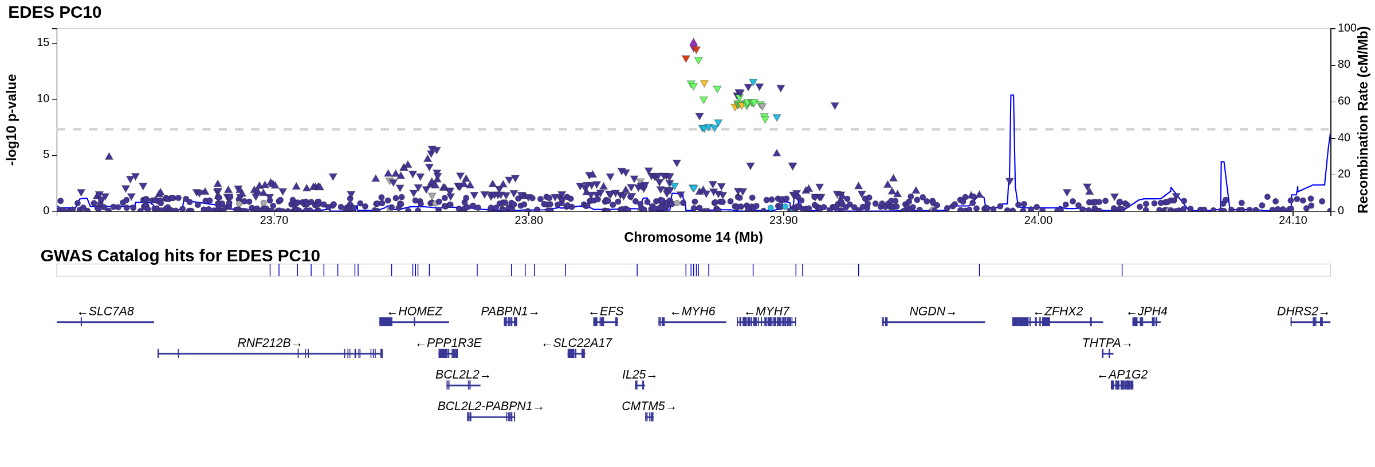

Supplementary Figure 87 – Locus zoom 18:34219777_G_GTT PC10

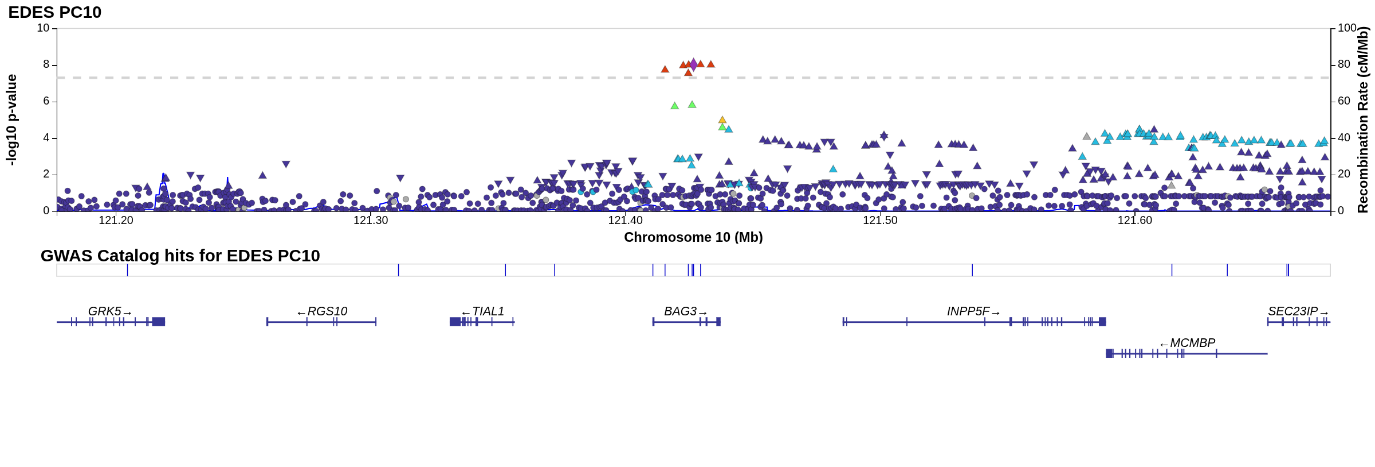

Supplementary Figure 88 – Locus zoom rs17617337 PC10

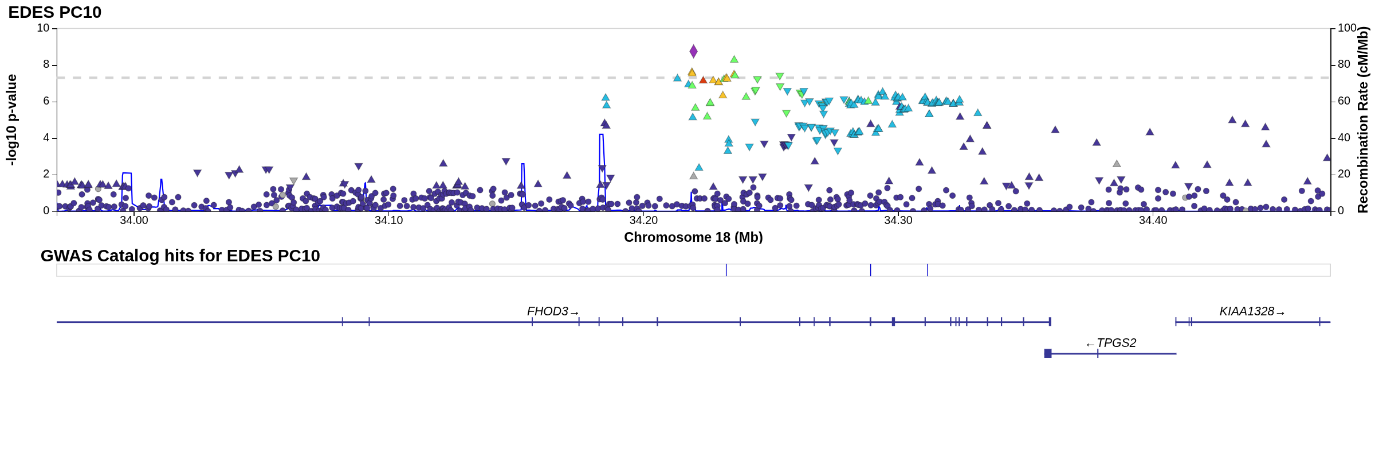

Supplementary Figure 89 – Locus zoom 16:66690988_AT_A PC10

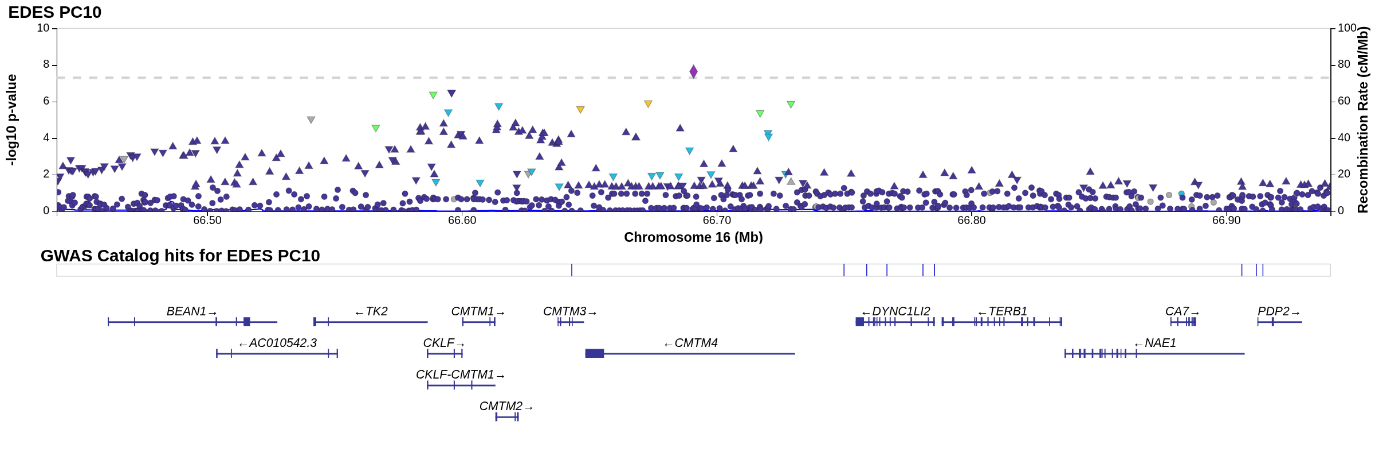

Supplementary Figure 90 – Locus zoom rs3176326 PC11

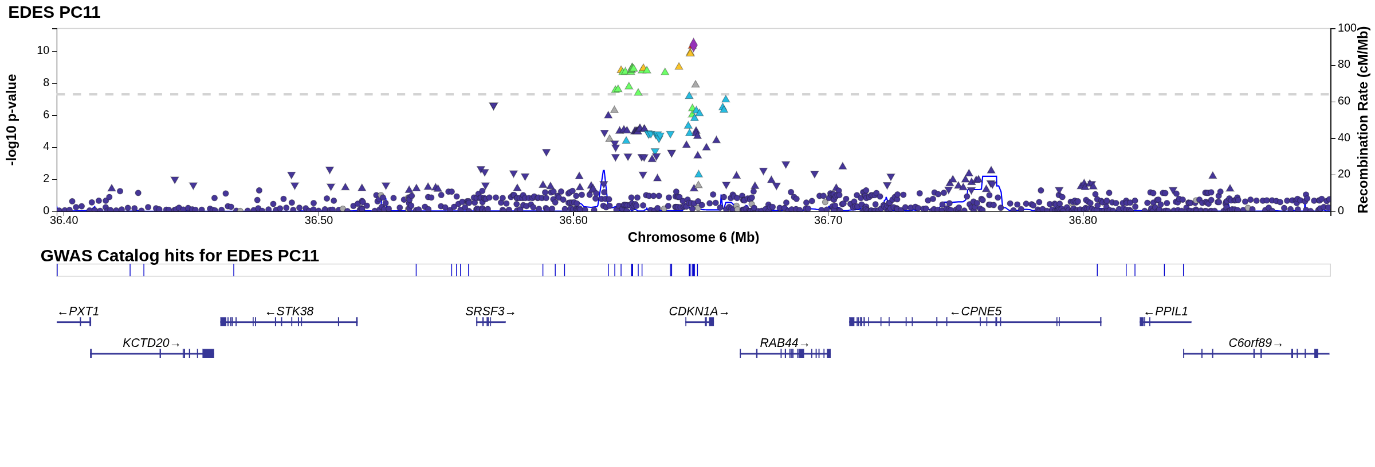

Supplementary Figure 91 – Locus zoom rs138934670 PC11

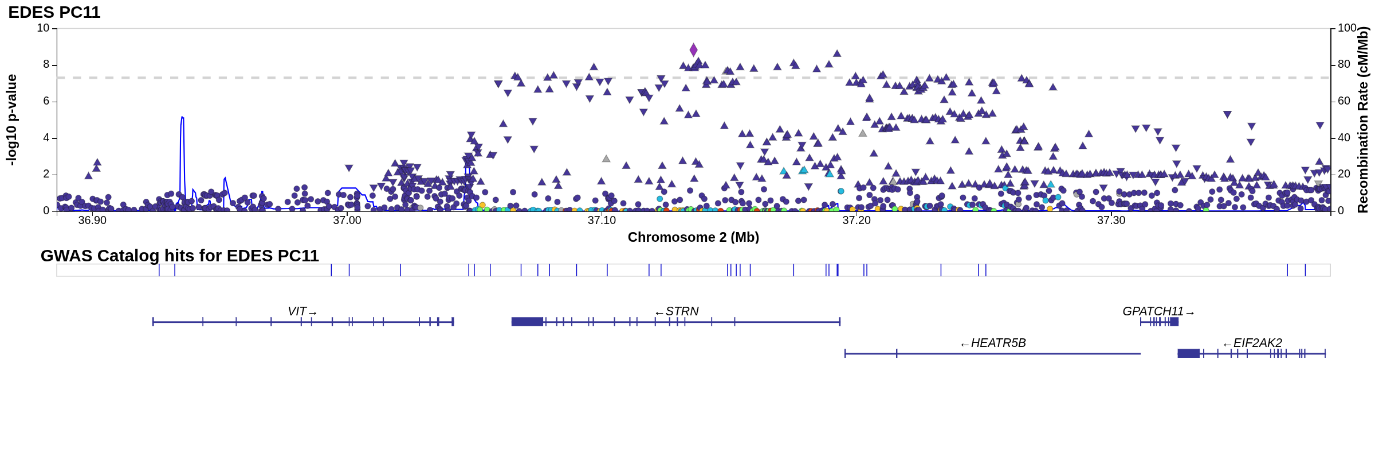

Supplementary Figure 92 – Locus zoom rs9932314 PC11

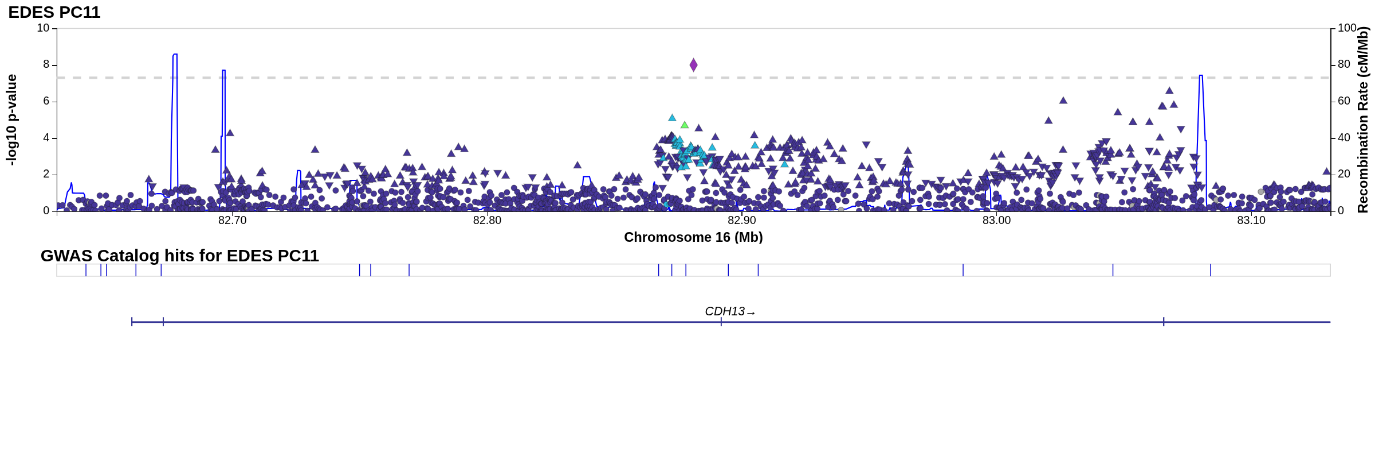

Supplementary Figure 93 – Locus zoom rs60757101 PC11

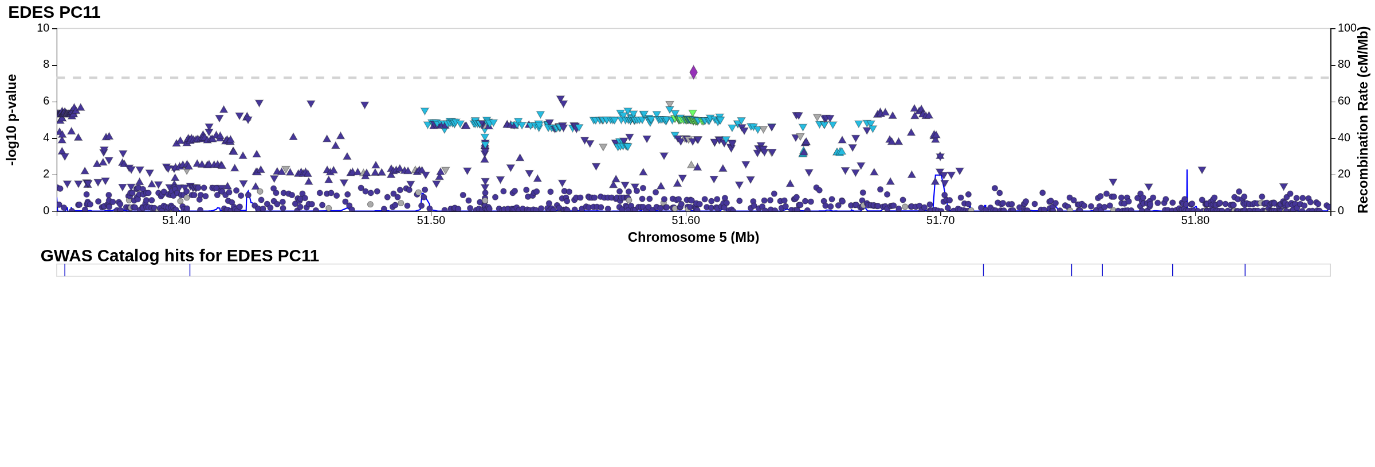

Supplementary Figure 94 – Locus zoom rs77504602 PC11

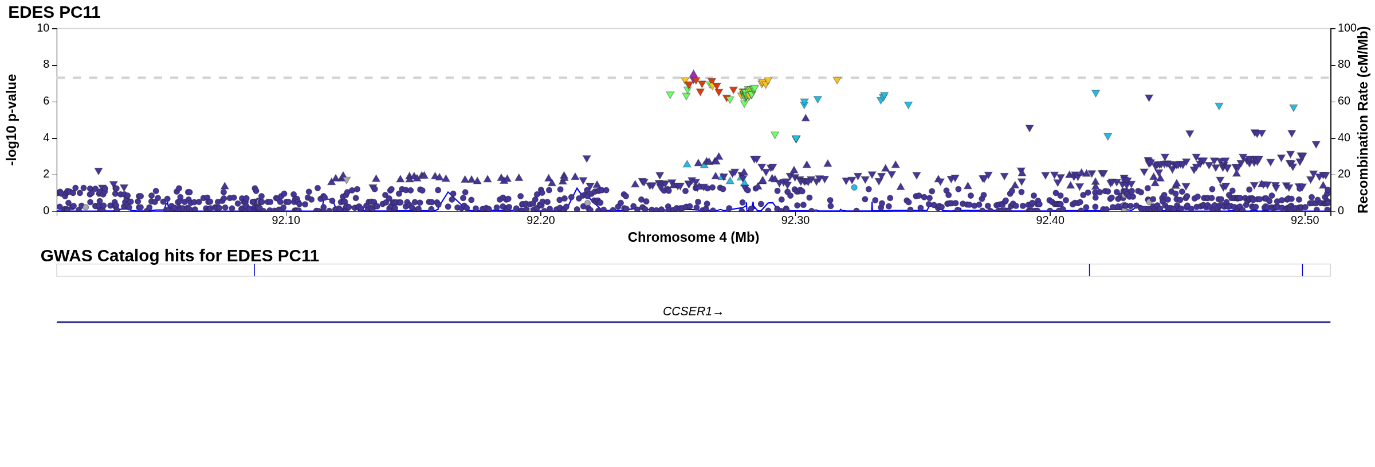

Supplementary Figure 95 – Locus zoom rs13185878 PC12

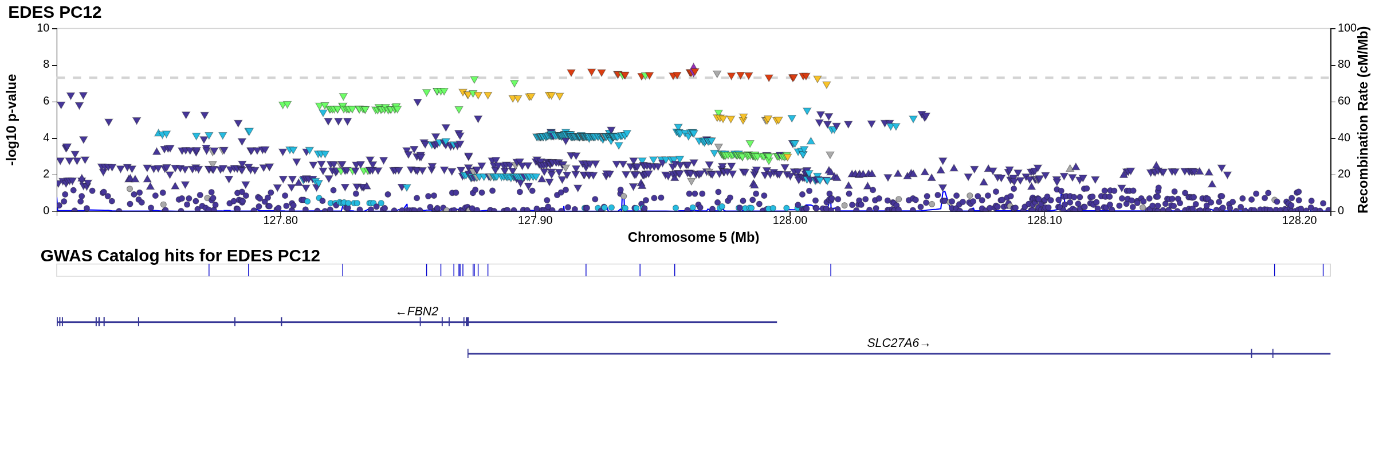

Supplementary Figure 96 – Locus zoom rs1105014 PC12

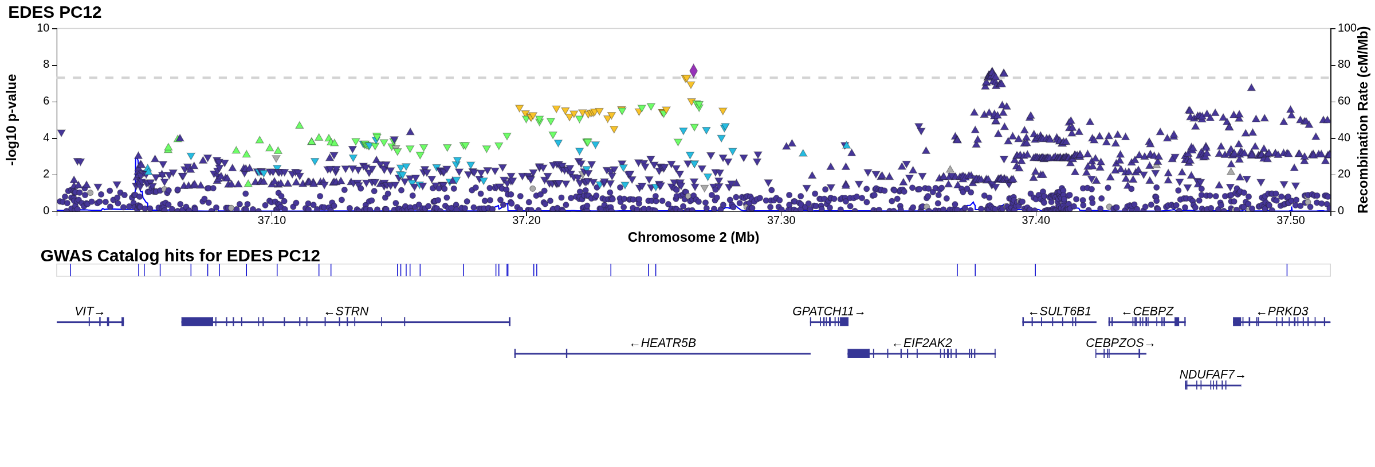

Supplementary Figure 97 – Locus zoom rs2234962 PC13

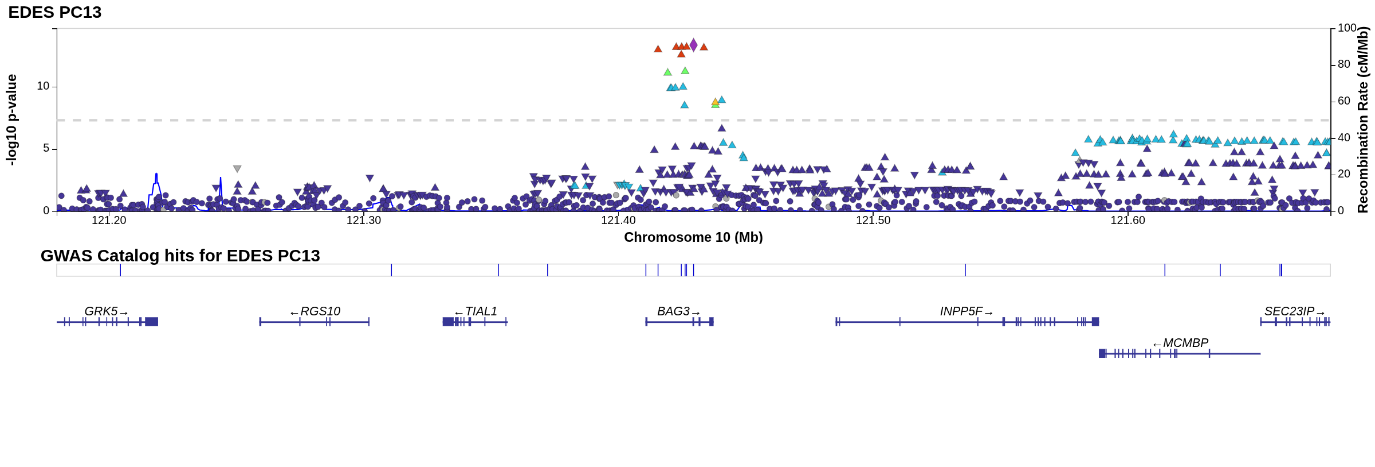

Supplementary Figure 98 – Locus zoom rs34866937 PC13

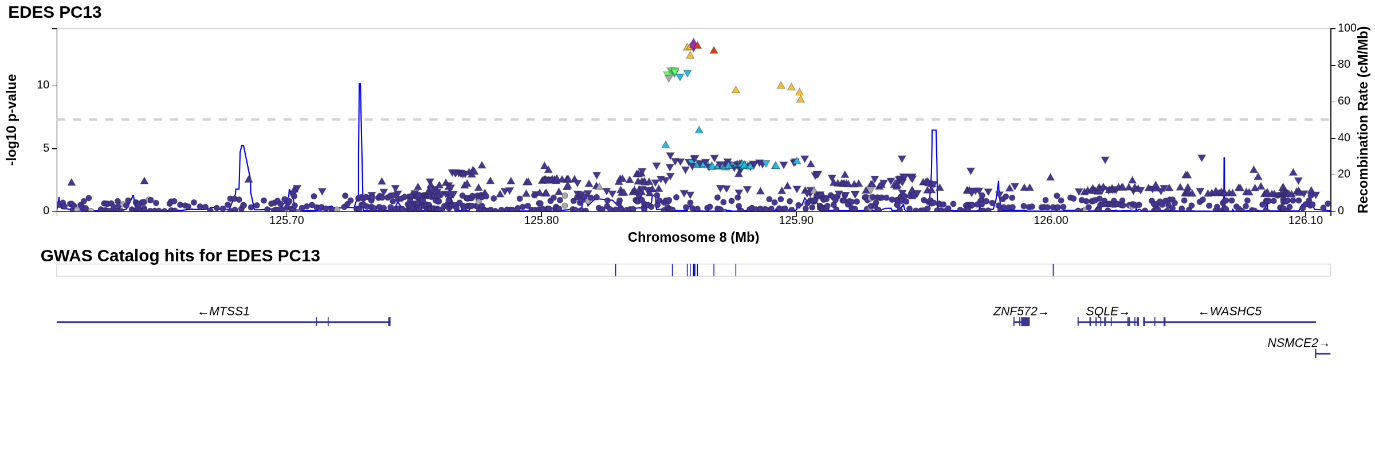

Supplementary Figure 99 – Locus zoom rs56242556 PC13

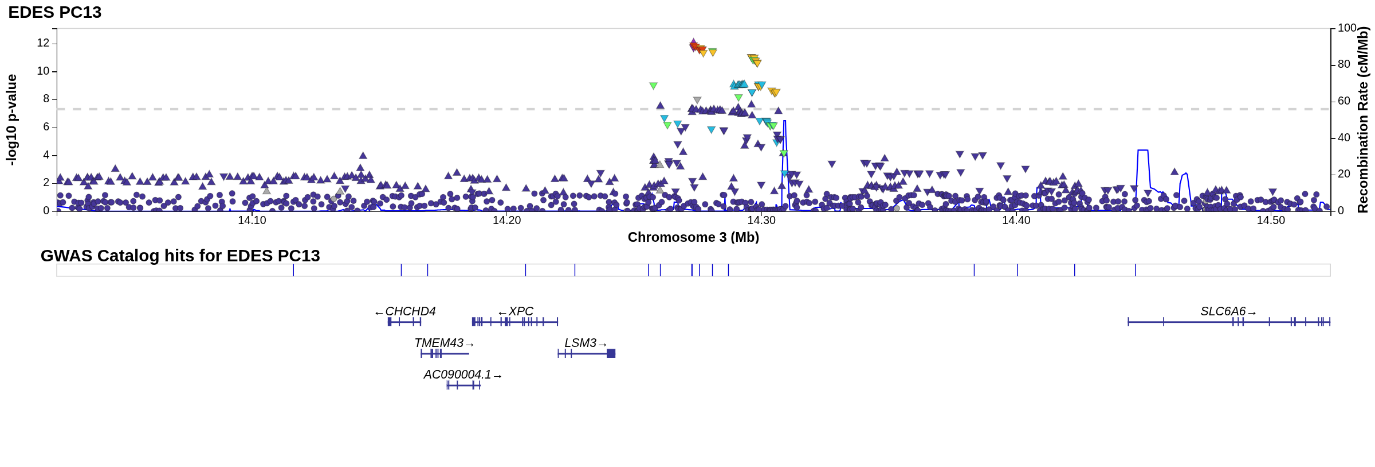

Supplementary Figure 100 – Locus zoom rs1048238 PC13

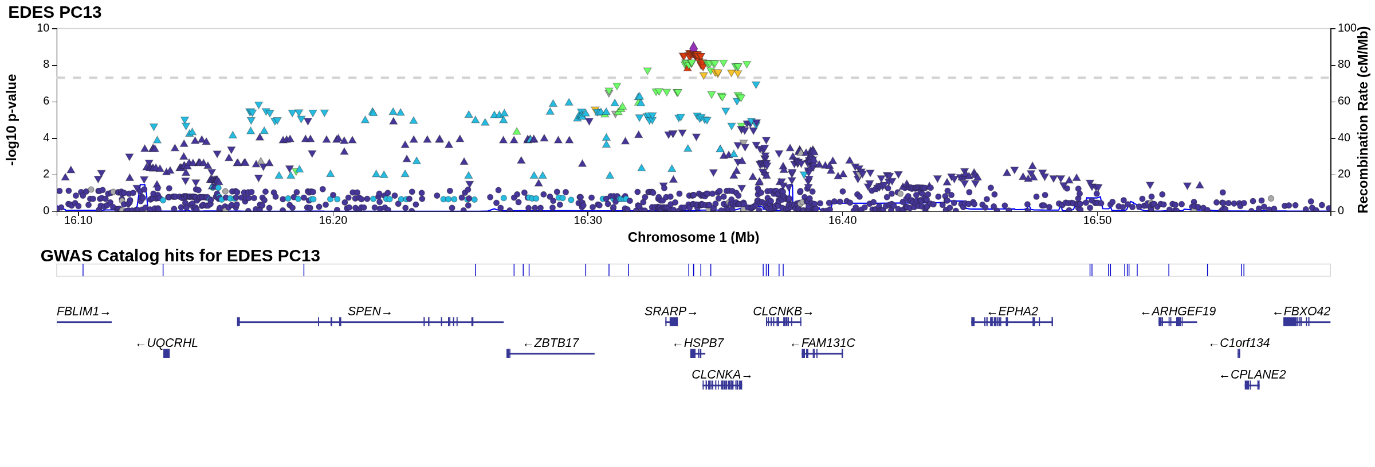

Supplementary Figure 101 – Locus zoom rs35605324 PC13

Supplementary Figure 102 – Locus zoom rs1915986 PC13

Supplementary Figure 103 – Locus zoom rs17079881 PC14

Supplementary Figure 104 – Locus zoom rs12724121 PC14

Supplementary Figure 105 – Locus zoom rs12488245 PC14

Supplementary Figure 106 – Locus zoom 18:34219777_G_GTT PC14

Supplementary Movie 1 – PC1 shape variation from the anterior view

*A plot of the biventricular model cycling between end-diastole and end-systole at standard deviations from the mean shape in the direction of PC1, from the anterior view. Left ventricle is shaded green, right ventricle is shaded blue, and the myocardium is shaded pink. The standard deviation shown is represented by s.*

Supplementary Movie 2 – PC1 shape variation from the basal view

*A plot of the biventricular model cycling between end-diastole and end-systole at standard deviations from the mean shape in the direction of PC1, from the basal view. Left ventricle is shaded green, right ventricle is shaded blue, and the myocardium is shaded pink. The standard deviation shown is represented by s.*

Supplementary Movie 3 – PC1 shape variation from the posterior view

*A plot of the biventricular model cycling between end-diastole and end-systole at standard deviations from the mean shape in the direction of PC1, from the posterior view. Left ventricle is shaded green, right ventricle is shaded blue, and the myocardium is shaded pink. The standard deviation shown is represented by s.*

Supplementary Movie 4 – PC2 shape variation from the anterior view

*A plot of the biventricular model cycling between end-diastole and end-systole at standard deviations from the mean shape in the direction of PC2, from the anterior view. Left ventricle is shaded green, right ventricle is shaded blue, and the myocardium is shaded pink. The standard deviation shown is represented by s.*

Supplementary Movie 5 – PC2 shape variation from the basal view

*A plot of the biventricular model cycling between end-diastole and end-systole at standard deviations from the mean shape in the direction of PC2, from the basal view. Left ventricle is shaded green, right ventricle is shaded blue, and the myocardium is shaded pink. The standard deviation shown is represented by s.*

Supplementary Movie 6 – PC2 shape variation from the posterior view

*A plot of the biventricular model cycling between end-diastole and end-systole at standard deviations from the mean shape in the direction of PC2, from the posterior view. Left ventricle is shaded green, right ventricle is shaded blue, and the myocardium is shaded pink. The standard deviation shown is represented by s.*

Supplementary Movie 7 – PC3 shape variation from the anterior view

*A plot of the biventricular model cycling between end-diastole and end-systole at standard deviations from the mean shape in the direction of PC3, from the anterior view. Left ventricle is shaded green, right ventricle is shaded blue, and the myocardium is shaded pink. The standard deviation shown is represented by s.*

Supplementary Movie 8 – PC3 shape variation from the basal view

*A plot of the biventricular model cycling between end-diastole and end-systole at standard deviations from the mean shape in the direction of PC3, from the basal view. Left ventricle is shaded green, right ventricle is shaded blue, and the myocardium is shaded pink. The standard deviation shown is represented by s.*

Supplementary Movie 9 – PC3 shape variation from the posterior view

*A plot of the biventricular model cycling between end-diastole and end-systole at standard deviations from the mean shape in the direction of PC3, from the posterior view. Left ventricle is shaded green, right ventricle is shaded blue, and the myocardium is shaded pink. The standard deviation shown is represented by s.*

Supplementary Movie 10 – PC4 shape variation from the anterior view

*A plot of the biventricular model cycling between end-diastole and end-systole at standard deviations from the mean shape in the direction of PC4, from the anterior view. Left ventricle is shaded green, right ventricle is shaded blue, and the myocardium is shaded pink. The standard deviation shown is represented by s.*

Supplementary Movie 11 – PC4 shape variation from the basal view

*A plot of the biventricular model cycling between end-diastole and end-systole at standard deviations from the mean shape in the direction of PC4, from the basal view. Left ventricle is shaded green, right ventricle is shaded blue, and the myocardium is shaded pink. The standard deviation shown is represented by s.*

Supplementary Movie 12 – PC4 shape variation from the posterior view

*A plot of the biventricular model cycling between end-diastole and end-systole at standard deviations from the mean shape in the direction of PC4, from the posterior view. Left ventricle is shaded green, right ventricle is shaded blue, and the myocardium is shaded pink. The standard deviation shown is represented by s.*

Supplementary Movie 13 – PC5 shape variation from the anterior view

*A plot of the biventricular model cycling between end-diastole and end-systole at standard deviations from the mean shape in the direction of PC5, from the anterior view. Left ventricle is shaded green, right ventricle is shaded blue, and the myocardium is shaded pink. The standard deviation shown is represented by s.*

Supplementary Movie 14 – PC5 shape variation from the basal view

*A plot of the biventricular model cycling between end-diastole and end-systole at standard deviations from the mean shape in the direction of PC5, from the basal view. Left ventricle is shaded green, right ventricle is shaded blue, and the myocardium is shaded pink. The standard deviation shown is represented by s.*

Supplementary Movie 15 – PC5 shape variation from the posterior view

*A plot of the biventricular model cycling between end-diastole and end-systole at standard deviations from the mean shape in the direction of PC5, from the posterior view. Left ventricle is shaded green, right ventricle is shaded blue, and the myocardium is shaded pink. The standard deviation shown is represented by s.*

Supplementary Movie 16 – PC6 shape variation from the anterior view

*A plot of the biventricular model cycling between end-diastole and end-systole at standard deviations from the mean shape in the direction of PC6, from the anterior view. Left ventricle is shaded green, right ventricle is shaded blue, and the myocardium is shaded pink. The standard deviation shown is represented by s.*

Supplementary Movie 17 – PC6 shape variation from the basal view

*A plot of the biventricular model cycling between end-diastole and end-systole at standard deviations from the mean shape in the direction of PC6, from the basal view. Left ventricle is shaded green, right ventricle is shaded blue, and the myocardium is shaded pink. The standard deviation shown is represented by s.*

Supplementary Movie 18 – PC6 shape variation from the posterior view

*A plot of the biventricular model cycling between end-diastole and end-systole at standard deviations from the mean shape in the direction of PC6, from the posterior view. Left ventricle is shaded green, right ventricle is shaded blue, and the myocardium is shaded pink. The standard deviation shown is represented by s.*

Supplementary Movie 19 – PC7 shape variation from the anterior view

*A plot of the biventricular model cycling between end-diastole and end-systole at standard deviations from the mean shape in the direction of PC7, from the anterior view. Left ventricle is shaded green, right ventricle is shaded blue, and the myocardium is shaded pink. The standard deviation shown is represented by s.*

Supplementary Movie 20 – PC7 shape variation from the basal view

*A plot of the biventricular model cycling between end-diastole and end-systole at standard deviations from the mean shape in the direction of PC7, from the basal view. Left ventricle is shaded green, right ventricle is shaded blue, and the myocardium is shaded pink. The standard deviation shown is represented by s.*

Supplementary Movie 21 – PC7 shape variation from the posterior view

*A plot of the biventricular model cycling between end-diastole and end-systole at standard deviations from the mean shape in the direction of PC7, from the posterior view. Left ventricle is shaded green, right ventricle is shaded blue, and the myocardium is shaded pink. The standard deviation shown is represented by s.*

Supplementary Movie 22 – PC8 shape variation from the anterior view

*A plot of the biventricular model cycling between end-diastole and end-systole at standard deviations from the mean shape in the direction of PC8, from the anterior view. Left ventricle is shaded green, right ventricle is shaded blue, and the myocardium is shaded pink. The standard deviation shown is represented by s.*

Supplementary Movie 23 – PC8 shape variation from the basal view

*A plot of the biventricular model cycling between end-diastole and end-systole at standard deviations from the mean shape in the direction of PC8, from the basal view. Left ventricle is shaded green, right ventricle is shaded blue, and the myocardium is shaded pink. The standard deviation shown is represented by s.*

Supplementary Movie 24 – PC8 shape variation from the posterior view

*A plot of the biventricular model cycling between end-diastole and end-systole at standard deviations from the mean shape in the direction of PC8, from the posterior view. Left ventricle is shaded green, right ventricle is shaded blue, and the myocardium is shaded pink. The standard deviation shown is represented by s.*

Supplementary Movie 25 – PC9 shape variation from the anterior view

*A plot of the biventricular model cycling between end-diastole and end-systole at standard deviations from the mean shape in the direction of PC9, from the anterior view. Left ventricle is shaded green, right ventricle is shaded blue, and the myocardium is shaded pink. The standard deviation shown is represented by s.*

Supplementary Movie 26 – PC9 shape variation from the basal view

*A plot of the biventricular model cycling between end-diastole and end-systole at standard deviations from the mean shape in the direction of PC9, from the basal view. Left ventricle is shaded green, right ventricle is shaded blue, and the myocardium is shaded pink. The standard deviation shown is represented by s.*

Supplementary Movie 27 – PC9 shape variation from the posterior view

*A plot of the biventricular model cycling between end-diastole and end-systole at standard deviations from the mean shape in the direction of PC9, from the posterior view. Left ventricle is shaded green, right ventricle is shaded blue, and the myocardium is shaded pink. The standard deviation shown is represented by s.*

Supplementary Movie 28 – PC10 shape variation from the anterior view

*A plot of the biventricular model cycling between end-diastole and end-systole at standard deviations from the mean shape in the direction of PC10, from the anterior view. Left ventricle is shaded green, right ventricle is shaded blue, and the myocardium is shaded pink. The standard deviation shown is represented by s.*

Supplementary Movie 29 – PC10 shape variation from the basal view

*A plot of the biventricular model cycling between end-diastole and end-systole at standard deviations from the mean shape in the direction of PC10, from the basal view. Left ventricle is shaded green, right ventricle is shaded blue, and the myocardium is shaded pink. The standard deviation shown is represented by s.*

Supplementary Movie 30 – PC10 shape variation from the posterior view

*A plot of the biventricular model cycling between end-diastole and end-systole at standard deviations from the mean shape in the direction of PC10, from the posterior view. Left ventricle is shaded green, right ventricle is shaded blue, and the myocardium is shaded pink. The standard deviation shown is represented by s.*

Supplementary Movie 31 – PC11 shape variation from the anterior view

*A plot of the biventricular model cycling between end-diastole and end-systole at standard deviations from the mean shape in the direction of PC11, from the anterior view. Left ventricle is shaded green, right ventricle is shaded blue, and the myocardium is shaded pink. The standard deviation shown is represented by s.*

Supplementary Movie 32 – PC11 shape variation from the basal view

*A plot of the biventricular model cycling between end-diastole and end-systole at standard deviations from the mean shape in the direction of PC11, from the basal view. Left ventricle is shaded green, right ventricle is shaded blue, and the myocardium is shaded pink. The standard deviation shown is represented by s.*

Supplementary Movie 33 – PC11 shape variation from the posterior view

*A plot of the biventricular model cycling between end-diastole and end-systole at standard deviations from the mean shape in the direction of PC11, from the posterior view. Left ventricle is shaded green, right ventricle is shaded blue, and the myocardium is shaded pink. The standard deviation shown is represented by s.*

Supplementary Movie 34 – PC12 shape variation from the anterior view

*A plot of the biventricular model cycling between end-diastole and end-systole at standard deviations from the mean shape in the direction of PC12, from the anterior view. Left ventricle is shaded green, right ventricle is shaded blue, and the myocardium is shaded pink. The standard deviation shown is represented by s.*

Supplementary Movie 35 – PC12 shape variation from the basal view

*A plot of the biventricular model cycling between end-diastole and end-systole at standard deviations from the mean shape in the direction of PC12, from the basal view. Left ventricle is shaded green, right ventricle is shaded blue, and the myocardium is shaded pink. The standard deviation shown is represented by s.*

Supplementary Movie 36 – PC12 shape variation from the posterior view

*A plot of the biventricular model cycling between end-diastole and end-systole at standard deviations from the mean shape in the direction of PC12, from the posterior view. Left ventricle is shaded green, right ventricle is shaded blue, and the myocardium is shaded pink. The standard deviation shown is represented by s.*

Supplementary Movie 37 – PC13 shape variation from the anterior view

*A plot of the biventricular model cycling between end-diastole and end-systole at standard deviations from the mean shape in the direction of PC13, from the anterior view. Left ventricle is shaded green, right ventricle is shaded blue, and the myocardium is shaded pink. The standard deviation shown is represented by s.*

Supplementary Movie 38 – PC13 shape variation from the basal view

*A plot of the biventricular model cycling between end-diastole and end-systole at standard deviations from the mean shape in the direction of PC13, from the basal view. Left ventricle is shaded green, right ventricle is shaded blue, and the myocardium is shaded pink. The standard deviation shown is represented by s.*

Supplementary Movie 39 – PC13 shape variation from the posterior view

*A plot of the biventricular model cycling between end-diastole and end-systole at standard deviations from the mean shape in the direction of PC13, from the posterior view. Left ventricle is shaded green, right ventricle is shaded blue, and the myocardium is shaded pink. The standard deviation shown is represented by s.*

Supplementary Movie 40 – PC14 shape variation from the anterior view

*A plot of the biventricular model cycling between end-diastole and end-systole at standard deviations from the mean shape in the direction of PC14, from the anterior view. Left ventricle is shaded green, right ventricle is shaded blue, and the myocardium is shaded pink. The standard deviation shown is represented by s.*

Supplementary Movie 41 – PC14 shape variation from the basal view

*A plot of the biventricular model cycling between end-diastole and end-systole at standard deviations from the mean shape in the direction of PC14, from the basal view. Left ventricle is shaded green, right ventricle is shaded blue, and the myocardium is shaded pink. The standard deviation shown is represented by s.*

Supplementary Movie 42 – PC14 shape variation from the posterior view

*A plot of the biventricular model cycling between end-diastole and end-systole at standard deviations from the mean shape in the direction of PC14, from the posterior view. Left ventricle is shaded green, right ventricle is shaded blue, and the myocardium is shaded pink. The standard deviation shown is represented by s.*
