## Supplementary Methods for "Biventricular cardiac dynamic shape: genetics and cardiometabolic disease associations"

### Genetic analyses

#### Heritability analysis

Heritability analysis was performed to estimate the percentage of variance explained by additive genetic variation for each dynamic cardiac shape PC on 36,992 white-European individuals (agreed between self-reported ancestry and genetically inferred ancestry) with high quality imaging data, no previous history of major cardiovascular disease, a left ventricular ejection fraction of >40%, and 558,994 model SNVs (of minor allele frequency > 1%, a Hardy-Weinberg exact test of < 1x10^-6^ and a missing rate of < 1.5% selected using PLINK^1^ (version 1.90b6.2)) using BOLT-REML^2^ (version 2.3) with a MAF threshold of ≥ 1% and INFO > 0.3, using the model SNVs and ~1.2 million imputed SNVs.

The heritability analysis was adjusted for height, age, sex, genotyping array used, a medication adjusted measure of systolic blood pressure averaged between automated and manual readings, BMI and resting heart rate.

#### Genome-wide association studies

Once a genetic basis for the dynamic shape PCs was established, each PC was analysed using a linear mixed model method (BOLT-LMM, version 2.4.1)^3^ to identify genetic loci. Multiple linear regression was applied to each PC with an additive genetic effect adjusted for height, age, sex, array type (UK Biobank/UK BiLEVE), systolic blood pressure (averaged between manual and automated readings and adjusted 15mmHg in the presence of blood pressure altering medication), resting heart rate and BMI*.* A P < 5x10^-8^ was used to declare genome-wide significance. LocusZoom region plots for all significant loci identified are available in *Supplementary Figures*.

To assess for population stratification and confounding, the LD score regression intercept was estimated using LD score software (LDSC, v1.0.1)^4^. Genomic inflation factor was reported from the mean lambda value output by BOLT-LMM, and quantile-quantile plots were generated from the GWAS summary statistics with the Functional Mapping and Annotation of Genome-Wide Association Studies (FUMA GWAS)^5^ (functional mapping and annotation of genome-wide association studies) functional annotation tool (version 1.4.0).

A genomic signal was defined as the most significant variant defined as the lead SNV and all SNVs within a 1mb window in linkage disequilibrium (LD) r^2^ > 0.1 in a 1mb region. A locus was defined as the 1mb window around the lead variant, disregarding LD.

The percent variance explained for each PC by all genome-wide significant variants and independent secondary signals was calculated^6^ as:

$$\frac{2\beta^{2}MAF\left( 1-MAF \right)}{2\beta^{2}MAF \left( 1-MAF \right) + SE^{2}\cdot2N\cdot MAF\left( 1-MAF \right)}$$

Per-variant estimates were summed to obtain the total PVE across all genome-wide significant and independent secondary variants. Effect estimates were derived from GWAS models that included the standard covariates.

#### Conditional analysis

To identify secondary signals at genome-wide significant loci, conditional analysis was performed using GCTA (version 1.94.1). A secondary signal would be declared if:

- The newly identified SNV original P-value was lower than 1x10^−6^
- There was less than a 1.5-fold difference between the main association and conditional association P-values on a –log_10_ scale, i.e., if -log_10_(P_sec_)/-log_10_(P_cond_) < 1.5
- The newly identified SNV is in LD < 0.1 with the main association

#### Variant Effect Prediction

Analyses were performed to annotate the identified lead SNVs and their proxies (r^2^ ≥ 0.8), and the secondary signals. Positional information and GWAS summary statistics were extracted, and each lead or conditionally independent SNV was assessed with Ensembl Variant Effect Predictor (release 105.0)^7^, a collation of tools used for assessing and predicting the effects of SNVs and their impacts on human biology (through SIFT version 5.2.2 and PolyPhen-2 version 2.2.2, release 405c)^8,9^.

#### Expression Quantitative Trait Locus (eQTL) analysis and colocalisation

To assess for potential effects of lead and conditionally independent variants (and their proxies [r^2^ ≥ 0.8]) on tissue specific gene expression in left ventricle (LV), aortic artery, coronary artery and atrial appendage tissues, they were first checked for overlap with lead eQTL variants at each tissue using the GTEx (version 8) database^10^. Subsequently, colocalization analyses were performed using the COLOC package (version 5.1.0.1)^11^ in R to analyse each eQTL-GWAS dataset pair. This tool uses Bayesian statistical methodology to test the pairwise colocalization of SNVs in a GWAS with eQTLs, and generates posterior probabilities for each locus, weighting the evidence for competing hypotheses of no evidence of colocalization or the sharing of a distinct SNV at each locus. A posterior probability of PP4 > 0.75 was used to indicate strong evidence of a tissue-specific eQTL-GWAS pair influencing both the expression and GWAS trait at a particular region for the specified GTEx tissues of interest.

#### Transcriptome-wide Association Study (TWAS)

The gene-based association tool S-PrediXcan (version 0.6.5, <https://github.com/hakyimlab/MetaXcan>^12^) was used to perform a TWAS to predict the effects of gene expression levels on each PC using GWAS summary statistics. S-PrediXcan provides a precalculated transcriptome model database from GTEX-based tissues and covariance matrices of SNVs within each gene model. A Bonferroni corrected threshold (0.05/number of tissue-gene pairs tested 0.05/16,097 = 3.1x10^-6^) was used to declare significant results.

#### Assessment of regulatory interactions (Hi-C)

The GWAS lead SNVs and high LD proxies (r^2^ > 0.8) were also assessed for their regulatory potential using RegulomeDB^13^, to find genes with which promoter regions form significant chromatin interactions in left and right ventricular and aortic tissues. This utilises long-range chromatin interaction (Hi-C) data from FUMA^14^ and Jung^15^ datasets, which use different methods and resolutions of Hi-C data and provide additional discovery power when used together. Target genes were only considered with evidence of significant enhancer-promoter interactions at FDR < 1x10^-6^ and filtered to regulatory GWAS variants with a RegulomeDB score of < 5 (where a lower score indicates greater evidence of functional significance, and chose the interactors of highest regulatory potential to annotate the loci.

#### Polygenic Priority Scoring (PoPS)

Gene prioritisation was performed using Polygenic Priority Scoring (PoPS)^16^. This tool computes gene level association statistics and correlations using GWAS summary statistics and LD information in the MAGMA tool^17^. Gene-level associations are adjusted for gene length and SNP density, then the PoPS tool performs marginal feature selection using MAGMA to perform enrichment analysis of each gene separately. Finally, PoPS computes a polygenic priority score for each gene based on the selected gene features. This outputs a list of genes, each with a polygenic priority score. This list is restricted to genes that fall within a 1Mb window of a genome-wide significant GWAS variant, and within each window the gene which has the highest polygenic priority score is considered as the candidate gene for that lead variant selected by PoPS.

### Causal gene prioritisation

All genome-wide significant variants were initially annotated by nearest gene using University of California, Santa Cruz Genome Browser^18^. Potential causal genes at each locus were shortlisted from results of bioinformatics analyses which included: variant effect prediction (VEP), Polygenic Priority Score (PoPS)^16^, expression quantitative trait locus (eQTL) colocalization analysis, Hi-C long-range chromatin interactions and a TWAS.

Genes were selected as candidate genes where they had the highest level of support from the following criteria:

- The gene was within ±500kb of the lead variant
- Either the lead variant or one of its high LD proxies (r^2^ > 0.8) had a predicted missense effect in that gene from the VEP tool
- The lead variant or a high LD proxy was an eQTL associated with that gene in cardiac tissues with evidence of colocalization (shared causal variants)
- The gene was identified through TWAS in cardiac tissues,
- The gene was identified through Hi-C analysis in cardiac tissues
- The gene was identified in the PoPS gene prioritisation tool

### Pathway analysis

Candidate genes were queried in the Gene2Func pathway analysis tool from FUMA to perform functional enrichment analysis and identify significantly associated gene ontology terms and biological pathways from Kyoto Encyclopaedia of Genes and Genomes^19^, Reactome^20^ and WikiPathways^21^ and results from GWAS catalog^22^. Significant enrichment was defined as an adjusted False Discovery Rate P-value < 0.05.

### Rare genetic variant analysis

To assess the contribution of rare genetic variants to dynamic cardiac shape, PCs were tested for burden of rare variants in UKB using Regenie (version 4.0^23^). Using PLINK^24^ samples were QC’ed, removing samples with missingness >2% and genotype missingness >1%. Variants failing HWE (P < 1×10⁻¹⁵) or outside the allele frequency window (MAF <2.2×10⁻⁵ or MAF >0.01) were also excluded. Gene-level rare variant association tests were performed using pre-specified variant masks and annotations, running burden and SKAT-O tests with a minimum minor allele count threshold of 5. Three variant masks were tested: one containing protein-truncating variants, one containing missense variants and one combining the previous two. Variants in these masks were defined using VEP. Variants are also stratified by allele frequency with three bins: ≤ 0.001, 0.001 < MAF ≤ 0.01, and MAF > 0.01. Significant findings were defined as P < 2.79x10^-6^ (0.05/17,902 tested gene sets). Leave one variant out (LOVO) analysis was performed to assess whether significant results were driven by single or multiple rare variants.

### Genetic relationship with end-diastolic shape atlas

To assess the genetic relationship between the dynamic cardiac shape atlas and previously published end-diastolic shape atlas, genetic correlations between PCs from each study were first calculated using the LD Score software tool LDSC (version 1.0.1^4^). Subsequently, all dynamic cardiac shape lead variants and high LD proxies (r^2^ > 0.8) were searched within the summary statistics of the end-diastolic shape GWAS, returning the end-diastolic GWAS statistics with the lowest P-value among the 11 end-diastolic PCs, and in doing so highlighting genome-wide significant signals that were shared between the studies alongside those which are specific to this dynamic cardiac shape GWAS.

### Pleiotropy analyses

To assess novelty of the GWAS results, lead signals were assessed through literature review of cardiac trait GWAS and lookup in GWAS catalog^22^*.* The literature review comprised publicly available GWAS results for cardiac traits, both cardiovascular diseases and measures of cardiac structure and function, and was last updated in March 2026. In the GWAS catalog, genome wide lead SNVs and high LD proxies (r^2^ > 0.8) were looked up, reporting all significant (P ≤ 5x10^-8^) GWAS associations, with a focus on those that are associated with the cardiovascular system (cardiac structural and functional measures, ECG measures, cardiac disease traits) for novelty assessment. If any of our lead variants fall within 500kb of a previously reported SNV associated with a cardiovascular trait, and is in linkage disequilibrium r^2^ ≥ 0.1, the association is reported. Any variants without prior information identified at the genetic signal level was considered novel. Identified prior information was assessed as to whether GWAS associations were cardiac for more detailed reporting.

#### PheWAS

To identify evidence of pleiotropy with clinical conditions, a phenome-wide association study (PheWAS) was performed using the R package PheWAS (version 0.99.6.1^25^). ICD9 and ICD10 codes from the UK Biobank HES data and mapped to phecodes. Lead and conditionally independent secondary variants from unrelated (kinship coefficient > 0.0884) UK Biobank individuals not included in the GWAS (n=412,774), and the PRS constructed for each dynamic cardiac shape PC were tested for association with these phecodes, adjusted for age, sex and the first 10 genetic principal components. Bonferroni corrected threshold (number of phecodes tested with ≥200; 0.05/325 = 1.54 x 10^-4^) was used to declare significance.

### Mendelian Randomization

Publicly available GWAS summary statistics used in the Mendelian randomization analysis were (alongside the case-control splits): AF^28^ (60,620 cases, 970,216 controls), T2D^29^ (154,194 cases, 278,454 controls), HF^30^ (90,653 cases, 1,188,957 controls), IHD^31^ (60,801 cases, 123,504 controls), stroke^32^ (5,386 cases, 343,560 controls) and atrioventricular block^33^ (9,046 cases, 721,907 controls). GWAS summary statistics were prioritised based on maximised sample size while not including individuals from UK Biobank do avoid biasing results.

For all MR analyses, lead SNVs (*P* < 5 × 10^−8^) were taken as genetic instruments. Effect alleles were harmonised between IVs and summary statistics. Where GWAS lead dynamic cardiac shape variants were not present in the outcome data summary statistics, high LD proxy variants which were present were used, prioritising the highest LD variant present in the outcome GWAS with LD > 0.1. To account for weak-instrument bias, the F-statistic was calculated for each lead SNV, and only those with an F > 10 were kept as instrumental variables. SNV’s were subsequently pruned using the ieugwasr package^34^ in R (version 1.1.0) to remove correlated variants (*r^2^* 0.001) within 10Mb, preserving the lowest P-value variant for association analyses. Five MR methods were used for this analysis: IVW^35^, weighted median^36,37^, MR-Egger^38,39^, Simple mode, and MR-PRESSO^40^.
