## Supplementary figures and images for "Biventricular cardiac dynamic shape: genetics and cardiometabolic disease associations"

### SM1.gif

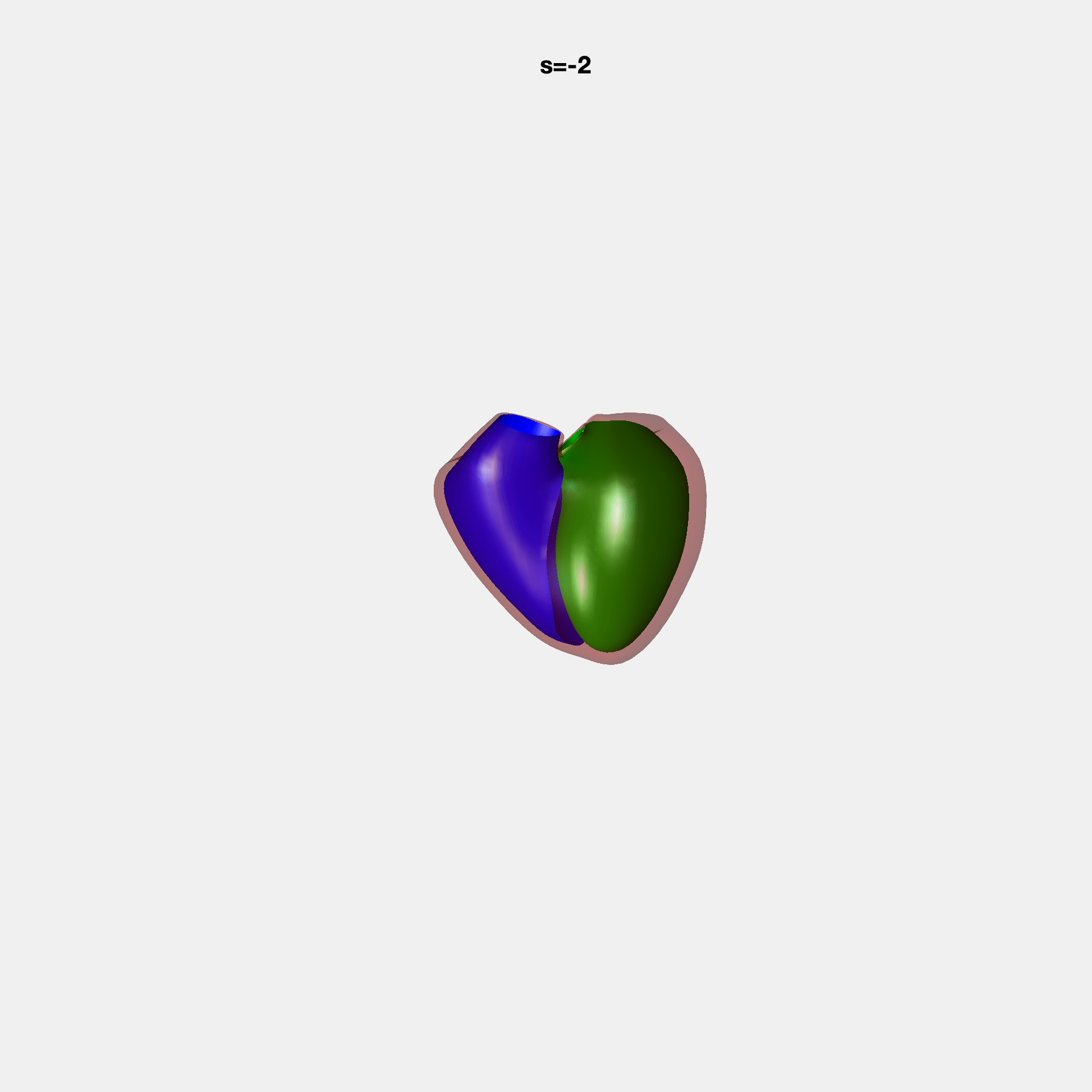

### SM2.gif

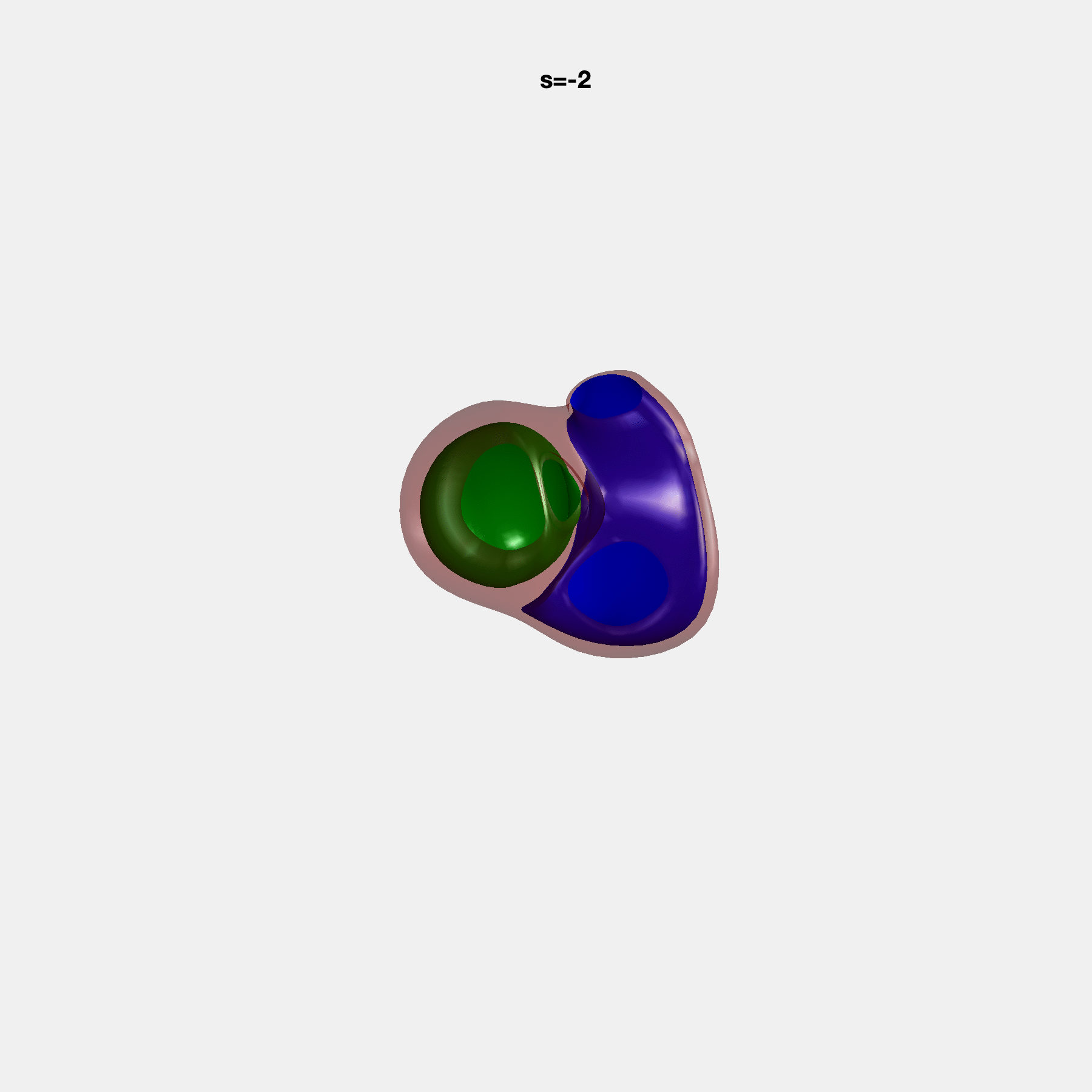

### SM3.gif

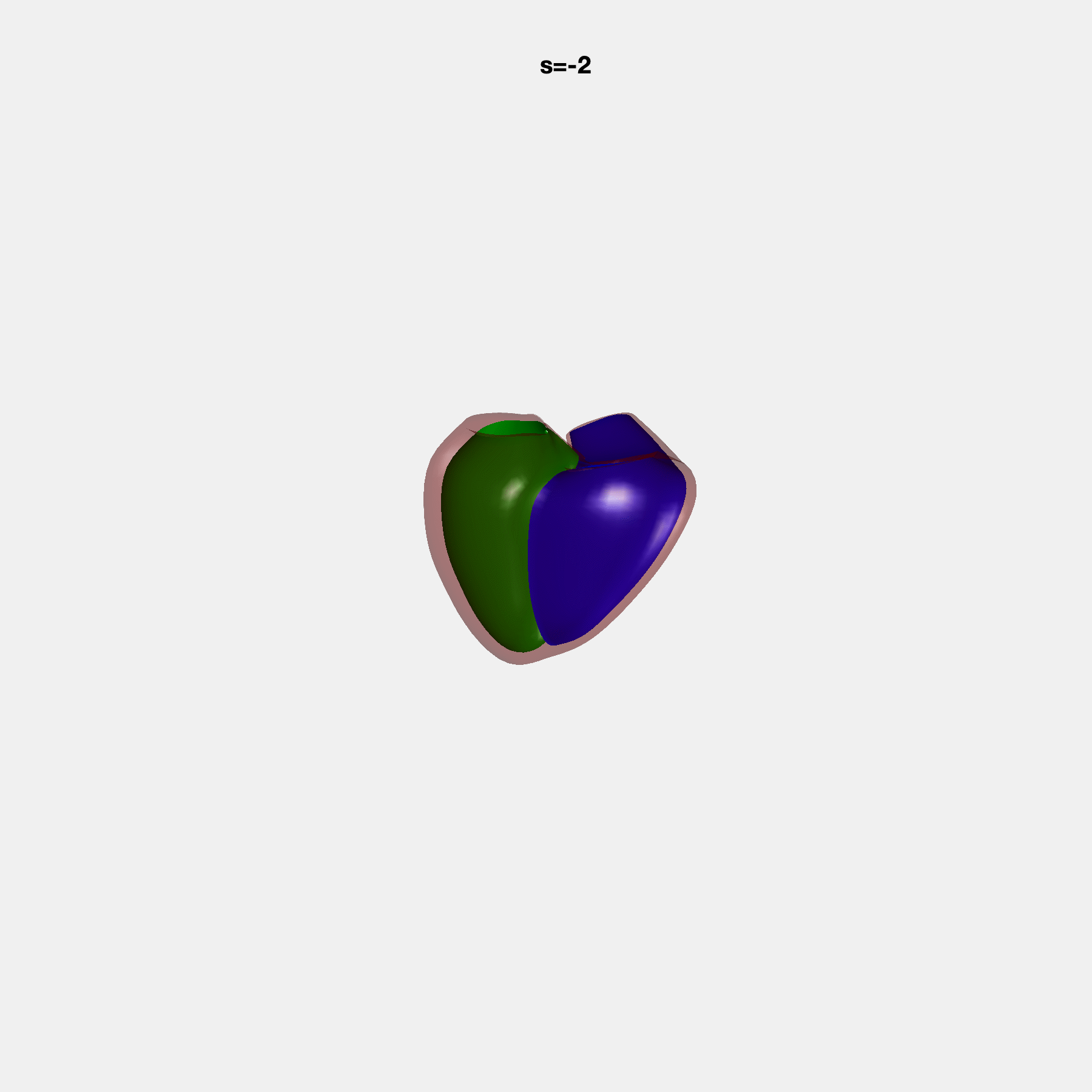

### SM4.gif

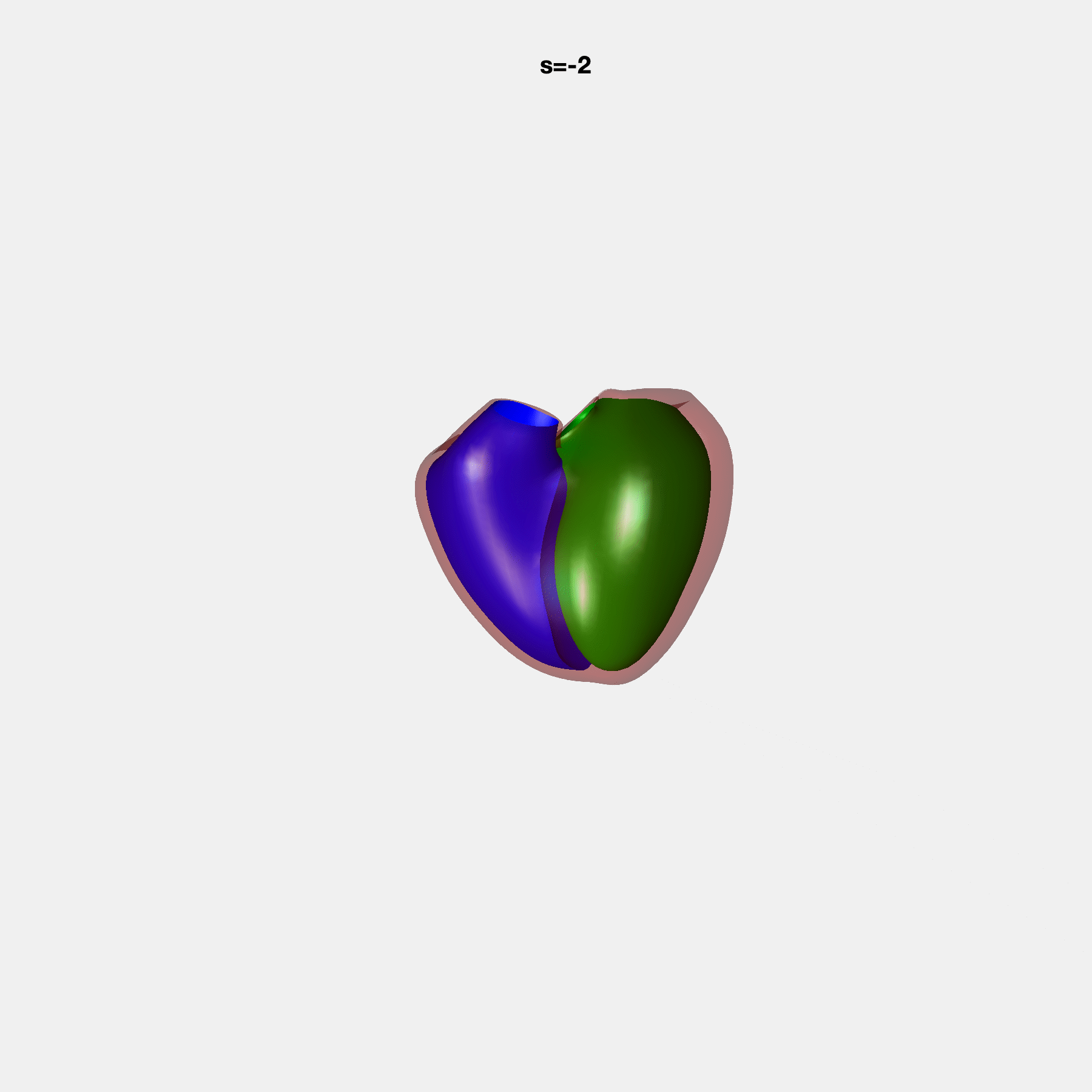

### SM5.gif

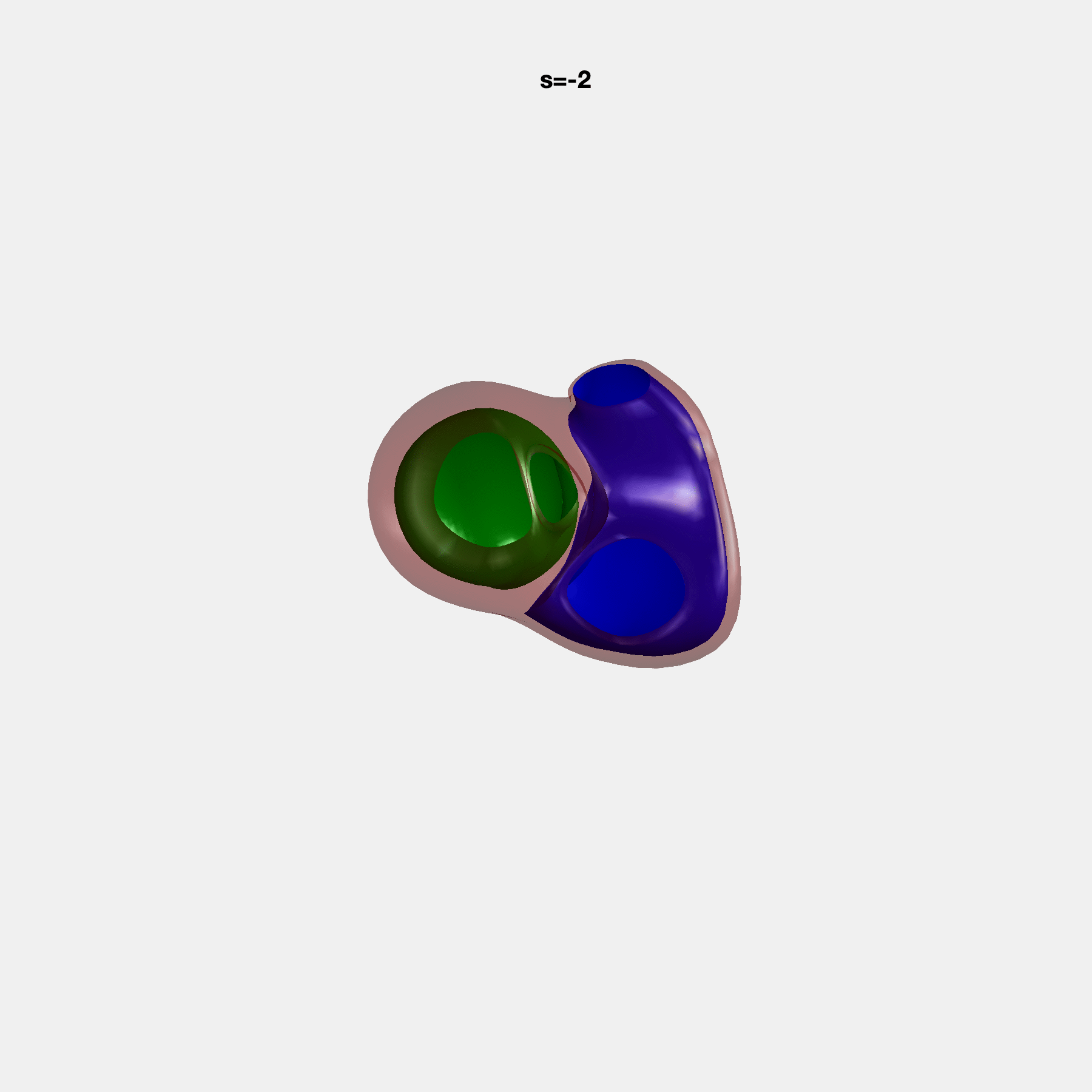

### SM6.gif

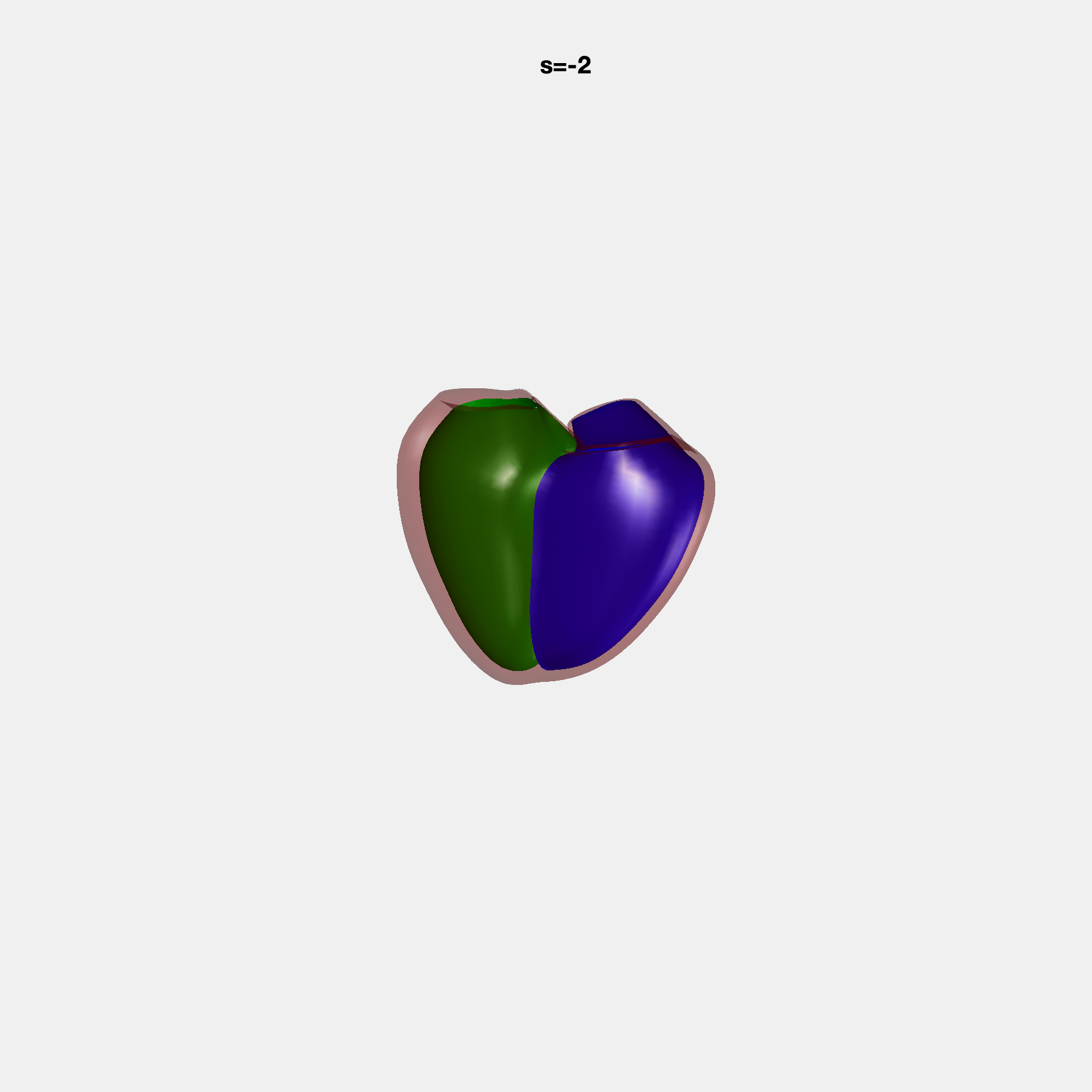

### SM7.gif

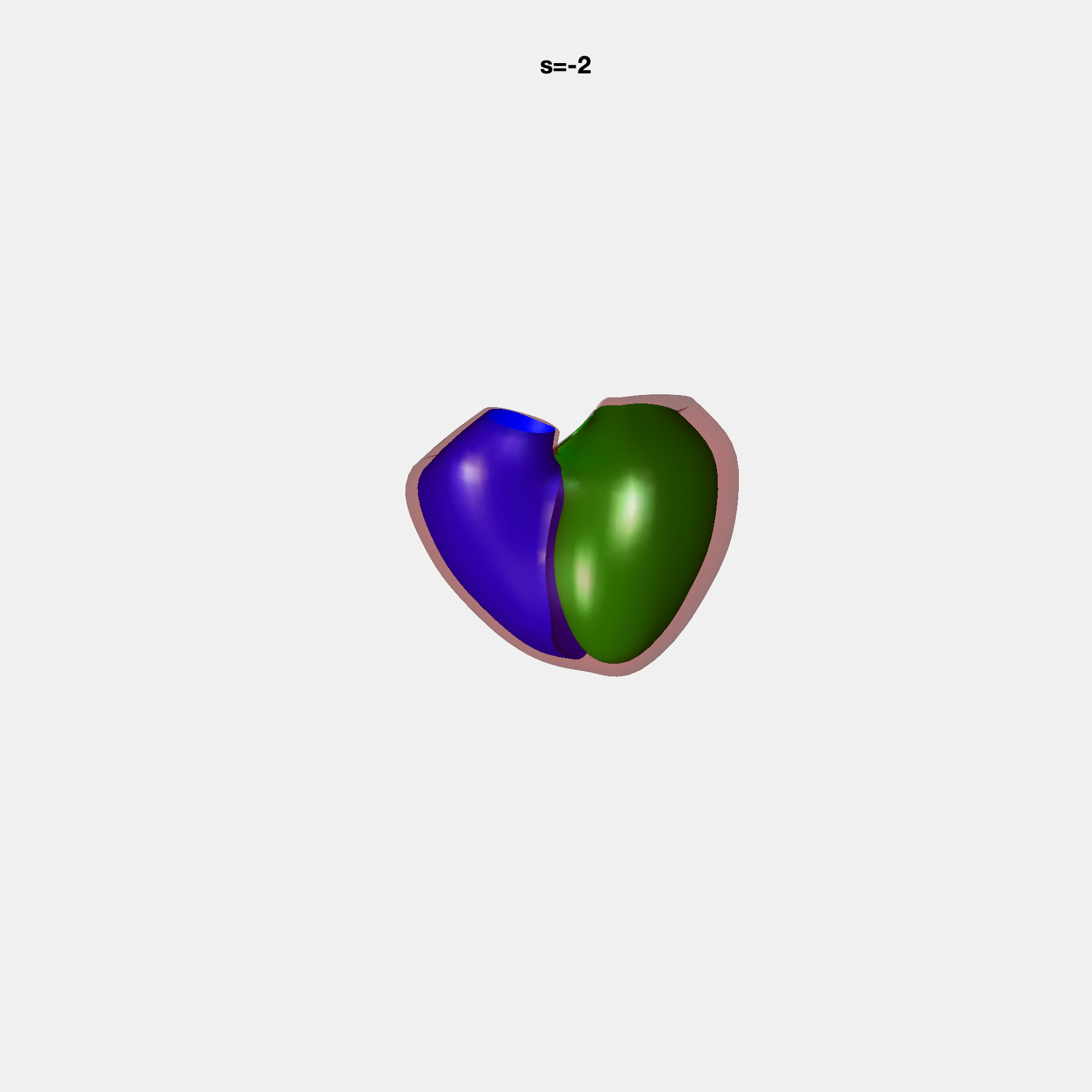

### SM8.gif

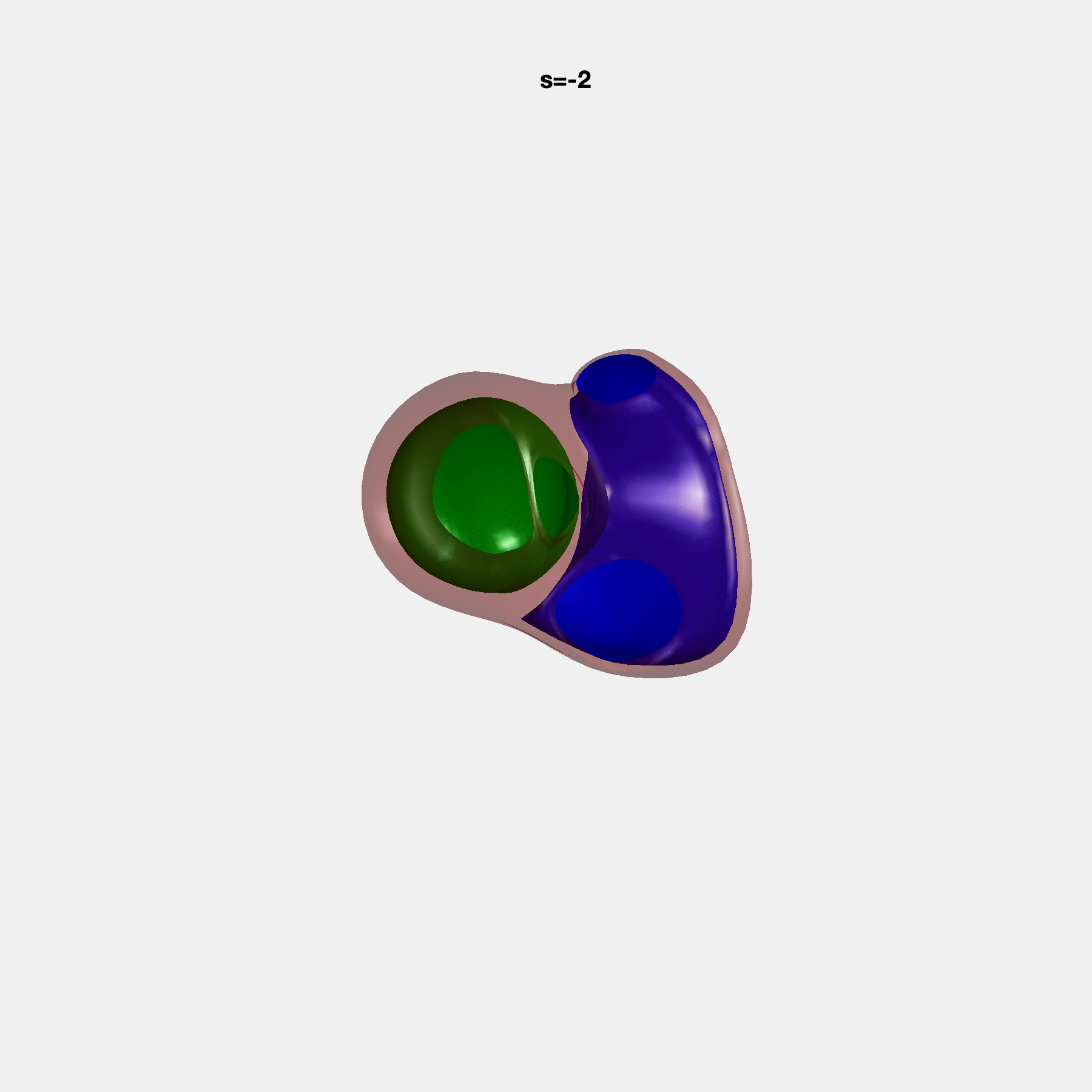

### SM9.gif

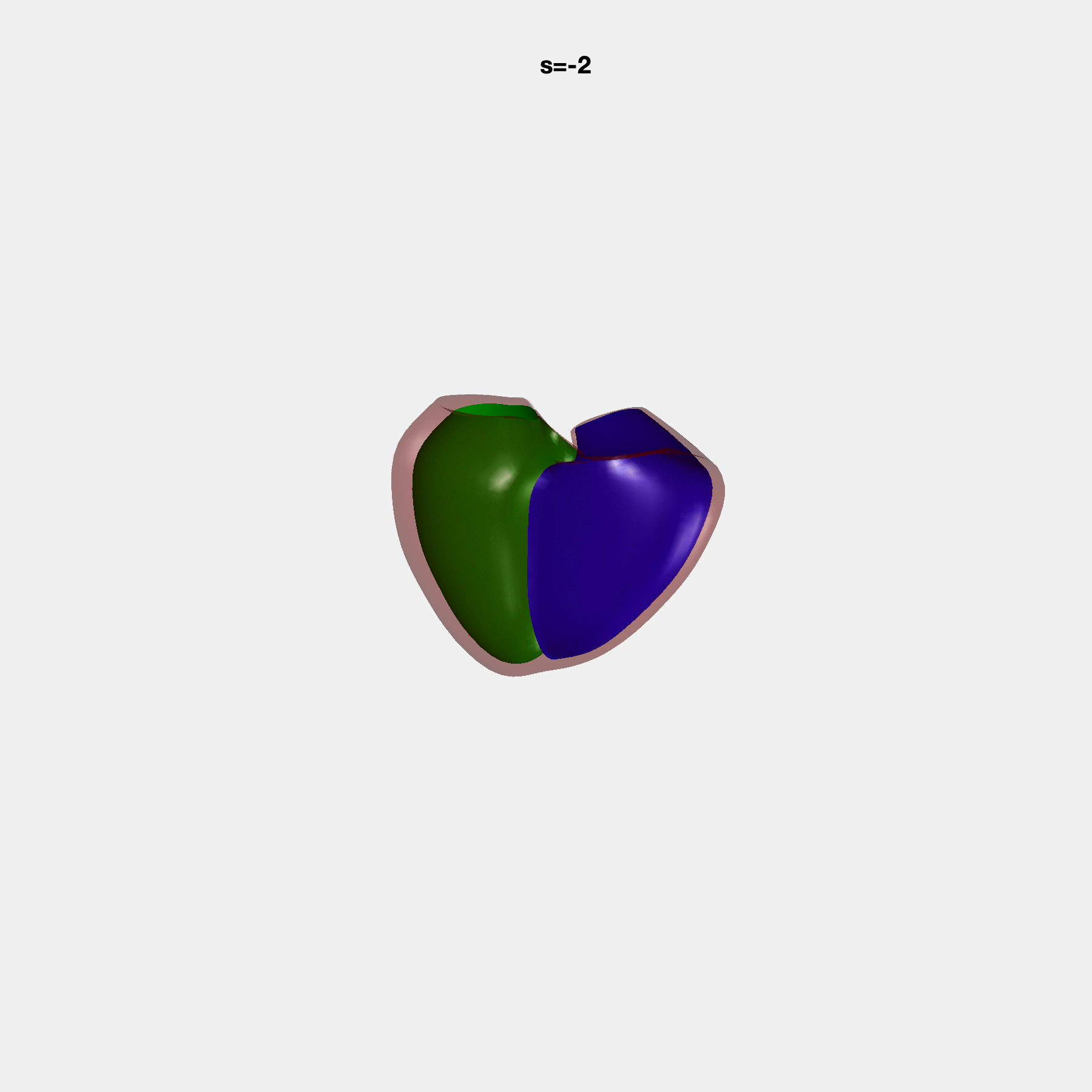

### SM10.gif

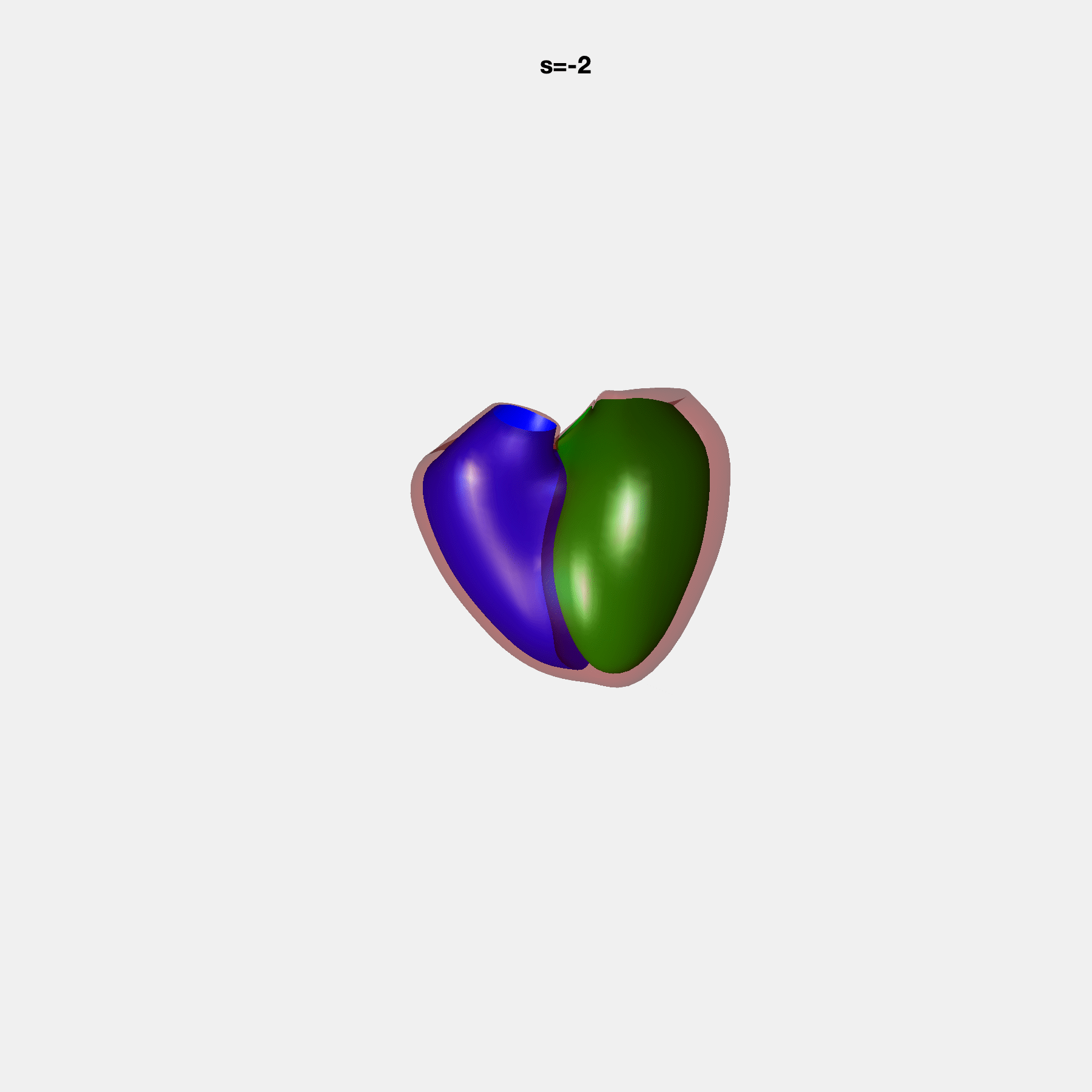

### SM11.gif

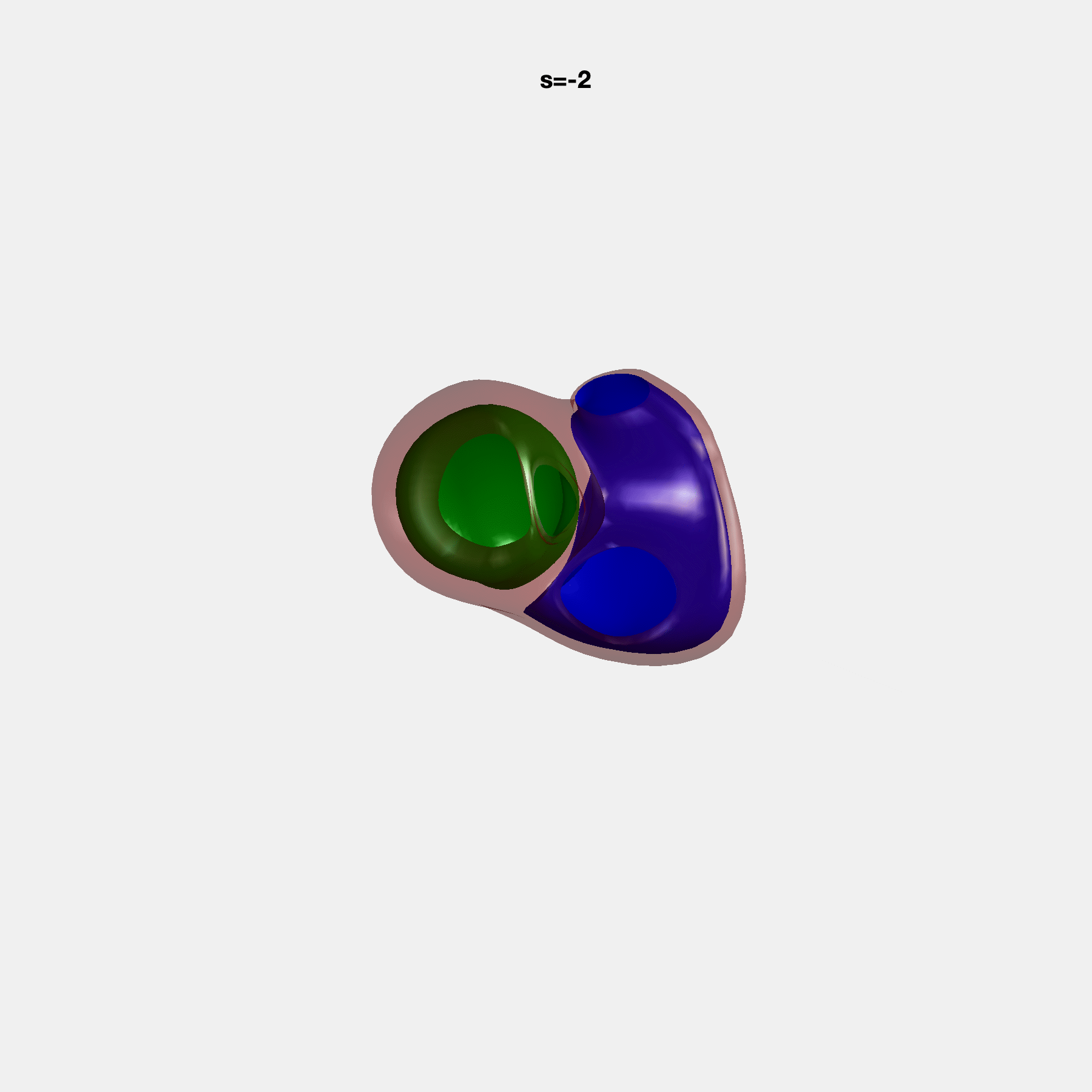

### SM12.gif

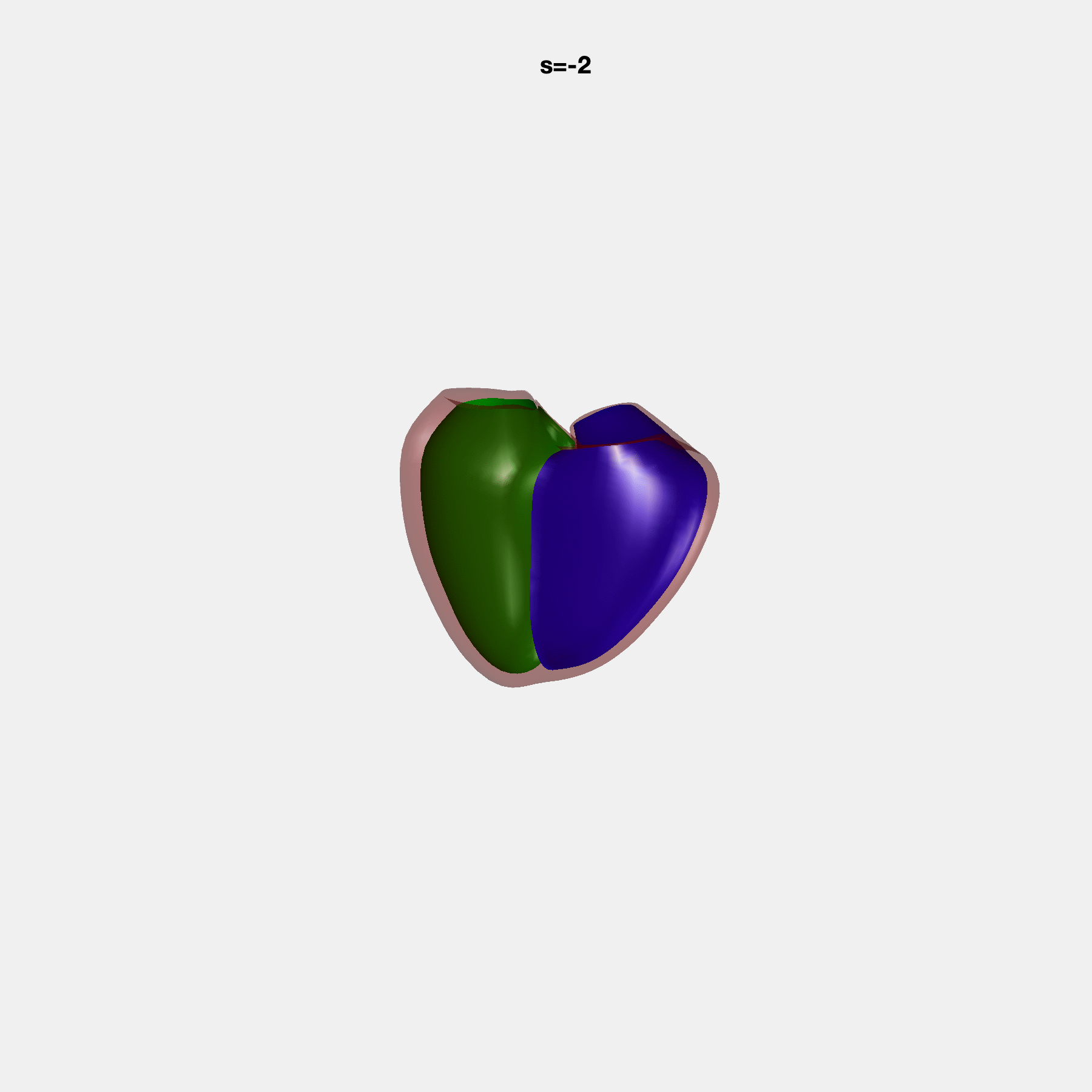

### SM13.gif

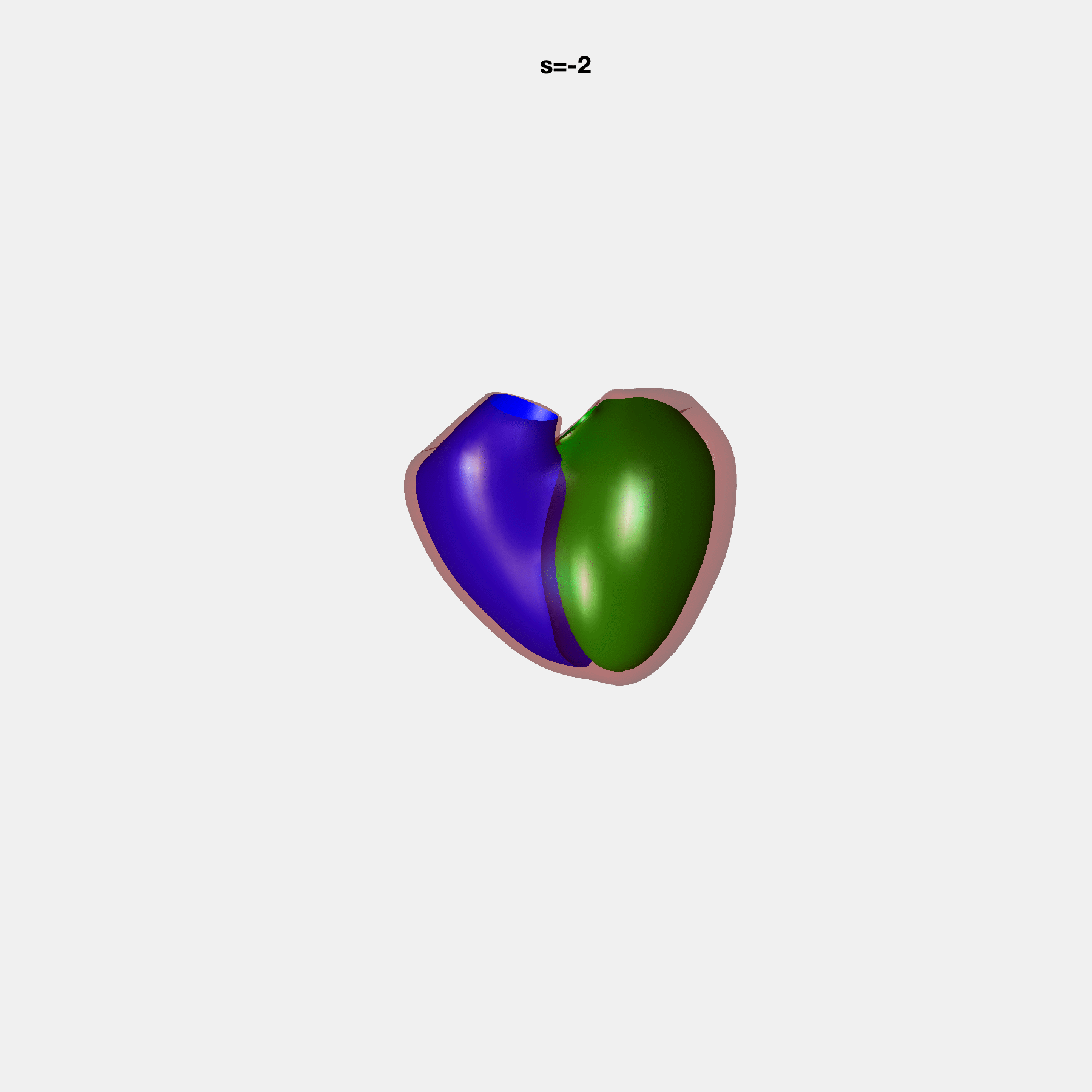

### SM14.gif

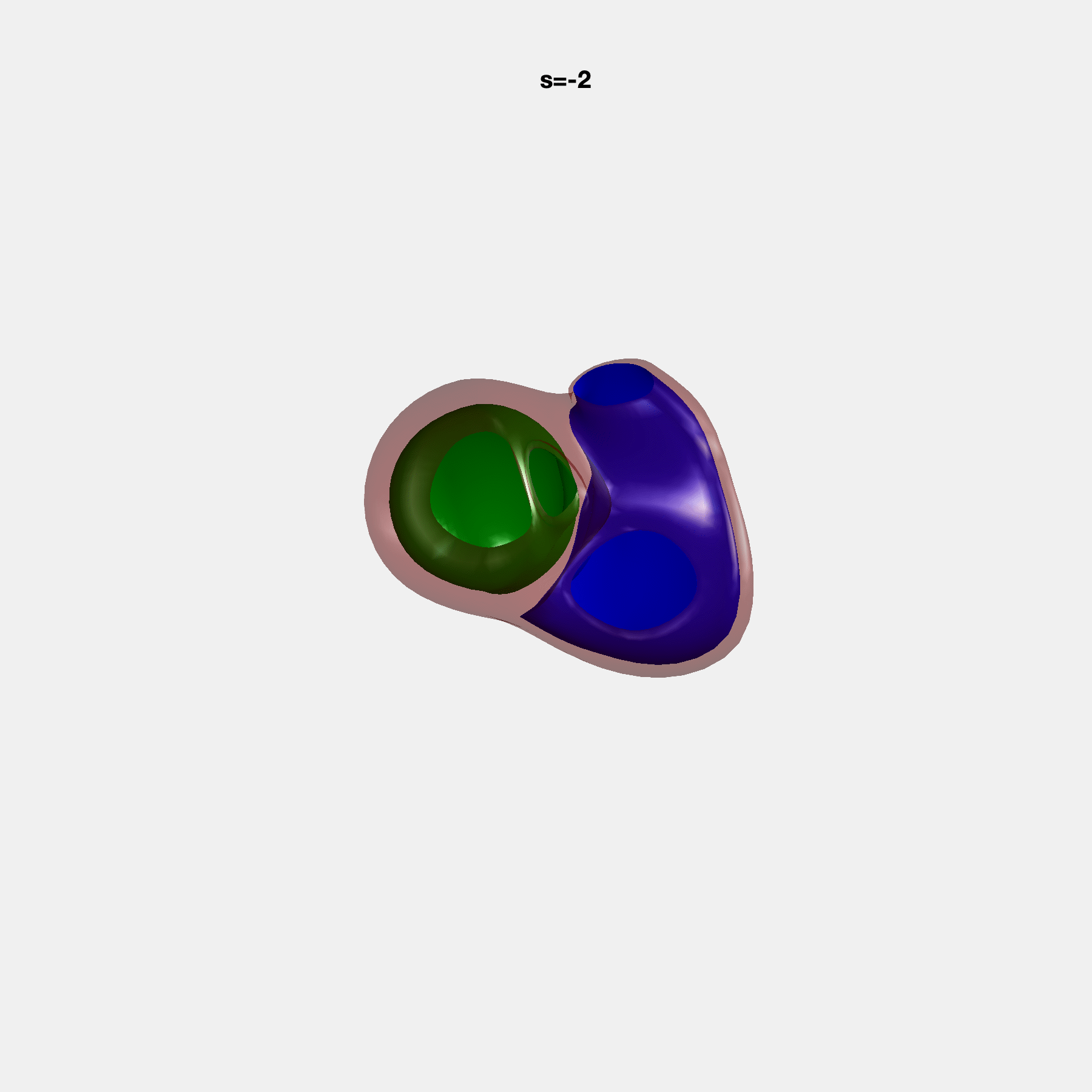

### SM15.gif

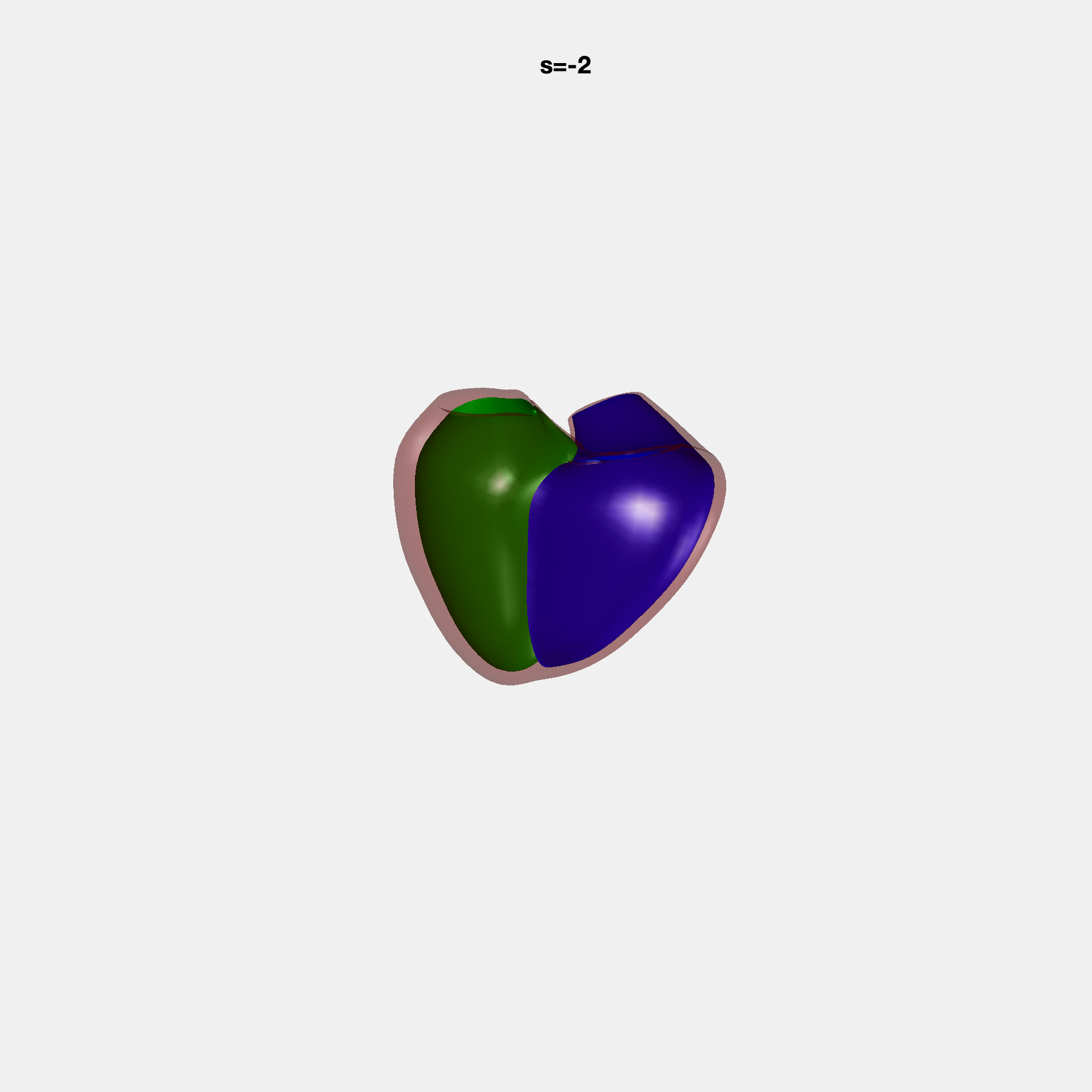

### SM16.gif

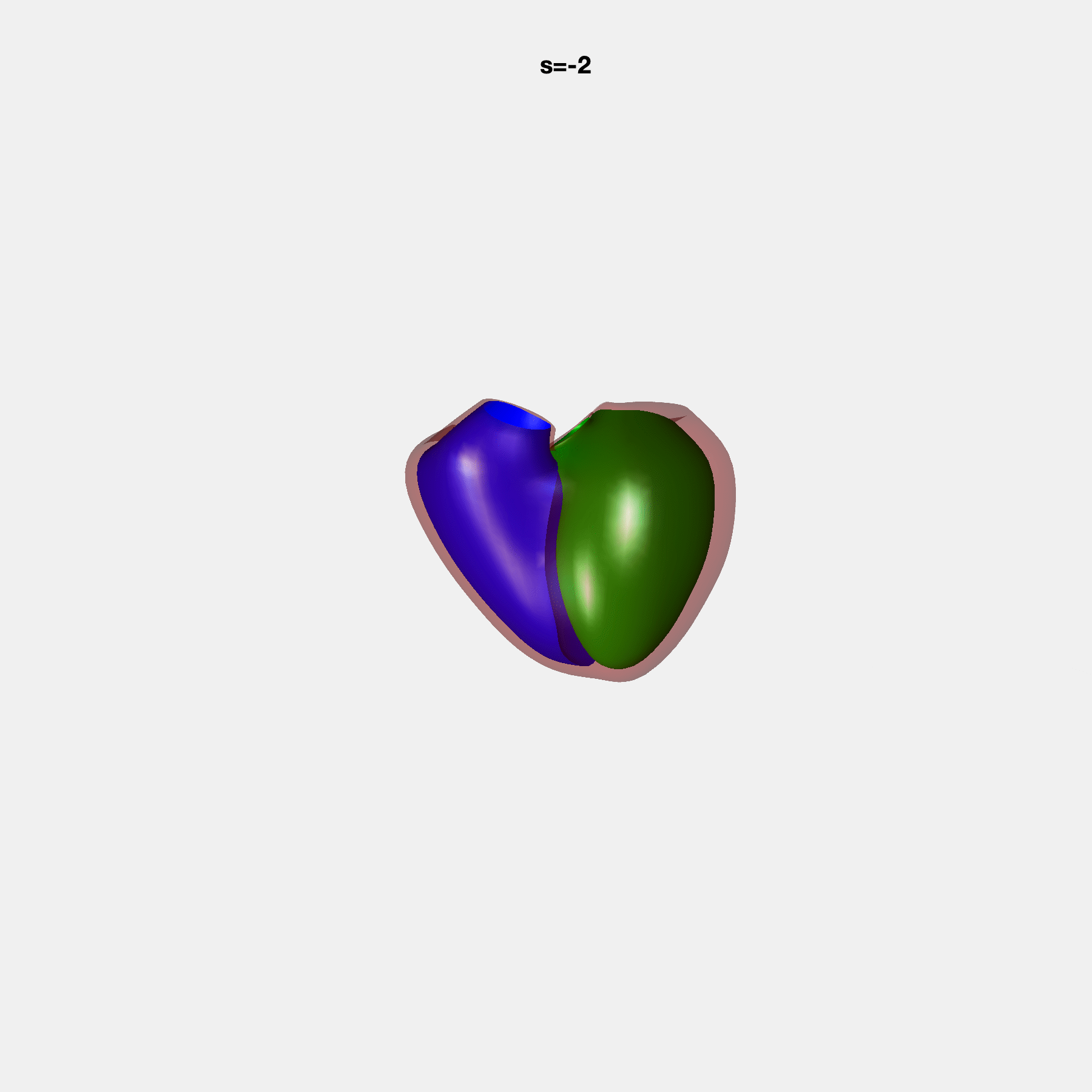

### SM17.gif

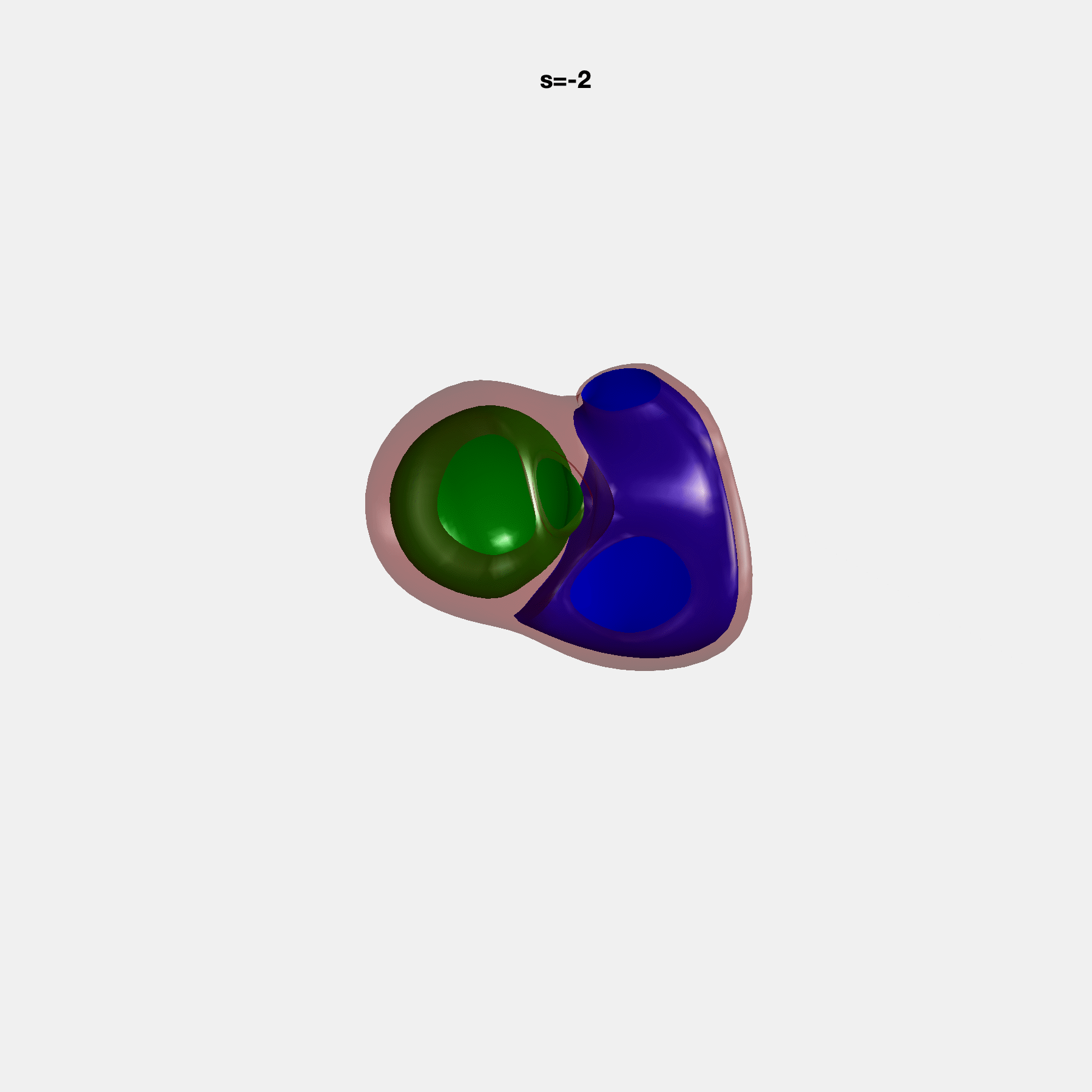

### SM18.gif

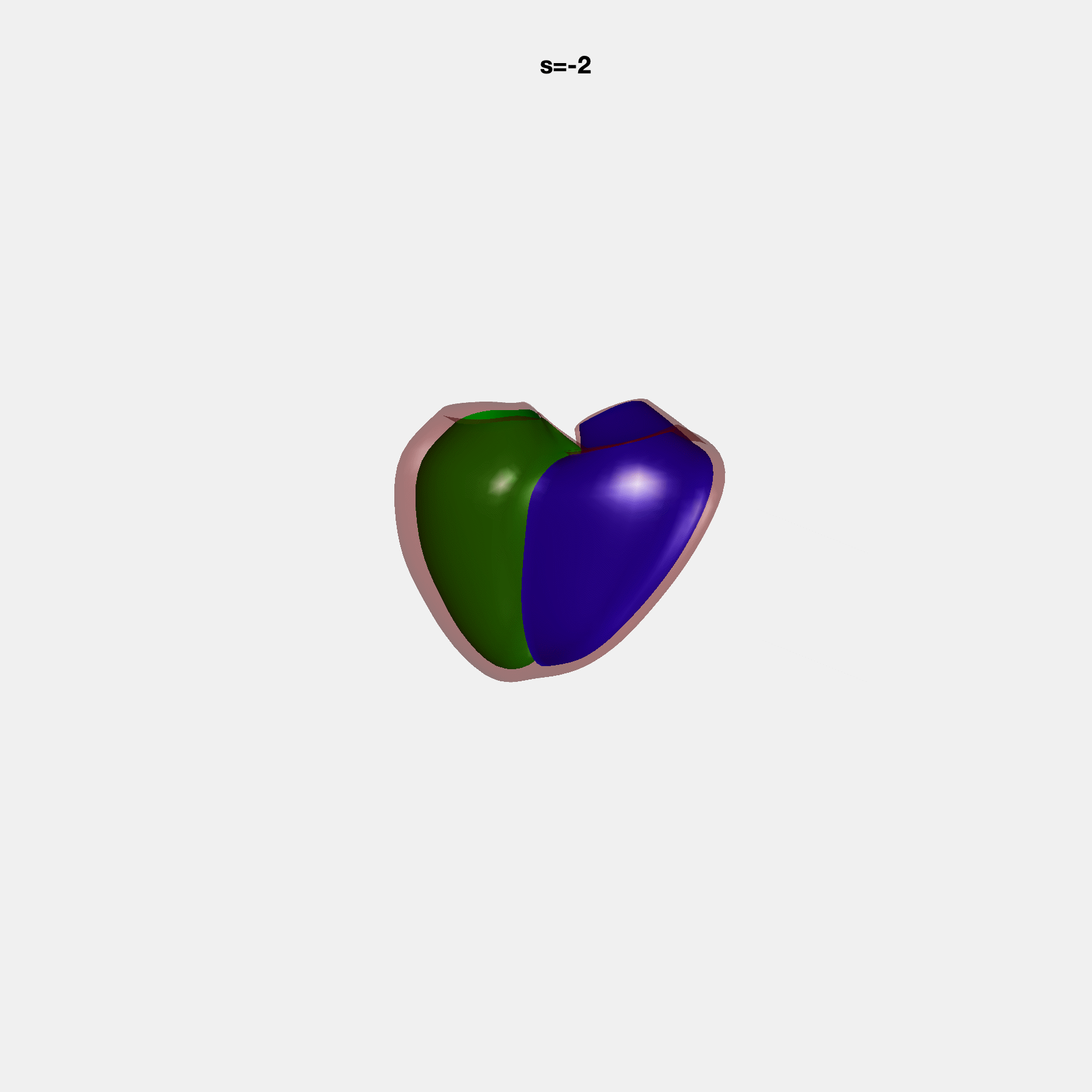

### SM19.gif

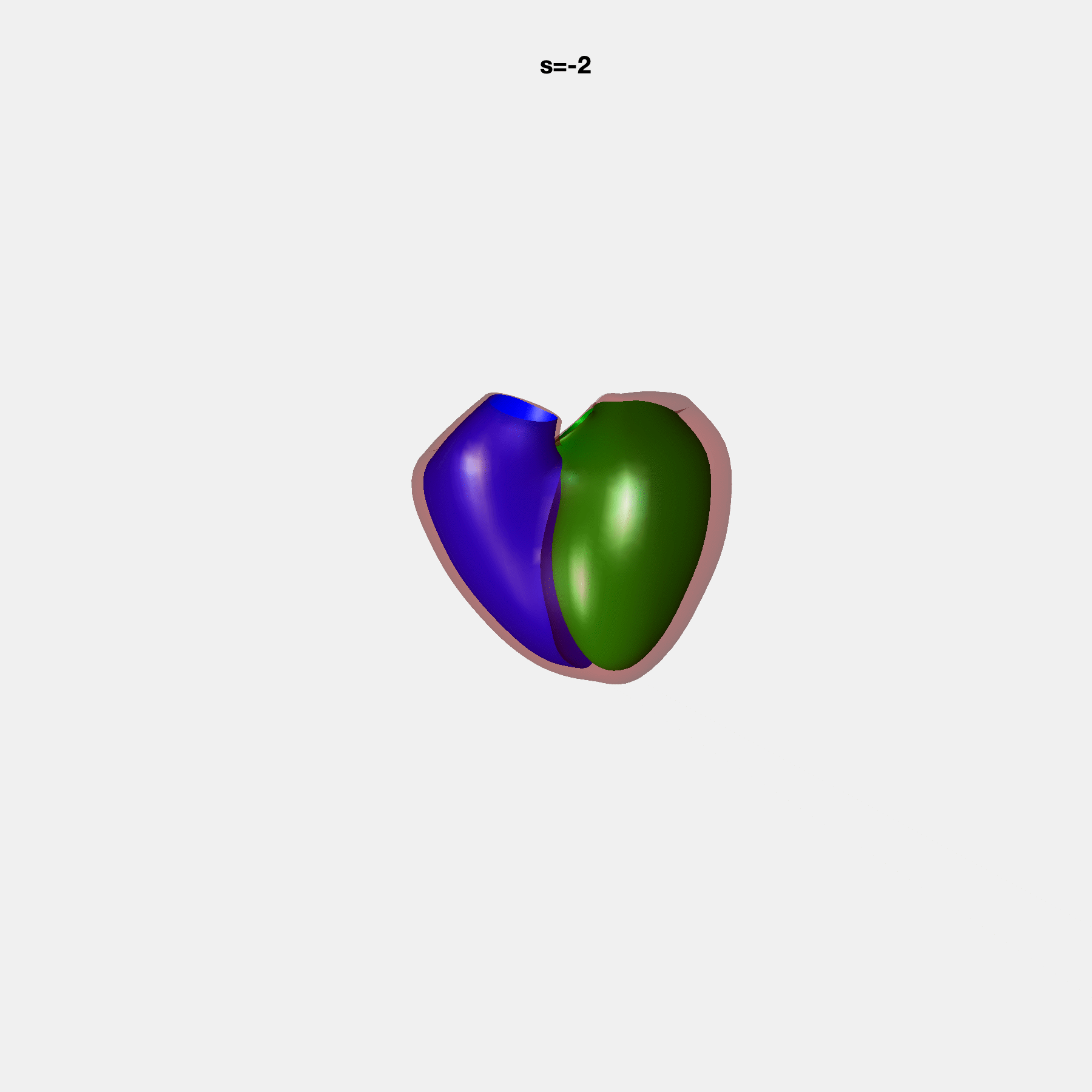

### SM20.gif

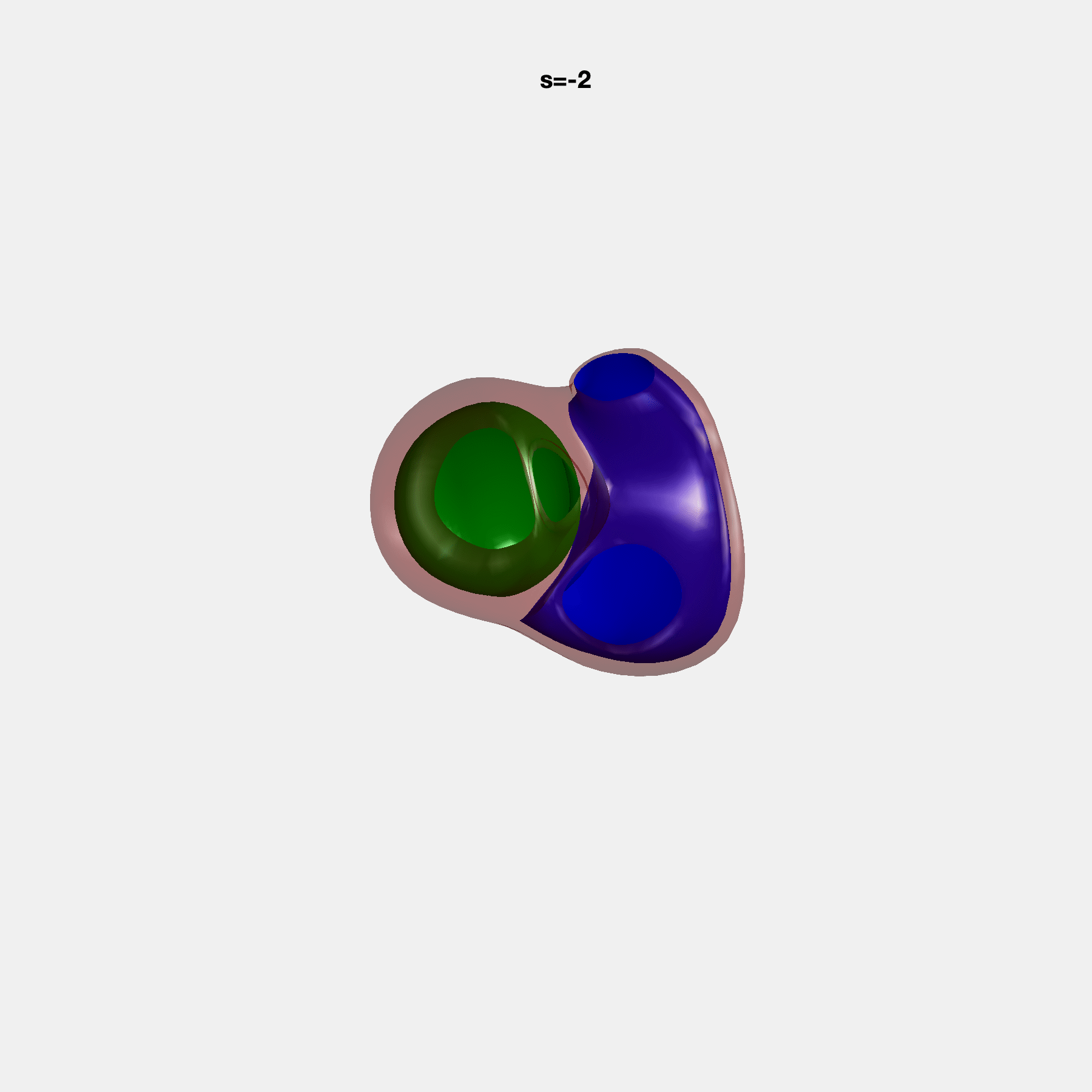

### SM21.gif

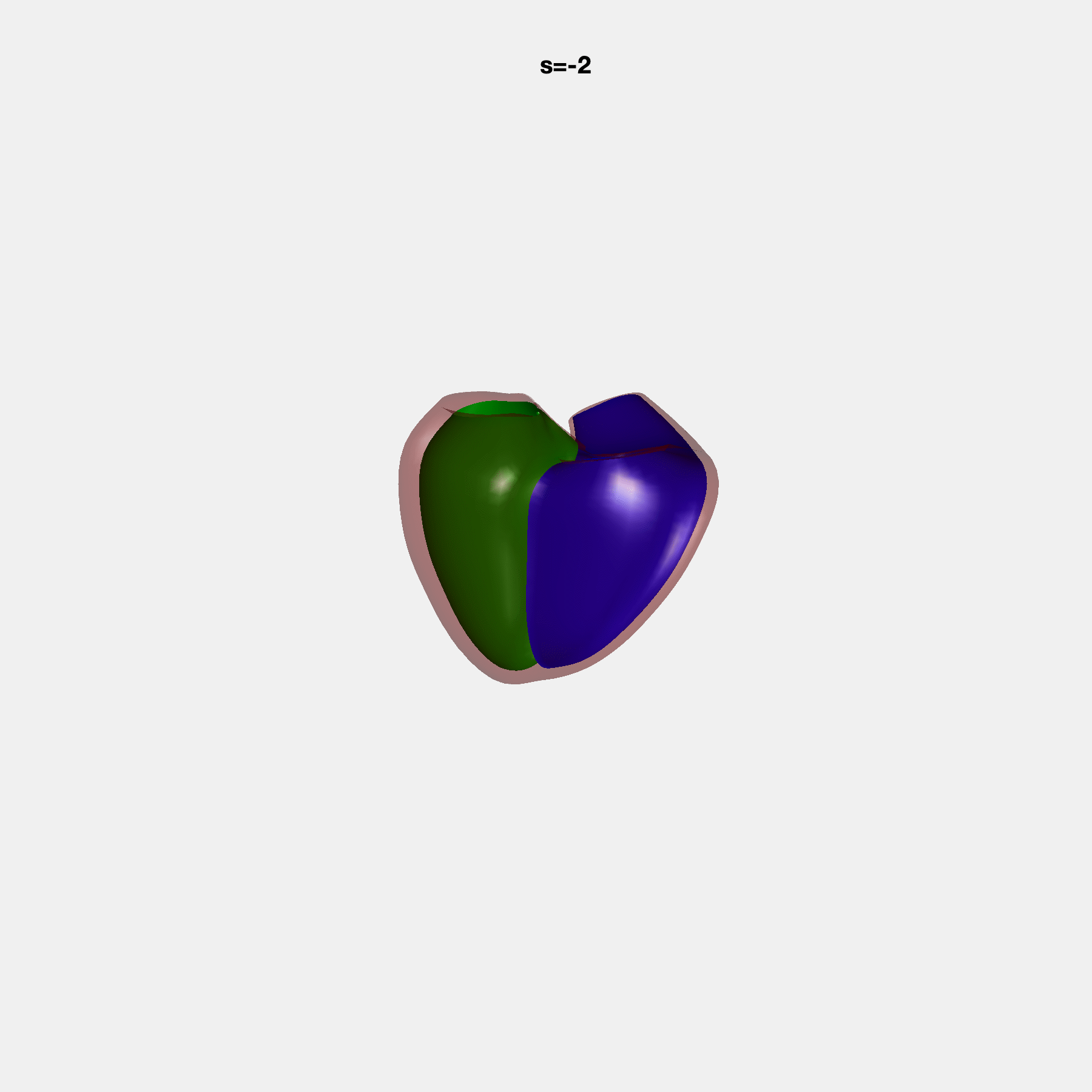

### SM22.gif

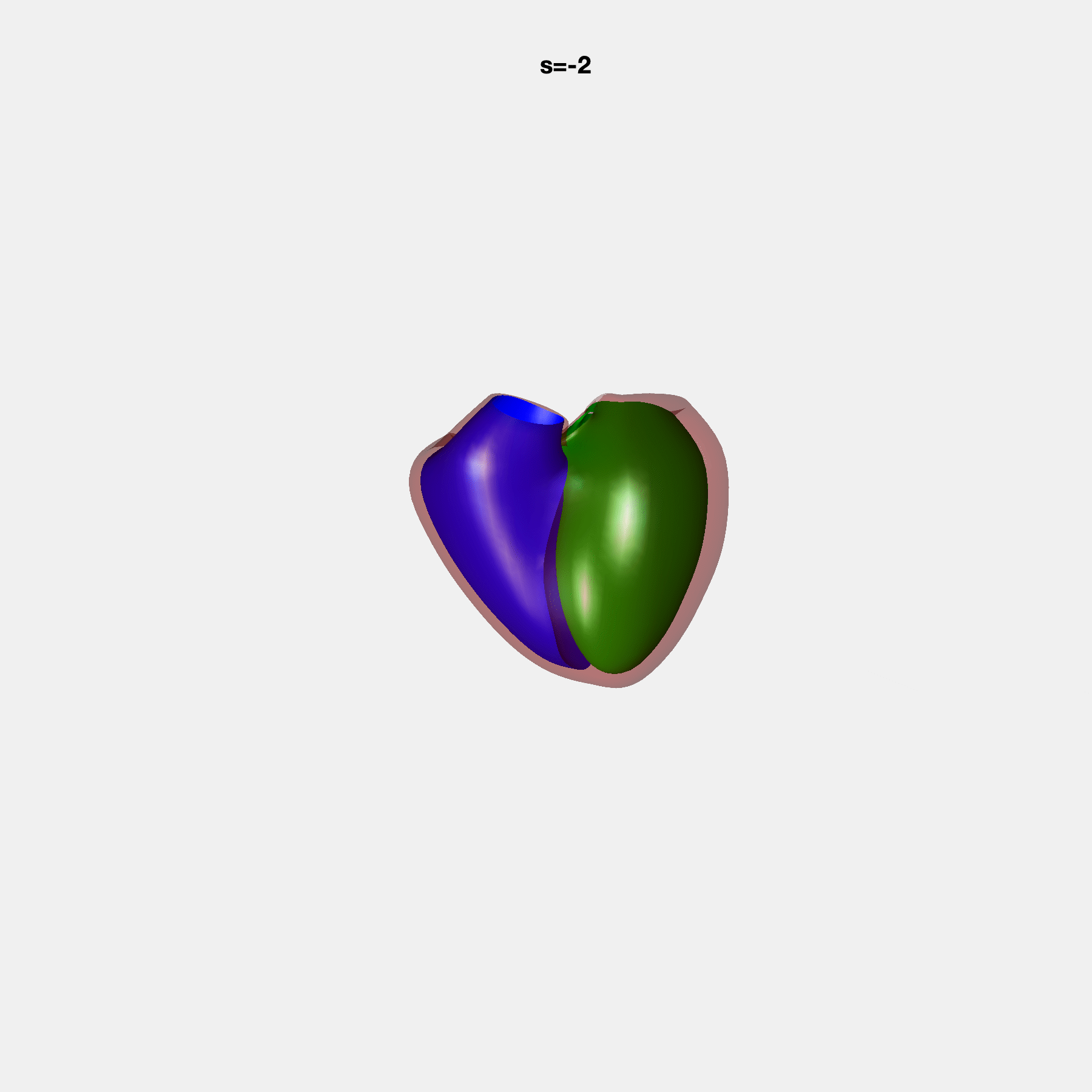

### SM23.gif

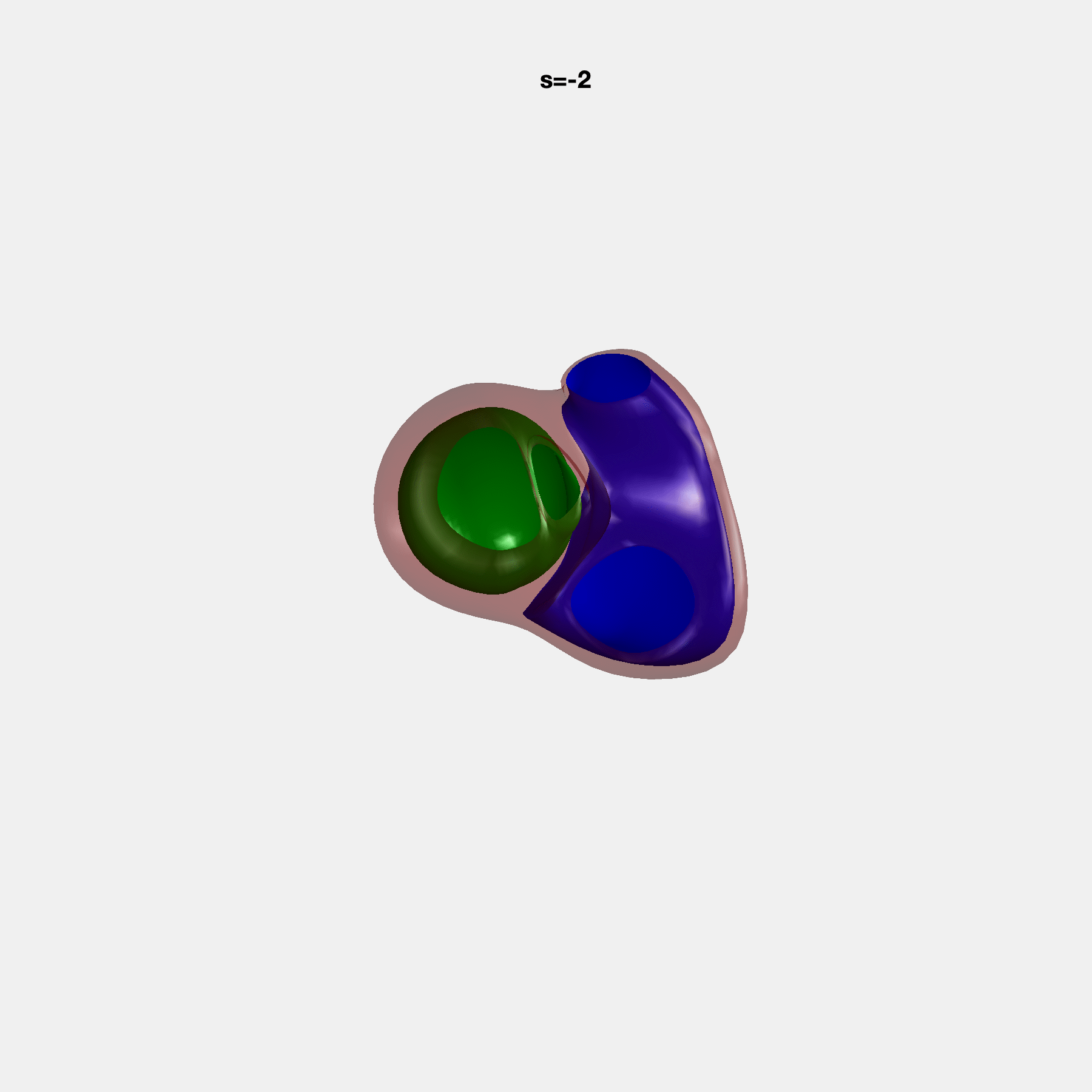

### SM24.gif

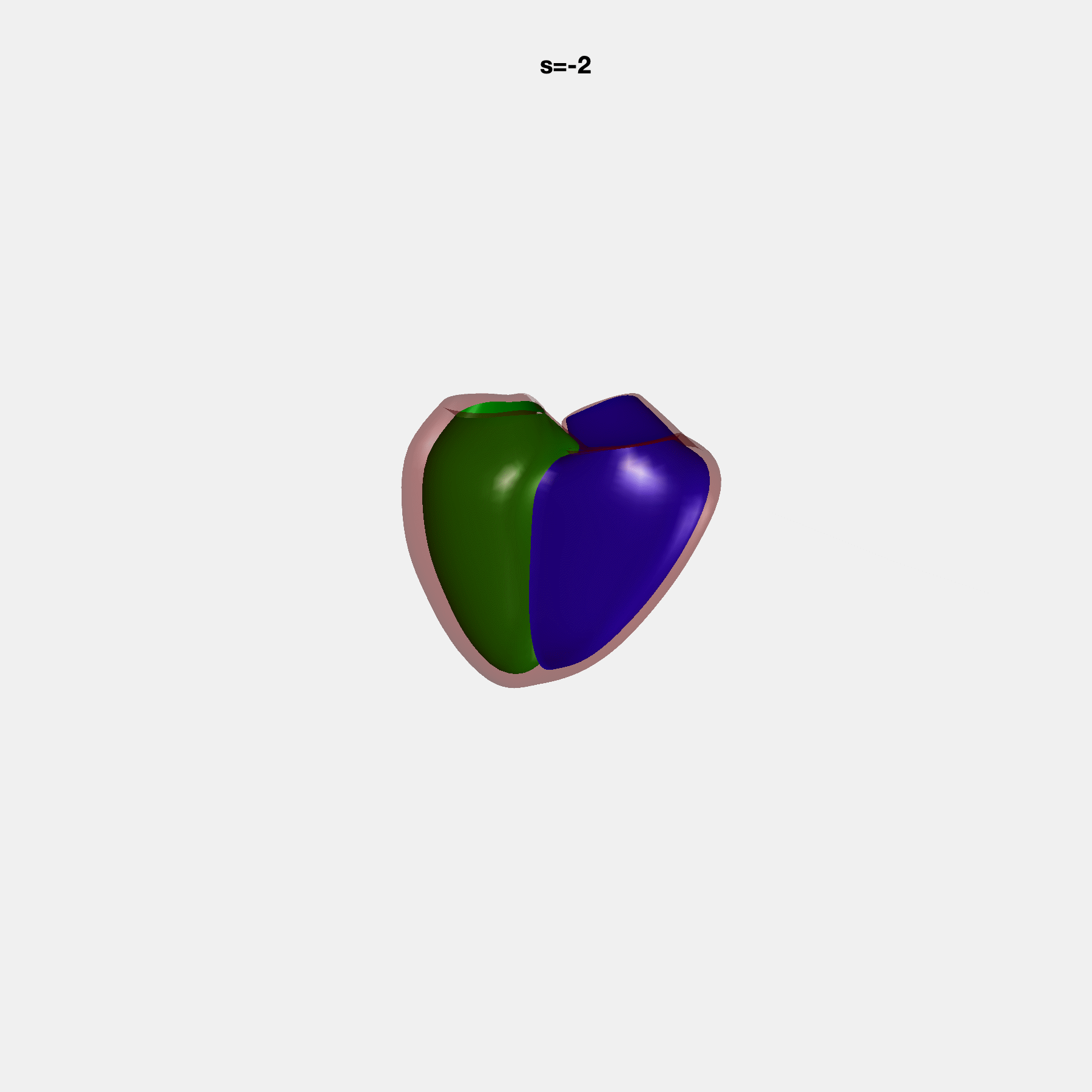

### SM25.gif

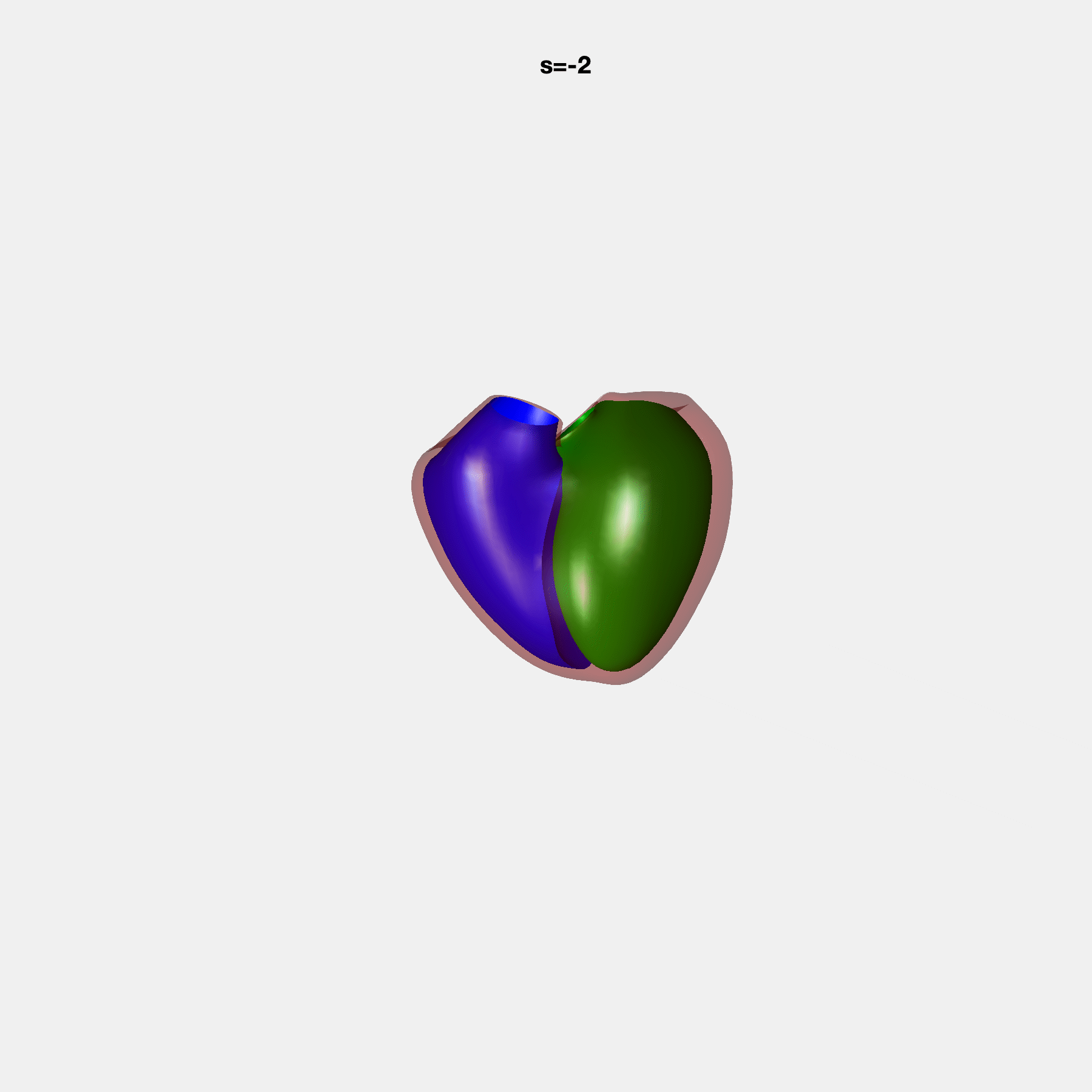

### SM26.gif

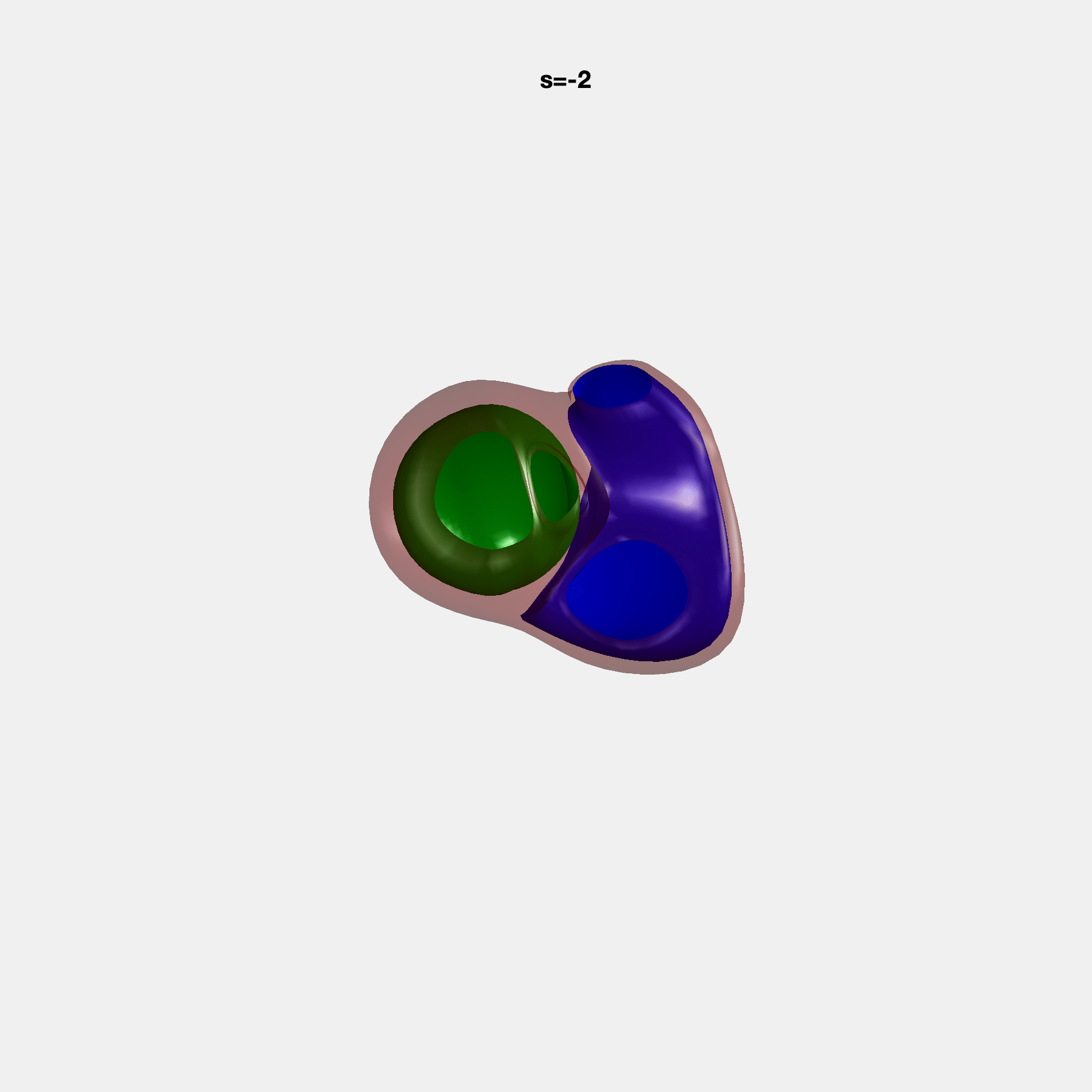

### SM27.gif

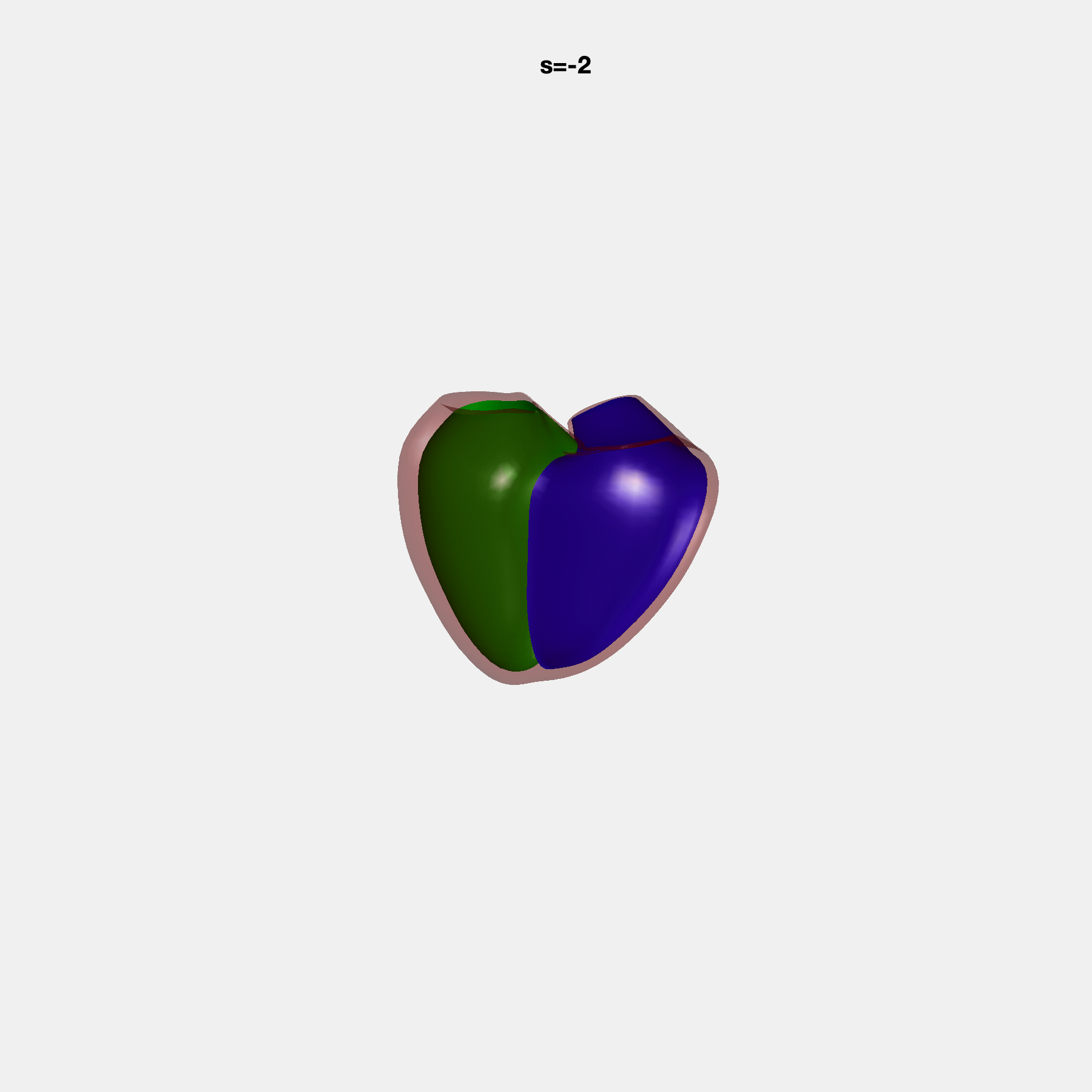

### SM28.gif

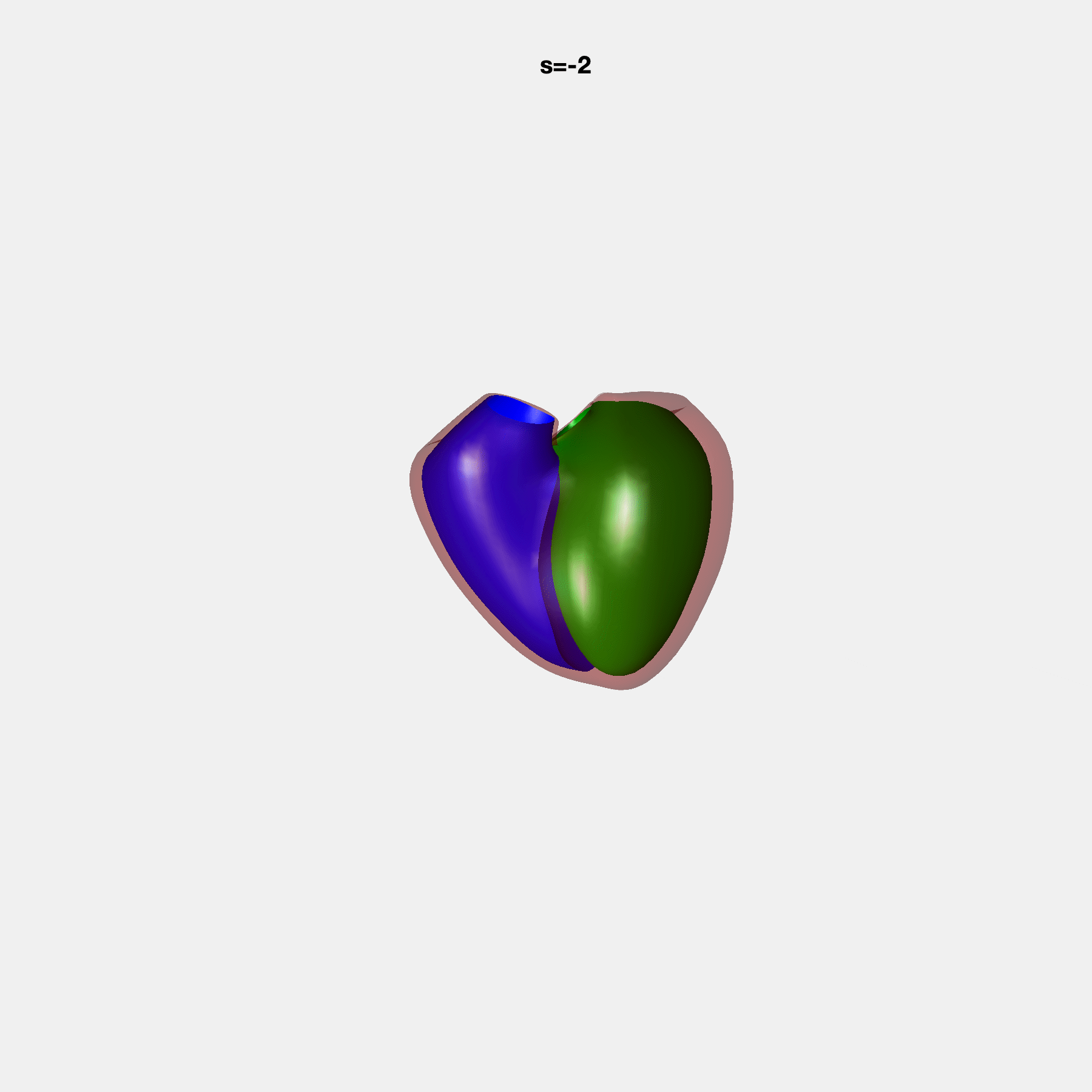

### SM29.gif

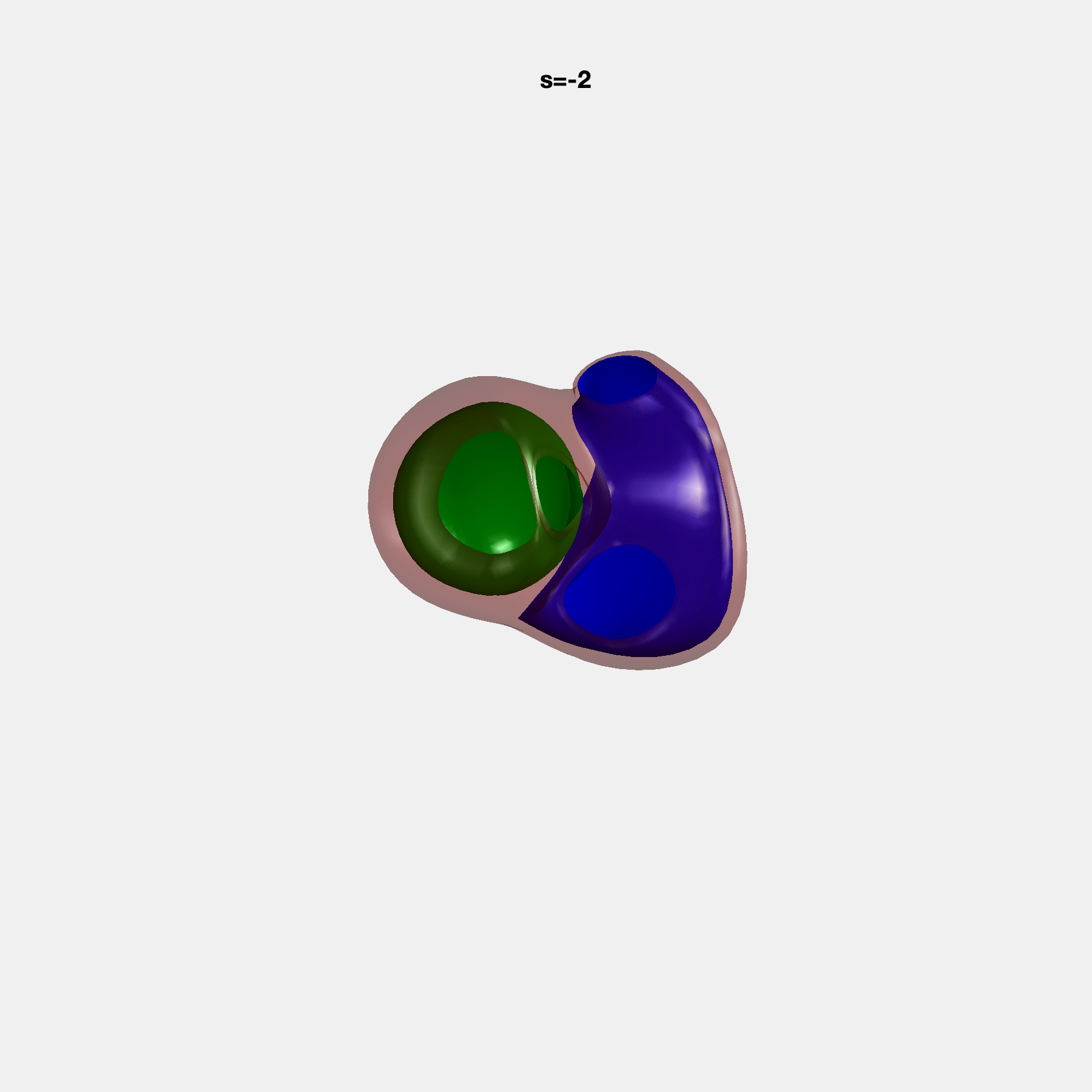

### SM30.gif

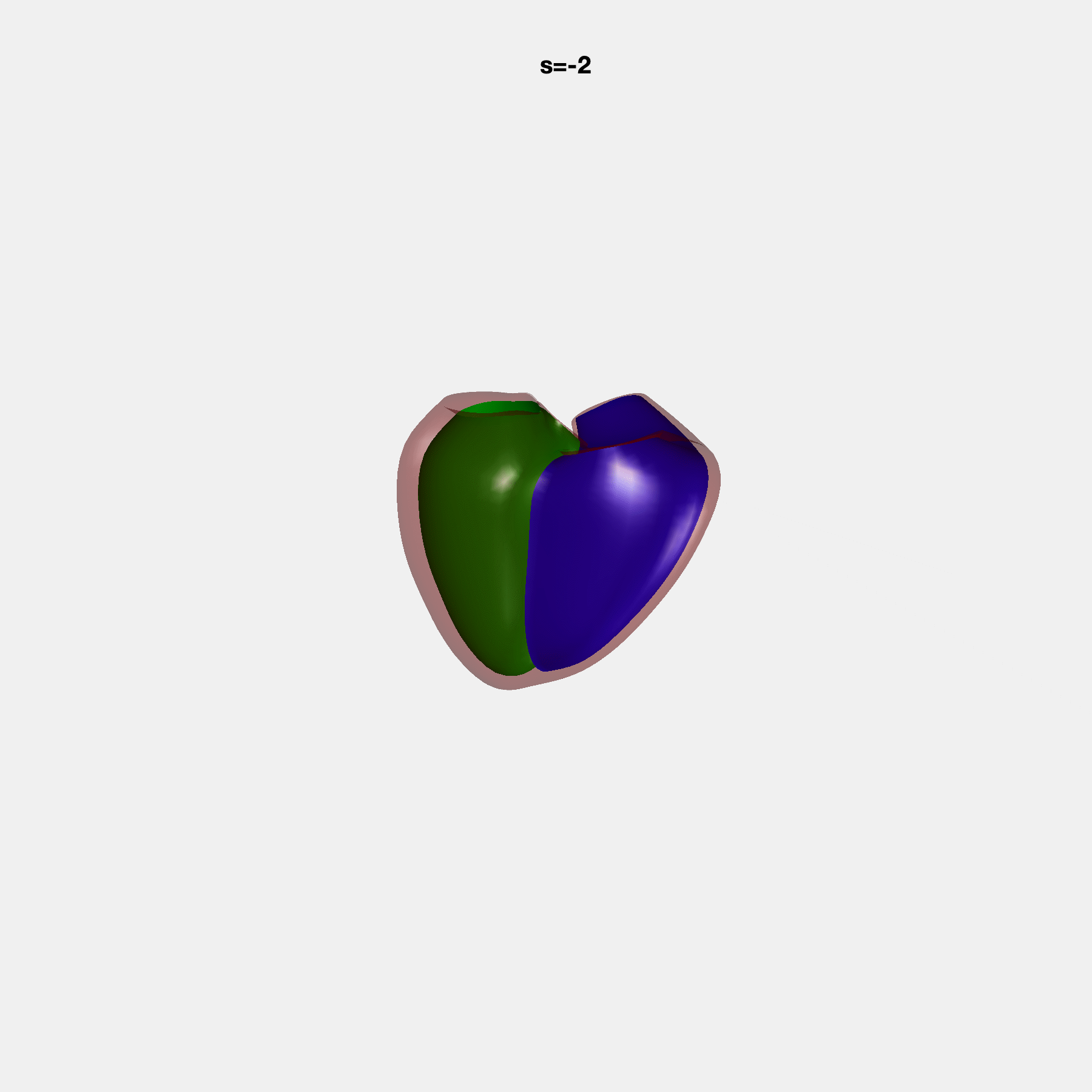

### SM31.gif

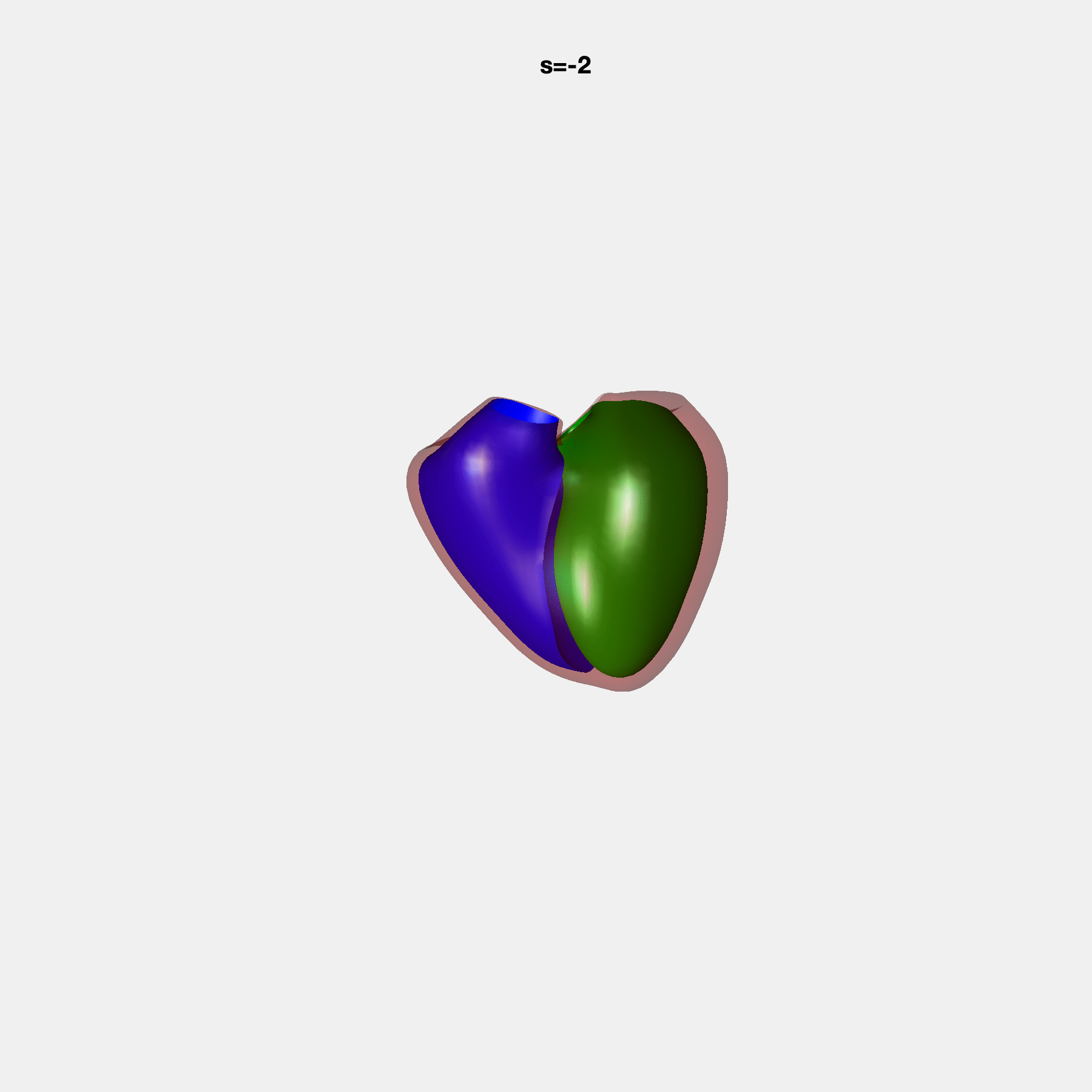

### SM32.gif

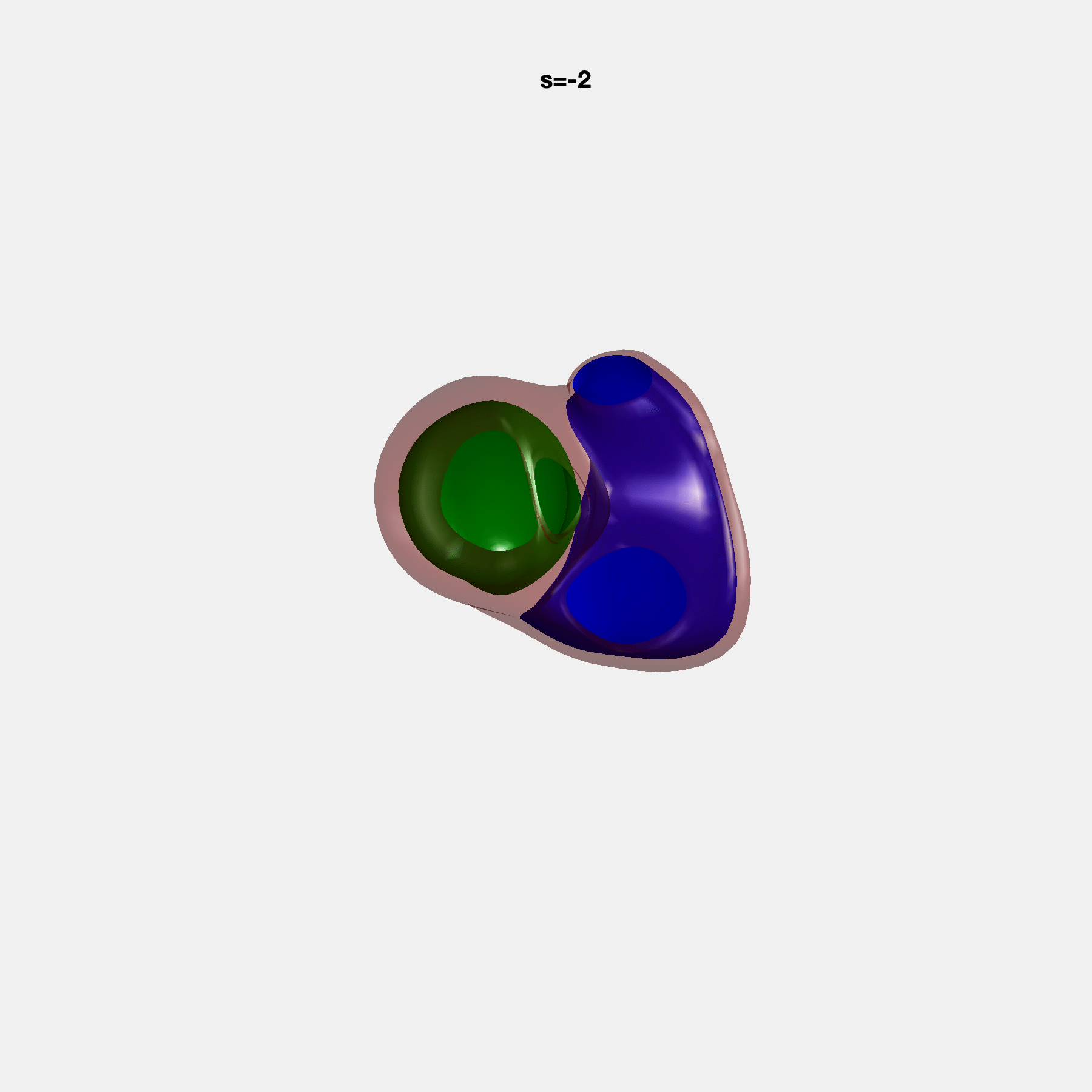

### SM33.gif

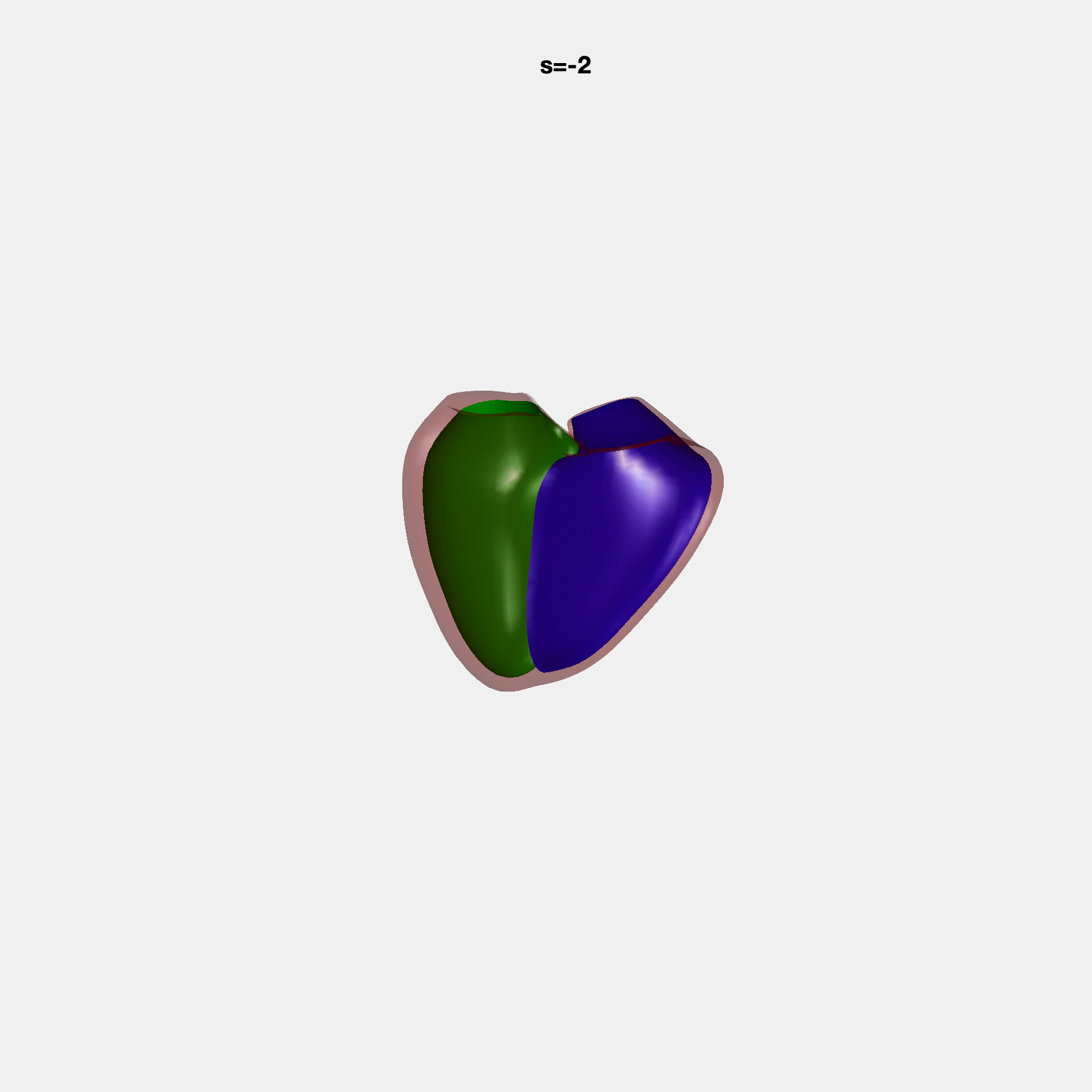

### SM34.gif

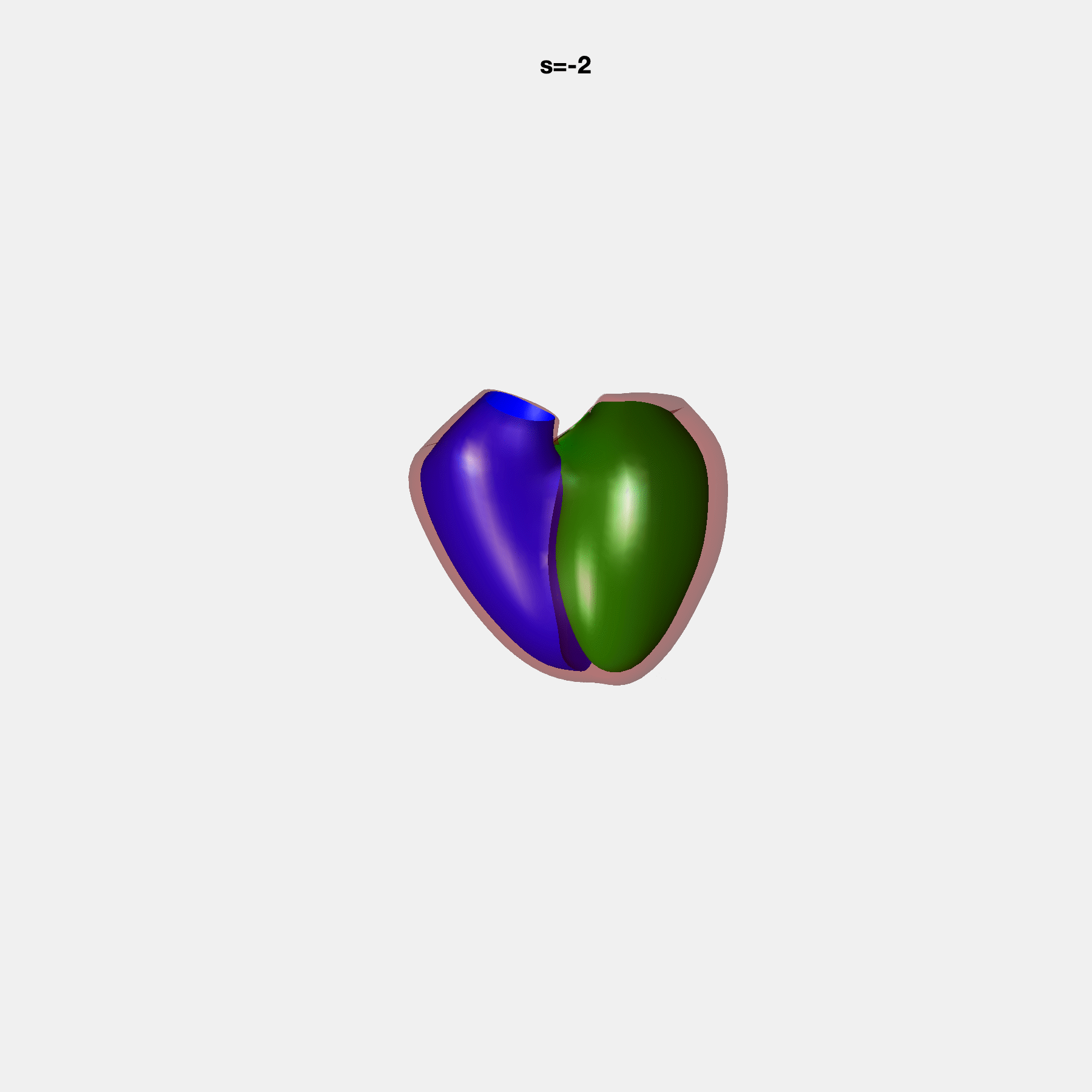

### SM35.gif

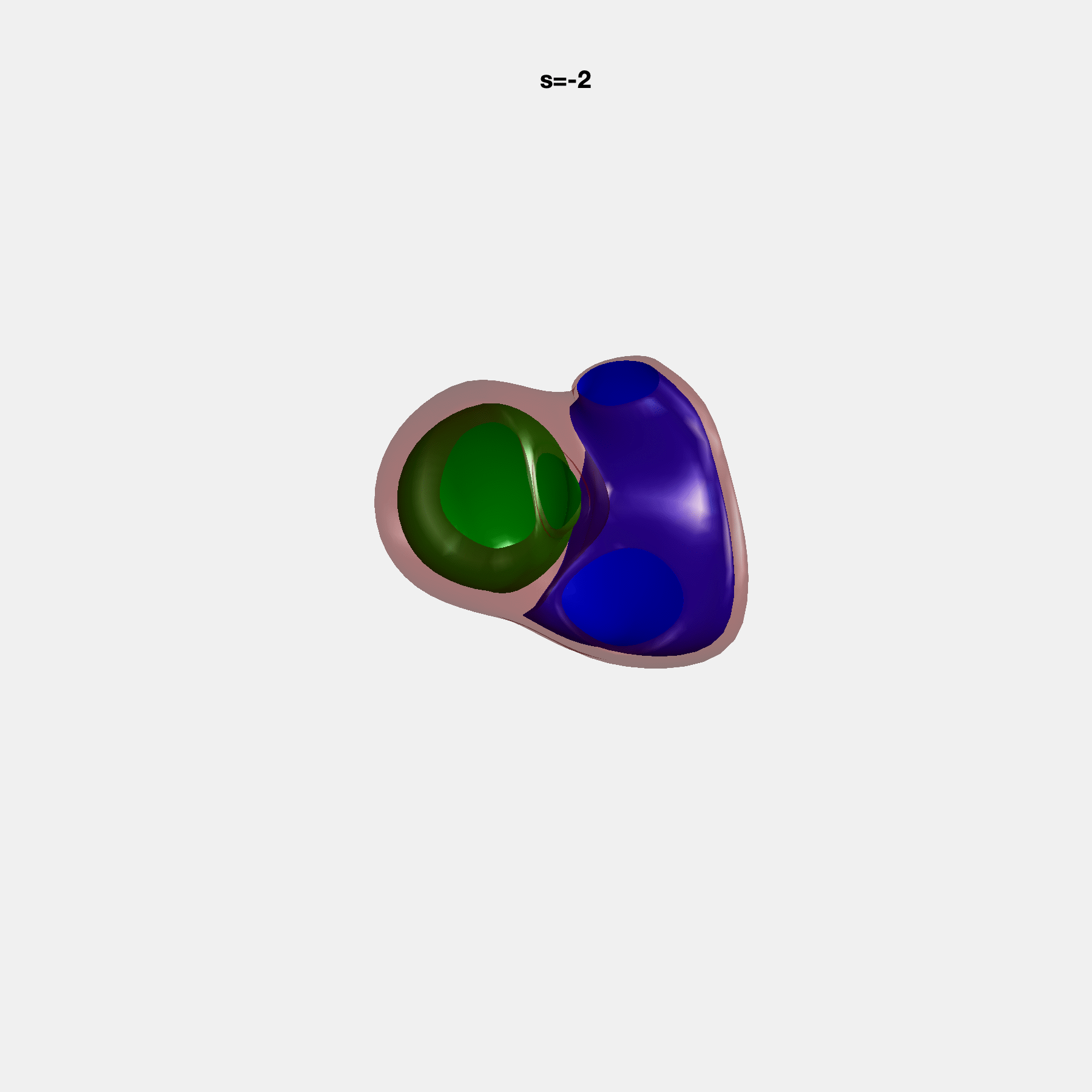

### SM36.gif

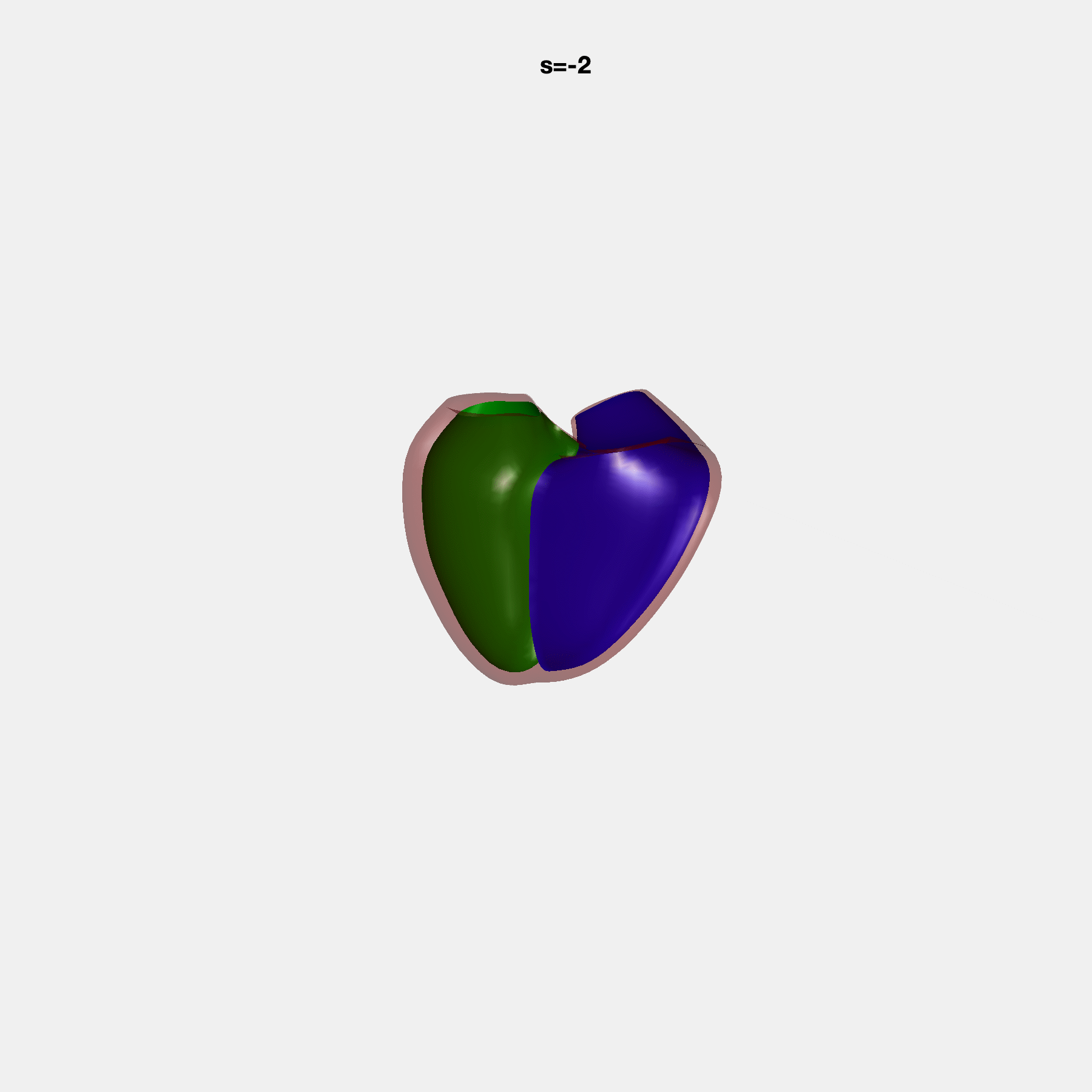

### SM37.gif

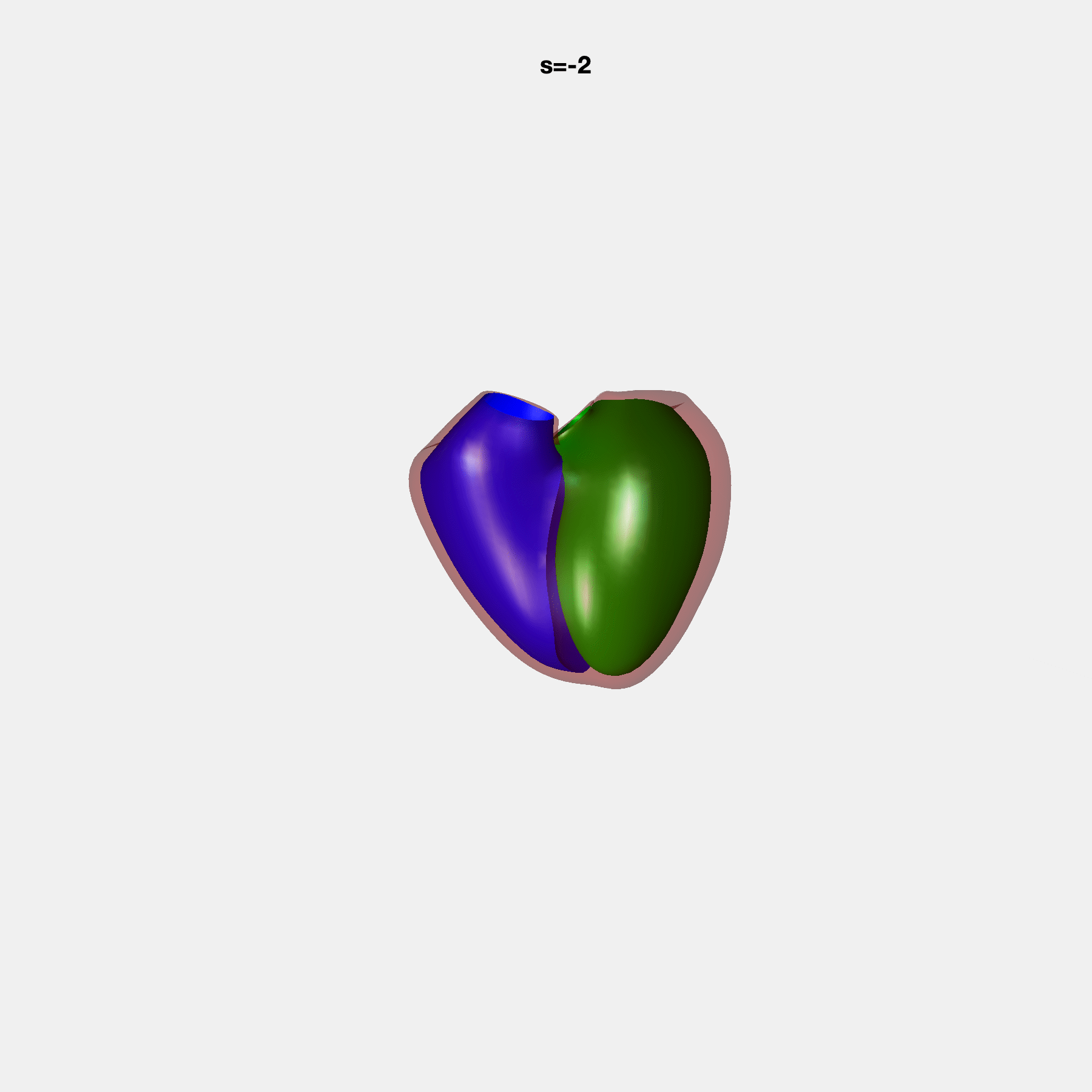

### SM38.gif

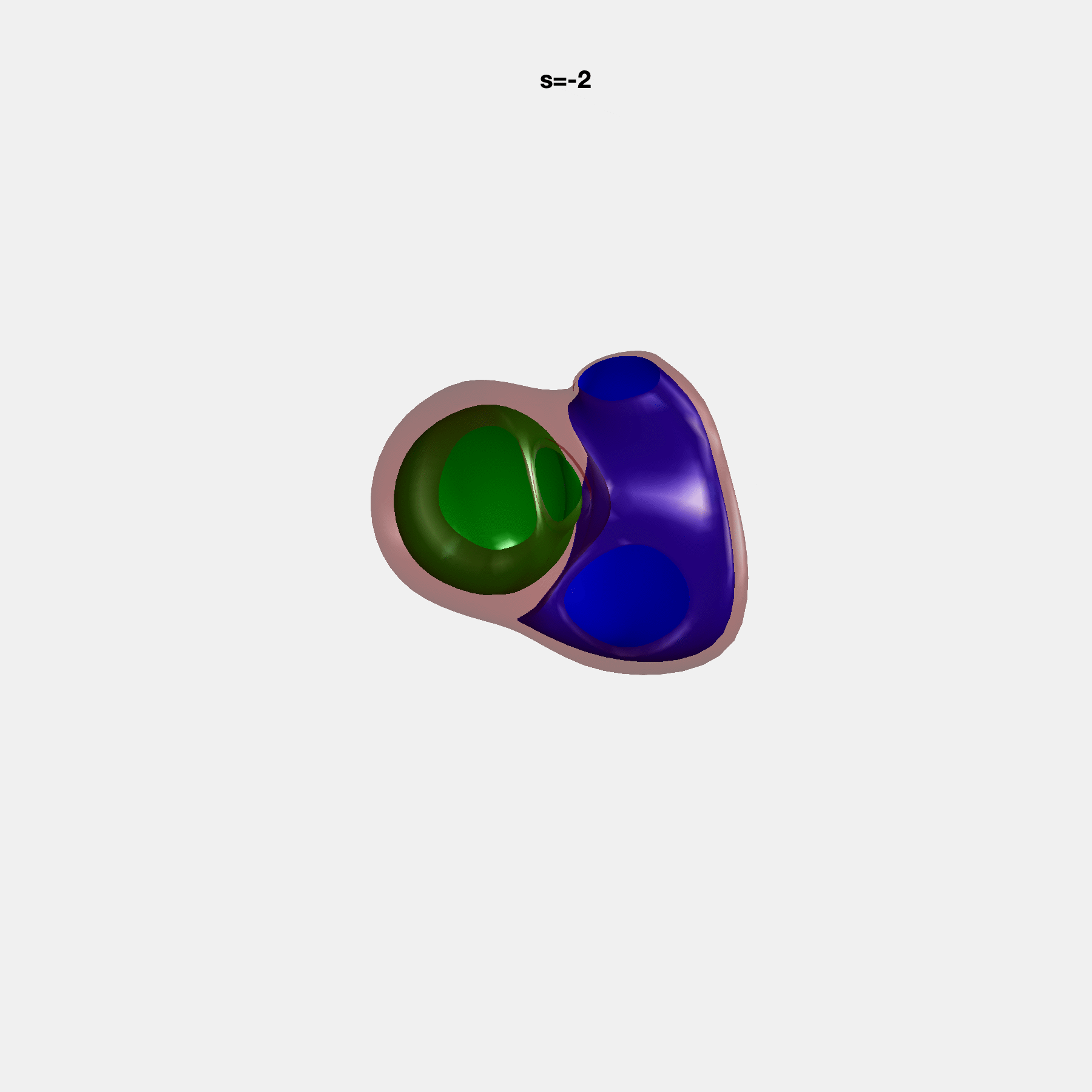

### SM39.gif

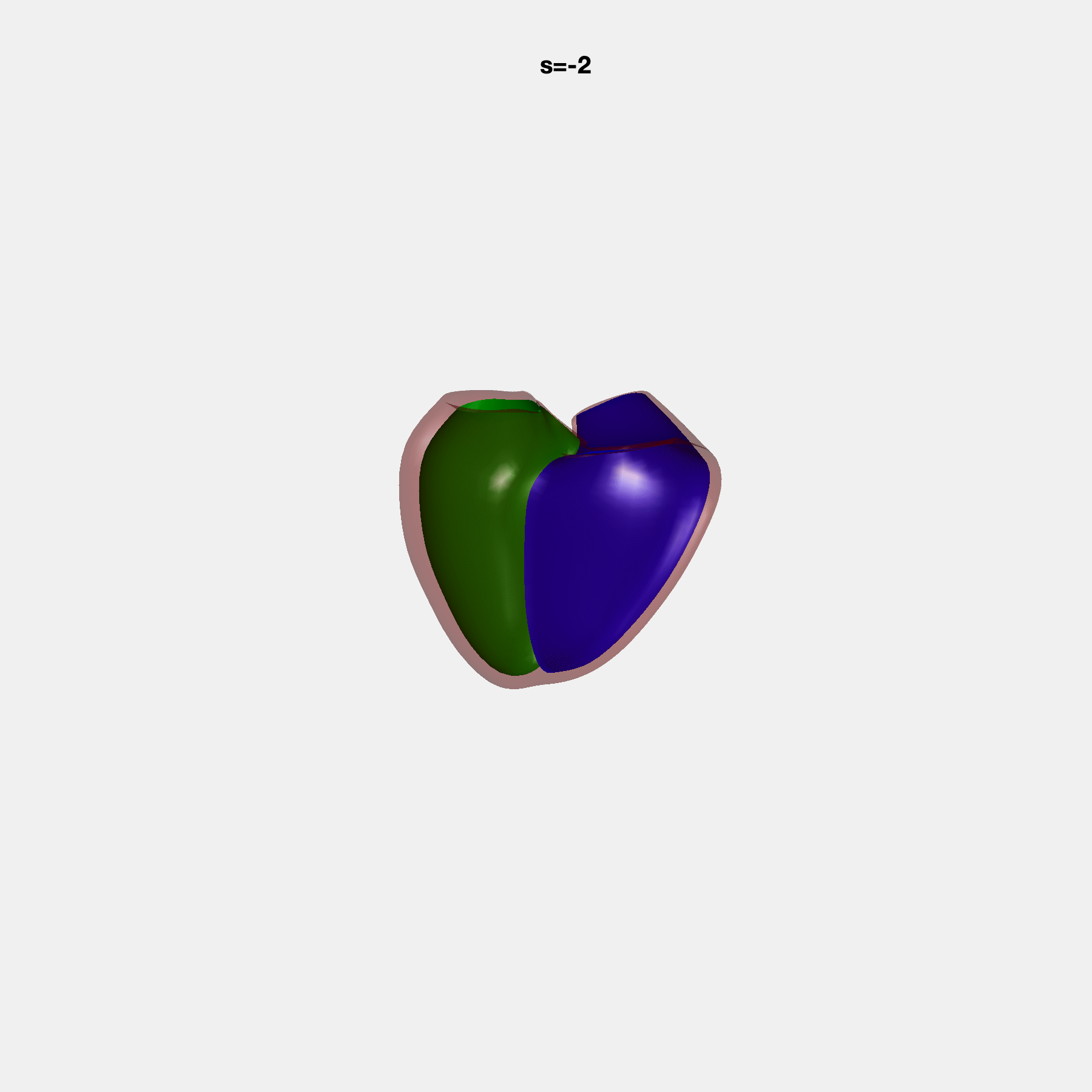

### SM40.gif

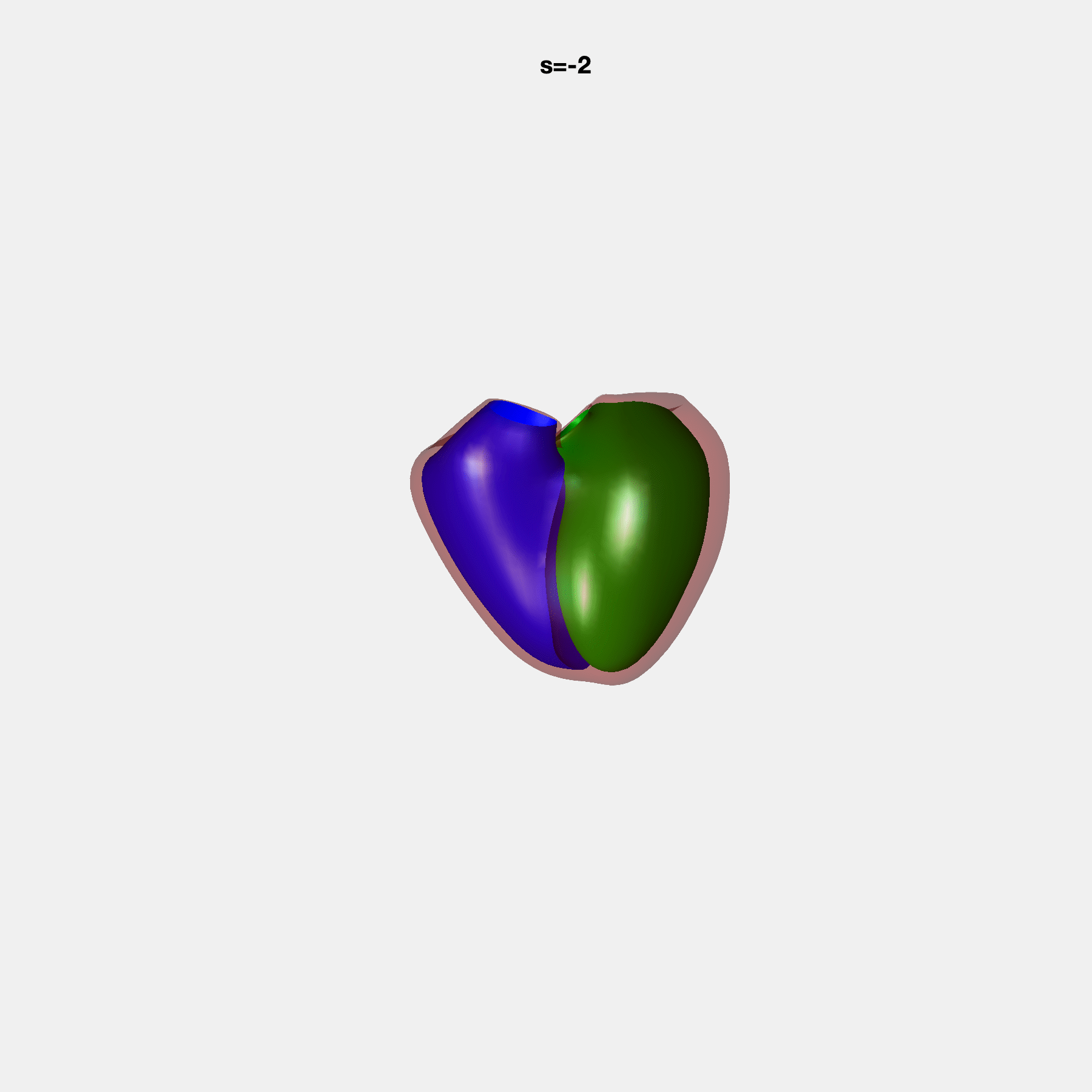

### SM41.gif

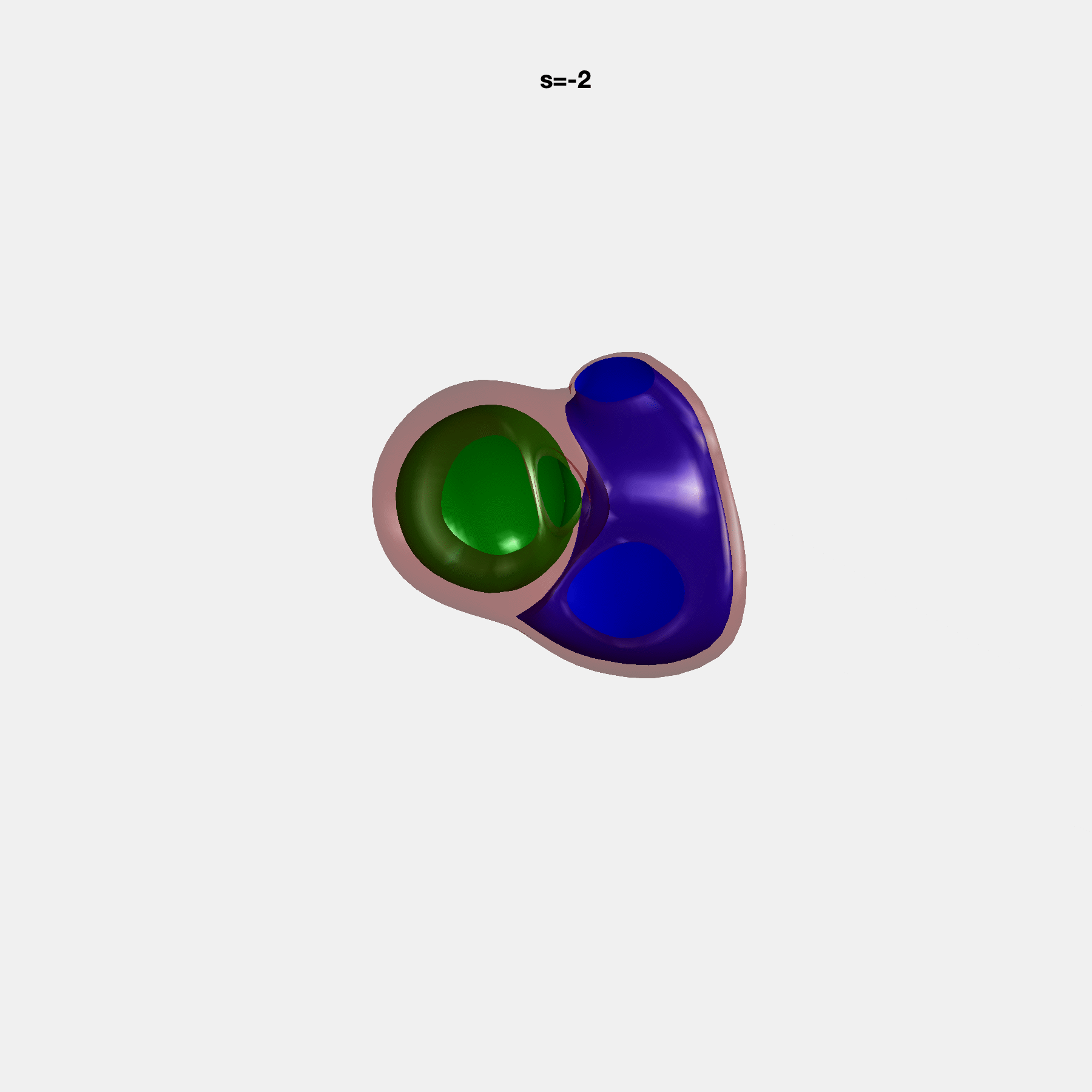

### SM42.gif

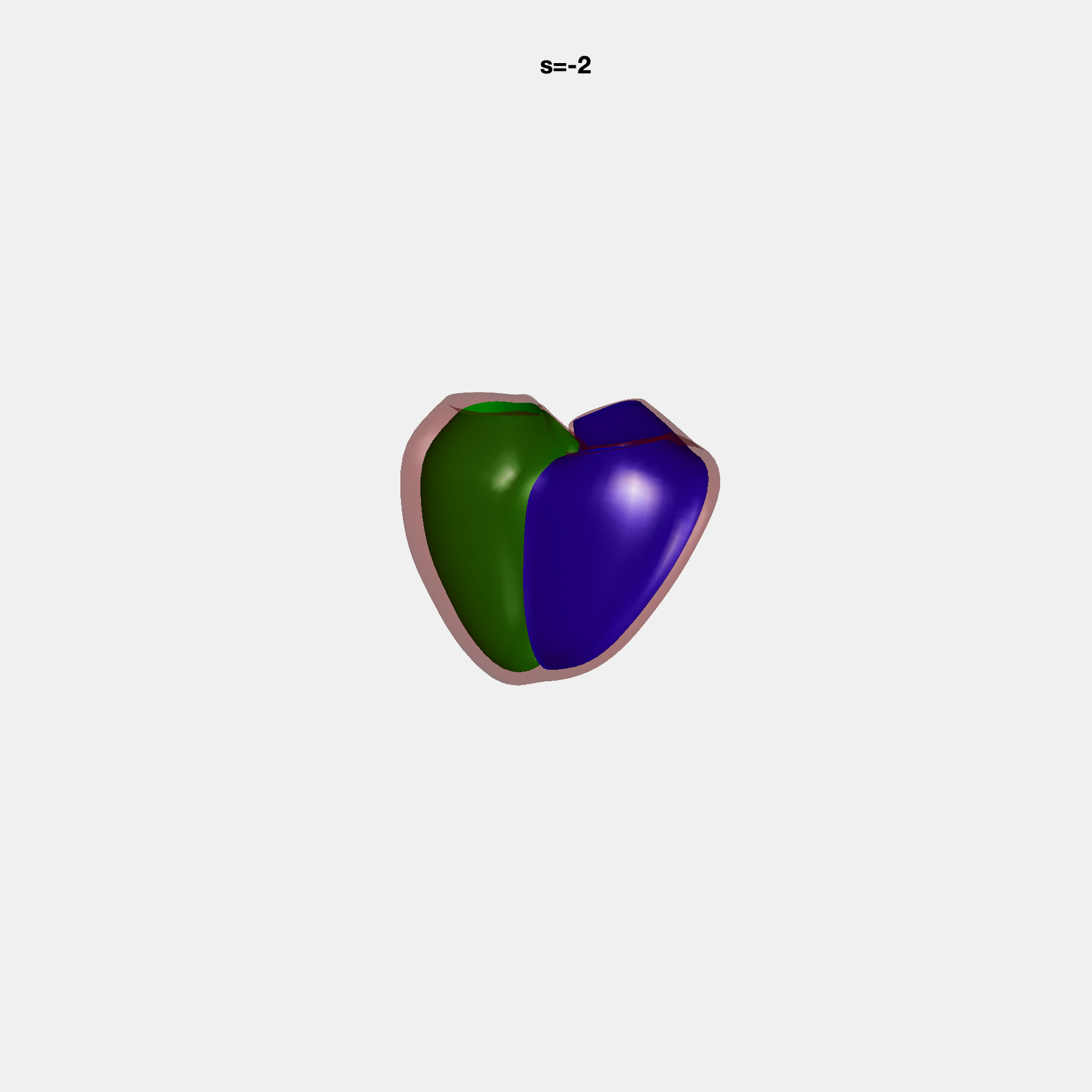
